# Adverse events of iron and/or erythropoiesis-stimulating agent therapy in preoperatively anaemic elective surgery patients: a systematic review

**DOI:** 10.1101/2022.03.21.22272600

**Authors:** Jorien Laermans, Hans Van Remoortel, Bert Avau, Geertruida Bekkering, Jørgen Georgsen, Paola Maria Manzini, Patrick Meybohm, Yves Ozier, Emmy De Buck, Veerle Compernolle, Philippe Vandekerckhove

## Abstract

**Background:** Iron supplementation and erythropoiesis-stimulating agent (ESA) administration represent the hallmark therapies in preoperative anaemia treatment, as reflected in a set of evidence-based treatment recommendations made during the 2018 International Consensus Conference on Patient Blood Management. However, little is known about the safety of these therapies. This systematic review investigated the occurrence of adverse events (AEs) during or after treatment with iron and/or ESAs.

**Methods:** Five databases (The Cochrane Library, MEDLINE, Embase, Transfusion Evidence Library, Web of Science) and two trial registries (ClinicalTrials.gov, WHO ICTRP) were searched until November 6 2020. Randomized controlled trials (RCTs), cohort and case-control studies investigating any AE during or after iron and/or ESAs administration in adult elective surgery patients with preoperative anaemia were eligible for inclusion, and judged using the Cochrane Risk of Bias tools. The GRADE approach was used to assess the overall certainty of evidence.

**Results:** 26 RCTs and 16 cohort studies involving a total of 6062 patients were included, providing data on 6 comparisons: (1) Intravenous (IV) versus oral iron, (2) IV iron versus usual care/no iron, (3) IV ferric carboxymaltose versus IV iron sucrose, (4) ESA+iron versus control (placebo and/or iron, no treatment), (5) ESA+IV iron versus ESA+oral iron, and (6) ESA+IV iron versus ESA+IV iron (different ESA dosing regimens). Most AE data concerned mortality/survival (n=24 studies), thromboembolic (n=22), infectious (n=20), cardiovascular (n=19) and gastrointestinal (n=14) AEs. Very low-certainty of evidence was assigned to all but one outcome category. This uncertainty results from both low quantity and quality of AE data due to high risk of bias caused by limitations in study design, data collection and reporting.

**Conclusions:** It remains unclear if ESA and/or iron therapy is associated with AEs in preoperatively anaemic elective surgery patients. Future trial investigators should pay more attention to the systematic collection, measurement, documenting and reporting of AE data.

## Introduction

Transfusion of blood components can be a life-saving intervention, but comes with the risks of transfusion reactions and transmission of bloodborne infections. To optimize the care of patients who might need transfusion and minimize the patient’s exposure to allogeneic blood products, a multidisciplinary approach has been developed and termed ‘Patient Blood Management’ (PBM)(1). PBM encompasses all aspects surrounding the transfusion decision-making process through three pillars: (1) addressing pre-existing preoperative anaemia, (2) minimizing intraoperative blood loss, and (3) applying appropriate transfusion triggers to ensure rational allogeneic blood product use. In an attempt to analyse the scientific evidence on all PBM aspects through systematic literature searches, and reach international consensus using transparent, rigorous and quality-controlled decision-making, an international consortium of scientific organizations in the field of blood transfusion coordinated a consensus meeting on evidence-based PBM. During this 2018 International Consensus Conference on Patient Blood Management (ICC-PBM), a set of 10 clinical and 12 research recommendations were formulated using the GRADE methodology (2, 3).

One PBM pillar focusses on addressing pre-existing preoperative anaemia. Preoperative anaemia is associated with increased perioperative blood transfusion requirements, increased risk of perioperative infection, mortality, postoperative complications and extended hospital stay (4-7). During the ICC-PBM, four clinical recommendations (Table 1) were formulated concerning the treatment of preoperative anaemia, using iron supplementation (in case of iron-deficiency anaemia) and/or erythropoiesis-stimulating agents (ESAs) (2).

**Table 1.** Clinical recommendations formulated during the ICC-PBM as published previously (2).

| Clinical recommendation | Strength of recommendation and certainty of evidence |
| --- | --- |
| 1. Detection and management of preoperative anaemia early enough before major elective surgery | Strong recommendation, low-certainty evidence |
| 2. Use of iron supplementation to reduce red blood cell transfusion rate in adult preoperative patients with iron-deficient anaemia undergoing elective surgery | Conditional recommendation, moderate-certainty evidence |
| 3. Do not use erythropoiesis-stimulating agents routinely in general for adult preoperative patients with anaemia undergoing elective surgery | Conditional recommendation, low-certainty evidence |
| 4. Consider short-acting erythropoietins in addition to iron supplementation to reduce transfusion rates in adult preoperative patients with hemoglobin concentrations < 13 g/dL undergoing elective major orthopaedic surgery | Conditional recommendation, low-certainty evidence |

As effectiveness is just one aspect to consider in making a balanced treatment recommendation, the expert panel recommended to investigate the use of short-acting erythropoietins and iron supplementation in adult preoperative patients undergoing elective surgery, with focus on long-term (un)desirable effects, optimal dose, type of surgery (particularly in cancer surgery), copresence of iron deficiency, and cost-effectiveness (2).

Therefore, in a follow-up project, three full systematic reviews were conducted to gather the best available scientific evidence on the effectiveness (review 1)(8), safety (review 2) and cost-effectiveness (review 3)(9) of iron and/or ESA therapy in adult patients with preoperative anaemia undergoing elective surgery. The current systematic review (review 2) focused on the occurrence of adverse events (AEs) during or after treatment with iron and/or ESAs.

## Methods

This systematic review was not prospectively registered, but was carried out in accordance with the pre-defined methodological standards of the Centre for Evidence-Based Practice (10). Eligibility criteria and data synthesis plans were established *a priori* by the reviewers (JL and HVR) and approved by a third external methodological expert (GB) and 4 PBM experts (JG, PM, PMM, YO). The reporting of this review adheres to the PRISMA harms checklist (11) (completed checklist in Additional file 1).

### Eligibility criteria

This review’s PICO question was: “In elective surgery patients with preoperative anaemia (P), is the use of iron and/or ESA therapy (I), linked to AEs (O)?”.

#### Population

Anaemic adults (≥18 years) scheduled for elective surgery were eligible for inclusion. Studies were included if the baseline haemoglobin (Hb) levels of the study participants were in line with the World Health Organization (WHO) criteria for anaemia, i.e. <13 g/dl for men and <12 g/dl for women. Different criteria for anaemia were accepted if the study investigators provided a clear definition of anaemia, or if no clear definition was provided but baseline Hb levels were <13 g/dl in all patients (based on the upper limit of the 99% confidence interval (CI) (mean Hb levels) or the 75^th^ percentile of the Interquartile Range (IQR) (median Hb levels)). Studies in pregnant women, children and non-elective surgery patients were excluded.

#### Intervention

Studies were eligible if they investigated the administration effect of iron and/or ESAs, regardless of treatment dose, duration and formulation (enteral or parenteral). Studies were only included if the administration was started, but not necessarily ended, during the preoperative period. If patients received other cointerventions (e.g. vitamins, folic acid, heparin), the study was only included if these cointerventions were identically administered to both (intervention and control) groups.

#### Comparison

Studies were included if they compared the intervention(s) of interest to at least one of the following control groups: placebo, no treatment, standard of care (as per each trial protocol), iron monotherapy, other type of iron therapy (e.g. IV versus oral), or other ESA dosing regimen.

#### Outcome

AEs were defined as “unfavourable or harmful outcomes that occur during, or after, the use of a drug or other intervention, but are not necessarily caused by it” (12).

Any AE occurring during/after iron and/or ESA administration was eligible for inclusion and classified by the reviewers (JL and HVR), a third external methodological expert (GB) and 4 PBM experts (JG, PM, PMM, YO) in one of 15 categories (listed in Table 2). The classification system of Szebeni, 2015 (13) served as a starting point and was supplemented with additional categories until consensus with the PBM experts was reached.

**Table 2.**
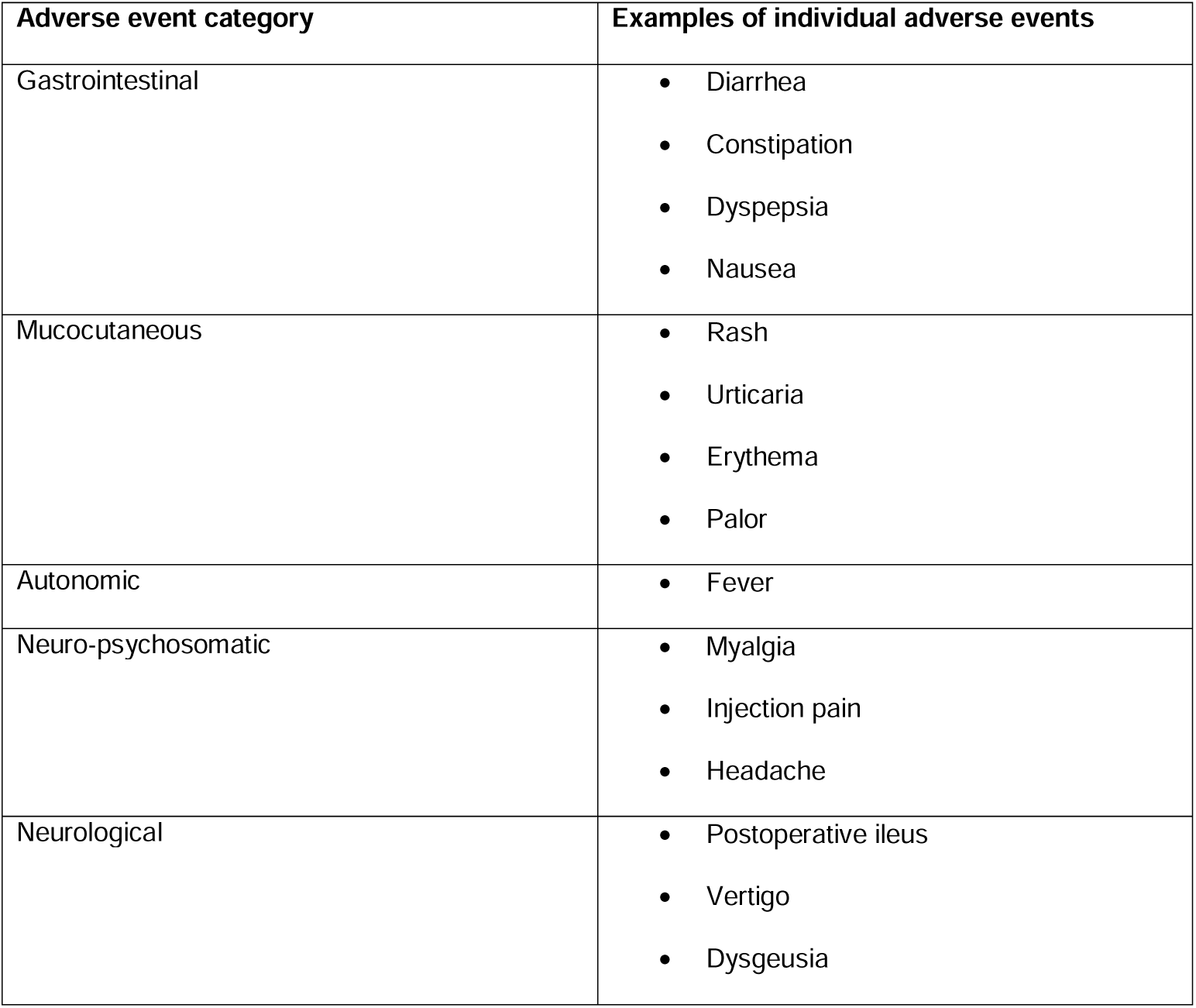
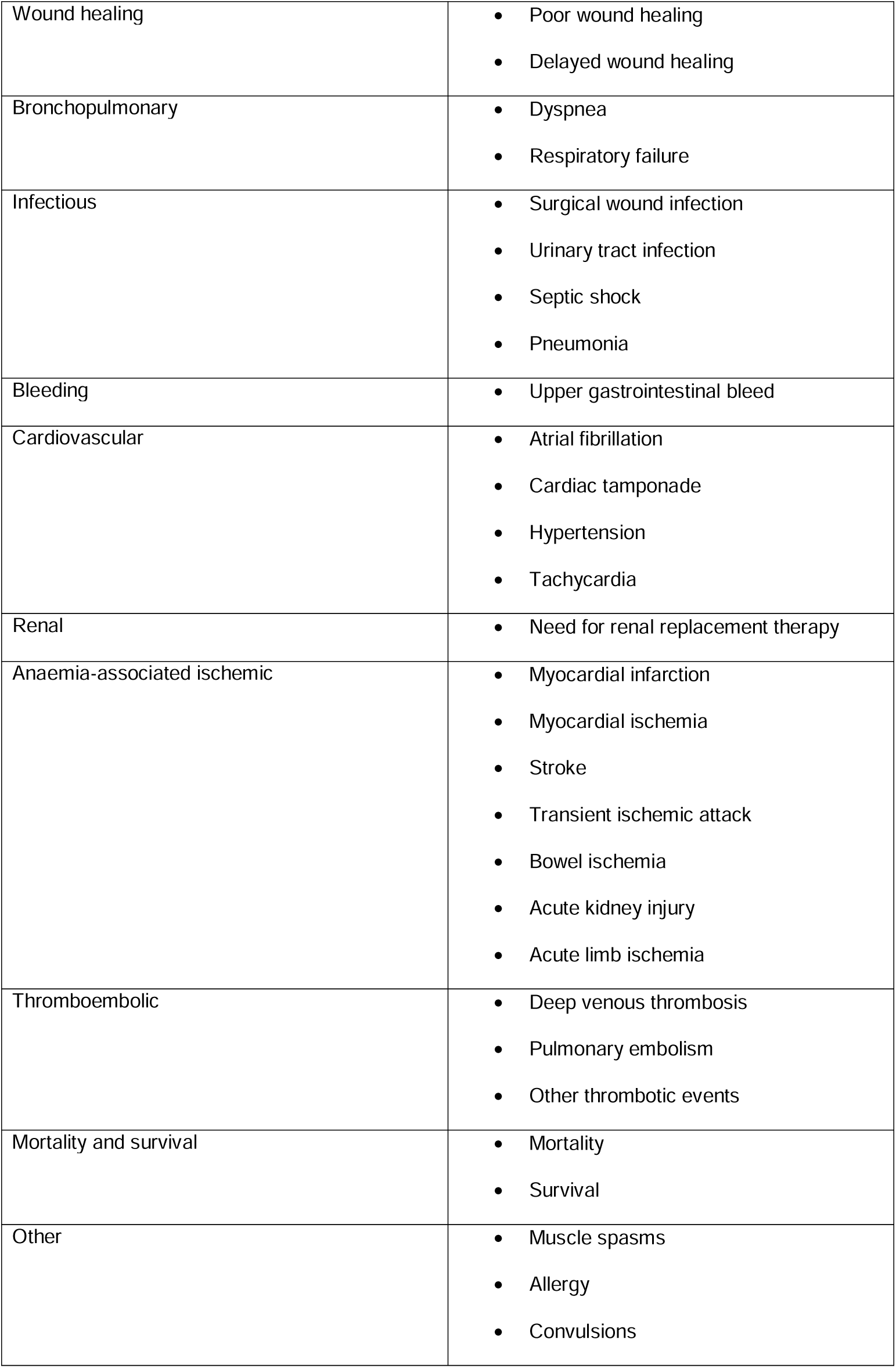
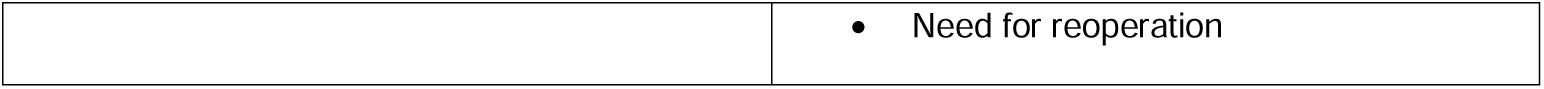
Adverse event categories.

| Adverse event category | Examples of individual adverse events |
| --- | --- |
| Gastrointestinal | • Diarrhea<br>• Constipation<br>• Dyspepsia<br>• Nausea |
| Mucocutaneous | • Rash<br>• Urticaria<br>• Erythema<br>• Palor |
| Autonomic | • Fever |
| Neuro-psychosomatic | • Myalgia<br>• Injection pain<br>• Headache |
| Neurological | • Postoperative ileus<br>• Vertigo<br>• Dysgeusia |
| Wound healing | • Poor wound healing<br>• Delayed wound healing |
| Bronchopulmonary | • Dyspnea<br>• Respiratory failure |
| Infectious | • Surgical wound infection<br>• Urinary tract infection<br>• Septic shock<br>• Pneumonia |
| Bleeding | • Upper gastrointestinal bleed |
| Cardiovascular | • Atrial fibrillation<br>• Cardiac tamponade<br>• Hypertension<br>• Tachycardia |
| Renal | • Need for renal replacement therapy |
| Anaemia-associated ischemic | • Myocardial infarction<br>• Myocardial ischemia<br>• Stroke<br>• Transient ischemic attack<br>• Bowel ischemia<br>• Acute kidney injury<br>• Acute limb ischemia |
| Thromboembolic | • Deep venous thrombosis<br>• Pulmonary embolism<br>• Other thrombotic events |
| Mortality and survival | • Mortality<br>• Survival |
| Other | • Muscle spasms<br>• Allergy<br>• Convulsions |
|  | • Need for reoperation |

AEs were classified into 15 categories (based on the classification system by Szebeni, 2015 (13)). Examples of individual adverse events listed are non-exhaustive, except for the anaemia-associated ischemic and thromboembolic events.

#### Study design

Since many AEs are too uncommon or long-term to be observed within randomized trials (12), both controlled experimental studies (e.g. RCTs) as well as observational cohort and case-control studies were included.

#### Publication status

Published and non-published data were included. Study authors of ongoing or prematurely ended registered trials were contacted to obtain the expected completion/publication date or reason for termination (if not specified).

#### Other criteria

No date or language restrictions were applied.

### Data sources and searches

The following databases and trial registries were searched from inception up to November 6 2020 (search date: November 6 2020): The Cochrane Library (Cochrane Database of Systematic Reviews and Cochrane Central Register of Controlled Trials), MEDLINE (using the PubMed interface), Embase (using the Embase.com interface), Transfusion Evidence Library, Web of Science, WHO International Clinical Trials Registry Platform and Clinicaltrials.Gov. Search strings comprising both index terms and free text words were tailored to each database (Supplementary Table 1 in Additional file 2).

Reference lists and the 20 first related citations in PubMed of the included records were scanned for additional studies.

### Study selection

Two reviewers (JL and HVR) independently screened the title and abstracts and subsequently the full-texts of the identified references guided by the eligibility criteria, using the EPPI-Reviewer Web software (14). Discrepancies were resolved by discussion. Where necessary, a third reviewer was consulted (BA).

As AE data are notorious for their incomplete/poor reporting, study authors of studies that did not report on AEs but did meet the other eligibility criteria were contacted via email at least twice. If the authors confirmed that no AE data were collected, or did not reply, the study was excluded. If the authors supplied the reviewers with unpublished findings on AEs, the study was included.

### Data extraction

Data extraction was performed independently by two reviewers (JL and HVR). For each individual study, the following data were extracted: source (peer-reviewed publication: author, publication year and country; trial registration: trial registry number), study design, description of the population, definition of anaemia and iron-deficiency applied by the study investigators, intervention(s), comparison(s), co-interventions, red blood cell transfusion trigger applied, AE outcome(s) of interest (+ method and timing of outcome assessment), raw event data for each of the reported AEs. If a study presented its data as both composite measures (e.g. ‘cardiovascular complications’) and separate individual AEs (e.g. ‘atrial fibrillation’ and ‘cardiac tamponade’), only the individual data were extracted. If a preregistered trial protocol was available, the trial registration webpage was scanned for additional information (e.g. End of Study Reports). In case of missing or insufficiently ambiguous data or composite measure data, study authors were contacted via email at least twice regarding additional or disaggregated data. If a study reported a general statement indicating the absence of an AE (e.g. “no serious AEs were identified in any group,” without defining seriousness), this was considered insufficiently ambiguous and authors were contacted to confirm the absence of events (true “zero events”), and to clarify the individual AEs that were studied/recorded. If they did not reply or were not able to specify the events, the study was excluded.

### Quality appraisal: risk of bias and GRADE assessment

Risk of bias and GRADE assessments were performed by two reviewers independently (JL and HVR). Discrepancies were resolved through discussion. The GRADE assessment was verified by a third external methodological expert (GB).

Risk of bias at the study level was assessed using the Cochrane risk-of-bias tool for randomized trials (15) or the GRADE key criteria for observational study limitations (‘Inappropriate eligibility criteria’, ‘Inappropriate methods for exposure variables’, ‘Not controlled for confounding’, ‘Incomplete or adequate follow-up’, ‘Other limitations’) for experimental and observational studies, respectively, except for the items regarding ‘Inadequate measurement of the outcomes’ and ‘Inadequate selection of the reported results’. For these items, domains 4 and 5 of the revised Cochrane risk-of-bias tool (RoB 2) (16) were used, as they cover the important aspects for assessing bias in AE studies more thoroughly. The signaling questions were answered in the Excel RoB 2 implementation tool (17). Whenever the tool’s algorithm proposed ‘low’, the reviewers judged the study at low risk of bias. If the algorithm proposed ‘some concerns’ or ‘high’, they judged it at high risk of bias.

Next, the GRADE approach (Grading of Recommendations, Assessment, Development and Evaluation) was used to assess the overall certainty of the evidence. The certainty of the evidence for each outcome (category) was graded as ‘high’, ‘moderate’, ‘low’ or ‘very low’. Experimental and observational studies receive an initial grade of ‘high’ and ‘low’, respectively. Subsequently, these initial levels may be downgraded (based on risk of bias, imprecision, inconsistency, indirectness, and selective non-reporting bias) or upgraded (based on large effect, dose-response gradient, plausible confounding)(18).

### Data synthesis

If at least 2 studies provided data on the same outcome within the same treatment comparison, and no large heterogeneity in outcome definitions and measurements was suspected, random effects meta-analysis was performed using the Review Manager 5.3 software (19). Heterogeneity was assessed through visual inspection of the forest plot and by using the X^2^-test and the I^2^ statistic. To investigate if AEs varied by administration route (i.e. oral versus IV iron), subgroup analyses were performed. Predefined sensitivity analyses were done to explore the influence of: (1) different definitions of anaemia, and (2) different risk of bias judgements concerning ‘Inadequate measurement of the outcomes’ and ‘Inadequate selection of the reported results’ (see Quality appraisal). The statistical significance threshold was set at 5%.

In case a meta-analysis was not possible (i.e. data were only reported by one study) or warranted (i.e. heterogeneity in outcome definitions was observed or suspected), outcome data were presented in a single forest plot per AE category (without calculating a total effect size) as a visual aid for result interpretation. Statistical synthesis of these results was deemed inappropriate and no statements about consistency of effects across studies or outcomes were made to avoid unintentional vote counting (20).

To formulate the overall review findings as clearly and simply as possible, informative statements were developed in accordance with the set of statements provided by the GRADE Working Group and the Cochrane GRADEing Methods Group (21, 22).

## Results

### Search results

Figure 1 shows the detailed PRISMA study selection flow diagram. The primary search yielded 10142 database records and 3163 trial registry records. After duplicate removal, the title and abstract of the remaining 8221 records were screened. After full text screening and resolving disagreements, 44 peer-reviewed publications and 3 clinical trial study reports reporting on 42 unique studies were included. A list of studies excluded during full text screening and the reasons for exclusion is provided in Supplementary Table 2 in Additional file 2. Moreover, we identified 21 unique relevant registered ongoing trials (24 trial registrations) and 6 unique prematurely ended or terminated trials (7 registrations), as well as 2 relevant protocol papers (Supplementary Table 3 in Additional file 2).

**Figure 1.**
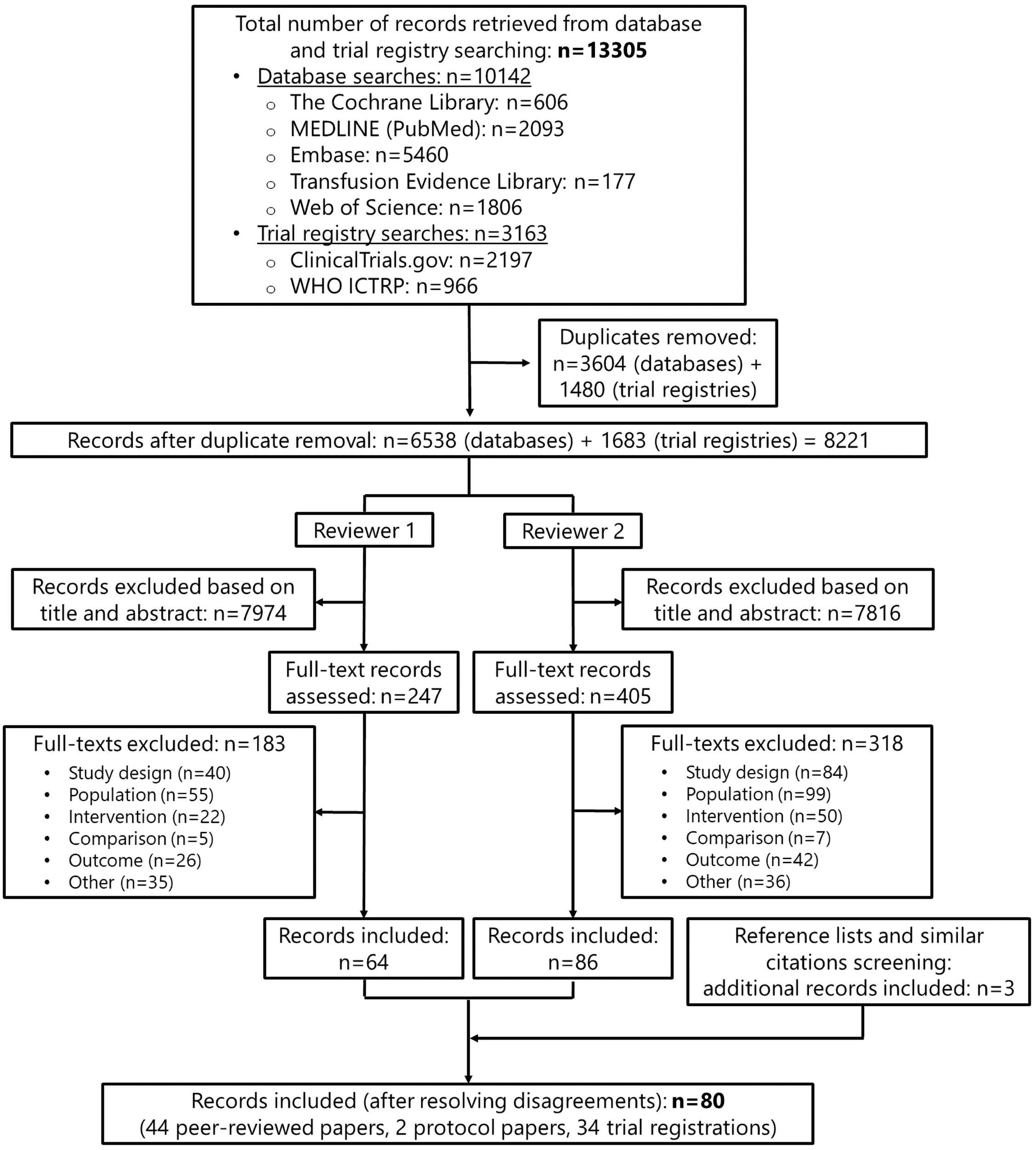
PRISMA study selection flow diagram. WHO: World Health Organization; ICTRP: International Clinical Trials Registry Platform

### Study characteristics

Of the 42 included studies involving a total of 6062 patients, 26 were RCTs (23-50) and 16 were cohort studies (51-66). Twenty-eight studies were conducted in Europe (Denmark, France, Germany, Greece, Italy, Spain, Sweden, The Netherlands, UK), whereas the other 14 were conducted in Australia (n=2) (30, 35), USA & Canada (n=6) (26, 45, 47, 48, 52, 53) and Asia (China & South Korea (n=6)) (24, 36, 39, 50, 66).

Only 15 studies (27, 30, 32, 33, 41, 44, 46, 48, 50, 51, 54, 57, 59, 61-63) applied the WHO definition of anaemia in determining patient study eligibility. Information on the patients’ iron-deficiency status was available from just over half the studies (55%) (23, 24, 26, 28-31, 34-36, 38-41, 44, 48, 50, 54, 55, 58, 59, 61, 63). In 8 studies, the entire study population suffered from iron-deficiency anaemia (30, 35, 36, 38, 39, 58, 61, 63). Patients were scheduled for the following types of elective surgery: colorectal cancer (n=12 studies)(25, 27, 31-34, 37, 42, 43, 56, 58, 61, 64, 65), orthopaedic (n=10)(23, 24, 26, 35, 40, 46, 52, 53, 60, 62), cardiac (n=7)(41, 48-51, 55, 57), gynaecologic (n=6)(28, 29, 36, 38, 39, 66), major head and neck oncologic (n=1)(45), abdominal (n=2)(30, 44), spinal (n=1)(47), vascular (n=1)(59)cardiac/thoracic/orthopaedic/gynaecologic/obstetric (n=1)(54) or visceral/vascular/gynaecologic/maxillofacial/cardiac/orthopedic/urologic/other major (n=1)(63) surgery.

Studies provided data on 6 different treatment comparisons. The majority of the studies (n=22; 52%) reported on the comparison of the combined therapy of ESAs and iron versus control (placebo and/or iron, no treatment) (24-26, 28, 29, 31, 34, 37, 38, 40, 42, 43, 45-53, 60, 66). Seven studies compared IV to oral iron monotherapy (27, 32, 33, 35, 36, 40, 41, 61). Data on the comparison of IV iron monotherapy versus control (usual care or no iron therapy) were provided by 12 studies (30, 44, 54-59, 63-66). Two RCTs compared the combined therapy of ESA+IV iron to that of ESA+oral iron (23, 40), whereas one cohort compared different dosing regimens of ESA during the combined therapy with IV iron (62). Finally, one RCT compared IV ferric carboxymaltose to IV iron sucrose monotherapy (39).

Most commonly, AE data concerned mortality/survival (n=24 studies) and the occurrence of thromboembolic (n=22), infectious (n=20), cardiovascular (n=19), gastrointestinal (n=14) and anaemia-associated ischemic AEs (n=10). Autonomic (n=8), bronchopulmonary (n=8), bleeding (n=7), renal (n=7), neuro-psychosomatic (n=6), mucocutaneous (n=6), neurological (n=5), AEs related to wound healing (n=4) and other types of AEs (n=24) were reported by less than 20% of the studies. Supplementary Table 4 in Additional file 2 provides an overview of the AE outcomes for which data were obtained and additionally depicts which RCTs provided data on these outcomes within the 6 treatment comparisons. An similar overview depicting which cohort studies provided data for each of the outcomes within the 6 treatment comparisons is provided in Supplementary Table 5 in Additional file 2. These tables clearly indicate that for any given AE outcome and treatment comparison, data were provided by a very limited number of studies, with the exception of deep venous thrombosis and mortality. In addition, some studies provided data on a multitude of outcomes, whereas others only studied (or at least reported on) a single outcome.

Detailed information on the characteristics of the included studies, including the specific AE outcomes and method and timing of their measurement, is presented in Supplementary Table 6 in Additional file 2.

### Risk of bias and certainty of evidence

Risk of bias in the individual studies is presented in Figures 2 and 3 (details in Supplementary Table 7 in Additional file 2). All but two RCTs (27, 49) and two cohort studies (55, 65) were found to be at high risk of bias in outcome measurement. Often, there was insufficient information available to determine if the measurement of the outcome was a pre-defined part of the study protocol or if data were added post-hoc (e.g. by analysing data on AE that must be reported to regulatory agencies such as the FDA). In addition, many studies failed to clearly mention the methods used for (some of) the actual outcome measurement, if these methods used were similar across all study participants, and if the outcome assessors were blinded. Only four RCTs (24, 27, 30, 47) and two cohort studies (55, 65), were judged to be at low risk of bias in the selection of reported results. Often, trial protocols or published papers did not describe how (expected or unexpected) adverse outcomes were collected and analysed. Therefore, the reported AE data may have been selected based on the finding being noteworthy.

**Figure 2.**
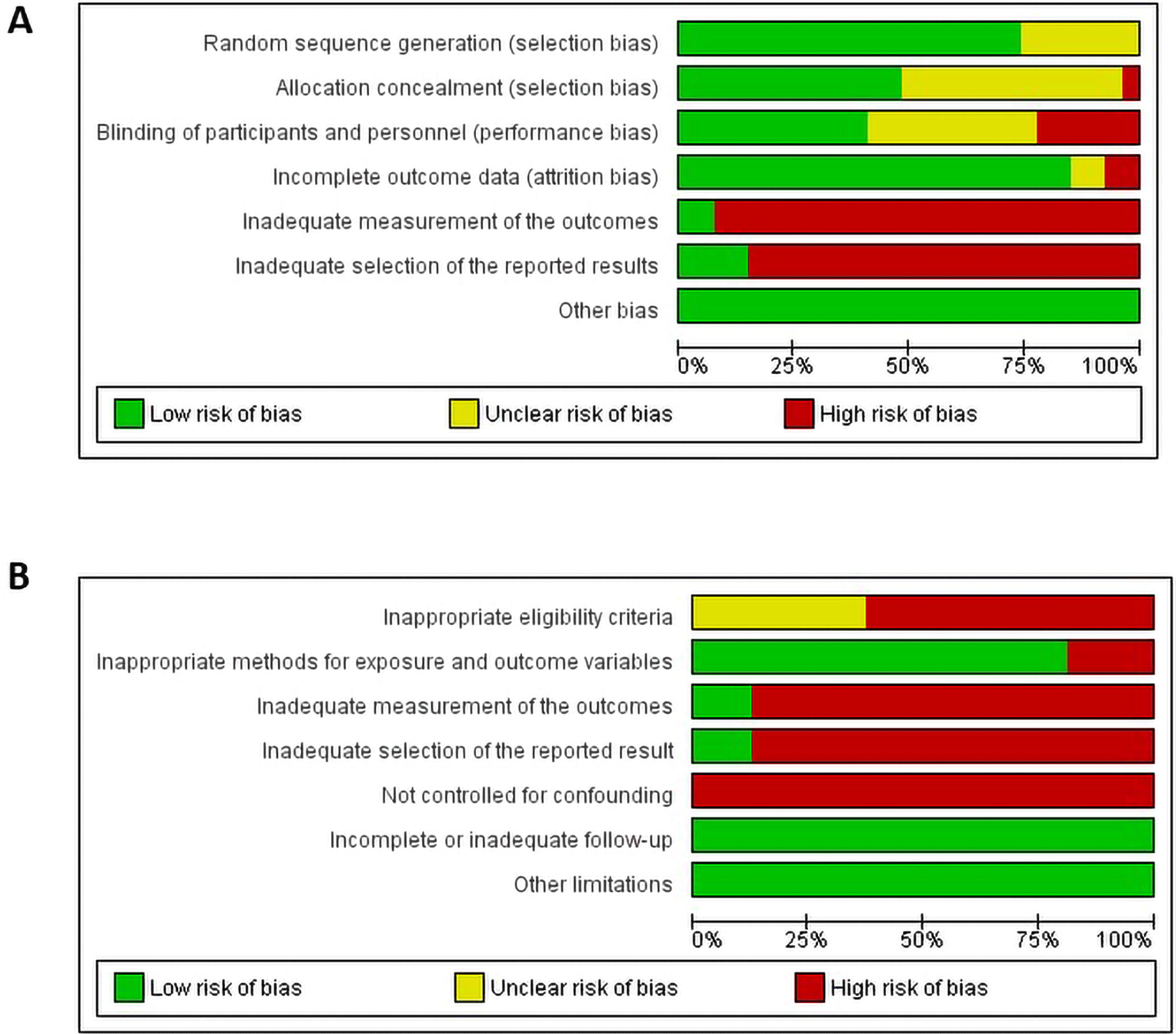
Risk of bias graph. Review authors’ judgments about each risk of bias item presented as percentage across all included (A) RCTs and (B) cohort studies.

**Figure 3.**
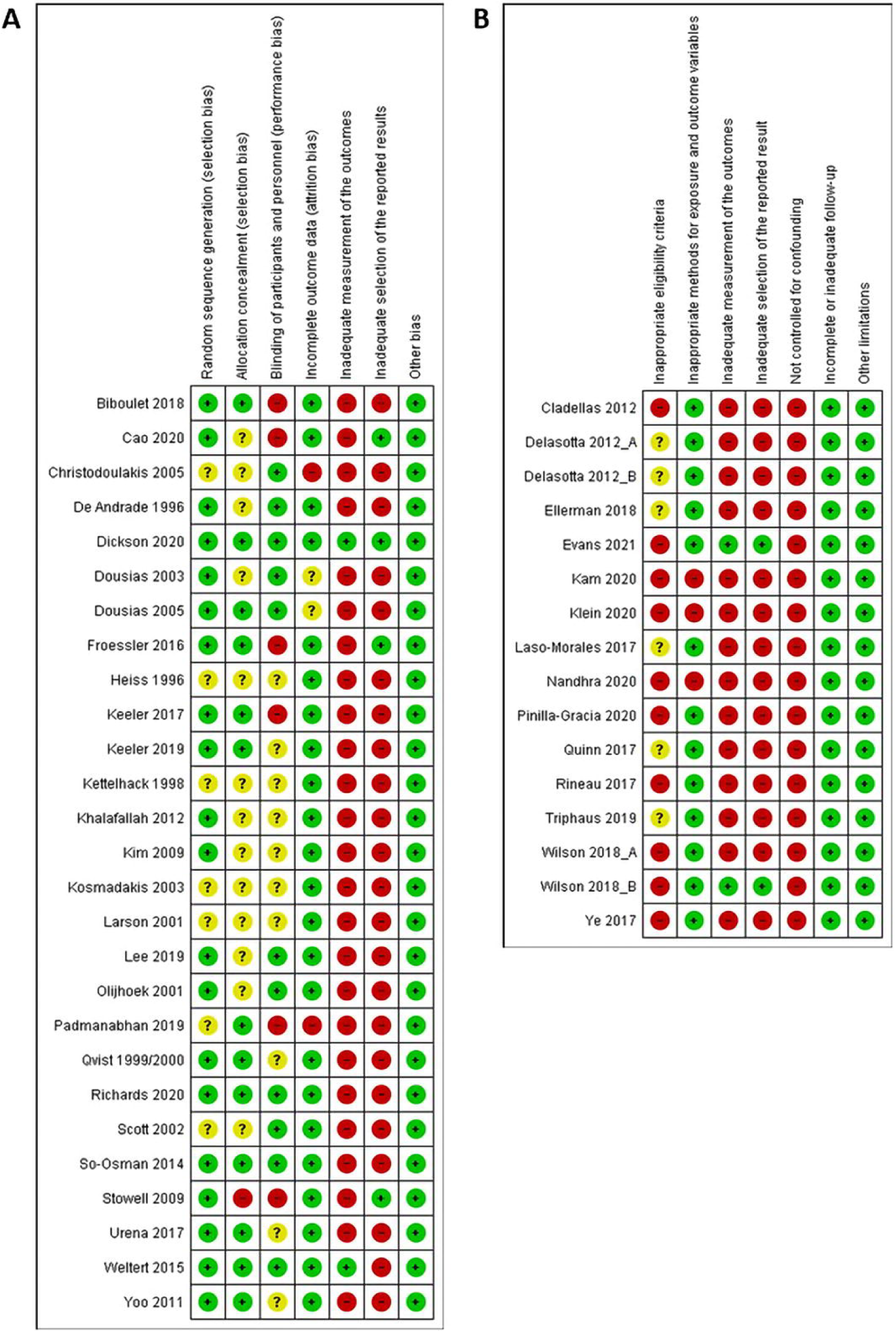
Risk of bias summary. Review authors’ judgments about each risk of bias item for each included (A) RCT and (B) cohort study 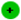. Low risk of bias, 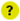 Unclear risk of bias, 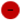 High risk of bias

Furthermore, 23% of the RCTs were at high risk of performance bias due to lack of blinding of study participants or personnel, whereas only 2 studies showed high risk of attrition bias. Random sequence generation, allocation concealment and blinding of study participants or personnel were unclear in 27%, 50% and 38% of the RCTs, respectively. Furthermore, none of the cohort studies adequately controlled for confounding, and 10 of them applied inappropriate eligibility, whereas the other 6 did not report sufficient information to make an appropriate judgement.

Based on the risk of bias assessment of the individual studies, the overall certainty of the entire body of evidence was downgraded by one level for each outcome (category) of treatment comparisons 2 to 6 (see Synthesis of results). Within the first treatment comparison (IV vs oral iron monotherapy), most outcomes were also downgraded by one level based on risk of bias assessment, except for ‘survival’. For this outcome, the reviewers found no reason to downgrade because of the well-designed and - executed RCT of Dickson *et al*. (27). Next, for all outcomes within the 6 treatment comparisons, the evidence was further downgraded by 2 levels due to imprecision because of the very low number of AEs, limited sample sizes and the wide 95% CIs around the effect estimates.

As a result, a very low certainty of evidence was assigned to all AE outcomes within treatment comparisons 2 to 6, indicating that we are uncertain about these effect estimates. Within the first treatment comparison, the overall certainty for ‘survival’ was judged to be low, in contrast to the other outcomes that were assigned a very low certainty of evidence.

### Synthesis of results

Supplementary Table 8 in Additional file 2 contains all AE outcome data extracted from the 42 included studies, classified into the 15 AE categories within the 6 treatment comparisons. Meta-analysis could only be performed for a total of 26 outcomes across 3 treatment comparisons: dyspepsia, postoperative infection and mortality (Comparison 1: IV versus oral iron monotherapy), nausea, headache, ileus, dyspnoea, surgical/superficial wound infection, deep venous thrombosis, mortality, hospital readmission (Comparison 2: IV iron vs usual care/no iron) and 15 outcomes within Comparison 4 on ESA+iron versus control (details listed below). For the vast majority of outcomes, meta-analysis was not warranted or feasible.

Causality assessment (67) was not feasible, as insufficient information was available to judge the relation between the intervention and the AEs reported, i.e. if AEs were related/probably related/possibly related/unlikely to be related/conditionally related to the studied intervention.

#### Comparison 1: IV vs oral iron monotherapy (7 studies, 312 participants)

Neither the meta-analyses on dyspepsia, postoperative infection and mortality data, nor the other separate analyses on any of the other available AE data (36 outcomes across 13 AE categories, detailed in Supplementary Table 8 in Additional file 2) revealed a statistically significant difference in the occurrence when administering IV compared to oral iron monotherapy.

#### Comparison 2: IV iron vs usual care/no iron (12 studies, 2298 participants)

One RCT revealed that IV iron monotherapy was associated with a statistically significantly lower readmission rate for wound infection between discharge and 8 weeks, compared to usual care/no iron therapy (44). One cohort study showed that IV iron monotherapy was associated with a statistically significantly lower occurrence of dyspepsia, as well as a statistically significantly lower 1-year infection rate and prevalence of infectious-related codes during hospital stay, compared to usual care/no iron therapy (54). Another cohort study showed that IV iron monotherapy was associated with statistically significantly lower 4-year and 5-year disease-free survival rates, as well as 4-year and 5-year overall survival rates, compared to usual care/no iron therapy (65).

Data on the following 8 AE outcomes were meta-analysed: nausea, headache, ileus, dyspnoea, surgical/superficial wound infection, deep venous thrombosis, mortality and hospital readmission. The meta-analysis on the dyspnoea data, provided by two cohort studies (54, 63), showed that IV iron monotherapy was associated with statistically significantly lower occurrence of dyspnoea, compared to usual care/no iron therapy. None of the other meta-analyses, nor the other separate analyses on any of the other available AE data (69 outcomes across 15 categories) revealed a statistically significant difference in the occurrence when administering IV iron monotherapy compared to usual care/no iron therapy.

#### Comparison 3: IV ferric carboxymaltose vs IV iron sucrose monotherapy (1 study, 101 participants)

One RCT could not reveal a statistically significant difference in mortality or the occurrence of anaphylactic reactions when administering IV ferric carboxymaltose monotherapy compared to IV iron sucrose monotherapy (39). Data on AEs in the other 13 AE categories were not available.

#### Comparison 4: ESA+iron vs control (placebo and/or iron, no treatment) (22 studies, 3152 participants)

One cohort study revealed that the combined therapy of ESA and IV iron was associated with a statistically significantly lower occurrence of acute renal failure, severe infection (composite measure of sepsis, pneumonia or mediastinitis) and major adverse cardiovascular events (composite measure), compared to no treatment (51). In addition, one RCT showed that the combined therapy of ESA and IV iron was associated with a statistically significantly lower postoperative complication rate (composite measure of anastomotic leak abscess/fistula formation, haemorrhage, wound infection, pulmonary complications, complications from blood transfusions), compared to the combination of placebo and IV iron therapy (37).

One RCT showed that the combined therapy of ESA and oral iron was associated with a statistically significant higher occurrence of back pain, compared to the combination of standard of care and oral iron therapy (47).

Data on the following 15 AE outcomes were meta-analysed: 3 gastrointestinal AEs (nausea, vomiting and obstipation), 2 infectious AEs (surgical/superficial wound infection and urinary tract infection), 4 cardiovascular AEs (atrial fibrillation, heart failure, hypertension and cardiac tamponade), 3 anaemia-associated ischemic AEs (acute kidney injury, myocardial infarction and stroke; see Figure 4), 2 thromboembolic AEs (pulmonary embolism and deep venous thrombosis; see Figure 5) and mortality. Interestingly, the meta-analysis on the mortality data provided by two small cohort studies (51, 60) showed that the combined therapy of ESA and IV iron was associated with a statistically significantly lower mortality rate compared to control (Risk Ratio (RR): 0.39, 95%CI [0.17;0.91], p=0.03), while this could not be demonstrated by the meta-analysis of the mortality data provided by two RCTs (37, 40) (RR: 0.48, 95%CI [0.21;1.09], p=0.08). When combining all RCTs (25, 31, 34, 37, 40, 45, 49, 53) comparing the combined therapy of ESA and iron (both oral and IV) to control in one meta-analysis, a statistically significant difference in mortality could not be demonstrated.

**Figure 4.**
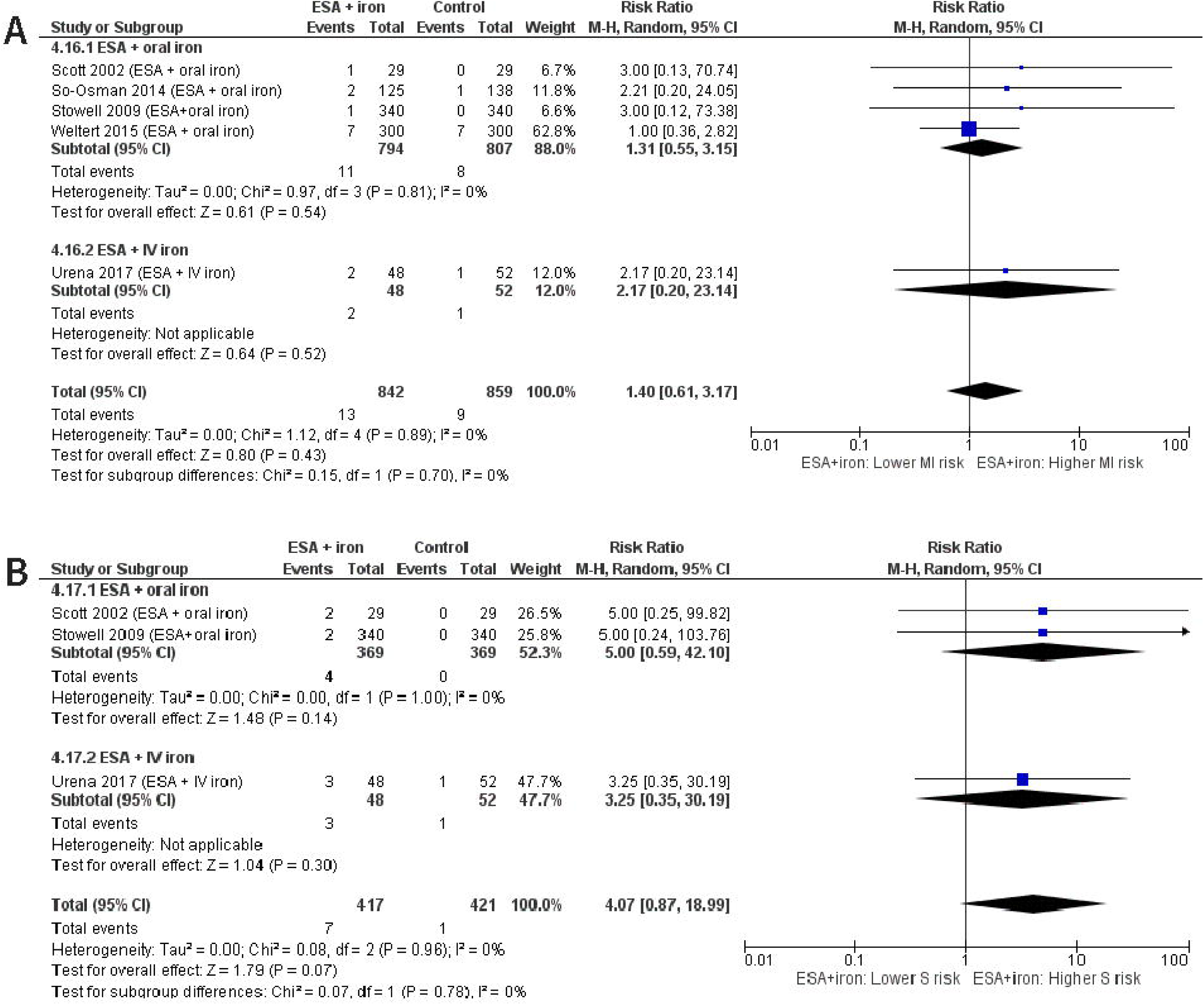
Meta-analyses on myocardial infarction (MI) and stroke (S) during ESA + iron treatment. Meta-analysis of data from RCTs on the occurrence of (A) myocardial infarction and (B) stroke in preoperatively anaemic patients scheduled for elective surgery undergoing the combined treatment therapy of ESA and iron, compared to a control (placebo and/or iron) or no treatment.

**Figure 5.**
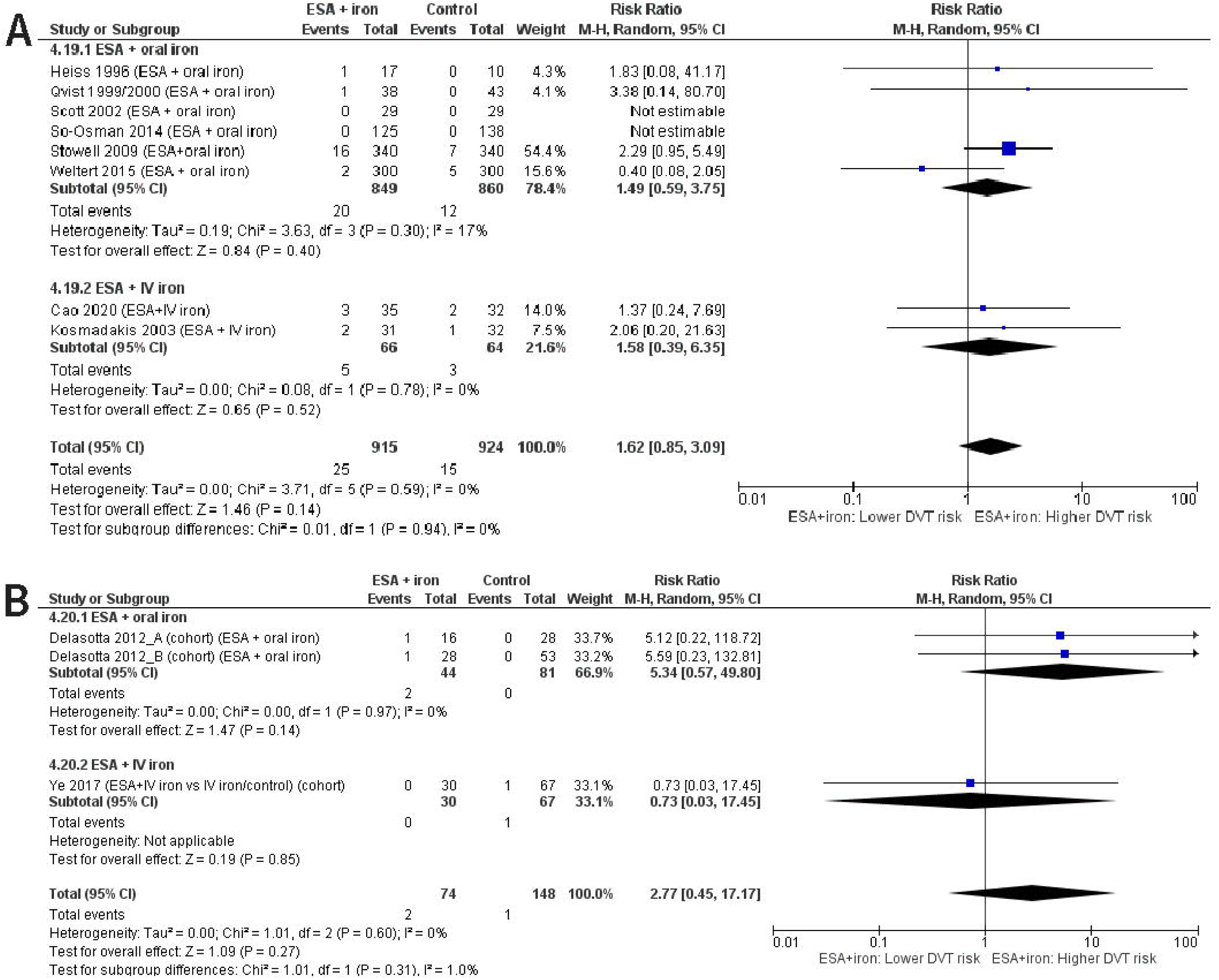
Meta-analyses on deep venous thrombosis (DVT) during ESA + iron treatment. Meta-analysis of data from RCTs on the occurrence of deep venous thrombosis in (A) RCTs and (B) cohort studies of preoperatively anaemic patients scheduled for elective surgery undergoing the combined treatment therapy of ESA and iron, compared to a control (placebo and/or iron) or no treatment.

None of the other meta-analyses, nor the other separate analyses on any of the other available AE data (90 outcomes across 14 categories) revealed a statistically significant difference in the occurrence when administering ESA+iron compared to control.

The findings related to atrial fibrillation and myocardial infarction were not influenced by excluding studies that were not in line with the WHO definition of anaemia.

To investigate if AEs differentially occurred during treatment with ESA+IV iron and ESA+oral iron, subgroups were created if possible. Subgroup analyses were performed for 3 cardiovascular AEs (atrial fibrillation, heart failure, hypertension), 2 anaemia-associated ischemic AEs (myocardial infarction, stroke), deep venous thrombosis and mortality. The only statistically significant difference was detected for the outcome of mortality reported by the RCTs, but there was considerable heterogeneity (p-value X^2^-test = 0.03, I^2^=78.8%). Two small RCTs (117 participants) that used ESA+IV iron as a treatment (37, 40) reported a lower mortality in the intervention group compared to the control group (Risk Ratio (RR): 0.48, 95%CI [0.21;1.09], p=0.08), whereas the 6 RCTs (1054 participants) that used ESA+oral iron (25, 31, 34, 40, 45, 49) demonstrated non-significantly higher mortality rates in the intervention group (RR: 1.75, 95%CI [0.76;4.02], p=0.18). This between-subgroup heterogeneity may result from the difference in sample size of and the follow-up period applied by the studies.

#### Comparison 5: ESA+IV iron vs ESA+oral iron (2 studies, 158 participants)

One RCT revealed that the combined therapy of ESA and IV iron was associated with statistically significantly less digestive complications, compared to the combined therapy of ESA and oral iron (23). Statistically significant differences in the occurrence of preoperative prostatitis, cardiac or respiratory failure, deep venous thrombosis or preoperative femoral vein thrombosis, thrombotic and/or vascular events, study mortality or mortality during hospitalization, when using the combined therapy of ESA and IV iron compared to ESA and oral iron, could not be demonstrated (23, 40). For 10 AE categories, data were not available.

#### Comparison 6: ESA+ IV iron vs ESA+IV iron (different ESA dosing regimens) (1 study, 127 participants)

The only included cohort study (62) could not reveal a statistically significant increase in the occurrence of intestinal obstruction, 3 bronchopulmonary AEs (pulmonary edema, the need for re-intubation, the need for prolonged ventilation), 3 infectious AEs (pneumonia, wound infection, urinary tract infection), 2 bleeding AEs (hemorrhagic shock, postoperative hematoma), 4 cardiovascular AEs (atrial fibrillation, acute coronary syndrome, arrhythmia, cardiac arrest), 3 anaemia-associated ischemic AEs (acute limb ischemia, stroke, acute kidney injury), 2 thromboembolic AEs (deep venous thrombosis, pulmonary embolism), epileptic seizures or allergy when comparing different ESA dosing regimens during the combined treatment of ESA and IV iron. Data on AEs in the other 7 AE categories were not available.

## Discussion

This systematic review identified 42 studies containing data on AEs occurring during/after the treatment with iron and/or ESAs, classified into 15 AE categories within 6 treatment comparisons.

Regarding iron monotherapy:

- IV iron monotherapy may not be associated with changes in survival compared to oral iron monotherapy (low-certainty evidence).
- We are uncertain (all very low-certainty evidence) whether:
  ∘ IV iron monotherapy is associated with an increased occurrence of AEs compared to oral iron monotherapy, or compared to usual care/no iron therapy.
  ∘ IV ferric carboxymaltose monotherapy is associated with an increased occurrence of AEs compared to IV iron sucrose monotherapy.

Regarding the combination treatment with ESAs and iron, we are uncertain (all very-low certainty evidence) whether:

- The combined administration of ESAs and iron is associated with an increased occurrence of AEs compared to placebo and/or iron or no treatment.
- The combined administration of ESAs and IV iron is associated with an increased occurrence of AEs compared to treatment with ESAs and oral iron.
- The use of different ESA dosing regimens during the combination treatment with ESAs and IV iron is associated with differences in the occurrence of AEs.

The majority of the overall certainty of the evidence was judged to be very low due to three main reasons. Firstly, the incidence of many of the studied AEs is low, thereby hindering the precision of the results. For example, incidence rates of anaphylactic reactions after IV iron administration are reported to lie around 0.1-1% (68, 69). Hence, several hundreds of patients would have been required to detect a difference between IV ferric carboxymaltose and IV iron sucrose patients in the study by Lee (39). The issue of low incidence is also illustrated in Figure 5 on deep vein thrombosis. In 4 of the 10 studies (31, 42, 52, 53), deep venous thrombosis was detected in just one treatment participant. Two other studies (45, 46) did not detect deep venous thrombosis in any of the treatment or control group participants. Although this highlights the successful use of thromboprophylaxis, the small number of events decreases the precision of the estimate and therefore renders the results fragile (70). A closer look at the entire dataset of treatment comparison 4 reveals that 17 of the 22 studies did not observe any event in the treatment group for at least one of their studied AEs (45 in total). The issue is further complicated by the fact that in 20% and 36% of the included peer-reviewed publications on iron monotherapy and the combination treatment with ESAs and iron, respectively, the study authors failed to properly report on study duration and patient follow-up time. Only 36% of the studies on the combination treatment with ESAs and iron certainly employed a follow-up period of at least 30 postoperative days. As a result, some longer-term AEs of these therapies (e.g. thromboembolic, cardiac AEs) may only have become apparent after follow-up of these patients had ended.

A second independent important reason for the very low certainty of the evidence is the lack of systematic surveillance of pre-defined AE outcomes: just 5 studies (27, 47, 49, 55, 65) explicitly indicated that patients were systematically assessed for AEs or that AE recording was part of the study protocol, whereas the others probably used spontaneous report monitoring and/or reporting. Moreover, there is heterogeneity in definitions and data collection methods used, as well as a lack of reporting on the exact methods used to ascertain the events.

Thirdly, selective non-reporting bias is likely to have occurred, for example due to conflicts of interest: entire study reports may be unpublished due to the unexpected findings of harms, or alternatively, particular study results may be selectively unavailable (e.g. because the magnitude, direction or p-value were considered unfavourable by the investigators). This may put the results of this review at risk, as these missing results may differ from the available results. Therefore, the results of these analyses should be interpreted with caution. Nevertheless, maximal effort has been put in to ensure minimal bias occurring at the review level by including both published and unpublished study data retrieved from an extensive array of databases and trial registries relevant to the topic of PBM, and by contacting study authors to request missing information.

This systematic review has several other strengths. By adopting comprehensive selection criteria, the review has captured any possible AE that may (have) occur(red) during or after administration of iron and/or ESA therapy in preoperatively anaemic elective surgery patients. In contrast to most systematic reviews of RCTs only, observational studies were eligible as well. No effort was spared in contacting study authors to obtain additional data or clarification. Special attention was paid to inadequate monitoring and incomplete reporting, both pivotal issues in assessing risk of bias for AE data. Finally, the GRADE assessment was checked by a third independent methodological expert.

To the best of our knowledge, this is the first systematic review to summarize the available direct evidence on potential AEs of iron and/or ESA therapy in preoperatively anaemic patients scheduled for any type of elective surgery.

A previous systematic review on the use of ESAs (whether or not augmented with preoperative autologous blood donation) in anaemic patients undergoing elective hip, knee and spine orthopaedic surgery demonstrated that recombinant human erythropoietin was associated with an increased risk of deep vein thrombosis (Peto Odds Ratio: 1.66, 95%CI [1.10;2.48]), but was inconclusive regarding the risk of mortality, myocardial infarction and cerebrovascular accidents (71). In contrast, in a more recent systematic review that only included RCTs that investigated preoperative erythropoietin administration in adult surgical patients and contained data on allogeneic transfusions as their primary outcomes, an association between preoperative erythropoietin and an increased risk of thromboembolic events could not be demonstrated (RR: 1.02, 95%CI [0.78;1.33], p=0.68) (72). In a third systematic review of RCTs investigating the combination therapy of ESAs and iron compared to iron monotherapy in adult surgical patients, an association could not be demonstrated between the combination therapy and an increased risk of deep vein thrombosis (RR: 1.48, 95%CI [0.95;2.31], p=0.09), pulmonary embolism, mortality, stroke, myocardial infarction or renal dysfunction (73). A recent Cochrane systematic review of RCTs comparing preoperative recombinant human erythropoietin plus iron therapy to control (placebo, no treatment, or standard of care with or without iron) in preoperatively anaemic adults undergoing non-cardiac surgery found moderate-certainty evidence indicating little or no difference in the risk of mortality within 30 days of surgery or of experiencing any adverse event (including local rash, fever, constipation, or transient hypertension) (74). Other prior systematic reviews on ESA safety have focused on other patient populations such as rheumatoid arthritis (75), chronic heart failure (76, 77), predialysis (78), chronic kidney disease (79) and critically-ill (80) patients. Similar to our review, these reviews concluded that the question of whether ESAs affect the risk of AEs in these patient populations remains unanswered. The very narrow scope of the population of interest (in our case preoperatively anaemic elective surgery patients) has certainly played a role in this, since it resulted in a limited number of included studies covering a wide range of AEs.

A recent systematic review has shed light on the safety of perioperative iron administration. Gómez-Ramírez *et al*. synthetized the evidence on the efficacy and safety of short-term perioperative intravenous iron, with or without erythropoietin, in both elective and non-elective major orthopaedic surgery (81). In 10 of the 14 studies identified, only 25 adverse drug effects were reported in patients treated with intravenous iron, which consisted mainly of gastrointestinal symptoms (nausea, vomiting, diarrhoea and/or constipation) and hypotension. There was no difference in the incidence of clinically relevant adverse drug effects in patients receiving iron, with or without EPO, compared to those assigned to control (1.13% vs. 0.85%; RR: 1.34, 95% CI[0.63;2.86], p=0.56). When focusing on elective surgery patients, one RCT and 3 observational studies were identified, of which 2 observational studies reported on 30-day mortality and infection rates. No statistically significant differences in postoperative infection rates were found. Mortality rates were 0 in both studies, hindering further statistical analysis. Similar to our review, a recent Cochrane systematic review of RCTs evaluating the effect of preoperative iron therapy (compared to placebo, no treatment, standard of care or another form of iron therapy) in anaemic patients undergoing surgery concluded that the effects on short-term mortality or postoperative morbidity (including infection and adverse events within 30 days) remain uncertain, and that the inclusion of new research in the future is therefore very likely to change the results (82).

Other previous systematic reviews investigating the safety and tolerability of iron therapy (83-85) have generally employed broader selection criteria at the population level, in combination with narrow criteria at the intervention (e.g. comparing IV iron supplements to each other) and/or outcome level (i.e. limiting the number of AEs of interest). For example, a systematic review and meta-analysis by Tolkien revealed that oral ferrous sulfate therapy in patients with iron-deficiency anaemia is associated with significantly more gastrointestinal-specific side-effects, compared to IV iron or placebo (86). By including patients with iron-deficiency anaemia of any cause (e.g. chronic kidney disease, pregnancy, blood donation), the reviewers were able to meta-analyse data from 43 RCTs comprising 6831 adults.

Therefore, future systematic review teams may benefit from employing broad selection criteria at the population level in combination with narrow intervention criteria. Expanding the population scope will facilitate the retrieval of AE data of regulated iron and ESA products published in regulatory agency databases (e.g. Food and Drug Administration AE Reporting System). If sufficient numbers of studies/data sources are included and significant heterogeneity is detected, the reviewers should consider performing subgroup analyses at the population level.

However, in order to be able to perform systematic reviews and meta-analyses that provide higher-certainty evidence that can influence decision-making in healthcare, further transparent post-marketing safety surveillance of iron and ESA products is warranted. In addition, authors of experimental studies on iron and/or ESA therapy should spend time thinking about expected AEs, how to measure and analyse them, and document this in an *a priori* (ideally published) protocol. They should keep track of expected and unexpected AEs and may want to consider providing all AE data online (e.g. in an End of Trial report, open-access) if reporting in a peer-reviewed publication is impeded by word limitations. Finally, publishers and funders should stress the importance of the collection, documenting and reporting of AEs and adopt rigorous conflict of interest policies.

## Conclusions

In conclusion, it remains unclear if ESA and/or iron therapy is associated with the occurrence of AEs in preoperatively anaemic elective surgery patients. This uncertainty results from both low quantity and quality of AE data due to limitations in study design, data collection and reporting.

## Data Availability

All data generated or analysed during this study are included in this published article and its additional file.

## List of abbreviations

AE: Adverse event
CI: Confidence interval
ESA: Erythropoiesis-stimulating agent
GRADE: Grading of Recommendations, Assessment, Development and Evaluation
ICC-PBM: International Consensus Conference – Patient Blood Management
IV: Intravenous
IQR: Interquartile range
PBM: Patient blood management
PICO: Population Intervention Comparison Outcome RCT Randomized controlled trial
RoB: Risk of Bias
RR: Risk Ratio
WHO: World Health Organization

## Declarations

### Ethics approval and consent to participate

Not applicable.

### Consent for publication

Not applicable.

### Availability of data and materials

All data generated or analysed during this study are included in this published article and its additional files.

### Competing interests

#### Financial competing interests directly related to this review

PMM received personal fees from Ethos Srl (Advisory Board on PBM) and SUMMEET Srl (Speaker in a meeting on PBM). PM received grants from Vifor Pharma, SerumWerke Bernburg, csl Behring, Fresenius Medical, and B.Baun. PM received personal fees from Vifor Pharma and Pharmacosmos. HV, JL, BA, EDB, GB, JG, VC, YO and PV declared not having any relevant direct financial competing interest.

#### Financial competing interests not directly related to this review

HV, JL, BA, EDB, VC and PV are employees of Belgian Red Cross-Flanders, which is responsible for supplying adequate quantities of safe blood products to hospitals in Flanders and Brussels on a continuous basis and is being paid for this activity by the Ministry of Social Affairs. Belgian Red Cross-Flanders received a grant from the European Blood Alliance to conduct this review. PMM received personal fees from Editree Srl (creation of training course on porphyria), Eleuthera Srl (AHP Advisory Board), Collage SpA (speaker in a meeting on Erythrocytosis), IQVIA (Advisory Board on porphyria), Alnylam (Advisory Board on porphyria). GB, JG, PM, YO declared not having any other financial competing interest.

### Authors’ contributions

JL: Investigation, Formal analysis, Writing – Original draft, Visualization. HVR: Conceptualization, Investigation, Formal analysis, Writing – Original draft, Visualization, Supervision, Project administration, Funding acquisition. BA: Validation, Writing – Review & Editing. GB: Validation, Writing – Review & Editing. JG: Validation, Writing – Review & Editing. PMM: Validation, Writing – Review & Editing. PM: Validation, Writing – Review & Editing. YO: Validation, Writing – Review & Editing. EDB: Conceptualization, Writing – Review & Editing, Supervision. VC: Conceptualization, Writing – Review & Editing. PV: Conceptualization, Resources, Writing – Review & Editing, Supervision.

## Acknowledgements

We are grateful to study authors Prof. Dr. Andrea Steinbicker, Dr. Emmanuel Rineau, Dr. Edel Quinn, Dr. Yunhwan Kim, Prof. Dr. Christoph Kettelhack, Prof. Dr. Niels Qvist, Prof. Dr. Cynthia So-Osman, Dr. Philip Ming Ho Kam, Dr. Andrew Klein, Mr. Sandip Nandhra, and Prof. Dr. Patrick Meybohm (also a co-author for this review), for providing additional data and information. We thank Dr. Daniela Junqueira (Cochrane Adverse Effects Methods Group) for the helpful methodological discussions and guidance, and Ehsan Sadeghian and Olga Krasnoukhova for helping us screen Persian and Russian papers, respectively.

## Additional files

**Additional file 1. Completed PRISMA harms checklist**.

**Additional file 2. Supplementary tables**.

