## Additional file 1 for "Adverse events of iron and/or erythropoiesis-stimulating agent therapy in preoperatively anaemic elective surgery patients: a systematic review"

### Additional file 1: Completed PRISMA harms checklist.

| Section /topic  (page no) | Item | PRISMA checklist item | | | PRISMA harms (minimum) | Recommendations for reporting harms in systematic reviews  (desirable) | Check if done |
| --- | --- | --- | --- | --- | --- | --- | --- |
| **Title** | | | | | | | |
| Title (3) | 1 | Identify the report as a systematic review, meta-analysis, or both. | | | Specifically mention “harms” or other related terms, or the harm of interest in the review. | — | X (see *Title,*  p 1) |
| **Abstract** | | | | | | | |
| Structured summary (4) | 2 | Provide a structured summary including, as applicable: background; objectives; data sources; study eligibility criteria, participants, and interventions; study appraisal and synthesis methods; results; limitations; conclusions and implications of key findings; systematic review registration number. | | | — | Abstracts should report any analysis of harms undertaken in the review, if harms are a primary or secondary outcome. | X (see *Abstract,* p 3) |
| **Introduction** | | | | | | | |
| Rationale (5) | 3 | Describe the rationale for the review in the context of what is already known. | | — | | It should clearly describe in introduction or in methods section which events are considered harms and provide a clear rationale for the specific harm(s), condition(s), and patient group(s) included in the review. | X (see *Introduction,*  p 4 and *Materials and Methods – Eligibility criteria,* p 5) |
| Objectives (5) | 4 | Provide an explicit statement of questions being addressed with reference to participants, interventions, comparisons, outcomes, and study design (PICOS). | | — | | PICOS format should be specified, although in systematic reviews of harms the selection criteria for P, C, and O may be very broad (same intervention may have been used for heterogeneous indications in a diverse range of patients) | X (see *Introduction,* p 4 and *Materials and Methods – Eligibility criteria,* p 5) |
| **Methods** | | | | | | | |
| Protocol and registration (6) | 5 | Indicate if a review protocol exists, if and where it can be accessed (eg, web address), and, if available, provide registration information including registration number. | | — | | No specific additional information is required for systematic reviews of harms. | X (see *Materials and Methods,* p 5) |
| Eligibility criteria (6) | 6 | Specify study characteristics (eg, PICOS, length of follow-up) and report characteristics (eg, years considered, language, publication status) used as criteria for eligibility, giving rationale. | | — | | Report how handled relevant studies (based on population and intervention) when the outcomes of interest were not reported.  Report choices for specific study designs and length of follow-up. | X (see  *Materials and Methods – Eligibility criteria,* p 5) |
| Information sources (7) | 7 | Describe all information sources (eg, databases with dates of coverage, contact with study authors to identify additional studies) in the search and date last searched. | | — | | Report if only searched for published data, or also sought data from unpublished sources, from authors, drug manufacturers and regulatory agencies. If includes unpublished data, provide the source and the process of obtaining it. | X (see  *Materials and Methods – Data sources and searches,*  p 8 and  *Materials and Methods – Study selection,* p 8) |
| Search (7) | 8 | Present full electronic search strategy for at least one database, including any limits used, such that it could be repeated. | | — | | If additional searches were used specifically to identify adverse events, authors should present the full search process so it can be replicated. | X (see *Table B*) |
| Study selection (8) | 9 | State the process for selecting studies (ie, screening, eligibility, included in systematic review, and, if applicable, included in the meta-analysis). | | — | | If only included studies reporting on adverse events of interest, defined if screening was based on adverse event reporting in title/abstract or full text. If no harms reported in the text, report if any attempt was made to retrieve relevant data from authors. | X (see  *Materials and Methods – Study selection,* p 8) |
| Data collection process (9) | 10 | Describe method of data extraction from reports (eg, piloted forms, independently, in duplicate) and any processes for obtaining and confirming data from investigators. | | — | | No specific additional information is required for systematic reviews of harms. | X (see  *Materials and Methods – Data extraction,* p 9) |
| Data items (9) | 11 | List and define all variables for which data were sought (eg, PICOS, funding sources) and any assumptions and simplifications made. | | — | | Report the definition of the harm and seriousness used by each included study (if applicable). Report if multiple events occurred in the same individuals, if this information is available. Consider if the harm may be related to factors associated with participants (eg, age, sex, use of medications) or provider (eg, years of practice, level of training). Specify if information was extracted and how it was used in subsequent results. Specify if extracted details regarding the specific methods used to capture harms (active/passive and timing of adverse event). | X (see  *Materials and Methods – Data extraction,* p 9 and  *Table D*) |
| Risk of bias in individual studies (10) | 12 | Describe methods used for assessing risk of bias of individual studies (including specification of whether this was done at the study or outcome level), and how this information is to be used in any data synthesis. | | — | | The risk of bias assessment should be considered separately for outcomes of benefit and harms. | X (see  *Materials and Methods – Quality appraisal,*  p 9-10) |
| Summary measures (11) | 13 | State the principal summary measures (eg, risk ratio, difference in means). | | — | | No specific additional information is required for systematic reviews of harms. | X (see  *Materials and Methods – Data extraction,* p 9) |
| Synthesis of results (11) | 14 | Describe the methods of handling data and combining results of studies, if done, including measures of consistency (eg, I^2^) for each meta-analysis. | | Specify how zero events were handled, if relevant. | |  | X (see  *Materials and Methods – Data synthesis,* p 10) |
| Risk of bias across studies (11) | 15 | Specify any assessment of risk of bias that may affect the cumulative evidence (eg, publication bias, selective reporting within studies). | | — | | Present the extent of missing information (studies without harms outcomes), any factors that may account for their absence, and whether these reasons may be related to the results. | X (see  *Materials and Methods – Quality appraisal,*  p 9-10) |
| Additional analyses (12) | 16 | Describe methods of additional analyses (eg, sensitivity or subgroup analyses, meta-regression), if done, indicating which were prespecified. | | — | | Sensitivity analyses may be affected by different definitions, grading, and attribution of adverse events, as adverse events are typically infrequent or reported using heterogeneous classifications. Report the number of participants and studies included in each subgroup. | X (see  *Materials and Methods – Data synthesis,* p 10) |
| **Results** | | | | | | | |
| Study selection (13) | 17 | Give numbers of studies screened, assessed for eligibility, and included in the review, with reasons for exclusions at each stage, ideally with a flow diagram. | | — | | If a review addresses both efficacy and harms, display a flow diagram specific for each (efficacy and harm). | X (see  *Results – Search results,* p 11 and  *Figure 1*) |
| Study characteristics (14) | 18 | For each study, present characteristics for which data were extracted (eg, study size, PICOS, follow-up period) and provide the citations. | | Define each harm addressed, how it was ascertained (eg, patient report, active search), and over what time period. | | Add additional characteristics to: “P” (population) patient risk factors that were considered as possibly affecting the risk of the harm outcome. “I” (intervention) professional expertise/skills if relevant (for example if the intervention is a procedure). “T” (time) timing of all harms assessments and the length of follow-up. | X (see  *Results – Study characteristics,* p 11-28 and  *Table D*) |
| Risk of bias within studies (15) | 19 | Present data on risk of bias of each study and, if available, any outcome level assessment (see item 12). | | — | | Consider the possible sources of biases that could affect the specific harm under consideration within the review. Sample selection, dropouts and measurement of adverse events should be evaluated separately from the outcomes of benefit as described in item 12, above. | X (see  *Results – Risk of bias and certainty of evidence,* p 29  and  *Table E*) |
| Results of individual studies (16) | 20 | For all outcomes considered (benefits or harms), present, for each study: (a) simple summary data for each intervention group (b) effect estimates and confidence intervals, ideally with a forest plot. | | — | | Report the actual numbers of adverse events in each study, separately for each intervention. | X (see *Table F*) |
| Synthesis of results (17) | 21 | Present results of each meta-analysis done, including confidence intervals and measures of consistency. | | Describe any assessment of possible causality. | | If included data from unpublished sources, report clearly the data source and the impact of these studies to the final systematic review. | X (see  *Results – Synthesis of results,* p 30-33*,  and Table F* and *Table 2,* p 13-28) |
| Risk of bias across studies (18) | 22 | Present results of any assessment of risk of bias across studies (see item 15). | | — | | No specific additional information is required for systematic reviews of harms. See item 15 above. | X (see  *Results – Risk of bias and certainty of evidence,* p 29) |
| Additional analysis (18) | 23 | Give results of additional analyses, if done (eg, sensitivity or subgroup analyses, meta-regression (see item 16)). | | — | | No specific additional information is required for systematic reviews of harms. | X (see  *Results – Synthesis of results,* p 30) |
| **Discussion** | | | | | | | |
| Summary of evidence (18) | 24 | Summarise the main findings including the strength of evidence for each main outcome; consider their relevance to key groups (eg, healthcare providers, users, and policy makers). | — | | | No specific additional information is required for systematic reviews of harms. | X (see *Discussion,*  p 34) |
| Limitations (18) | 25 | Discuss limitations at study and outcome level (eg, risk of bias), and at review level (eg, incomplete retrieval of identified research, reporting bias). | — | | | Recognise possible limitations of meta-analysis for rare adverse events (ie, quality and quantity of data), issues noted previously related to collection and reporting. | X (see *Discussion,*  p 34-38) |
| Conclusions (19) | 26 | Provide a general interpretation of the results in the context of other evidence, and implications for future research. | — | | | State conclusions in coherence with the review findings. When adverse events were not identified we caution against the conclusion that the intervention is “safe,” when, in reality, its safety remains unknown. | X (see *Discussion,*  p 34-38) |
| **Funding** | | | | | | | |
| Funding (19) | 27 | Describe sources of funding for the systematic review and other support (eg, supply of data); role of funders for the systematic review. | — | | | No specific additional information is required for systematic reviews of harms. | X (see *Acknowledgements*, p 39 and  *Online submission*) |
