## Additional file 2 for "Adverse events of iron and/or erythropoiesis-stimulating agent therapy in preoperatively anaemic elective surgery patients: a systematic review"

**Supplementary Table 1. Search string.**

**Supplementary Table 2. List of excluded studies and reasons for exclusion.**

**Supplementary Table 3. List of ongoing/prematurely ended trials and published protocols.**

**Supplementary Table 4. Overview of adverse events for which data were obtained from the 26 included randomized controlled trials (RCTs).**

**Supplementary Table 5. Overview of adverse events for which data were obtained from the 16 included cohort studies.**

**Supplementary Table 6. Characteristics of included studies.**

**Supplementary Table 7. Risk of bias assessment.**

**Supplementary Table 8. Synthesis of findings.**

**References**

### Supplementary Table 1. Search string.

**Databases**

The Cochrane Library (systematic reviews and controlled trials) using the following search strategy:

1. [mh “Perioperative Period”] OR [mh “Perioperative care”] OR preoperat*:ti,ab,kw OR pre-operat*:ti,ab,kw OR presurg*:ti,ab,kw OR pre-surg*:ti,ab,kw OR (before NEXT surger*):ti,ab,kw OR (before NEXT surgical*):ti,ab,kw OR (before NEXT operati*):ti,ab,kw OR (“before the” NEXT operation*):ti,ab,kw OR (“prior to” NEXT surger*):ti,ab,kw OR (“prior to” NEXT surgical*):ti,ab,kw OR (“prior to” NEXT operati*):ti,ab,kw OR (undergoing NEXT surger*):ti,ab,kw OR (undergoing NEXT surgical*):ti,ab,kw OR perioperat*:ti,ab,kw OR peri-operat*:ti,ab,kw OR postoperat*:ti,ab,kw OR post-operat*:ti,ab,kw
2. [mh “Anemia”] OR anemi*:ti,ab,kw OR anaemi*:ti,ab,kw OR [mh “Hemoglobins”] OR hemoglobin*:ti,ab,kw OR haemoglobin*:ti,ab,kw OR [mh “Hematocrit”] OR hematocrit*:ti,ab,kw OR haematocrit*:ti,ab,kw
3. [mh “Iron”] OR [mh “Iron Compounds”] OR iron:ti,ab,kw OR dextran:ti,ab,kw OR Venofer:ti,ab,kw OR ferrous:ti,ab,kw OR ferric:ti,ab,kw OR ferrlecit:ti,ab,kw OR femiron:ti,ab,kw OR hemocyte:ti,ab,kw OR feosol:ti,ab,kw OR bifera:ti,ab,kw OR auryxia:ti,ab,kw OR Losferron:ti,ab,kw OR Ferricure:ti,ab,kw OR Fero-Grad:ti,ab,kw OR Fero-Gradumet:ti,ab,kw OR Tardyferon:ti,ab,kw OR Fercayl:ti,ab,kw OR Injectafer:ti,ab,kw OR [mh “Erythropoietin”] OR [mh “Hematinics”] OR epo:ti,ab,kw OR erythropo*:ti,ab,kw OR (“erythropoiesis-stimulating” NEXT agent*):ti,ab,kw OR hematopoiet*:ti,ab,kw OR haematopoiet*:ti,ab,kw OR hemopoiet*:ti,ab,kw OR haemopoiet*:ti,ab,kw OR hematinic*:ti,ab,kw OR haematinic*:ti,ab,kw OR Procrit:ti,ab,kw OR NeoRecormon:ti,ab,kw OR darbepoetin:ti,ab,kw OR Mircera:ti,ab,kw OR epoetin:ti,ab,kw OR rhuepo:ti,ab,kw OR epogen*:ti,ab,kw OR eprex:ti,ab,kw OR filgrastim:ti,ab,kw OR G-CSF:ti,ab,kw OR neupogen:ti,ab,kw OR Aranesp:ti,ab,kw OR Darbecept:ti,ab,kw OR Epocept:ti,ab,kw OR Nanokine:ti,ab,kw OR Epofit:ti,ab,kw OR Epogin:ti,ab,kw OR Binocrit:ti,ab,kw OR PDpoetin:ti,ab,kw OR Bioyetin:ti,ab,kw OR Recormon:ti,ab,kw OR Dynepo:ti,ab,kw OR Epomax:ti,ab,kw OR Silapo:ti,ab,kw OR Retacrit:ti,ab,kw OR EPOTrust:ti,ab,kw OR “Erypro Safe”:ti,ab,kw OR Repoitin:ti,ab,kw OR Vintor:ti,ab,kw OR Erykine:ti,ab,kw OR Wepox:ti,ab,kw OR Espogen:ti,ab,kw OR ReliPoietin:ti,ab,kw OR Shanpoietin:ti,ab,kw OR Zyrop:ti,ab,kw OR EPIAO:ti,ab,kw OR Cinnapoietin:ti,ab,kw OR Peginesatide:ti,ab,kw OR Omontys:ti,ab,kw OR “Methoxy polyethylene glycol-epoetin beta”:ti,ab,kw
4. 1-3 AND

MEDLINE (via PubMed interface) for experimental and observational studies using the following search strategy:

1. “Perioperative Period”[Mesh] OR “Perioperative Care”[Mesh] OR preoperat*[TIAB] OR pre-operat*[TIAB] OR presurg*[TIAB] OR pre-surg*[TIAB] OR before surger*[TIAB] OR before surgical*[TIAB] OR before operati*[TIAB] OR before the operation*[TIAB] OR prior to surger*[TIAB] OR prior to surgical*[TIAB] OR prior to operati*[TIAB] OR undergoing surger*[TIAB] OR undergoing surgical*[TIAB] OR perioperat*[TIAB] OR peri-operat*[TIAB] OR postoperat*[TIAB] OR post-operat*[TIAB]
2. “Anemia”[Mesh] OR anemi*[TIAB] OR anaemi*[TIAB] OR “Hemoglobins”[Mesh] OR hemoglobin*[TIAB] OR haemoglobin*[TIAB] OR Hematocrit[Mesh] OR hematocrit*[TIAB] OR haematocrit*[TIAB]
3. “Iron”[Mesh] OR “Iron Compounds”[Mesh] OR iron[TIAB] OR dextran[TIAB] OR Venofer[TIAB] OR ferrous[TIAB] OR ferric[TIAB] OR ferrlecit[TIAB] OR femiron[TIAB] OR hemocyte[TIAB] OR feosol[TIAB] OR bifera[TIAB] OR auryxia[TIAB] OR Losferron[TIAB] OR Ferricure[TIAB] OR Fero-Grad[TIAB] OR Fero-Gradumet[TIAB] OR Tardyferon[TIAB] OR Fercayl[TIAB] OR Injectafer[TIAB] OR“Erythropoietin”[Mesh] OR “Hematinics”[Mesh] OR epo[TIAB] OR erythropo*[TIAB] OR erythropoiesis-stimulating agent*[TIAB] OR hematopoiet*[TIAB] OR haematopoiet*[TIAB] OR hemopoiet*[TIAB] OR haemopoiet*[TIAB] OR hematinic*[TIAB] OR haematinic*[TIAB] OR Procrit[TIAB] OR Epogen*[TIAB] OR NeoRecormon[TIAB] OR darbepoetin[TIAB] OR Mircera[TIAB] OR epoetin[TIAB] OR rhuepo[TIAB] OR eprex[TIAB] OR filgrastim[TIAB] OR G-CSF[TIAB] OR neupogen[TIAB] OR Aranesp[TIAB] OR Darbecept[TIAB] OR Epocept[TIAB] OR Nanokine[TIAB] OR Epofit[TIAB] OR Epogin[TIAB] OR Binocrit[TIAB] OR PDpoetin[TIAB] OR Bioyetin[TIAB] OR Recormon[TIAB] OR Dynepo[TIAB] OR Epomax[TIAB] OR Silapo[TIAB] OR Retacrit[TIAB] OR EPOTrust[TIAB] OR “Erypro Safe”[TIAB] OR Repoitin[TIAB] OR Vintor[TIAB] OR Erykine[TIAB] OR Wepox[TIAB] OR Espogen[TIAB] OR ReliPoietin[TIAB] OR Shanpoietin[TIAB] OR Zyrop[TIAB] OR EPIAO[TIAB] OR Cinnapoietin[TIAB] OR Peginesatide[TIAB] OR Omontys[TIAB] OR “Methoxy polyethylene glycol-epoetin beta”[TIAB]
4. 1-3 AND

Embase (via Embase.com interface) using the following search strategy:

1. ‘Preoperative period’/exp OR ‘Perioperative period’/exp OR ‘Postoperative period’/exp OR preoperat*:ab,ti OR pre-operat*:ab,ti OR presurg*:ab,ti OR pre-surg*:ab,ti OR (before NEXT/1 surger*):ab,ti OR (before NEXT/1 surgical*):ab,ti OR (before NEXT/1 operati*):ab,ti OR (‘before the’ NEXT/1 operation*):ab,ti OR (‘prior to’ NEXT/1 surger*):ab,ti OR (‘prior to’ NEXT/1 surgical*):ab,ti OR (‘prior to’ NEXT/1 operati*):ab,ti OR (undergoing NEXT/1 surger*):ab,ti OR (undergoing NEXT/1 surgical*):ab,ti OR perioperat*:ab,ti OR peri-operat*:ab,ti OR postoperat*:ab,ti OR post-operat*:ab,ti
2. ‘Anemia’/exp OR anemi*:ab,ti OR anaemi*:ab,ti OR ‘Hemoglobin’/exp OR hemoglobin*:ab,ti OR haemoglobin*:ab,ti OR ‘Hematocrit’/exp OR hematocrit*:ab,ti OR haematocrit*:ab,ti
3. ‘Antianemic agent’/exp OR ‘Iron’/exp OR ‘Iron derivative’/exp OR iron:ab,ti OR dextran:ab,ti OR Venofer:ab,ti OR ferrous:ab,ti OR ferric:ab,ti OR ferrlecit:ab,ti OR femiron:ab,ti OR hemocyte:ab,ti OR feosol:ab,ti OR bifera:ab,ti OR auryxia:ab,ti OR Losferron:ab,ti OR Ferricure:ab,ti OR Fero-Grad:ab,ti OR Fero-Gradumet:ab,ti OR Tardyferon:ab,ti OR Fercayl:ab,ti OR Injectafer:ab,ti OR epo:ab,ti OR erythropo*:ab,ti OR (‘erythropoiesis-stimulating’ NEXT/1 agent*):ab,ti OR hematopoiet*:ab,ti OR haematopoiet*:ab,ti OR hemopoiet*:ab,ti OR haemopoiet*:ab,ti OR hematinic*:ab,ti OR haematinic*:ab,ti OR Procrit:ab,ti OR Epogen*:ab,ti OR NeoRecormon:ab,ti OR darbepoetin:ab,ti OR Mircera:ab,ti OR epoetin:ab,ti OR rhuepo:ab,ti OR eprex:ab,ti OR filgrastim:ab,ti OR G-CSF:ab,ti OR neupogen:ab,ti OR Aranesp:ab,ti OR Darbecept:ab,ti OR Epocept:ab,ti OR Nanokine:ab,ti OR Epofit:ab,ti OR Epogin:ab,ti OR Binocrit:ab,ti OR PDpoetin:ab,ti OR Bioyetin:ab,ti OR Recormon:ab,ti OR Dynepo:ab,ti OR Epomax:ab,ti OR Silapo:ab,ti OR Retacrit:ab,ti OR EPOTrust:ab,ti OR ‘Erypro Safe’:ab,ti OR Repoitin:ab,ti OR Vintor:ab,ti OR Erykine:ab,ti OR Wepox:ab,ti OR Espogen:ab,ti OR ReliPoietin:ab,ti OR Shanpoietin:ab,ti OR Zyrop:ab,ti OR EPIAO:ab,ti OR Cinnapoietin:ab,ti OR Peginesatide:ab,ti OR Omontys:ab,ti OR ‘Methoxy polyethylene glycol-epoetin beta’:ab,ti
4. 1-3 AND

Transfusion Evidence Library using the following search strategy:

1. Subject Area < Clinical Practice < Management of anaemia
2. perioperative OR peri-operative OR postoperative OR post-operative OR preoperative OR pre-operative OR surgical OR surgery OR operating OR operation OR surgically OR surgeries
3. Study design < Randomized Controlled Trial or Economic study
4. 1-3 AND

Web of Science using the following search strategy:

1. TS=(“preoperat*”) OR TI=(“preoperat*”) OR TS=(“pre-operat*”) OR TI=(“preoperat*”) OR TS=(“presurg*”) OR TI=(“presurg*”) OR TS=(“pre-surg*”) OR TI=(“pre-surg*”) OR TS=(“before surger*”) OR TI=(“before surger*”) OR TS=(“before surgical*”) OR TI=(“before surgical*”) OR TS=(“before operati*”) OR TI=(“before operati*”) OR TS=(“before the operation*”) OR TI=(“before the operation*”) OR TS=(“prior to surger*”) OR TI=(“prior to surger*”) OR TS=(“prior to surgical*”) OR TI=(“prior to surgical*”) OR TS=(“prior to operati*”) OR TI=(“prior to operati*”) OR TS=(“undergoing surger*”) OR TI=(“undergoing surger*”) OR TS=(“undergoing surgical*”) OR TI=(“undergoing surgical*”) OR TS=(“perioperat*”) OR TI=(“perioperat*”) OR TS=(“peri-operat*”) OR TI=(“peri-operat*”) OR TS=(“postoperat*”) OR TI=(“postoperat*”) OR TS=(“post-operat*”) OR TI=(“post-operat*”)
2. TS=(“anemi*”) OR TI=(“anemi*”) OR TS=(“anaemi*”) OR TI=(“anaemi*”) OR TS=(“hemoglobin*”) OR TI=(“hemoglobin*”) OR TS=(“haemoglobin*”) OR TI=(“haemoglobin*”) OR TS=(“hematocrit*”) OR TI=(“hematocrit*”) OR TS=(“haematocrit*”) OR TI=(“haematocrit*”)
3. TS=(“iron”) OR TI=(“iron”) OR TS=(“dextran”) OR TI=(“dextran”) OR TS=(“Venofer”) OR TI=(“Venofer”) OR TS=(“ferrous”) OR TI=(“ferrous”) OR TS=(“ferric”) OR TI=(“ferric”) OR TS=(“ferrlecit”) OR TI=(“ferrlecit”) OR TS=(“femiron”) OR TI=(“femiron”) OR TS=(“hemocyte”) OR TI=(“hemocyte”) OR TS=(“feosol”) OR TI=(“feosol”) OR TS=(“bifera”) OR TI=(“bifera”) OR TS=(“auryxia”) OR TI=(“auryxia”) OR TS=(“Losferron”) OR TI=(“Losferron”) OR TS=(“Ferricure”) OR TI=(“Ferricure”) OR TS=(“Fero-Grad”) OR TI=(“Fero-Grad”) OR TS=(“Fero-Gradumet”) OR TI=(“Fero-Gradumet”) OR TS=(“Tardyferon”) OR TI=(“Tardyferon”) OR TS=(“Fercayl”) OR TI=(“Fercayl”) OR TS=(“Injectafer”) OR TI=(“Injectafer”) OR TS=(“epo”) OR TI=(“epo”) OR TS=(“erythropo*”) OR TI=(“erythropo*”) OR TS=(“erythropoiesis-stimulating agent*”) OR TI=(“erythropoiesis-stimulating agent*”) OR TS=(“hematopoiet*”) OR TI=(“hematopoiet*”) OR TS=(“haematopoiet*”) OR TI=(“haematopoiet*”) OR TS=(“hemopoiet*”) OR TI=(“hemopoiet*”) OR TS=(“haemopoiet*”) OR TI=(“haemopoiet*”) OR TS=(“hematinic*”) OR TI=(“hematinic*”) OR TS=(“haematinic*”) OR TI=(“haematinic*”) OR TS=(“Procrit”) OR TI=(“Procrit”) OR TS=(“Epogen*”) OR TI=(“Epogen*”) OR TS=(“NeoRecormon”) OR TI=(“NeoRecormon”) OR TS=(“darbepoetin”) OR TI=(“darbepoetin”) OR TS=(“Mircera”) OR TI=(“Mircera”) OR TS=(“epoetin”) OR TI=(“epoetin”) OR TS=(“rhuepo”) OR TI=(“rhuepo”) OR TS=(“eprex”) OR TI=(“eprex”) OR TS=(“filgrastim”) OR TI=(“filgrastim”) OR TS=(“G-CSF”) OR TI=(“G-CSF”) OR TS=(“neupogen”) OR TI=(“neupogen”) OR TS=(“Aranesp”) OR TI=(“Aranesp”) OR TS=(“Darbecept”) OR TI=(“Darbecept”) OR TS=(“Epocept”) OR TI=(“Epocept”) OR TS=(“Nanokine”) OR TI=(“Nanokine”) OR TS=(“Epofit”) OR TI=(“Epofit”) OR TS=(“Epogin”) OR TI=(“Epogin”) OR TS=(“Binocrit”) OR TI=(“Binocrit”) OR TS=(“PDpoetin”) OR TI=(“PDpoetin”) OR TS=(“Bioyetin”) OR TI=(“Bioyetin”) OR TS=(“Recormon”) OR TI=(“Recormon”) OR TS=(“Dynepo”) OR TI=(“Dynepo”) OR TS=(“Epomax”) OR TI=(“Epomax”) OR TS=(“Silapo”) OR TI=(“Silapo”) OR TS=(“Retacrit”) OR TI=(“Retacrit”) OR TS=(“EPOTrust”) OR TI=(“EPOTrust”) OR TS=(“Erypro Safe”) OR TI=(“Erypro Safe”) OR TS=(“Repoitin”) OR TI=(“Repoitin”) OR TS=(“Vintor”) OR TI=(“Vintor”) OR TS=(“Erykine”) OR TI=(“Erykine”) OR TS=(“Wepox”) OR TI=(“Wepox”) OR TS=(“Espogen”) OR TI=(“Espogen”) OR TS=(“ReliPoietin”) OR TI=(“ReliPoietin”) OR TS=(“Shanpoietin”) OR TI=(“Shanpoietin”) OR TS=(“Zyrop”) OR TI=(“Zyrop”) OR TS=(“EPIAO”) OR TI=(“EPIAO”) OR TS=(“Cinnapoietin”) OR TI=(“Cinnapoietin”) OR TS=(“Peginesatide”) OR TI=(“Peginesatide”) OR TS=(“Omontys”) OR TI=(“Omontys”) OR TS=(“Methoxy polyethylene glycol-epoetin beta”) OR TI=(“Methoxy polyethylene glycol-epoetin beta”)
4. 1-3 AND

**Trial registries**

WHO International Clinical Trials Registry Platform using the following search strategy:

Condition: Anaemia OR anemia

AND

Intervention: Iron or epo or erythropoiesis-stimulating agents or epogen or neorecormon or darbepoetin or epoetin

AND

Recruitment status: ALL

Clinicaltrials.Gov using the following search strategy:

Anemia [DISEASE]

Adult_Older adult [AGE GROUP]

“Interventional” [STUDY TYPE]

Iron or epo or erythropoiesis-stimulating agents or epogen or NeoRecormon or darbepoetin or epoetin [INTERVENTION]

**Reference lists and related citations**

Included articles, retrieved with the above searches, were used to identify other studies by searching (1) reference lists and (2) 20 first related citations in MEDLINE (via PubMed interface).

### Supplementary Table 2. List of excluded studies and reasons for exclusion.

| **Reference** | **Reason for exclusion** |
| --- | --- |
| Lower transfusion rate with similar outcomes for symptomatic triggers of transfusion versus hemoglobin level target in elderly surgical patients with cardiovascular disease. Clinical Geriatrics. 2009;17(12):13-6. | Design; Type of news report |
| A multicentre comparative study on the efficacy of intravenous ferric carboxymaltose and iron sucrose for correcting preoperative anaemia in patients undergoing major elective surgery British Journal of Anaesthesia, 2015; 107(3): 477-78, DOI 10.1093/bja/aer242. Br J Anaesth. 2015;115(1):154. doi: 10.1093/bja/aev158. Epub 2015 May 7. | Design |
| Correction to: the impact of pre-operative intravenous iron on quality of life after colorectal cancer surgery: outcomes from the intravenous iron in colorectal cancer-associated anaemia (IVICA) trial (Anaesthesia, (2019), 74, 6, (714-725), 10.1111/anae.14659). Anaesthesia. 2019;74(9):1191‐. doi: 10.1111/anae.14785. PubMed PMID: CN-01993863. | Outcome; Correction to Keeler 2019, that was already included. Corrections concern the outcomes of quality of life, which were not extracted for our review. |
| Abbott TEF, Gillies MA. The PREVENNT randomised, double-blind, controlled trial of preoperative intravenous iron to treat anaemia before major abdominal surgery: an independent discussion. Br J Anaesth. 2020. Epub 2020/10/18. doi: 10.1016/j.bja.2020.08.053. PubMed PMID: 33066972. | Design; discusses the results of the trial by Richards et al. |
| Abdullah HR, Sim YE, Sim YTM, Lamoureux E. Preoperative ANemiA among the elderly undergoing major abdominal surgery (PANAMA) study: Protocol for a single-center observational cohort study of preoperative anemia management and the impact on healthcare outcomes. Medicine. 2018;97(21). doi: 10.1097/MD.0000000000010838. | Intervention |
| Acheson AG, Brookes MJ, Spahn DR. Effects of Allogeneic Red Blood Cell Transfusions on Clinical Outcomes in Patients Undergoing Colorectal Cancer Surgery A Systematic Review and Meta-Analysis. Annals of Surgery. 2012;256(2):235-44. doi: 10.1097/SLA.0b013e31825b35d5. | Design |
| Actrn. Assessment of a single intravenous iron infusion versus standard care in the management of Post-OPerative Iron (POPi) deficiency anaemia patients undergoing elective surgery at the Launceston General Hospital. Http://wwwanzctrorgau/actrn12614001261606aspx. 2014. | Population |
| Actrn. Assessment of a single intravenous (IV) iron therapy versus oral iron in the management of preoperative anaemia patients undergoing elective surgery at the Launceston General Hospital. 2011. | Other; Full text paper already included |
| Actrn. Safety of iron polymaltose infusion given over 30 and 15 minutes for treatment of iron deficiency. 2017. | Population; no elective surgery patients |
| Actrn. An observational study of perioperative anaemia and transfusion in surgical patients at St George Hospital. 2018. | Intervention |
| Adamson J, Messmer, Monk, Mercuriali, D'Ambra. Perisurgical use of epoetin alfa in orthopedic surgery patients. Seminars in Hematology. 1996;33(2 SUPPL. 2):55-9. | Design |
| Agostini V, Delucia M, Gagliardi S, Gasperoni A, Santarelli R. Preoperative use of erythropoietin as an alternative to preoperative blood donation programme in elective orthopedic surgery. Transfusion Alternatives in Transfusion Medicine. 2012;12(2):22. doi: 10.1111/j.1778-428X.2012.01163.x. | Design |
| Albion, Sprim Advanced L, Sciences. A Study to Compare the Gastrointestinal Tolerability of Ferrochel®, Sumalate®,Ferrous Fumarate, Ferrous Sulfate, Ferric Glycinate, and Placebo. 2014. | Population; no elective surgery patients |
| Althoff FC, Neb H, Herrmann E, Trentino KM, Vernich L, Fullenbach C, et al. Multimodal Patient Blood Management Program Based on a Three-pillar Strategy A Systematic Review and Meta-analysis. Annals of Surgery. 2019;269(5):794-804. doi: 10.1097/SLA.0000000000003095. PubMed PMID: WOS:000472682400015. | Intervention; Multimodal PBM programs. Not possible to distinguish pure effects of iron/EPO. |
| Amag P, Inc. A Trial Comparing Ferumoxytol With Placebo for the Treatment of Iron Deficiency Anemia. 2012. | Population; no elective surgery patients |
| Amag P, Inc. A Trial of Ferumoxytol for the Episodic Treatment of Iron Deficiency Anemia. 2012. | Population; no elective surgery patients |
| Amag P, Inc. A Phase III Safety Study of Ferumoxytol Compared to Ferric Carboxymaltose for the Treatment of Iron Deficiency Anemia (IDA). 2017. | Population; no elective surgery patients |
| American R, Inc. Safety and Tolerability of a Single Dose of FCM vs. Standard of Care in Treating Iron Deficiency Anemia. 2009. | Population; no elective surgery patients |
| American R, Inc. Safety and Tolerability of Ferric Carboxymaltose (FCM) Versus Standard of Care in Treating Iron Deficiency Anemia. 2009. | Population; no elective surgery patients |
| Andrews CM, Lane DW, Bradley JG. Iron pre-load for major joint replacement. Transfusion Medicine. 1997;7(4):281-6. doi: 10.1046/j.1365-3148.1997.d01-42.x. | Comparison |
| Aniteye EA, Sereboe L, Kotei D, Frimpong-Boateng K, Adu-Gyamfi Y. The efficacy of pre-operative erythropoietin therapy. East African medical journal. 2007;84(6):279-82. | Design |
| ApoPharma. Efficacy and Safety of Ferriprox® in Patients With Sickle Cell Disease or Other Anemias. 2019. | Population; no elective surgery patients |
| Asuero MS, Rubial M. Methods of blood saving in the surgical patient. Revista española de anestesiología y reanimación. 1995;42(7):290-5. | Design; Narrative review |
| Auerbach Hematology Oncology A, P C, Amag P, Inc. Total Dose Infusion of Ferumoxytol(1020mg) in 15 Minutes for Iron Deficiency Anemia. 2012. | Design; Uncontrolled study |
| Auerbach M, Pappadakis JA, Bahrain H, Auerbach SA, Ballard H, Dahl NV. Safety and efficacy of rapidly administered (one hour) one gram of low molecular weight iron dextran (INFeD) for the treatment of iron deficient anemia. American Journal of Hematology. 2011;86(10):860-2. doi: 10.1002/ajh.22153. | Design |
| Bailey W, Bourne R, Feagan B, Grainger R, Laupacis A, Phillips T, et al. Effectiveness of perioperative recombinant human erythropoietin in elective hip replacement. Lancet. 1993;341(8855):1227-32. | Outcome; nothing on adverse events |
| Basora M, Colomina MJ, Tio M, Mora L, Salazar F, Ciercoles E. Optimizing preoperative haemoglobin with intravenous iron. Br J Anaesth. 2013;110(3):488-90. doi: 10.1093/bja/aes587. | Design |
| Basora M, Colomina MJ, Tio M, Mora L, Sánchez-Etayo G, Salazar F, et al. Optimizing preoperative haemoglobin in major orthopaedic surgery using intravenous iron with or without erythropoietin. An epidemiologic study. Revista espanola de anestesiologia y reanimacion. 2015;62(6):313-21. doi: 10.1016/j.redar.2014.07.011. | Other; duplicate |
| Basora M, Pereira A, Coca M, Tio M, Lozano L. Cost-effectiveness analysis of ferric carboxymaltose in pre-operative haemoglobin optimisation in patients undergoing primary knee arthroplasty. Blood Transfusion. 2018;16(5):438-42. doi: 10.2450/2018.0031-18. | Design |
| Bedair H, Yang J, Dwyer MK, McCarthy JC. Preoperative Erythropoietin Alpha Reduces Postoperative Transfusions in THA and TKA but May Not Be Cost-effective. Clinical Orthopaedics and Related Research. 2015;473(2):590-6. doi: 10.1007/s11999-014-3819-z. | Outcome; nothing on adverse events |
| Biboulet P, Motais C, Pencole M, Karam O, Dangelser G, Smilevitch P, et al. Preoperative erythropoietin within a patient blood management program decreases both blood transfusion and postoperative anemia: a prospective observational study. Transfusion. 2020;60(8):1732-40. Epub 2020/07/19. doi: 10.1111/trf.15900. PubMed PMID: 32681743. | Design; Post-hoc data analysis on a subset of patients included in Biboulet 2018, which was already included. |
| Bisbe E, Rodríguez C, Ruiz A, Sáez M, Castillo J, Santiveri X. Preoperative use of intravenous iron: a new transfusional therapy. Revista española de anestesiología y reanimación. 2005;52(9):536-40. | Population |
| Bisbe E, Sáez M, Pérez C, Castillo J, García-Erce JA. Delayed improvement of anemia treated with intravenous iron and epoetin alfa after hip replacement surgery. Revista española de anestesiología y reanimación. 2005;52(8):507-9. | Population |
| Bisbe V. Treatment of anemia in Patient Blood Management from an economic perspective. Revista espanola de anestesiologia y reanimacion. 2015;62:80-5. doi: 10.1016/S0034-9356(15)30013-X. | Other; Not available |
| Bonnet V, Lienhart A. Iron deficiency treatment, erythropoietin and post-partum. Annales Francaises D Anesthesie Et De Reanimation. 2002;21(6):FI84-FI92. | Design; Narrative review |
| Borstlap WAA, Buskens CJ, Tytgat K, Dijkgraaf MGW, Tanis PJ, Bemelman WA. FIT trial: multicentre randomised controlled trial comparing Ferric (III) carboxymaltose infusion with oral iron supplementation in the treatment of preoperative anaemia in colorectal cancer patients. Colorectal disease. 2015;17:104‐. doi: 10.1111/codi.13054. | Design |
| Borstlap WA, Buskens CJ, Tytgat KM, Tuynman JB, Consten EC, Tolboom RC, et al. Erratum to: Multicentre randomized controlled trial comparing ferric(III) carboxymaltose infusion with oral iron supplementation in the treatment of preoperative anaemia in colorectal cancer patients. BMC Surg. 2015;15:110. doi: 10.1186/s12893-015-0090-5. | Design |
| Braga M, Gentilini O, Gianotti L, Vignali A, Di C. Preoperative treatment with recombinant human erythropoietin in anemic patients suffering from digestive tract neoplasms. Chirurgia. 1997;10(6):515-8. | Other; Not available |
| Braga M, Gianotti L, Gentilini O, Vignali A, Corizia L, Di C. Erythropoiesis after therapy with recombinant human erythropoietin: a dose-response study in anemic cancer surgery patients. Vox Sanguinis. 1999;76((1):):38-42. doi: 10.1046/j.1423-0410.1999.7610038. | Comparison |
| Braga M, Gianotti L, Gentilini O, Vignali A, DiCarlo V. Erythropoietic response induced by recombinant human erythropoietin in anemic cancer patients candidate to major abdominal surgery. Hepato-Gastroenterology. 1997;44(15):685-90. | Outcome; nothing on adverse events |
| Briguglio M, Hrelia S, Malaguti M, De Vecchi E, Lombardi G, Banfi G, et al. Oral supplementation with sucrosomial ferric pyrophosphate plus l-ascorbic acid to ameliorate the martial status: a randomized controlled trial. Nutrients. 2020;12(2). doi: 10.3390/nu12020386. | Population; Only a small portion of the included participants was anaemic at baseline (8,2% according to the WHO criteria). No separate data available for these anaemic patients. |
| Cahill C, Blumberg N, Melvin A, Knight P, Gloff M, Robinson R, et al. Pre-operative anemia management program reduces blood transfusion in elective cardiac surgical patients. Transfusion. 2018;58:39A. doi: 10.1111/trf.14903. | Design |
| Calleja JL, Delgado S, del V, Hervás A, Larraona JL, Terán Á, et al. Ferric carboxymaltose reduces transfusions and hospital stay in patients with colon cancer and anemia. International Journal of Colorectal Disease. 2016;31(3):543-51. doi: 10.1007/s00384-015-2461-x. | Outcome; nothing on adverse events |
| Calvet X, Gené E, Àngelruíz M, Figuerola A, Villoria A, Cucala M, et al. Cost-minimization analysis favours intravenous ferric carboxymaltose over ferric sucrose or oral iron as preoperative treatment in patients with colon cancer and iron deficiency anaemia. Technology and Health Care. 2016;24(1):111-20. doi: 10.3233/THC-151074. | Design |
| Candiano G. Efficacy and tolerability of oral Sucrosomial® iron supplementation in urologic patients presenting with pre-operative iron deficiency anemia. Blood Transfusion. 2020;18:s31. | Design; conference abstract |
| Centre F, Baclesse, Hospira now a wholly owned subsidiary of P, Roche P, A G, Vifor P. Randomized Study Evaluating Agents Stimulants Erythropoiesis (ASE) Associated With Ferric Carboxymaltose (Ferinject ®) in Concomitant or Sequential Patients Treated for Cancer and With Anemia Associated With Functional Iron Deficiency. 2014. | Population; no elective surgery patients |
| Chan PTY, Corallo CE, Dooley MJ, Poole SG, Gibson PR. Safety of rapid infusion of iron polymaltose: Comparative study in 300 patients. Journal of Pharmacy Practice and Research. 2016;46(4):324-30. doi: 10.1002/jppr.1158. | Population |
| Chan MY, Kam PMH, Chu CWH, Kwok KH. Comparing the efficacy and safety of monofer and venofer in colorectal cancer patients with iron deficiency anemia. Surgical Practice. 2019;23:18. doi: 10.1111/1744-1633.12389. | Design; conference abstract |
| Charbonneau H, Pasquié M, Berthoumieu P, Savy N, Autones G, Anglès O, et al. Patient blood management in elective bypass cardiac surgery: A 2-step single-centre interventional trial to analyse the impact of an educational programme and erythropoiesis stimulation on red blood cell transfusion. Contemporary clinical trials communications. 2020;19:100617. Epub 2020/07/23. doi: 10.1016/j.conctc.2020.100617. | Intervention; Treatment given will depend on the Hb and iron status of the patients, i.e. if Hb<13 g/dl, the patient will receive EPO. If the patient has iron deficiency, he will receive IV iron. Hence, if a group of people receiving EPO + iron is compared to a group only receiving EPO or iron, the populations of interest will not be comparable. |
| Cho BC, Serini J, Zorrilla-Vaca A, Scott MJ, Gehrie EA, Frank SM, et al. Impact of Preoperative Erythropoietin on Allogeneic Blood Transfusions in Surgical Patients: Results From a Systematic Review and Meta-analysis. Anesth Analg. 2019;128(5):981-92. Epub 2019/01/17. doi: 10.1213/ane.0000000000004005. | Other; all relevant individual studies covered by our review. |
| Chu MWA, Losenno KL, Moore K, Berta D, Hewitt J, Ralley F. Blood conservation strategies reduce the need for transfusions in ascending and aortic arch surgery. Perfusion (United Kingdom). 2013;28(4):315-21. doi: 10.1177/0267659113479816. | Population; Combination of both elective and emergent procedures. Moreover, Hb levels of both groups not clear. |
| Courtney JB, Cushner F, Long WJ, Nett MP. An effective bloodless surgery protocol. Techniques in Knee Surgery. 2011;10(4):188-97. doi: 10.1097/BTK.0b013e31823b31ea. | Design; Narrative review |
| Couvret C, Laffon M, Baud A, Payen V, Burdin P, Fusciardi J. A restrictive use of both autologous donation and recombinant human erythropoietin is an efficient policy for primary total hip or knee arthroplasty. Anesthesia and Analgesia. 2004;99(1):262-71. doi: 10.1213/01.ane.0000118165.70750.78. | Intervention; depends on the baseline Hct levels: - study 1:  * baseline Hct levels >33%: preoperative autologous blood donation (PADB) + oral iron.  *Other patients only receive oral iron. - study 2:  * baseline Hct levels < or = 37%: EPO + oral iron * baseline Hct levels between 37 and 39%: PABD. * baseline Hct levels >39%: no PADB. |
| Coyle D, Lee KM, Fergusson DA, Laupacis A. Economic analysis of erythropoietin use in orthopaedic surgery. Transfusion Medicine. 1999;9((1):):21-30. doi: 10.1046/j.1365-3148.1999.009001021.x. | Design |
| Cuenca J, García-Erce JA, Martínez F, Cardona R, Pérez-Serrano L, Muñoz M. Preoperative haematinics and transfusion protocol reduce the need for transfusion after total knee replacement. International Journal of Surgery. 2007;5(2):89-94. doi: 10.1016/j.ijsu.2006.02.003. | Intervention; folic acid / vit C |
| Cuenca J, García-Erce JA, Martínez F, Pérez-Serrano L, Herrera A, Muñoz M. Perioperative intravenous iron, with or without erythropoietin, plus restrictive transfusion protocol reduce the need for allogeneic blood after knee replacement surgery. Transfusion. 2006;46(7):1112-9. doi: 10.1111/j.1537-2995.2006.00859.x. | Comparison |
| Cushner FD, Locker JR, Hanssen AD, Jacosky DJ, Scott WN, Scuderi GR, et al. Use of recombinant human erythropoietin in two-stage total knee arthroplasty for infection. Clinical Orthopaedics and Related Research. 2001;(392):116-23. | Outcome; nothing on adverse events |
| D'Ambra M, Messmer, Gombotz. Perioperative epoetin alfa reduces transfusion requirements in coronary artery bypass graft surgery. Seminars in Hematology. 1996;33(2 SUPPL. 2):73-4. | Design |
| D'Ambra MN, Gray RJ, Hillman R, Jones JW, Kim HC, Rawitscher R, et al. Effect of recombinant human erythropoietin on transfusion risk in coronary bypass patients. Annals of Thoracic Surgery. 1997;64((6):):1686-93. doi: 10.1016/s0003-4975(97)00839-4. | Population; Non-anemic population |
| de Andrade, J R, Frei D, Guilfoyle M. Integrated analysis of thrombotic/vascular event occurrence in epoetin alfa-treated patients undergoing major, elective orthopedic surgery. Orthopedics. 1999;22(1):S113-S8. | Design; Pooled results from individual studies |
| De Bellis M, Girelli D, Ruzzenente A, Bagante F, Ziello R, Campagnaro T, et al. Pancreatic resections in patients who refuse blood transfusions. The application of a perioperative protocol for a true bloodless surgery. Pancreatology : official journal of the International Association of Pancreatology (IAP) [et al]. 2020;20(7):1550-7. Epub 2020/09/21. doi: 10.1016/j.pan.2020.08.020. | Intervention; No comparison between people who received preoperative EPO therapy (n=19) and those who did not. |
| Delaforce A, Galeel L, Poon E, Hurst C, Duff J, Munday J, et al. Preoperative Anemia Screening and Treatment Practices in Patients Having Total Joint Replacement Surgery: A Retrospective, Observational Audit. Journal of blood medicine. 2020;11:259-65. Epub 2020/08/22. doi: 10.2147/jbm.S254116. | Population; Treatment given depends on the urgency of the surgery (non-urgent surgery: oral iron vs urgent surgery: iv iron). Both groups will not be comparable. |
| Delasotta LA, Orozco F, Jafari SM, Blair JL, Ong A. Should We Use Preoperative Epoetin-alpha in the Mildly Anemic Patient Undergoing Simultaneous Total Knee Arthroplasty? Open Orthop J. 2013;7:47-50. doi: 10.2174/1874325001307010047. | Outcome; nothing on adverse events |
| Derzon J, Alford A, Clarke N, Gross I, Shander A, Thurer R. Anemia Management and Audit Feedback Practices for Reducing Overuse of RBC Transfusion: A Laboratory Medicine Best Practice Systematic Review and Meta-Analysis. American Journal of Clinical Pathology. 2019;151(1):18-28. doi: 10.1093/ajcp/aqy123. | Population; Mix of surgical and non-surgical patients |
| Dialysis C, Inc. Iron Indices and Intravenous Ferumoxytol: Time to Steady State. 2010. | Design; Uncontrolled study |
| Dickson EA, Keeler BD, Ng O, Kumar A, Brookes MJ, Acheson AG. Reply to comment on "Preoperative Intravenous Iron Therapy and Survival after Colorectal Cancer Surgery: Long Term Results from the IVICA Randomised Controlled Trial". Colorectal disease : the official journal of the Association of Coloproctology of Great Britain and Ireland. 2020. Epub 2020/10/26. doi: 10.1111/codi.15414. | Design; letter |
| Djavan B, Laze J, Eckersberger E, Finkelstein J, Agalliu I, Lepor H. The short-term use of erythropoetin-stimulating agents: impact on the biochemical recurrence of prostate cancer. Bju International. 2011;108(10):1582-7. doi: 10.1111/j.1464-410X.2011.10173.x. | Population; Non-anemic (all patients with a baseline haematocrit ≤48) |
| Doodeman HJ, van H, I MM, Egberts TCG, Bennis M, Traast HS, et al. The effect of a preoperative erythropoietin protocol as part of a multifaceted blood management program in daily clinical practice (CME). Transfusion. 2013;53(9):1930-9. doi: 10.1111/trf.12016. | Outcome; nothing on adverse events |
| dos Santos, A A, da S, J P, da S, L D, et al. Therapeutic options to minimize allogeneic blood transfusions and their adverse effects in cardiac surgery: A systematic review. Revista Brasileira De Cirurgia Cardiovascular. 2014;29(4):606-21. doi: 10.5935/1678-9741.20140114. | Design; SR did not search Embase; no adequate search strategy; no clear selection criteria. |
| Drabinski T, Zacharowski K, Meybohm P, Rüger AM, Ramirez de Arellano A. Estimating the Epidemiological and Economic Impact of Implementing Preoperative Anaemia Measures in the German Healthcare System: The Health Economic Footprint of Patient Blood Management. Advances in therapy. 2020;37(8):3515-36. Epub 2020/06/21. doi: 10.1007/s12325-020-01372-4. | Intervention; Unclear what is meant by preoperative anaemia measures (PAMs). In the paper, it is called "principally iron therapy"), but not possible to determine which treatments were compared. |
| DRKS00015857. Reduction of the red cell concentrate use by patient blood management. Changing the pretransfusional hemoglobin trigger by education of transfusion triggers and modification of the transfusion ordering form. 2018. | Intervention; Broad PBM intervention, consisting of training of the physicians and making changes to the request form for blood products. No specific mentioning of ESA/iron. |
| Duce L, Cooter ML, McCartney SL, Lombard FW, Guinn NR. Outcomes in Patients Undergoing Cardiac Surgery Who Decline Transfusion and Received Erythropoietin Compared to Patients Who Did Not: A Matched Cohort Study. Anesthesia and Analgesia. 2018;127(2):490-5. doi: 10.1213/ANE.0000000000002418. | Population |
| Dunphy FR, Dunleavy TL, Harrison BR, Boyd JH, Varvares MA, Dunphy CH, et al. Erythropoietin reduces anemia and transfusions after chemotherapy with paclitaxel and carboplatin. Cancer. 1997;79(8):1623-8. doi: 10.1002/(SICI)1097-0142(19970415)79:8<1623::AID-CNCR28>3.0.CO;2-Z. | Population |
| Dunphy FR, Harrison BR, Dunleavy TL, Rodriguez JJ, Hilton JG, Boyd JH. Erythropoietin reduces anemia and transfusions - A randomized trial with or without erythropoietin during chemotherapy. Cancer. 1999;86(7):1362-7. doi: 10.1002/(SICI)1097-0142(19991001)86:7<1362::AID-CNCR36>3.0.CO;2-T. | Population |
| Edwards TJ, Noble EJ, Durran A, Mellor N, Hosie KB. Randomized clinical trial of preoperative intravenous iron sucrose to reduce blood transfusion in anaemic patients after colorectal cancer surgery. The British Journal of Surgery. 2009;96((10):):1122-8. doi: 10.1002/bjs.6688. | Outcome; nothing on adverse events |
| Eeles A, Baikady RR. Peri-operative blood management. Indian Journal of Anaesthesia. 2017;61(6):456-62. doi: 10.4103/ija.IJA_341_17. | Design; Narrative review |
| Emmert MY, Salzberg SP, Theusinger OM, Felix C, Plass A, Hoerstrup SP, et al. How good patient blood management leads to excellent outcomes in Jehovah's witness patients undergoing cardiac surgery. Interactive Cardiovascular and Thoracic Surgery. 2011;12(2):183-8. doi: 10.1510/icvts.2010.242552. | Design; Uncontrolled study |
| Enko D, Wallner F, Von-Goedecke A, Hirschmugl C, Auersperg V, Halwachs-Baumann G. The Impact of an Algorithm-Guided Management of Preoperative Anemia in Perioperative Hemoglobin Level and Transfusion of Major Orthopedic Surgery Patients. Anemia. 2013. doi: 10.1155/2013/641876. | Outcome; nothing on adverse events |
| Eramishantsev AK, Vinnitsky LI, Gordeev PS, Ischanov MA. Perioperative correction of hypochromic anemia and iron metabolism disturbances in patients with liver cirrhosis and portal hypertension. Anesteziologiya i Reanimatologiya. 1992;(1):13-7. | Intervention |
| Espallardo CD, Morales MJL, Calvet CC, Lopez LM, Martinez IR, Marin MTM. The multidisciplinary approach is useful for optimising preoperative haemoglobin in colorectal cancer surgery. Cirugia Espanola. 2011;89(6):392-9. doi: 10.1016/j.ciresp.2011.01.013. | Design; Uncontrolled study |
| EUCTR2005-001412-50-ES. Randomised, parallel clinical trial, to compare efficacy and safety of sucrose intravenous iron versus oral ferrous sulfate for the treatment of perioperative lack of iron in patients with colorrectal neoplasia and forropenic anaemia. E.C aleatorizado, con grupos paralelos, comparativo de eficacia y seguridad con hierro sacarosa endovenoso vs. sulfato ferroso vía oral para el tratamiento del déficit perioperatorio de hierro en pacientes con neoplasia colorrectal y anemia ferropénica. In: amp, Compañía SA, editors. 2006. | Other; Duplicate of NCT00199277 (same sponsor study ID: DM01VEN/4/03) |
| EUCTR2005-003608-13-GB. A Prospective Double-blind Placebo Controlled Randomised Trial of Intravenous Iron Supplementation in Patients Undergoing Colorectal Cancer Surgery - RCT IV iron in colorectal cancer patients. 2005. | Outcome; nothing on adverse events |
| EUCTR2012-002786-35-GB. PREVENTT (Preoperative intravenous iron to treat anaemia in major surgery). 2012. | Other; study protocol already included (Richards 2015) |
| EUCTR2017-002972-15-GR. Comparison of the efficacy of anew oral formulation of iron aspartylate to oral ferrous sulfate in patients with Iron Deficiency Anemia for the restoration of low hemoglobin. 2017. | Population; no elective surgery patients |
| EUCTR2017-003439-12-DE. Clinical trial to demonstrate safety and efficacy of an intravenous (i.v.) administration of Feramyl compared to i.v. Ferinject and to iron tablets in patients with iron deficiency diganosis before a planned operation who develop anaemia during or after surgery. In: Applied E, editor. 2017. | Population |
| Faris P. Use of recombinant human erythropoietin in the perioperative period of orthopedic surgery. American Journal of Medicine. 1996;101((2A):):28S-32S. doi: 10.1016/s0002-9343(96)00164-7. | Other; duplicate |
| Faris PM, Ritter MA. Epoetin alfa: A bloodless approach for the treatment of perioperative anemia. Clinical Orthopaedics and Related Research. 1998;(357):60-7. | Design |
| Faris PM, Ritter MA. Epoetin alfa - A bloodless approach for the treatment of perioperative anemia. Clinical Orthopaedics and Related Research. 1998;(357):60-7. | Other; duplicate |
| Faris PM, Ritter MA, Abels RI, Ball GV, Bernini PM, Bryant GL, et al. The effects of recombinant human erythropoietin on perioperative transfusion requirements in patients having a major orthopaedic operation. Journal of Bone and Joint Surgery - Series A. 1996;78(1):62-72. | Population |
| Feagan BG, Wong CJ, Kirkley A, Johnston DWC, Smith FC, Whitsitt P, et al. Erythropoietin with iron supplementation to prevent allogeneic blood transfusion in total hip joint arthroplasty - A randomized, controlled trial. Annals of Internal Medicine. 2000;133(11):845-54. doi: 10.7326/0003-4819-133-11-200012050-00008. | Population |
| Feng S, Greenberg J, Moloo H, Thavorn K, McIsaac DI. Hospital cost associated with anemia in elective colorectal surgery: a historical cohort study. Canadian Journal of Anesthesia-Journal Canadien D Anesthesie. 2019;66(8):877-85. doi: 10.1007/s12630-019-01379-8. | Population; Compares costs in anemic vs non-anemic patients. Nothing on treated anemic vs non-treated anemic patients. |
| Ferrari P, Nicolini A, Manca ML, Rossi G, Anselmi L, Conte M, et al. Treatment of mild non-chemotherapy-induced iron deficiency anemia in cancer patients: comparison between oral ferrous bisglycinate chelate and ferrous sulfate. Biomedicine & Pharmacotherapy. 2012;66((6):):414-8. doi: 10.1016/j.biopha.2012.06.003. | Population |
| Franzini C, Pera E, Casali L, Rollo A, Santi C, Tarantino G. Perioperative sucrosomial® iron supplementation in gastrointestinal cancer surgery. Blood Transfusion. 2019;17:s8. doi: 10.2450/2019.S1. | Design; conference abstract |
| Fred Hutchinson Cancer R, Center, National C, Institute. Deferasirox in Treating Patients With Very Low, Low, or Intermediate-Risk Red Blood Cell Transfusion Dependent Anemia or Myelodysplastic Syndrome. | Design; Uncontrolled study |
| Freedman J, Luke K, Escobar M, Vernich L, Chiavetta JA. Experience of a network of transfusion coordinators for blood conservation (Ontario Transfusion Coordinators [ONTraC]). Transfusion. 2008;48(2):237-50. doi: 10.1111/j.1537-2995.2007.01515.x. | Design; Uncontrolled study |
| Fregonesi A, Ciari C, Melotti L, Zani EL, Ferreira U, Reis LO. Strategies for transfusion-free radical retropubic prostatectomy in Jehovah's witnesses. Actas Urologicas Espanolas. 2010;34(5):440-3. doi: 10.1016/j.acuro.2009.10.004. | Design; Uncontrolled study |
| Frew N, Alexander D, Hood J, Acornley A. Impact of a blood management protocol on transfusion rates and outcomes following total hip and knee arthroplasty. Annals of the Royal College of Surgeons of England. 2016;98(6):380-6. doi: 10.1308/rcsann.2016.0139. | Design; Uncontrolled study |
| Froessler B, Rueger AM, Connolly MP. Assessing the costs and benefits of perioperative iron deficiency anemia management with ferric carboxymaltose in Germany. Risk Manag Healthc Policy. 2018;11:77-82. doi: 10.2147/rmhp.s157379. | Design |
| Fuertinger DH, Kappel F, Thijssen S, Levin NW, Kotanko P. A model of erythropoiesis in adults with sufficient iron availability. J Math Biol. 2013;66(6):1209-40. doi: 10.1007/s00285-012-0530-0. | Design; Purely mathematical simulation study |
| Garcia-Erce JA, Cuenca J, Martinez F, Cardona R, Perez-Serrano L, Munoz M. Perioperative intravenous iron preserves iron stores and may hasten the recovery from post-operative anaemia after knee replacement surgery. Transfus Med. 2006;16(5):335-41. doi: 10.1111/j.1365-3148.2006.00682.x. | Population |
| Garcia Ruiz A, Ortega López AB, Morente Constantín E, García Sánchez MJ, Jurado Chacón M. Results of patient blood management (PBM) strategies in gynecological surgery in our center for 10 years. HemaSphere. 2020;4:1087. doi: 10.1097/HS9.0000000000000404. | Design; conference abstract |
| Garrido-Martin P, Nassar-Mansur MI, de la L-D, Virgos-Aller TM, Fortunez PM, Avalos-Pinto R, et al. The effect of intravenous and oral iron administration on perioperative anaemia and transfusion requirements in patients undergoing elective cardiac surgery: a randomized clinical trial. Interactive Cardiovascular and Thoracic Surgery. 2012;15((6):):1013-8. doi: 10.1093/icvts/ivs344. | Population |
| Gaston KE, Kouba E, Moore DT, Pruthi RS. The use of erythropoietin in patients undergoing radical prostatectomy: effects on hematocrit, transfusion rates and quality of life. Urol Int. 2006;77(3):211-5. doi: 10.1159/000094811. | Population |
| Gill KS, Antigua AD, Barnett AK, Hall AJ, Klodell CT. Evaluation of Erythropoietin Stimulating Agents (ESA) and Their Effect on Blood Optimization for Cardiac Surgery. Journal of pharmacy practice. 2020:897190020969274. Epub 2020/11/07. doi: 10.1177/0897190020969274. | Intervention; Compares patients receiving ESAs to patients receiving ESA+blood transfusion |
| Goldberg MA, McCutchen JW, Jove M, Di C, Friedman RJ, Poss R, et al. A safety and efficacy comparison study of two dosing regimens of epoetin alfa in patients undergoing major orthopedic surgery. Am J Orthop (Belle Mead NJ). 1996;25(8):544-52. | Comparison |
| Gómez-Ramírez S, Maldonado-Ruiz MÁ, Campos-Garrigues A, Herrera A, Muñoz M. Short-term perioperative iron in major orthopedic surgery: state of the art. Vox Sanguinis. 2019;114(1):3-16. doi: 10.1111/vox.12718. | Design; SR in which only PubMed was searched. Moreover, no additional relevant studies included. |
| Gómez-Sánchez A, Fuente-Alonso E. Management of iron deficiency anaemia by the preanaesthesia nurse with respect to perioperative transfusion rates. Enfermeria clinica. 2020;30(1):47-52. Epub 2019/04/23. doi: 10.1016/j.enfcli.2019.02.002. | Intervention; All patients received some form of IV iron. Not possible to distinguish groups receiving different types of IV iron or different doses of IV iron. |
| Gonzalez-Porras JR, Colado E, Conde MP, Lopez T, Nieto MJ, Corral M. An individualized pre-operative blood saving protocol can increase pre-operative haemoglobin levels and reduce the need for transfusion in elective total hip or knee arthroplasty. Transfusion Medicine. 2009;19(1):35-42. doi: 10.1111/j.1365-3148.2009.00908.x. | Population; Matched historic control group is not anemic (mean Hb 13.8± 1.4). |
| Grant MC, Lester L, Cho BC. In Response. Anesthesia and Analgesia. 2019;129(3):E111. doi: 10.1213/ANE.0000000000004297. | Design; letter |
| Green D, Lawler M, Rosen M, Bloom S, Duerden M, Turba R, et al. Recombinant human erythropoietin: effect on the functional performance of anemic orthopedic patients. Archives of Physical Medicine & Rehabilitation. 1996;77((3):):242-6. doi: 10.1016/s0003-9993(96)90105-7. | Population |
| Green WS, Toy P, Bozic KJ. Cost minimization analysis of preoperative erythropoietin vs autologous and allogeneic blood donation in total joint arthroplasty. J Arthroplasty. 2010;25(1):93-6. doi: 10.1016/j.arth.2008.10.005. | Design |
| Gromova VV, Imaev AA, Lubnin AI. [Use of recombinant human erythropoietin preparations as a blood-saving method in neurosurgery]. Anesteziologiia i reanimatologiia. 2010;(4):19-23. | Design; Uncontrolled study |
| Gross I, Seifert B, Hofmann A, Spahn DR. Patient blood management in cardiac surgery results in fewer transfusions and better outcome. Transfusion. 2015;55(5):1075-81. doi: 10.1111/trf.12946. | Population; Study participants only partially preoperative anemic (38% and 41%) |
| Guan XZ, Wang LL, Pan X, Liu L, Sun XL, Zhang XJ, et al. Clinical Indications of Recombinant Human Erythropoietin in a Single Center: A 10-Year Retrospective Study. Frontiers in Pharmacology. 2020;11. doi: 10.3389/fphar.2020.01110. | Intervention; Only looked at all patients who received EPO over a 10-year course. No comparison with no-EPO use. |
| Guarino S, Di M, Sorrenti S, Greco R, Nardi M, Favoriti P, et al. Bloodless surgery in geriatric surgery. International Journal of Surgery. 2014;12:S82-S5. doi: 10.1016/j.ijsu.2014.08.374. | Design; Purely descriptive study |
| Guinn NR, Guercio JR, Hopkins TJ, Grimsley A, Kurian DJ, Jimenez MI, et al. How do we develop and implement a preoperative anemia clinic designed to improve perioperative outcomes and reduce cost? Transfusion. 2016;56(2):297-303. doi: 10.1111/trf.13426. | Design; Descriptive overview of how a preoperative anemia clinic was developed and implemented; no real data on effectiveness included. |
| Gupta S, Panchal P, Gilotra K, Wilfred AM, Hou W, Siegal D, et al. Intravenous iron therapy for patients with preoperative iron deficiency or anaemia undergoing cardiac surgery reduces blood transfusions: a systematic review and meta-analysis. Interact Cardiovasc Thorac Surg. 2020;31(2):141-51. Epub 2020/07/10. doi: 10.1093/icvts/ivaa094. | Other; all relevant individual studies covered by our review. |
| Hafeez S, Mehboob S, Ashfaq B. Study to Determine the Safety and Efficacy of Iron Sucrose Given Intravenously in Anemic Women. Indo American Journal of Pharmaceutical Sciences. 2018;5(9):8581-5. doi: 10.5281/zenodo.1419574. | Design; Uncontrolled study |
| Hajjar LA, Vincent JL, Galas FR, Nakamura RE, Silva CM, Santos MH, et al. Transfusion requirements after cardiac surgery: the TRACS randomized controlled trial. JAMA. 2010;304(14):1559-67. Epub 2010/10/14. doi: 10.1001/jama.2010.1446. | Population; critically ill patients |
| Hamilton W, Coleman MG, Rubin G. Colorectal cancer. BMJ (Online). 2013;347(7924). doi: 10.1136/bmj.f3172. | Design; Narrative |
| Hardesty DA, Doerfler S, Sandhu S, Whitmore RG, Ford P, Rushton S, et al. “Bloodless” Neurosurgery Among Jehovah's Witnesses: A Comparison with Matched Concurrent Controls. World Neurosurgery. 2017;97:132-9. doi: 10.1016/j.wneu.2016.09.028. | Population; Only a small portion of "intervention" group is anaemic (5 of the 68 Jehova patients; 4 received EPO treatment, 1 received iron supplementation alone). No information on Hb status or treatment of "control" group (i.e. matched non-Jehova patients). |
| Harwin SF, Issa K, Naziri Q, Johnson AJ, Mont MA. Results of Primary Total Knee Arthroplasty in Jehovah's Witness Patients. Journal of Arthroplasty. 2013;28(1):49-55. doi: 10.1016/j.arth.2012.05.021. | Population; Unclear reporting of baseline Hemoglobin levels; unclear if this involves anaemic people. |
| Harwin SF, Pivec R, Johnson AJ, Naziri Q, Mont MA. Revision Total Hip Arthroplasty in Jehovah's Witnesses. Orthopedics. 2012;35(8):E1145-E51. doi: 10.3928/01477447-20120725-11. | Population; No clear description of baseline Hb levels in Jehova witness patients. Unclear who received which type of intervention. |
| Harwin SF, Pivec R, Naziri Q, Issa K, Mont MA. Is total hip arthroplasty a successful and safe procedure in Jehovah's Witnesses? Mean five-year results. Hip International. 2014;24(1):69-76. doi: 10.5301/hipint.5000106. | Intervention |
| Heschl M, Gombotz H, Haslinger-Eisterer B, Hofmann A, Bohler N, Meier J. The efficacy of pre-operative preparation with intravenous iron and/or erythropoietin in anaemic patients undergoing orthopaedic surgery: An observational study. European Journal of Anaesthesiology. 2018;35(4):289-97. doi: 10.1097/EJA.0000000000000752. | Outcome; nothing on adverse events |
| Hönemann C, Bierbaum M, Heidler J, Doll D, Schöffski O. Costs of delivering allogenic blood in hospitals. Chirurg. 2013;84(5):426-32. doi: 10.1007/s00104-012-2464-x. | Design; Economic analysis |
| Hourlier H, Fennema P. Surgical prescription of epoetin alfa in contemporary total hip arthroplasty: a prospective comparative study. Int Orthop. 2020;44(2):261-6. Epub 2019/08/31. doi: 10.1007/s00264-019-04399-7. | Design; Only one person in the group receiving EPO + IV iron. Not possible to compare to group receiving EPO + oral iron. |
| Ionescu A, Sharma A, Kundnani NR, Mihăilescu A, David VL, Bedreag O, et al. Intravenous iron infusion as an alternative to minimize blood transfusion in peri-operative patients. Scientific reports. 2020;10(1):18403. Epub 2020/10/29. doi: 10.1038/s41598-020-75535-2. | Outcome; Contacted authors twice to obtain information on possible collection of adverse events. No response obtained. |
| ISRCTN22158788. Can administration of iron supplement injection reduce blood transfusion rates in people with a low red blood cell count (anaemia) who are undergoing heart operation? 2013. | Other |
| Jafari-Fesharaki M, Toy P. Effect and cost of subcutaneous recombinant human erythropoietin in preoperative patients. Orthopedics. 1997;20(12):1159-67. | Design |
| Jahn MR, Andreasen HB, Fütterer S, Nawroth T, Schünemann V, Kolb U, et al. A comparative study of the physicochemical properties of iron isomaltoside 1000 (Monofer®), a new intravenous iron preparation and its clinical implications. European Journal of Pharmaceutics and Biopharmaceutics. 2011;78(3):480-91. doi: 10.1016/j.ejpb.2011.03.016. | Design; Comparative study of properties of polynuclear iron formulations (particle size, size and structure of the core, ferrous content). |
| Jans O, Nielsen CS, Khan N, Gromov K, Troelsen A, Husted H. Iron deficiency and preoperative anaemia in patients scheduled for elective hip- and knee arthroplasty - an observational study. Vox Sang. 2018;113(3):260-7. Epub 2018/02/07. doi: 10.1111/vox.12630. | Intervention; no iron/EPO |
| Janssen TL, Steyerberg EW, van Gammeren AJ, Ho GH, Gobardhan PD, van der Laan L. Intravenous Iron in a Prehabilitation Program for Older Surgical Patients: Prospective Cohort Study. Journal of Surgical Research. 2021;257:32-41. doi: 10.1016/j.jss.2020.07.059. | Intervention; All anaemic patients received IV iron, all non-anaemic patients received standard care |
| Janssens M, Lamy M. [Role of recombinant erythropoietin during the preoperative period]. Ann Fr Anesth Reanim. 1995;14 Suppl 1:98-106. | Design; Narrative review |
| Johnson, Johnson P, Research, Development LLC, Ortho Biotech P, L P. Study of the Efficacy and Safety of Epoetin Alfa Administered Weekly in Patients With Gastric or Rectal Cancers Undergoing a Treatment Plan of Preoperative Chemotherapy and Radiation Therapy, Followed by Surgery. | Other |
| Joseph SA, Berekashvili K, Mariller MM, Rivlin M, Sharma K, Casden A, et al. Blood conservation techniques in spinal deformity surgery: A retrospective review of patients refusing blood transfusion. Spine. 2008;33(21):2310-5. doi: 10.1097/BRS.0b013e31818047f2. | Population; No clear reporting of baseline Hb levels of participants; probably both anaemic and non-anaemic patients. |
| Jung R, Yoo JJ, Tan R, Kim S. PSU8 Health Economic Evaluation of Protocol Change Including IV Iron Ferric Carboxymaltose Versus Control Group for HIP Surgery Patients with Iron Deficiency Anemia (IDA) in Korea. Value in Health Regional Issues. 2020;22:S105. doi: 10.1016/j.vhri.2020.07.550. | Design; conference abstract |
| Juraszek A, Kolsut P, Szymanski J, Kuriata J, Kusmierski K, Sitkowska-Rysiak E, et al. Results of open heart surgery in Jehovah's Witness patients. Single centre experience. Kardiochirurgia I Torakochirurgia Polska. 2017;14(3):164-9. doi: 10.5114/kitp.2017.70529. | Design; Uncontrolled study |
| Kang T, Park SY, Nam JJ, Lee SH, Park JH, Suh SW. Patient Mood Management During Lumbar Spinal Fusion Surgery. World Neurosurgery. 2019;130:E566-E72. doi: 10.1016/j.wneu.2019.06.153. | Intervention; PBM protocol = use of IV iron supplementation + introduction of restrictive transfusion trigger (Hb<7 g/dl) + use of tranexamic acid. |
| Karkouti K, McCluskey SA, Evans L, Mahomed N, Ghannam M, Davey R. Erythropoietin is an effective clinical modality for reducing RBC transfusion in joint surgery. Canadian Journal of Anaesthesia-Journal Canadien D Anesthesie. 2005;52(4):362-8. | Outcome; nothing on adverse events |
| Kato M, Sawada T, Kita J, Shimoda M, Kubota K. Erythropoietin ameliorates early ischemia-reperfusion injury following the Pringle maneuver. World J Gastroenterol. 2010;16(38):4838-45. | Population; No information on preoperative Hb levels, because study focuses on other non-haematopoietic properties of EPO; hence, probably mixture of non-anaemic and anaemic participants. |
| Kaufner L, Heymann C, Henkelmann A, Pace NL, Weibel S, Kranke P, et al. Erythropoietin plus iron versus control treatment including placebo or iron for preoperative anaemic adults undergoing non‐cardiac surgery. Cochrane Database of Systematic Reviews. 2020;(8). doi: 10.1002/14651858.CD012451.pub2. | Other; all relevant individual studies covered by our review. |
| Kearney B, To J, Southam K, Howie D, To B. Anaemia in elective orthopaedic surgery - Royal Adelaide Hospital, Australia. Intern Med J. 2016;46(1):96-101. doi: 10.1111/imj.12945. | Design; Purely descriptive study |
| Keeler BD, Simpson JA, Ng S, Tselepis C, Iqbal T, Brookes MJ, et al. The feasibility and clinical efficacy of intravenous iron administration for preoperative anaemia in patients with colorectal cancer. Colorectal disease : the official journal of the Association of Coloproctology of Great Britain and Ireland. 2014;16(10):794-800. Epub 2014/06/12. doi: 10.1111/codi.12683 | Design; Uncontrolled study |
| Keeler BD, Mishra A, Stavrou CL, Beeby S, Simpson JA, Acheson AG. A cohort investigation of anaemia, treatment and the use of allogeneic blood transfusion in colorectal cancer surgery. Annals of Medicine and Surgery. 2016;6:6-11. doi: 10.1016/j.amsu.2015.12.052. | Outcome; nothing on adverse events |
| Kei T, Mistry N, Curley G, Pavenski K, Shehata N, Tanzini RM, et al. Efficacy and safety of erythropoietin and iron therapy to reduce red blood cell transfusion in surgical patients: a systematic review and meta-analysis. Canadian journal of anaesthesia = Journal canadien d'anesthesie. 2019;66(6):716-31. Epub 2019/03/30. doi: 10.1007/s12630-019-01351-6. | Other; all relevant individual studies covered by our review. |
| Kepler U, Hospital. Oral Iron Substitution for Orthopedic Surgery. 2017. | Outcome; nothing on adverse events |
| Kim JE, Song SW, Kim JY, Lee HJ, Chung KH, Shim YH. Effect of a Single Bolus of Erythropoietin on Renoprotection in Patients Undergoing Thoracic Aortic Surgery With Moderate Hypothermic Circulatory Arrest. Annals of Thoracic Surgery. 2016;101(2):690-6. doi: 10.1016/j.athoracsur.2015.08.007. | Population |
| Kim JH, Shim JK, Song JW, Song Y, Kim HB, Kwak YL. Effect of erythropoietin on the incidence of acute kidney injury following complex valvular heart surgery: a double blind, randomized clinical trial of efficacy and safety. Crit Care. 2013;17(5):R254. doi: 10.1186/cc13081. | Population |
| Kleinerüschkamp AG, Zacharowski K, Ettwein C, Müller MM, Geisen C, Weber CF, et al. Cost analysis of patient blood management. Anaesthesist. 2016;65(6):438-48. doi: 10.1007/s00101-016-0152-9. | Design; Economic analysis |
| Koo CH, Shin HJ, Cho H, Ryu JH. The Effect of Perioperative Intravenous Iron on Hemoglobin in Surgical Patients: A Meta-Analysis. J Surg Res. 2020;246:42-51. Epub 2019/09/29. doi: 10.1016/j.jss.2019.08.023. | Other; all relevant individual studies covered by our review. |
| Kopanidis P, Hardidge A, McNicol L, Tay S, McCall P, Weinberg L. Perioperative blood management programme reduces the use of allogenic blood transfusion in patients undergoing total hip and knee arthroplasty. J Orthop Surg Res. 2016;11:28. doi: 10.1186/s13018-016-0358-1. | Intervention; Multicomponent intervention, including intraoperative tranexamic acid in intervention group, but not in control group. |
| Kotzé A, Carter LA, Scally AJ. Effect of a patient blood management programme on preoperative anaemia, transfusion rate, and outcome after primary hip or knee arthroplasty: A quality improvement cycle. British Journal of Anaesthesia. 2012;108(6):943-52. doi: 10.1093/bja/aes135. | Population; Both cohorts (before or after implementation of a PBM programme) contain both anaemic (Hb <12 g/dl for women and Hb <13g/dl for men) and non-anaemic people. In the comparison between the 2 cohorts, there is no subgroup analysis for anaemic people. In the post-implementation cohort, no subgroup analysis was performed for the treatment they were given. |
| Kurian DJ, Guinn NR, Hunting J, Gamble JF, Hopkins TJ, Grimsley A, et al. Preoperative Blood Management Strategy for Elective Hip and Knee Arthroplasty. Journal for healthcare quality : official publication of the National Association for Healthcare Quality. 2019;41(6):376-83. Epub 2019/06/19. doi: 10.1097/jhq.0000000000000207. | Outcome; Did not look at adverse events. |
| Kwon HY, Kim BR, Kim YW. Association of preoperative anemia and perioperative allogenic red blood cell transfusion with oncologic outcomes in patients with nonmetastatic colorectal cancer. Curr Oncol. 2019;26(3):e357-e66. Epub 2019/07/10. doi: 10.3747/co.26.4983. | Intervention; no iron/EPO |
| Kyo S, Omoto R, Hirashima K, Eguchi S, Fujita T. Effect of human recombinant erythropoietin on reduction of homologous blood transfusion in open-heart surgery. A Japanese Multicenter study. Circulation. 1992;86(5 Suppl):Ii413-8. | Population; Controls are non-anaemic (mean baseline Hb 13.8 +/- 2 g/dl) |
| Lachance K, Savoie M, Bernard M, Rochon S, Fafard J, Robitaille R, et al. Oral ferrous sulfate does not increase preoperative hemoglobin in patients scheduled for hip or knee arthroplasty. Ann Pharmacother. 2011;45(6):764-70. doi: 10.1345/aph.1P757. | Design |
| Laffosse JM, Minville V, Chiron P, Colombani A, Gris C, Pourrut JC, et al. Preoperative use of epoietin beta in total hip replacement: a prospective study. Arch Orthop Trauma Surg. 2010;130(1):41-5. doi: 10.1007/s00402-009-0863-3. | Outcome; nothing on adverse events |
| Lagos S, University, Health Forever Product Limited L, Nigeria. The Safety and Effectiveness of Jobelyn in Pre-operative Management of Anaemia in Gynaecological Patients. 2013. | Intervention |
| Lakkawar NJ, Sankaran S, Rangaswamy T. Efficacy of intravenous administration of iron sucrose for treatment of iron deficiency anaemia in patients with abnormal uterine bleeding. Acta Facultatis Medicae Naissensis. 2012;29(2):59-68. doi: 10.2478/v10283-012-0009-3. | Design; Uncontrolled study |
| Laman CA, Silverstein SB, Rodgers GM. Parenteral iron therapy: A single institution's experience over a 5-year period. JNCCN Journal of the National Comprehensive Cancer Network. 2005;3(6):791-5. | Design |
| Laupacis A, Monk, Messmer, McClelland, Beris. Effectiveness of perioperative epoetin alfa in patients scheduled for elective hip surgery. Seminars in Hematology. 1996;33(2 SUPPL. 2):51-4. | Design |
| Leahy MF, Roberts H, Mukhtar SA, Farmer S, Tovey J, Jewlachow V, et al. A pragmatic approach to embedding patient blood management in a tertiary hospital. Transfusion. 2014;54(4):1133-45. doi: 10.1111/trf.12362. | Population; Compares blood transfusion rates before and after implementation of a PBM programme. However, concerns both preoperative and postoperative anemia. |
| Lee BW, Park MG, Cho DY, Park SS, Yeo JK. Preoperative erythropoietin administration in patients with prostate cancer undergoing radical prostatectomy without transfusion. Korean J Urol. 2014;55(2):102-5. doi: 10.4111/kju.2014.55.2.102. | Intervention |
| Lee ES, Kim MJ, Park BR, Kim JS, Choi GY, Lee JJ, et al. Avoiding unnecessary blood transfusions in women with profound anaemia. Australian & New Zealand Journal of Obstetrics & Gynaecology. 2015;55(3):262-7. doi: 10.1111/ajo.12329. | Population; no elective surgery patients |
| Lee SH, Kim JI, Choi W, Kim TW, Lee YS. Effectiveness of iron supplementation in the perioperative management of total knee arthroplasty: a systematic review. Knee surgery & related research. 2020;32(1):44. Epub 2020/08/30. doi: 10.1186/s43019-020-00064-1. | Other; all relevant individual studies covered by our review. |
| Lee B, Kim EJ, Song J, Jung YS, Koo BN. A randomised trial evaluating the effect of intraoperative iron administration. Scientific reports. 2020;10(1):15853. Epub 2020/09/29. doi: 10.1038/s41598-020-72827-5. | Intervention; intra-operative administration of iron |
| Lentschener C, Gomola A, Grabar S, Soubrane O, Dousset B, Massault PP, et al. The effect of erythropoietin on allogeneic blood requirement in patients undergoing elective liver resection: a model simulation. Anesth Analg. 2004;98(4):921-6. | Design; Mathematical modeling study |
| Levine EA, Laborde C, Hambrick E, McKnight CA, Vijayakumar S. Influence of erythropoietin on transfusion requirements in patients receiving preoperative chemoradiotherapy for rectal cancer. Dis Colon Rectum. 1999;42(8):1065-9. | Intervention |
| Li RP, Xue FS, Liu GP, Sun C. Association of Preoperative Anemia With Complications and Mortality Following Total Joint Arthroplasty. Journal of Arthroplasty. 2015;30(11):2043-4. doi: 10.1016/j.arth.2015.06.065. | Design; Letter to the editor |
| Lidder PG, Sanders G, Whitehead E, Douie WJ, Mellor N, Lewis SJ, et al. Pre-operative oral iron supplementation reduces blood transfusion in colorectal surgery - a prospective, randomised, controlled trial. Annals of the Royal College of Surgeons of England. 2007;89((4):):418-21. doi: 10.1308/003588407x183364. | Outcome; nothing on adverse events |
| Lim C, Salloum C, Esposito F, Giakoustidis A, Moussallem T, Osseis M, et al. Safety and feasibility of elective liver resection in adult Jehovah's Witnesses: the Henri Mondor Hospital experience. Hpb. 2018;20(9):823-8. doi: 10.1016/j.hpb.2018.02.642. | Design; Case series |
| Lin Y, Howell A, Vernich L, Pavenski K, Freedman J. Oral iron versus intravenous iron for preoperative anemia management: the ONTraC experience. Transfusion Medicine Reviews. 2020;34(1):64. doi: 10.1016/j.tmrv.2019.11.004. | Design; conference abstract |
| Lofthouse RA, Boitano MA, Davis JR, Jinnah RH. Preoperative administration of epoetin alfa to reduce transfusion requirements in elderly patients having primary total hip or knee reconstruction. J South Orthop Assoc. 2000;9(3):175-81. | Outcome; nothing on adverse events |
| Lopez Soques, M M, Leon A, Garcia A, Garces P, Saez M. [Benefit of a blood conservation program in elective orthopaedic surgery]. Med Clin (Barc). 2002;119(17):650-2. | Population; Intervention is based on baseline Hb levels: Hb = or < 13: EPO+iron Hb > 13: autologous donation + iron --> therefore, control group is non-anaemic |
| Loughnane F, Pollock R, Muduma G, Cook L. PSY6 A RESOURCE IMPACT ANALYSIS OF IRON ISOMALTOSIDE IN THE TREATMENT OF IRON DEFICIENCY ANEMIA IN IRELAND. Value in Health. 2019;22:S375. doi: 10.1016/j.jval.2019.04.1831. | Design; conference abstract |
| Lowry B, Hardy K, Vergis A. Iron deficiency in bariatric surgery patients: a single-centre experience over 5 years. Canadian journal of surgery Journal canadien de chirurgie. 2020;63(4):E365-e9. Epub 2020/08/20. doi: 10.1503/cjs.001818. | Population; Only a subset of patients were anemic before the operation. No separate data on these anemic patients. |
| Luo CT, Shi YQ, Lin Y, Ma RH, Xia Q, Ding WJ. Intravenous transfusion of iron sucrose reduces blood transfusions and improves postoperative anaemia after a second thoracotomy: a propensity-score matching study. Journal of International Medical Research. 2020;48(2). doi: 10.1177/0300060520902912. | Population; Only a subset of patients were anemic before the operation. No separate data on these anemic patients. |
| Luporsi E, Mahi L, Morre C, Wernli J, de P, Bugat R. Evaluation of cost savings with ferric carboxymaltose in anemia treatment through its impact on erythropoiesis-stimulating agents and blood transfusion: French healthcare payer perspective. J Med Econ. 2012;15(2):225-32. doi: 10.3111/13696998.2011.639823. | Design |
| Mabry C, Perelman S, Kim JT, Blitz JD. Implementation of a Preoperative Anemia Clinic Utilizing a Minimal Staffing Model. A&A practice. 2020;14(3):90-4. Epub 2019/11/27. doi: 10.1213/xaa.0000000000001131. | Design; uncontrolled study |
| MacLaren R, Sullivan PW. Cost-effectiveness of recombinant human erythropoietin for reducing red blood cells transfusions in critically ill patients. Value in Health. 2005;8(2):105-16. doi: 10.1111/j.1524-4733.2005.04006.x. | Design |
| Magali S, A M, Pampillón N, Abaurre M, Omelanczuk PE. PRE-OPERATIVE IRON DEFICIENCY IN BARIATRIC SURGERY: DIAGNOSIS AND TREATMENT. Nutricion hospitalaria. 2015;32(1):75-9. doi: 10.3305/nh.2015.32.1.8871. | Population; Only 6% of patient population is anaemic, no subgroup analyses. |
| Mazzeffi M, Chow JH, Tanaka K. Preoperative Erythropoietin in Cardiac Surgery: Evolving Standard of Care or Aggregation of Marginal Gain? Anesthesia and Analgesia. 2019;129(3):E110-E1. doi: 10.1213/ANE.0000000000004296. | Design; letter |
| McSorley ST, Anderson JH, Whittle T, Roxburgh CS, Horgan PG, McMillan DC, et al. The impact of preoperative systemic inflammation on the efficacy of intravenous iron infusion to correct anaemia prior to surgery for colorectal cancer. Perioperative medicine (London, England). 2020;9:17. Epub 2020/06/17. doi: 10.1186/s13741-020-00146-4. | Intervention; No data on patients who had recevied oral iron supplementation, so not able to compare oral iron to IV iron |
| Meara JG, Smith EM, Harshbarger RJ, Farlo JN, Matar MM, Levy ML. Blood-conservation techniques in craniofacial surgery. Ann Plast Surg. 2005;54(5):525-9. | Population; Children (mean age 13.1 and 11.0 months) |
| Medert HA, Mahler S. Preoperative treatment with erythropoetin: dose-effect relationships for haemoglobin, hematocrit value and reticulocytes before and after surgery. Annals of hematology. 1997;74 Suppl 4:A179. | Design: Conference abstract |
| Meir M, Center. Iron and Vitamin Adminstration Prior to Joint Replacement to Prevent Transfusion. 2013. | Intervention; Intervention group receives iron, folic acid and vitamin B12. Control group does not receive folic acid and vitamin B12. Therefore, not possible to determine the effect of iron. |
| Meybohm P, Lindau S, Treskatsch S, Francis R, Spies C, Velten M, et al. Liberal transfusion strategy to prevent mortality and anaemia-associated, ischaemic events in elderly non-cardiac surgical patients - the study design of the LIBERAL-Trial. Trials. 2019;20(1):101. Epub 2019/02/06. doi: 10.1186/s13063-019-3200-3. | Population; patients with Hb <9 g/dl during and after surgery |
| Meybohm P, Straub N, Fullenbach C, Judd L, Kleineruschkamp A, Taeuber I, et al. Health economics of Patient Blood Management: a cost-benefit analysis based on a meta-analysis. Vox Sanguinis. 2020;115(2):182-8. doi: 10.1111/vox.12873. PubMed PMID: WOS:000501730600001. | Design; no original data, based on meta-analysis of Althoff |
| Mihaleva R, Tenchev P, Frangov P, Maskov P. Preoperative recombinant human erythropoietin in elective spine surgery. Anaesthesiology and Intensive Care. 2007;34(2):17-22. | Intervention |
| Milton SHMC. A Pilot Study Comparing Tolerance of Oral Heme Iron Polypeptide With Oral Ionic Iron. 2016. | Population; no elective surgery patients |
| Minoda Y, Sakawa A, Fukuoka S, Tada K, Takaoka K. Blood management for patients with hemoglobin level lower than 130 g/l in total knee arthroplasty. Arch Orthop Trauma Surg. 2004;124(5):317-9. doi: 10.1007/s00402-004-0647-8. | Intervention; Investigates the effect of hematinics, including epoetin beta and/or prednisolone for rheumatoid arthritis patients. No subgroup analysis on epoetin beta only. |
| Moonen AF, Thomassen BJ, Knoors NT, van O, J J, Verburg AD, et al. Pre-operative injections of epoetin-alpha versus post-operative retransfusion of autologous shed blood in total hip and knee replacement: a prospective randomised clinical trial. J Bone Joint Surg Br. 2008;90(8):1079-83. doi: 10.1302/0301-620x.90b8.20595. | Intervention |
| Muller H, Ratschiller T, Schimetta W, Meier J, Gombotz H, Zierer A. Open Heart Surgery in Jehovah's Witnesses: A Propensity Score Analysis. Annals of Thoracic Surgery. 2020;109(2):526-33. doi: 10.1016/j.athoracsur.2019.06.065. | Intervention; Not clear which treatments were given to non-Jehovah's witnesses, that serve as a control group. |
| Mundy GM, Birtwistle SJ, Power RA. The effect of iron supplementation on the level of haemoglobin after lower limb arthroplasty. J Bone Joint Surg Br. 2005;87(2):213-7. | Population |
| Munoz M, Garcia-Erce JA, Cuenca J, Bisbe E. Pharmacological management of perioperative anaemia: our experience with intravenous iron in orthopaedic surgery. Isbt Science Series, Vol 2, No 1: State of the Art Presentations. 2. Malden: Wiley-Blackwell; 2007. p. 257. | Design; no actual study, overview of results |
| Munoz M, Garcia-Erce JA, Cuenca J, Bisbe E, Naveira E. On the role of iron therapy for reducing allogeneic blood transfusion in orthopaedic surgery. Blood Transfusion. 2012;10(1):8-22. doi: 10.2450/2011.0061-11. | Design; Narrative review |
| Munoz M, Garcia-Erce JA, Diez-Lobo AI, Campos A, Sebastianes C, Bisbe E. Usefulness of the administration of intravenous iron sucrose for the correction of preoperative anemia in major surgery patients. Medicina Clinica. 2009;132(8):303-6. doi: 10.1016/j.medcli.2008.04.011. | Design; Uncontrolled study |
| Munoz M, Gomez-Ramirez S, Cuenca J, Garcia-Erce JA, Iglesias-Aparicio D, Haman-Alcober S, et al. Very-short-term perioperative intravenous iron administration and postoperative outcome in major orthopedic surgery: a pooled analysis of observational data from 2547 patients. Transfusion. 2014;54(2):289-99. doi: 10.1111/trf.12195. | Design |
| Myers E, Grady PO, Dolan AM. The influence of preclinical anaemia on outcome following total hip replacement. Archives of Orthopaedic and Trauma Surgery. 2004;124(10):699-701. doi: 10.1007/s00402-004-0754-6. | Outcome; nothing on adverse events |
| Na HS, Shin SY, Hwang JY, Jeon YT, Kim CS, Do SH. Effects of intravenous iron combined with low-dose recombinant human erythropoietin on transfusion requirements in iron-deficient patients undergoing bilateral total knee replacement arthroplasty. Transfusion. 2011;51((1):):118-24. doi: 10.1111/j.1537-2995.2010.02783.x. | Population |
| Na N, Hong LQ, Miao B, Hua XF, Huang ZY. Effect of pre-transplantation hemoglobin concentration on prognosis of renal transplant recipients. Chinese Medical Journal. 2011;124(8):1213-6. doi: 10.3760/cma.j.issn.0366-6999.2011.08.017. | Population |
| Nasir A, Yasmin H, Korejo R. Management of iron deficiency anaemia in gynaecological patients at Jinnah Postgraduate Medical Centre, Karachi. J Pak Med Assoc. 2011;61(10):998-1001. | Design; Uncontrolled study |
| National Cancer C, Korea. Ferinject® Assessment In GastRectomy Patients With Acute Isovolemic Anemia (FAIRY). 2016. | Population |
| National Cancer C, Korea, Pharmaceutical JW. Efficacy of Ferric Carboxymaltose (Ferinject®) in Anemic Patients Anticipating Pancreatoduodenectomy. 2018. | Design |
| NCT04141631. Transfusion Savings in Heart Surgery: impact of Individual Strategy by Erythropoietin and Metabolic Adjustment (ScvO2). https://clinicaltrialsgov/show/NCT04141631. 2019. | Intervention; Experimental group and control group do not receive the same co-interventions (i.e. postop iron sucrose administration in control group vs postop EPO + ferric carboxymaltose administration in intervention group). |
| NCT02862665. Effect of Perioperative Iron Isomaltoside 1000 Administration on Transfusion Requirements in Patients Undergoing Complex Valvular Heart Surgery: a Randomized Clinical Trial. Https://clinicaltrialsgov/show/nct02862665. 2016. | Population; probably no anaemic patients |
| NCT03888768. ProPBM : a Modified Patient Blood Management Protocol. https://clinicaltrialsgov/show/NCT03888768. 2019. | Intervention; Not the same co-interventions in both groups |
| NCT04035902. Intraoperative IV Iron on Postoperative Red Blood Cell Recovery. https://clinicaltrialsgov/show/NCT04035902. 2019. | Intervention; intra-operative administration of iron |
| NCT00270179. A Study to Evaluate the Effectiveness and Safety of Epoetin Alfa During the Period When One is Donating One's Own Blood Before Surgery. Https://clinicaltrialsgov/show/nct00270179. 2005. | Population |
| NCT00269958. A Study to Evaluate the Safety and Efficacy of Epoetin Alfa Versus Placebo to Reduce the Need for Blood Transfusions and Reduce the Occurrence of Severe Anemia During the Time Period Surrounding Total Hip Replacement Surgery. Https://clinicaltrialsgov/show/nct00269958. 2005. | Population |
| NCT01265680. Blood Sparing Strategies: single Shot High Dose Erythropoietin Two Days Before Heart Surgery. Https://clinicaltrialsgov/show/nct01265680. 2010. | Population; Nothing on anaemia. |
| NCT01912261. Effects of Oral Iron on Postoperative Fatigue Upon Coronary Artery Bypass Graft Patients. Https://clinicaltrialsgov/show/nct01912261. 2013. | Population |
| NCT02862665. Effect of Perioperative Iron Isomaltoside 1000 Administration on Transfusion Requirements in Patients Undergoing Complex Valvular Heart Surgery: a Randomized Clinical Trial. Https://clinicaltrialsgov/show/nct02862665. 2016. | Population |
| NCT02800746. Intravenous Iron Pre-treatment in Prognathic Surgery. Https://clinicaltrialsgov/show/nct02800746. 2016. | Population |
| NCT03094182. Efficacy of Intravenous Iron Therapy in Maintaining Hemoglobin Concentration on Patients Undergoing Bimaxillary Orthognathic Surgery. Https://clinicaltrialsgov/show/nct03094182. 2017. | Population |
| NCT00270062. A Study to Evaluate the Safety of Epoetin Alfa and Its Effectiveness in Facilitating the Presurgical Collection of Blood From Anemic Patients for Possible Self-transfusion During and After Scheduled Joint Surgery. Https://clinicaltrialsgov/show/nct00270062. 2005. | Intervention |
| NCT00270140. A Study to Evaluate the Safety and Effectiveness of Epoetin Alfa Versus Placebo in Facilitating the Presurgical Collection of Blood to be Used for Self-donation During Surgery on the Spine, Hip, or Knee. Https://clinicaltrialsgov/show/nct00270140. 2005. | Intervention |
| NCT02031289. Impact of Preoperative Treatment of Anemia and Iron Deficiency in Cardiac Surgery on Outcome. https://clinicaltrials.gov/ct2/show/NCT02031289. 2013. | Comparison; Intervention group receives a combination treatment of iron, EPO, vitamin B12 and folic acid. Control group does not receive vitamin B12 and folic acid. Hence, not possible to extract the effect of iron+EPO. |
| NCT00254436. A Double-Blind, Randomized, Placebo-Controlled Study of the Efficacy and Safety of Weekly Procrit Given to Gastric or Rectal Patients. Https://clinicaltrialsgov/show/nct00254436. 2005. | Outcome; nothing on adverse events |
| NCT02802592. EPOgen and Restrictive Transfusions in Patients Undergoing Cardiac Surgery (EPORT). Https://clinicaltrialsgov/show/nct02802592. 2016. | Outcome; nothing on adverse events |
| NCT01692418. Preoperative Intravenous Iron to Treat Anaemia in Major Surgery. Https://clinicaltrials.gov/ct2/show/NCT01692418. 2014. | Other; Duplicate of PREVENTT trial by Richards 2015 (already included). |
| NCT00083434. Treatment of Anemic Patients With Cancer Who Are Not Receiving Chemotherapy or Radiotherapy. In: amp, Johnson P, Research, amp, Development LLC, editors. 2004. | Population; no elective surgery patients |
| NCT00083486. Treatment of Anemia in Patients With Cancer Who Are Not Currently Receiving Chemotherapy or Radiotherapy. In: amp, Johnson P, Research, amp, Development LLC, editors. 2004. | Population; no elective surgery patients |
| NCT00199277. Iron Therapy in Colo-Rectal Neoplasm and Iron Deficiency Anemia: Intravenous Iron Sucrose Versus Oral Ferrous Sulphate. In: Company, editor. 2005. | Other; Duplicate |
| NCT00211146. A Study to Confirm the Safety and Efficacy of Epoetin Alfa (PROCRIT) Administered Perioperatively vs. the Standard of Care in Blood Conservation in Patients Undergoing Major Elective Spinal Surgery (SPINE Study). In: amp, Johnson P, Research, amp, Development LLC, Ortho B, et al., editors. 2005. | Other; full text paper already included (Stowell 2009) |
| NCT00236405. PROCRIT and Short-Term Outcomes in Orthopedic Surgery. In: amp, Johnson P, Research, amp, Development LLC, Ortho Biotech P, et al., editors. 2005. | Comparison |
| NCT00269958. A Study to Evaluate the Safety and Efficacy of Epoetin Alfa Versus Placebo to Reduce the Need for Blood Transfusions and Reduce the Occurrence of Severe Anemia During the Time Period Surrounding Total Hip Replacement Surgery. 2005. | Other |
| NCT00269971. A Study to Determine an Effective Dose of Epoetin Alfa to Decrease the Number of Units of Blood Required to be Transfused During Hip Replacement Surgery. 2005. | Other; full text paper already included (Feagan 2000) |
| NCT00270036. A Study to Determine the Safety of Epoetin Alfa and Whether Epoetin Alfa Can Reduce the Need for Blood Transfusions in Patients After Major Orthopedic Surgery. In: amp, Johnson P, Research, amp, Development LLC, editors. 2005. | Other |
| NCT00270088. A Study to Determine Whether Epoetin Alfa Can Reduce the Need for Blood Transfusions in Patients During the Period of Time Around Major Orthopedic Surgery. In: amp, Johnson P, Research, amp, Development LLC, editors. 2005. | Other |
| NCT00593619. Trial Comparing the Safety of Two Different Intravenous Iron Formulations. 2008. | Population; Consists of people who received iron for different indications: the most frequent indication for intravenous iron therapy was pre-operative iron supplementation in 117/143 (81.8 %), but there were also people who received iron because of bleeding or malignancy. No separate subgroup-analyses. |
| NCT01012063. Perioperative Iron With Erythropoietin in Bilateral Total Knee Replacement Arthroplasty (TKRA). 2009. | Other |
| NCT01888003. The Benefits of a Preoperative Anemia Management Program. 2013. | Outcome; nothing on adverse events |
| NCT02172001. A Randomized, Double-blind, Comparative Study of Intravenous Iron Isomaltoside 1000 (Monofer®) Against Placebo. 2014. | Outcome; nothing on adverse events |
| NCT02210949. Pre-operative Treatment With Erythropoietin and Iron Supplement in Cardiac Surgery. 2014. | Outcome; nothing on adverse events |
| NCT02390102. Erythropoietin + Iron Therapy for Anemic Patients Undergoing Aortic Valve Replacement. 2013. | Other |
| NCT02496377. Cross Iron (Comparative Randomized Oral Versus Systemic IRON). 2015. | Other; full text already included (Biboulet 2018) |
| NCT02632760. Intravenous Iron for Treatment of Anaemia Before Cardiac Surgery. In: National H, Medical Research C, Australia, Monash U, editors. 2015. | Design |
| NCT02637102. The UK CAVIAR Study. 2015. | Outcome; nothing on adverse events |
| NCT02999217. Intravenous Iron for Correction of Anaemia After Colorectal Surgery. In: Orivas L, Pharmacosmos AS, editors. 2016. | Intervention |
| NCT03470649. Effect of IV Iron Isomaltoside on Postoperative Anemia in Total Knee Arthroplasty Patients. 2018. | Intervention; Post-operative administration of iron. |
| NCT03528564. Hemoglobin Optimization to Prevent Transfusion and Adverse Events in Perioperative Patients With Iron Restricted Anemia. 2018. | Other; duplicate (already included from databases) |
| NCT03560687. CardioSideral Heart Surgery: Randomized Study on Sucrosomial Iron Supplementation Before Heart Surgery. 2018. | Other; duplicate (already included from databases) |
| NCT03565354. Efficacy of Preoperative Intravenous Iron in Anaemic Colorectal Cancer Surgical Patients. 2018. | Other; duplicate (already included from databases) |
| Newton R, Mc Guckin S, Dick J, Chapman M, Westwood JP, Scully M. Administration of intravenous iron to optimise haemoglobin pre-operatively. British Journal of Haematology. 2019;185:64. doi: 10.1111/bjh.15854. | Design; conference abstract |
| Ng O, Keeler BD, Mishra A, Simpson JA, Neal K, Al‐Hassi HO, et al. Iron therapy for preoperative anaemia. Cochrane Database of Systematic Reviews. 2019;(12). doi: 10.1002/14651858.CD011588.pub3. | Other; all relevant individual studies covered by our review. |
| NICE. Evidence review for preoperative management of anaemia: Perioperative care in adults: Evidence review E. London: National Institute for Health and Care Excellence (UK); 2020. | Other; all relevant individual studies covered by our review. |
| Nieder AM, Rosenblum N, Lepor H. Comparison of two different doses of preoperative recombinant erythropoietin in men undergoing radical retropubic prostatectomy. Urology. 2001;57(4):737-41. doi: 10.1016/S0090-4295(00)01056-6. | Population |
| Norager CB, Jensen MB, Madsen MR, Qvist N, Laurberg S. Effect of darbepoetin alfa on physical function in patients undergoing surgery for colorectal cancer - A randomized, double-blind, placebo-controlled study. Oncology. 2006;71(3-4):212-20. doi: 10.1159/000106071. | Population |
| Nottingham University H, Trust NHS. Intravenous Iron: Measuring Response in Anemic Surgical Patients. 2011. | Design; Uncontrolled study |
| Nottingham University H, Trust NHS, National Institute for Health R, United K. IVICA: Intravenous Iron in Colorectal Cancer Associated Anaemia. 2014. | Other; duplicate of EUCTR2011-002185-21 |
| Nova Scotia H, Authority, Capital H, Canada, Dalhousie U. Effects of Oral Iron on Postoperative Fatigue Upon Coronary Artery Bypass Graft Patients. 2017. | Intervention |
| Okuyama M, Ikeda K, Shibata T, Tsukahara Y, Kitada M, Shimano T. Preoperative iron supplementation and intraoperative transfusion during colorectal cancer surgery. Surgery Today. 2005;35(1):36-40. doi: 10.1007/s00595-004-2888-0. | Outcome; nothing on adverse events |
| Olshove V, Berndsen N, Sivarajan V, Nawathe P, Phillips A. Comprehensive blood conservation program in a new congenital cardiac surgical program allows bloodless surgery for the Jehovah Witness and a reduction for all patients. Perfusion-Uk. 2018;33(3):194-202. doi: 10.1177/0267659117733810. | Population; Consists of both children and adults; no subgroup analysis performed on adults. |
| Osorio J, Jerico C, Miranda C, Garsot E, Luna A, Miro M, et al. Perioperative transfusion management in gastric cancer surgery: Analysis of the Spanish subset of the EURECCA oesophago-gastric cancer registry. Cirugia Espanola. 2018;96(9):546-54. doi: 10.1016/j.ciresp.2018.03.010. | Intervention; implementation of a PBM programme, consisting of different dosing/routes of iron administration and use of a more restrictive transfusion trigger. Not possible to determine effect of iron administration alone. |
| Panarese A, D’Andrea V, Pontone S, Kyriacou KA, Tonda M, Brighi M, et al. Can bloodless surgery be applied to every patient undergoing major abdominal surgical intervention? Archives of Hellenic Medicine. 2016;33(6):826-30. | Comparison |
| Pappa E, Vergados N, Spiridakis E, Chountas G, Apostolopoulou A, Sourmelis S. A Retrospective Comparative Study of Different Methods of Blood Management in Total Knee Replacement. J Knee Surg. 2018. doi: 10.1055/s-0038-1675217. | Design; Analysis includes data of another study (Goodnough NEJM 1999) that investigated the effect of EPO administration. As these are the only data of interest in this paper, and these data are not original, this paper cannot be included. |
| Parc de S, Mar, Vifor P. Comparing Intravenous and Oral Iron in Postoperative Anemia. 2012. | Population; Postoperatively anemic patients (and postoperative treatment with iron) |
| Park HS, Kim TY, Kim HJ, Ro YJ, Jang HY, Koh WU. The Effect of Intraoperative Ferric Carboxymaltose in Joint Arthroplasty Patients: A Randomized Trial. Journal of clinical medicine. 2019;8(10). Epub 2019/10/17. doi: 10.3390/jcm8101674. | Intervention; intra-operative administration of iron |
| Petis SM, Lanting BA, Vasarhelyi EM, Naudie DDR, Ralley FE, Howard JL. Is There a Role for Preoperative Iron Supplementation in Patients Preparing for a Total Hip or Total Knee Arthroplasty? J Arthroplasty. 2017;32(9):2688-93. doi: 10.1016/j.arth.2017.04.029. | Outcome; nothing on adverse events (Discussion: "Owing to the retrospective study design, we could not capture patients' compliance with the oral iron therapy and whether iron therapy discontinuation was because of adverse side effects.") |
| Pierson JL, Hannon TJ, Earles DR. A blood-conservation algorithm to reduce blood transfusions after total hip and knee arthroplasty. Journal of Bone and Joint Surgery-American Volume. 2004;86A(7):1512-8. doi: 10.2106/00004623-200407000-00022. | Population; Consists of both anemic and non-anaemic patients. Mean baseline Hb levels = 13.8 g/dl. |
| Ploem JE, Bloem JA. Compensation for blood loss in plastic surgery with ferastral and autologous blood transfusion. Scand J Haematol Suppl. 1977;32:298-302. | Population; Non-anaemic ("Hb-concentrations at first attendance, before operation, ranged from 12.0 to 17.3 g/100 ml (mean 13.7 g/100 ml), i.e. all were within normal limits.") |
| Podestà A, Carmagnini E, Parodi E, Dottori V, Crivellari R, Barberis L, et al. Elective coronary and valve surgery without blood transfusion in patients treated with recombinant human erythropoietin (epoetin-α). Minerva Cardioangiologica. 2000;48(11):341-7. | Population |
| Polanco-García M, Capielo AM, Miret X, Chamero A, Sainz J, Revilla E, et al. Effectiveness of a patient blood management protocol on reduction of allogeneic red blood cell transfusions in orthopedic surgery. Med Clin (Barc). 2019;152(3):90-7. Epub 2018/06/12. doi: 10.1016/j.medcli.2018.04.021. | Outcome; nothing on adverse events |
| Politano N, Jaskolka M, Blakey G, Turvey T, White R, Phillips C. The Effect of Preoperative Recombinant Erythropoietin on Postoperative Hematocrit Level After Orthognathic Surgery. Journal of Oral and Maxillofacial Surgery. 2012;70(11):E625-E30. doi: 10.1016/j.joms.2012.07.021. | Population; Consists of people aged = or >13 years; therefore children and adults. |
| Pompei E, Tursi V, Guzzi G, Vendramin I, Ius F, Muzzi R, et al. Mid-term clinical outcomes in cardiac surgery of Jehovah's witnesses. J Cardiovasc Med (Hagerstown). 2010;11(3):170-4. doi: 10.2459/JCM.0b013e3283330752. | Population; No subgroup analysis on anaemic population (16/34 patients) |
| Poon E, Pache D, Delaforce A, Abdalla L, McGuire T. Anaemia in patients undergoing major bowel surgery - Prevalence and current practice: A public and private institution experience. Journal of perioperative practice. 2020:1750458920934321. Epub 2020/07/09. doi: 10.1177/1750458920934321. | Population; Table 3 provides an overview of the number of anaemic patients receiving preoperative treatment, but there is no comparison between those patients and the ones who did not receive treatment. The paper further on only provides data on comparison anaemic vs non-anaemic patients. |
| Poulsen KA, Qvist N, Winther K, Boesby S. Haemostatic aspects of recombinant human erythropoietin in colorectal surgery. Eur J Surg. 1998;164(3):211-5. doi: 10.1080/110241598750004661. | Population |
| Poulsen TD, Andersen LW, Steinbrüchel D, Gøtze JP, Jørgensen OS, Olsen NV. Two large preoperative doses of erythropoietin do not reduce the systemic inflammatory response to cardiac surgery. Journal of cardiothoracic and vascular anesthesia. 2009;23(3):316‐23. doi: 10.1053/j.jvca.2008.08.018. | Population |
| Pujol-Nicolas A, Morrison R, Casson C, Khan S, Marriott A, Tiplady C, et al. Preoperative screening and intervention for mild anemia with low iron stores in elective hip and knee arthroplasty. Transfusion. 2017;57(12):3049-57. doi: 10.1111/trf.14372 | Outcome; nothing on adverse events |
| Quarterman C, Shaw M, Hughes S, Wallace V, Agarwal S. Anaemia in cardiac surgery - a retrospective review of a centre's experience with a pre-operative intravenous iron clinic. Anaesthesia. 2020. Epub 2020/11/06. doi: 10.1111/anae.15271. | Population; Table 2 provides an overview of data of anaemic patients who received iv iron vs anaemic patients who did not receive iv iron. However, this control group consists of both people who did not receive iv iron due to logistic reasons ánd people who were not iron-deficient. Therefore, no comparable groups. |
| Quinn M, Drummond RJ, Ross F, Murray J, Murphy J, Macdonald A. Short course pre-operative ferrous sulphate supplementation--is it worthwhile in patients with colorectal cancer? Ann R Coll Surg Engl. 2010;92(7):569-72. doi: 10.1308/003588410x12699663904277. | Outcome; No specific adverse effects data. |
| Quintana-Diaz M, Fabra-Cadenas S, Gomez-Ramirez S, Martinez-Virto A, Garcia-Erce JA, Munoz M. A fast-track anaemia clinic in the Emergency Department: feasibility and efficacy of intravenous iron administration for treating sub-acute iron deficiency anaemia. Blood Transfusion. 2016;14(2):126-33. doi: 10.2450/2015.0176-15. | Population; all patients presenting to the emergency department; no mentioning of elective surgery anywhere in the paper. |
| Radia D, Momoh I, Dillon R, Francis Y, Cameron L, Fagg TL, et al. Anemia management: Development of a rapid-access anemia and intravenous iron service. Risk Management and Healthcare Policy. 2013;6:13-22. doi: 10.2147/RMHP.S41818. | Design; No original study. Overview that includes results of a pilot study by Momoh. |
| Rambam Health C, Campus, Carmel M, Center, The Baruch Padeh Medical C, Poriya. The Effect of Ferric Carboxymaltose on Hemoglobin and Blood Transfusion in Cardiac Surgery. 2018. | Population |
| Ranucci M, Pavesi M, Pistuddi V, Baryshnikova E. Preoperative Anemia Correction in Cardiac Surgery: A Propensity-Matched Study. J Cardiothorac Vasc Anesth. 2020. Epub 2020/08/04. doi: 10.1053/j.jvca.2020.07.015. | Population; Only provides data on comparison anaemic vs non-anaemic patients. |
| Rauh MA, Bayers-Thering M, LaButti RS, Krackow KA. Preoperative administration of epoetin alfa to total joint arthroplasty patients. Orthopedics. 2002;25(3):317-20. | Outcome; nothing on adverse events |
| Razurel A, Dupont X, Politis B, Chauny JV, Boyeldieu D, Benhamou F, et al. Pre-operative oral iron supplementation in the prevention of post-operative acute anaemia in orthopaedic surgery. Journal de Pharmacie Clinique. 2014;33(1):21-31. doi: 10.1684/jpc.2014.0272. | Outcome; nothing on adverse events |
| Reddy SM, Talwar S, Velayoudam D, Gharde P, Mallick V, Jha RK, et al. Multi-modality blood conservation strategy in open-heart surgery: an audit. Interact Cardiovasc Thorac Surg. 2009;9(3):480-2. doi: 10.1510/icvts.2009.203034. | Population; Patients are aged >15 years, so both children and adults included. Moreover, mixture of elective and emergency surgery. No subgroup analyses. |
| Rheude T, Pellegrini C, Lessmann L, Wiebe J, Mayr NP, Michel J, et al. Prevalence and Clinical Impact of Iron Deficiency in Patients With Severe Aortic Stenosis Referred for Transcatheter Aortic Valve Implantation. The American journal of cardiology. 2019;124(9):1442-8. Epub 2019/09/03. doi: 10.1016/j.amjcard.2019.07.051. | Population; See figure 3 and 4; only data provided before vs after IV iron administration. No comparison with control group that did not get IV iron. |
| Richards T, Clevenger B, Keidan J, Collier T, Klein AA, Anker SD, et al. Erratum to: PREVENTT: preoperative intravenous iron to treat anaemia in major surgery: study protocol for a randomised controlled trial. Trials. 2015;16:312. doi: 10.1186/s13063-015-0869-9. | Design; Erratum only corrects "Acknowledgements" section; no data. |
| Rineau E, Chaudet A, Chassier C, Bizot P, Lasocki S. Implementing a blood management protocol during the entire perioperative period allows a reduction in transfusion rate in major orthopedic surgery: a before-after study. Transfusion. 2016;56(3):673-81. doi: 10.1111/trf.13468. | Outcome; nothing on adverse events |
| Ripolles-Melchor J, Jerico-Alba C, Quintana-Diaz M, Garcia-Erce JA. From blood saving programs to patient blood management and beyond. Medicina Clinica. 2018;151(9):368-73. doi: 10.1016/j.medcli.2018.02.027. | Design; Narrative review |
| Ritter MA. Blood management in total joint replacement: the need for erythropoietin alpha. Orthopedics. 2002;25(9):915. | Design; Editorial/comment |
| Rogers BA, Cowie A, Alcock C, Rosson JW. Identification and treatment of anaemia in patients a waiting hip replacement. Annals of the Royal College of Surgeons of England. 2008;90(6):504-7. doi: 10.1308/003588408X301163. | Outcome; nothing on adverse events |
| Rogers SN, Horisk K, Groom P, Lowe D. Management of anaemia and blood in patients having neck dissections or free flaps for head and neck cancer. The British journal of oral & maxillofacial surgery. 2019;57(6):543-9. Epub 2019/05/28. doi: 10.1016/j.bjoms.2019.05.001. | Population; No separate data on iron use in the anaemic proportion of the patients |
| Rohling RG, Zimmermann AP, Biro P, Haers PE, Sailer HF. Alternative methods for reduction of blood loss during elective orthognathic surgery. Int J Adult Orthodon Orthognath Surg. 1999;14(1):77-82. | Population; Same study as Rohling 1998. The only interesting intervention is the administration of EPO. However, this was only administered to people whose preoperative Hct was = or > 45%. Therefore, non-anaemic population. |
| Rohling RG, Zimmermann AP, Haers PE, Locher MC, Major A, Sailer HF. [Allo-transfusion saving measures in maxillofacial surgery]. Swiss Surg. 1998;4(3):133-40. | Population; The only interesting intervention is the administration of EPO. However, this was only administered to people whose preoperative Hct was = or > 45%. Therefore, non-anaemic population. |
| Rosenblum N, Levine MA, Handler T, Lepor H. The role of preoperative epoetin alfa in men undergoing radical retropubic prostatectomy. J Urol. 2000;163(3):829-33. | Design; Uncontrolled study |
| Rössler J, Hegemann I, Schoenrath F, Seifert B, Kaserer A, Spahn GH, et al. Efficacy of quadruple treatment on different types of pre-operative anaemia: secondary analysis of a randomised controlled trial. Anaesthesia. 2020;75(8):1039-49. Epub 2020/04/29. doi: 10.1111/anae.15062. | Intervention; see study Spahn |
| Rössler J, Kaserer A, Spahn GH, Spahn DR. Not all anemia is solely due to iron deficiency. Journal of Thoracic Disease. 2020;12(3):1130-2. doi: 10.21037/jtd.2019.12.129. | Design; letter to the editor |
| Rutherford CJ, Schneider TJ, Dempsey H, Kirn DH, Brugnara C, Goldberg MA. Efficacy of Different Dosing Regimens for Recombinant-Human-Erythropoietin in a Simulated Perisurgical Setting - the Importance of Iron Availability in Optimizing Response. American Journal of Medicine. 1994;96(2):139-45. doi: 10.1016/0002-9343(94)90134-1. | Population; no elective surgery patients |
| Sanchez AMM, Pampillon N, Abaurre M, Omelanczuk P. Pre-Operative Iron Deficiency in Bariatric Surgery: Diagnosis and Treatment. Nutricion Hospitalaria. 2015;32(1):75-9. doi: 10.3305/nh.2015.32.1.8871. | Other; missed duplicate |
| Sanquin R, Blood B, Divisions, Erasmus M, Center. Well Being of Obstetric Patients on Minimal Blood Transfusions. 2011. | Population; no elective surgery patients |
| Santoro JE, Eastlack RK, Mirocha JM, Bugbee WD. Impact of erythropoietin on allogenic blood exposure in orthopedic surgery. Am J Orthop (Belle Mead NJ). 2007;36(11):600-4. | Outcome; nothing on adverse events |
| Scardino M, Di M, Martorelli F, Tanzi D, Kon E, D’Amato T. Improved patient blood management and cost saving in hip replacement surgery through the implementation of pre-operative Sucrosomial® iron supplementation: a quality improvement assessment study. International Orthopaedics. 2019;43(1):39-46. doi: 10.1007/s00264-018-4149-7. | Population; no anaemia |
| Scardino M, D'Amato T, Martorelli F, Fenocchio G, Pera E, Tarantino G. A cost-effective implementation of pre-operative protocol with sucrosomial® iron supplementation. Blood Transfusion. 2019;17:s7-s8. doi: 10.2450/2019.S1. | Design; conference abstract |
| Scolletta S, Simioni P, Campagnolo V, Celiento M, Fontanari P, Guadagnucci A, et al. Patient blood management in cardiac surgery: The “Granducato algorithm”. International Journal of Cardiology. 2019;289:37-42. doi: 10.1016/j.ijcard.2019.01.025. | Population; Mix of elective and non-elective surgery patients. No separate data on elective surgery. |
| Scrimshire AB, Booth A, Fairhurst C, Kotze A, Reed M, McDaid C. Preoperative iron treatment in anaemic patients undergoing elective total hip or knee arthroplasty: a systematic review and meta-analysis. BMJ open. 2020;10(10):e036592. Epub 2020/11/02. doi: 10.1136/bmjopen-2019-036592. | Other; all relevant individual studies covered by our review. |
| Sesti F, Ticconi C, Bonifacio S, Piccione E. Preoperative administration of recombinant human erythropoietin in patients undergoing gynecologic surgery. Gynecol Obstet Invest. 2002;54(1):1-5. doi: 10.1159/000064688. | Outcome; No specific data on adverse events/side effects. |
| Sheth SS, Das SB. Preoperative management of anemia to avoid blood transfusion. International Journal of Gynecology and Obstetrics. 2002;77(3):245-7. doi: 10.1016/S0020-7292(02)00031-0. | Design; Uncontrolled study |
| Shevchenko IL, Khubulava GG, Chechetkin AV, Belevitin AB, Solov'eva NI. Erythropoietin in the prevention and treatment of anemia in heart surgery patients. Vestnik khirurgii imeni I I Grekova. 1999;158(6):69-71. | Intervention |
| Shin HW, Park JJ, Kim HJ, You HS, Choi SU, Lee MJ. Efficacy of perioperative intravenous iron therapy for transfusion in orthopedic surgery: A systematic review and meta-analysis. PLoS One. 2019;14(5):e0215427. Epub 2019/05/07. doi: 10.1371/journal.pone.0215427. | Other; all relevant individual studies covered by our review. |
| Sinclair RC, Duffield KE, de Pennington JH. Improving preoperative haemoglobin using a quality improvement approach to treat iron deficiency anaemia. BMJ open quality. 2020;9(1). Epub 2020/01/28. doi: 10.1136/bmjoq-2019-000776. | Intervention; Broad PBM intervention, including changes in the day ward facilities of hospitals. Not possible to distinguish the effect of iron administration only. |
| Singapore G, Hospital, Duke NUSGMS. Preoperative Intravenous Iron Infusion to Reduce Post-surgical Complications: a Pilot Randomised Control Trial. 2019. | Other; duplicate (already included from databases) |
| Singer MB, Sheckley M, Menon VG, Sundaram V, Donchev V, Voidonikolas G, et al. Can Transfusions Be Eliminated in Major Abdominal Surgery? Analysis of a Five-Year Experience of Blood Conservation in Patients Undergoing Pancreaticoduodenectomy. Am Surg. 2015;81(10):983-7. | Population; no elective surgery patients |
| Slappendel R, Dirksen R, Weber EWG, Van Der S, D B. An algorithm to reduce allogenic red blood cell transfusions for major orthopedic surgery. Acta Orthopaedica Scandinavica. 2003;74(5):569-75. doi: 10.1080/00016470310017974. | Design; Narrative overview of PBM policy |
| Slappendel R, Weber EWG, Hémon YJM, Mähler S, Dalén T, Rouwet EFAM, et al. Patients with and without rheumatoid arthritis benefit equally from preoperative epoetin-α treatment. Acta Orthopaedica. 2006;77(4):677-83. doi: 10.1080/17453670610012782. | Design; Post-hoc analysis of previously published trial (Weber 2005). |
| Smith D, Delmore B. New York University Medical Center's pilot "Epo-Depot" program: a win-win for patients and health care. J Assoc Nurses AIDS Care. 2004;15(6):23-30. doi: 10.1177/1055329004271123. | Design; Purely descriptive study. No actual data. |
| Smith A, Moon T, Pak T, Park B, Urman RD. Preoperative Anemia Treatment With Intravenous Iron in Patients Undergoing Major Orthopedic Surgery: A Systematic Review. Geriatric orthopaedic surgery & rehabilitation. 2020;11:2151459320935094. Epub 2020/07/09. doi: 10.1177/2151459320935094. | Other; all relevant individual studies covered by our review. |
| So-Osman C. Cost-effectiveness of patient blood management methods in elective orthopaedic surgery. State of the Art Presentations 33rd International Congress of the International Society of Blood Transfusion, in Conjunction with the 33rd Congress of the Ksbt and 2014 Congress of the Korean Hematology Societies, Vol 10, No S1. 10. Oxford: Blackwell Science Publ; 2015. p. 146-9. | Outcome; nothing on adverse events |
| Sowade O, Gross J, Sowade B, Warnke H, Franke W, Messinger D, et al. Evaluation of oxygen availability with oxygen status algorithm in patients undergoing open heart surgery treated with epoetin beta. J Lab Clin Med. 1997;129(1):97-105. | Population; Non-anaemic population (see other study Sowade 1997 "Avoidance of allogeneic blood transfusions..."). |
| Sowade O, Messinger D, Franke W, Sowade B, Scigalla P, Warnke H. The estimation of efficacy of oral iron supplementation during treatment with epoetin beta (recombinant human erythropoietin) in patients undergoing cardiac surgery. Eur J Haematol. 1998;60(4):252-9. | Population; no preop anaemia |
| Sowade O, Sowade B, Brilla K, Franke W, Stephan P, Gross J, et al. Kinetics of reticulocyte maturity fractions and indices and iron status during therapy with epoetin beta (recombinant human erythropoietin) in cardiac surgery patients. Am J Hematol. 1997;55(2):89-96. | Population; Non-anaemic population (see other study Sowade 1997 "Avoidance of allogeneic blood transfusions..."). |
| Sowade O, Sowade B, Gross J, Brilla K, Ziemer S, Franke W, et al. Evaluation of erythropoietic activity on the basis of the red cell and reticulocyte distribution widths during epoetin beta therapy in patients undergoing cardiac surgery. Acta Haematol. 1998;99(1):1-7. doi: 10.1159/000040720. | Non-anaemic population (mean Hb levels before treatment 8.9 and 8.6 mmol/l). |
| Sowade O, Warnke H, Scigalla P. [Operations with a heart-lung machine in adult members of Jehovah's Witnesses]. Anaesthesist. 1995;44(4):257-64. | Outcome; nothing on adverse events |
| Sowade O, Warnke H, Scigalla P, Sowade B, Franke W, Messinger D, et al. Avoidance of allogeneic blood transfusions by treatment with epoetin beta (recombinant human erythropoietin) in patients undergoing open-heart surgery. Blood. 1997;89(2):411-8. | Population; no preop anaemia |
| Sowade O, Ziemer S, Sowade B, Franke W, Messinger D, Ziebell E, et al. The effect of preoperative recombinant human erythropoietin therapy on platelets and hemostasis in patients undergoing cardiac surgery. J Lab Clin Med. 1997;129(3):376-83. | Population; Non-anaemic population (see other study Sowade 1997 "Avoidance of allogeneic blood transfusions..."). |
| Spahn DR, Schoenrath F, Spahn GH, Seifert B, Stein P, Theusinger OM, et al. Effect of ultra-short-term treatment of patients with iron deficiency or anaemia undergoing cardiac surgery: a prospective randomised trial. Lancet. 2019;393(10187):2201-12. Epub 2019/05/01. doi: 10.1016/s0140-6736(18)32555-8. | Intervention; Not the same co-interventions in both groups |
| Steinbicker AU, Zurheiden NJ, Buckmann A, Venherm S, Schopper C, Geissler GR, et al. Patient blood management - Implementation within the anaesthesia clinic. Anasthesiologie & Intensivmedizin. 2015;56:64-74. | Intervention; implementation of the PBM programme. |
| Sturgis EM, Gianoli GJ, Miller RH, Fisher JW. Avoiding transfusion in head and neck surgery: Feasibility study of erythropoietin. Laryngoscope. 2000;110(1):51-7. doi: 10.1097/00005537-200001000-00011. | Intervention; No actual administration of erythropoietin. Hypothetical study. |
| Styron JF, Klika AK, Szubski CR, Tolich D, Barsoum WK, Higuera CA. Relative efficacy of tranexamic acid and preoperative anemia treatment for reducing transfusions in total joint arthroplasty. Transfusion. 2017;57(3):622-9. doi: 10.1111/trf.13955. | Intervention; PBM programme (iron, EPO, tranexamic acid, aspirin use). Not possible to extract relevant data on people who only received iron and/or EPO. |
| Sunnybrook Health S, Centre. Oral Iron vs. Placebo in Newly Diagnosed Gynecologic Oncology Patients Who Are Surgical Candidates. 2017. | Outcome; nothing on adverse events |
| Tahir S, Saeed K, Nazeer S, Rehman M, Rasul S. Comparison of intravenous iron sucrose alone versus intravenous iron sucrose along with erythropoietin for management of anemia for gynecological patients waiting for surgery. Pakistan journal of medical and health sciences. 2019;13(3):566‐9. | Intervention; contacted authors twice to obtain additional information on specifics concerning (co-)interventions (e.g. timing, duration). No response obtained. |
| Tamir L, Fradin Z, Fridlander M, Ashkenazi U, Zeidman A, Cohen AM, et al. Recombinant human erythropoietin reduces allogeneic blood transfusion requirements in patients undergoing major orthopedic surgery. Haematologia (Budap). 2000;30(3):193-201. | Population |
| Tanaka A, Ota T, Uriel N, Asfaw Z, Onsager D, Lonchyna VA, et al. Cardiovascular surgery in Jehovah's Witness patients: The role of preoperative optimization. J Thorac Cardiovasc Surg. 2015;150(4):976-83.e1. doi: 10.1016/j.jtcvs.2015.06.059. | Intervention; Preoperative optimization protocol included supplementation with oral (intravenous) iron or vitamin (vitamin B12, folic acid) and subcutaneous (intravenous) erythropoietin to increase preoperative hemoglobin greater than 12 g/dL. Patients are categorized as optimized or non-optimized. No subgroup analyses on use of iron and/or EPO. |
| Tang GH, Dhir V, Scheer AS, Tricco AC, Sholzberg M, Brezden-Masley C. Intravenous iron versus oral iron or observation for gastrointestinal malignancies: a systematic review. European journal of gastroenterology & hepatology. 2019;31(7):799-808. Epub 2019/05/15. doi: 10.1097/meg.0000000000001433. | Other; all relevant individual studies covered by our review. |
| Tasanarong A, Duangchana S, Sumransurp S, Homvises B, Satdhabudha O. Prophylaxis with erythropoietin versus placebo reduces acute kidney injury and neutrophil gelatinase-associated lipocalin in patients undergoing cardiac surgery: a randomized, double-blind controlled trial. Bmc Nephrology. 2013;14. doi: 10.1186/1471-2369-14-136. | Population |
| Tayo AO, Dosunmu AO, Akinola IO, Adewunmi A, Oloyede OA, Akinbami AA, et al. An open-label, randomized, parallel-group comparative study of the efficacy of sorghum bicolor extract in preoperative anemia. Nutrition. 2017;33:113-7. doi: 10.1016/j.nut.2016.05.005. | Population |
| Theusinger OM, Kind SL, Seifert B, Borgeat A, Gerber C, Spahn DR. Patient blood management in orthopaedic surgery: A four-year follow-up of transfusion requirements and blood loss from 2008 to 2011 at the Balgrist University Hospital in Zurich, Switzerland. Blood Transfusion. 2014;12(2):195-203. doi: 10.2450/2014.0306-13. | Intervention; EPO+iron+folic acid+vitamin B12.  In addition: "Complications and serious adverse events could not be analysed due to changes in codification with the introduction of the Swiss DRG system and due to the fact that in 2008 no data on complications were electronically available." |
| Țigliș M, Neagu TP, Niculae A, Lascăr I, Grințescu IM. Incidence of Iron Deficiency and the Role of Intravenous Iron Use in Perioperative Periods. Medicina (Kaunas, Lithuania). 2020;56(10). Epub 2020/10/16. doi: 10.3390/medicina56100528. | Design; no actual systematic review |
| Tomeczkowski J, Stern S, Muller A, von H. Potential cost saving of Epoetin alfa in elective hip or knee surgery due to reduction in blood transfusions and their side effects: a discrete-event simulation model. PLoS One. 2013;8(9):e72949. doi: 10.1371/journal.pone.0072949. | Design |
| Tovey FI, Clark CG. Anaemia after partial gastrectomy: a neglected curable condition. Lancet. 1980;1(8175):956-8. | Population; post-operative anaemia |
| Trentino KM, Mace H, Symons K, Sanfilippo FM, Leahy MF, Farmer SL, et al. Associations of a Preoperative Anemia and Suboptimal Iron Stores Screening and Management Clinic in Colorectal Surgery With Hospital Cost, Reimbursement, and Length of Stay: A Net Cost Analysis. Anesth Analg. 2020. Epub 2020/10/27. doi: 10.1213/ane.0000000000005241. | Intervention; Broad PBM intervention. Not possible to distinguish the effect of iron administration only. |
| Trentino KM, Mace HS, Symons K, Sanfilippo FM, Leahy MF, Farmer SL, et al. Screening and treating pre-operative anaemia and suboptimal iron stores in elective colorectal surgery: a cost effectiveness analysis. Anaesthesia. 2020. Epub 2020/08/28. doi: 10.1111/anae.15240. | Intervention; Broad PBM intervention. Not possible to distinguish the effect of iron administration only. |
| Tsuji Y, Kambayashi J, Shiba E, Sakon M, Kawasaki T, Mori T. Effect of recombinant human erythropoietin on anaemia after gastrectomy: a pilot study. Eur J Surg. 1995;161(1):29-33. | Population |
| Tsuji Y, Kambayashi JI, Shiba E, Sakon M, Kawasaki T, Mori T. Effect of Recombinant-Human-Erythropoietin on Anemia after Gastrectomy - a Pilot-Study. European Journal of Surgery. 1995;161(1):29-33. | Other; duplicate |
| Umstadt HE, Weippert-Kretschmer M, Austermann KH, Kretschmer V. Need for transfusions in orthognathic surgery. No general indication for preoperative autologous blood donation. Mund-, Kiefer- und Gesichtschirurgie : MKG. 2000;4(4):228-33. | Intervention; No administration of EPO or iron. |
| University C, London. Preoperative Intravenous Iron to Treat Anaemia in Major Surgery. 2018. | Other; full text paper already included (Richards 2015) |
| University of Zurich, Vifor I. Intravenous Ferric Carboxymaltose (Ferinject®) With or Without Erythropoietin in Patients Undergoing Orthopaedic Surgery. 2012. | Other; duplicate (already included from databases) |
| Vaislic C, Bical O, Deleuze P, Khoury W, Gaillard D, Ponzio O, et al. Cardiac surgery without transfusion in 2005. Archives Des Maladies Du Coeur Et Des Vaisseaux. 2005;98(1):7-12. | Population; Control group: all patients with preoperative Hb levels of <14 g/dl were excluded. --> non-anaemic population |
| Vaislic CD, Dalibon N, Ponzio O, Ba M, Jugan E, Lagneau F, et al. Outcomes in cardiac surgery in 500 consecutive Jehovah's Witness patients: 21 year experience. J Cardiothorac Surg. 2012;7:95. doi: 10.1186/1749-8090-7-95. | Population; In control group: all patients with Hb levels <14g/dl were excluded from the study, as they could not undergo surgery. |
| Van der Linden, De H, Daper A, Trenchant A, Jacobs D, De B, et al. A standardized multidisciplinary approach reduces the use of allogeneic blood products in patients undergoing cardiac surgery. Canadian Journal of Anesthesia. 2001;48(9):894-901. | Population; Non-anaemic (see fig 3) |
| Van Der Linden, Hardy JF. Implementation of patient blood management remains extremely variable in Europe and Canada: The NATA benchmark project: An observational study. European Journal of Anaesthesiology. 2016;33(12):913-21. doi: 10.1097/EJA.0000000000000519. | Intervention; Not possible to compare people who received EPO with people who only received iron. |
| van Wijk L, Bos J, Klaase JM. Prehabilitation worth it? A theoretical algorithmic for the cost-effectiveness of a multimodal prehabilitation program for complex abdominal surgery. HPB. 2020;22:S64. doi: 10.1016/j.hpb.2020.04.525. | Design; conference abstract |
| Verma V, Schwarz RE. Factors influencing perioperative blood transfusions in patients with gastrointestinal cancer. J Surg Res. 2007;141(1):97-104. doi: 10.1016/j.jss.2007.03.032. | Intervention; No actual EPO or iron administration. |
| von Bormann, Weiler J, Aulich S. Acute treatment with recombinant erythropoietin in patients with pre- and postoperative anemia: a clinical report. Clin Investig. 1994;72(6 Suppl):S31-5. | Design; Uncontrolled study |
| Voorn VMA, Marang-van de M, P J, van der H, Hofstede SN, So-Osman C, et al. The effectiveness of a de-implementation strategy to reduce low-value blood management techniques in primary hip and knee arthroplasty: a pragmatic cluster-randomized controlled trial. Implementation science. 2017;12(1):72. doi: 10.1186/s13012-017-0601-0. | Intervention; de-implementation strategy |
| Wan S, Sparring V, Cabrales DA, Jansson KÅ, Wikman A. Clinical and Budget Impact of Treating Preoperative Anemia in Major Orthopedic Surgery—A Retrospective Observational Study. Journal of Arthroplasty. 2020. doi: 10.1016/j.arth.2020.06.018. | Design; Cost simulation model; no actual data on effectiveness (let alone adverse events) of IV iron administration |
| Warm M, Kates R, Dick M, Thomas A, Mallmann P, Hoopmann M, et al. Optimized pro-active management of anemia by Epoetin α in pre-operative chemotherapy for primary breast cancer. Oncology Reports. 2009;21(3):777-85. doi: 10.3892/or_00000284. | Outcome; nothing on adverse events |
| Watanabe Y, Fuse K, Naruse Y, Kobayashi T, Yamamoto S, Konishi H, et al. Subcutaneous Use of Erythropoietin in Heart-Surgery. Annals of Thoracic Surgery. 1992;54(3):479-84. doi: 10.1016/0003-4975(92)90438-A. | Population |
| Weber EW, Slappendel R, Hemon Y, Mahler S, Dalen T, Rouwet E, et al. Effects of epoetin alfa on blood transfusions and postoperative recovery in orthopaedic surgery: the European Epoetin Alfa Surgery Trial (EEST). Eur J Anaesthesiol. 2005;22(4):249-57. | Population |
| Weltert L, D'Alessandro S, Nardella S, Girola F, Bellisario A, Maselli D, et al. Preoperative very short-term, high-dose erythropoietin administration diminishes blood transfusion rate in off-pump coronary artery bypass: a randomized blind controlled study. J Thorac Cardiovasc Surg. 2010;139(3):621-6; discussion 6. doi: 10.1016/j.jtcvs.2009.10.012. | Population |
| Weltert LP. Pre-operative cardio surgery protocol implementation with Sucrosomial® iron. Blood Transfusion. 2020;18:s7. | Design; conference abstract |
| Wolfson TS, Novikov D, Chen KK, Kim KY, Anoushiravani AA, Deshmukh AJ, et al. Total Knee Arthroplasty Is Safe in Jehovah's Witness Patients-A 12-Year Perspective. J Knee Surg. 2020;33(1):34-41. Epub 2019/01/09. doi: 10.1055/s-0038-1676372 | Population; Mix of anemic and non-anaemic patients. |
| Wong CJ, Vandervoort MK, Vandervoort SL, Donner A, Zou G, MacDonald JK, et al. A cluster-randomized controlled trial of a blood conservation algorithm in patients undergoing total hip joint arthroplasty. Transfusion. 2007;47(5):832-41. doi: 10.1111/j.1537-2995.2007.01197.x. | Intervention; introduction of a PBM algorithm. |
| Wong S, Tang H, de Steiger R. Blood management in total hip replacement: an analysis of factors associated with allogenic blood transfusion. ANZ J Surg. 2015;85(6):461-5. Epub 2015/03/18. doi: 10.1111/ans.13048. | Intervention; no iron/EPO. Examines the risk of anemia on complications. |
| Wu YG, Zeng Y, Shen B, Si HB, Cao F, Yang TM, et al. Combination of erythropoietin and tranexamic acid in bilateral simultaneous total hip arthroplasty: a randomised, controlled trial. Hip Int. 2016;26(4):331-7. doi: 10.5301/hipint.5000356. | Population; Non-anaemic ("The inclusion criterion for patients was a preoperative Hb level of 120-150 g/L for females and 130-150 g/L for males.") |
| Wurnig C, Schatz K, Noske H, Hemon Y, Dahlberg G, Josefsson G, et al. Subcutaneous low-dose epoetin beta for the avoidance of transfusion in patients scheduled or elective surgery not eligible for autologous blood donation. European Surgical Research. 2001;33(5-6):303-10. doi: 10.1159/000049723. | Population |
| Xu H, Duan Y, Yuan X, Wu H, Sun H, Ji H. Intravenous Iron Versus Placebo in the Management of Postoperative Functional Iron Deficiency Anemia in Patients Undergoing Cardiac Valvular Surgery: A Prospective, Single-Blinded, Randomized Controlled Trial. J Cardiothorac Vasc Anesth. 2019;33(11):2941-8. Epub 2019/04/02. doi: 10.1053/j.jvca.2019.01.063. | Population; patients with preoperative anemia are excluded |
| Xi YL, Duan YP, Yang Q, Yang Y, Zhang ZX, Zhao XD, et al. Tranexamic acid combined with iron sucrose reduces blood transfusion in primary total hip arthroplasty: a prospective, randomized, controlled trial. Chinese journal of tissue engineering research. 2018;22(27):4288‐93. doi: 10.3969/j.issn.2095-4344.0341. | Population; See inclusion criteria: 1.4.1.: "There are no obvious abnormalities in blood coagulation function, hemoglobin, and platelets before surgery" + table 2 Hb values >13 g/dl |
| Yanagawa B, Rocha RV, Mazine A, Verma S, Mazer CD, Vernich L, et al. Hemoglobin Optimization for Coronary Bypass: A 10-Year Canadian Multicenter Experience. Ann Thorac Surg. 2019;107(3):711-7. Epub 2018/12/12. doi: 10.1016/j.athoracsur.2018.10.067. | Intervention; implementation of a multifaceted PBM programme. |
| Yazicioǧlu L, Eryilmaz S, Širlak M, Bahadir I, Aral A, Taşöz R, et al. Recombinant human erythropoietin administration in cardiac surgery. Journal of Thoracic and Cardiovascular Surgery. 2001;122(4):741-5. doi: 10.1067/mtc.2001.115426. | Outcome; nothing on adverse events |
| Zhang S, Huang Q, Xu B, Ma J, Cao G, Pei F. Effectiveness and safety of an optimized blood management program in total hip and knee arthroplasty: A large, single-center, retrospective study. Medicine (Baltimore). 2018;97(1):e9429. doi: 10.1097/md.0000000000009429. | Outcome; nothing on adverse events |
| Zhou LX, Wu WX, Li M, Yu WJ. Clinical observation of recombinant human erythropoietin in treatment of preoperative anemia in patients with gastric cancer. Pharmaceutical care and research. 2006;6(5):355‐7. | Other; full text not available |
| Ziabakhsh-Tabary S, Mokhtari-Esbuie F. Effects of single dose of erythropoietin on blood transfusion requirement in patients undergoing coronary artery bypass graft surgery. Journal of babol university of medical sciences. 2013;15(2):18‐24. | Intervention |
| Zwiep TM, Gilbert RWD, Moloo H, Touchie D, Martel G, Wallace T, et al. Improving the treatment of pre-operative anemia in hepato-pancreato-biliary patients: a quality improvement initiative. Patient safety in surgery. 2020;14:18. Epub 2020/04/30. doi: 10.1186/s13037-020-00239-5. | Intervention; Broad PBM intervention. Not possible to distinguish the effect of oral iron prescription only. |

### Supplementary Table 3. List of ongoing/prematurely ended trials and published protocols.

| **ONGOING/PREMATURELY ENDED TRIALS** | | | |
| --- | --- | --- | --- |
| **Trial registry number** | **Trial title** | **Year of registration** | **Trial status** |
| **Ongoing** | | | |
| ISRCTN41421863 / EUCTR2011-003695-36-GB | Treatment of a low blood count before heart surgery comparing iron supplements taken by mouth and the combination of iron given by intravenous drip and a drug to increase the blood count | 2012 | Completed; full text paper in preparation (personal communication with Kong) |
| NCT02243735 | Trial Comparing Ferric(III)Carboxymaltose Infusion With Oral Iron Suppletion as Treatment of Anaemia | 2014 | Unknown (last recruitment status was 'recruiting', verified August 2018;  trial still ongoing, obtained via personal communication with Bemelman) |
| NCT02632760 | Intravenous Iron for Treatment of Anaemia Before Cardiac Surgery | 2015 | Recruiting |
| NCT02385383 | An Intravenous Iron Based Protocol for Preoperative Anaemia in Hip and Knee Surgery - An Observational Study | 2015 | Unknown  (study investigators contacted, but no response obtained) |
| NCT03295851 | Preoperative Intravenous Iron Infusion to Reduce Post-surgical Complications: a Pilot Randomised Control Trial | 2017 | Unknown (last recruitment status was 'recruiting', verified December 2018; study investigators contacted, but no response obtained) |
| EUCTR2017-003416-38-DE | Clinical trial to demonstrate safety and efficacy of an intravenous (i.v.) administration of Feramyl compared to i.v. Ferinject and to iron tablets in patients with iron deficiency anaemia diagnosed before a planned non-cardiac surgery | 2017 | Temporarily halted |
| NCT03528564 | Hemoglobin Optimization to Prevent Transfusion and Adverse Events in Perioperative Patients With Iron Restricted Anemia | 2018 | Recruiting |
| NCT03561506 | Effect of Preoperative Ferric Carboxymaltose After Simultaneous Bilateral Total Knee Arthroplasty | 2018 | Unknown (last recruitment status was 'Not yet recruiting', verified June 2018; study investigators contacted, but not response obtained) |
| NCT03560687 | CardioSideral Heart Surgery: randomized Study on Sucrosomial Iron Supplementation Before Heart Surgery | 2018 | Completed |
| NCT03565354 / ChiCTR1800017010 | Efficacy of Preoperative Intravenous Iron in Anaemic Colorectal Cancer Surgical Patients | 2018 | Completed |
| ChiCTR1900022790 | Peri-operative Effect of EPO Combined with Iron in Total Hip Arthroplasty: A prospective Randomized, Controlled Trial | 2019 | Recruiting |
| ChiCTR1900026064 | Effect of recombinant human erythropoietin combined with iron on anemia treatment indifferent time points during perioperative period of total hip arthroplasty: a prospective randomized controlled trial | 2019 | Recruiting |
| NCT04083755 / EUCTR2018-003714-40 | Impact of intravenous iron treatment of preoperative anemia in patients with lower extremity peripheral artery disease (IRONPAD) | 2019 | Recruiting |
| IRCT20170705034908N3 | Comparison of the Effectiveness of Ferric carboxymaltose versus Iron sucrose in Anemia correction/impact on Transfusion Requirements in patients undergoing cardiac surgery | 2019 | Recruitment complete |
| IRCT20190121042447N1 | Evaluating the efficacy of Erythropoietin and Intravenous Iron on transfusion requirements in patients undergoing cardiac surgery | 2019 | Recruitment complete |
| KCT0004015 | Can Preoperative Ferric Carboxymaltose (FCM) Reduce Postoperative Blood Transfusion in Bilateral Total Knee Arthroplasty : Randomized controlled trial | 2019 | Not yet recruiting |
| NCT03915327 | Short-term Intravenous Iron Dextran for IDA | 2019 | Not yet recruiting |
| NCT04168346 | Preoperative Intravenous Iron Therapy in Patients With Gastric Cancer (IRONSTOMACH) | 2019 | Not yet recruiting |
| NCT04087993 | Polyglucoferron Compared to i.v. Ferric Carboxymaltose and Oral Iron Substitution in Preoperative Treatment of Iron Deficiency Anaemia in Patients (IDA-I) | 2019 | Suspended (difficult recruitment - redesign in discussion) |
| NCT04475497 | Role of Blood Management in Perioperative Outcomes | 2020 | Recruiting |
| NCT04351607 | Preoperative Supplementation of Sucrosomal Iron as Hematopoietic Support. (Preop Iron) | 2020 | Recruiting |
| **Prematurely ended** | | | |
| NCT00199277 | Iron Therapy in Colo-Rectal Neoplasm and Iron Deficiency Anemia: intravenous Iron Sucrose Versus Oral Ferrous Sulphate | 2005 | Never really began  (personal communication with García-Erce) |
| NCT00706667 / EUCTR2009-015799-92-AT | Intravenous Ferric Carboxymaltose (Ferinject®) With or Without Erythropoietin in Patients Undergoing Orthopaedic Surgery | 2008 | Terminated; not able to recruit sufficient patients due to lack of compliance |
| NCT01345968 | Intravenous Ferric Carboxymaltose (Ferinject) in Patients Undergoing Orthopaedic Surgery | 2011 | Terminated; less patients than expected for inclusion, therefore recruitment level too low |
| EUCTR2013-004979-13-DK | A Placebo Study Comparing Intravenous Iron with Saline in Treatment of Low Blood Count before Surgery in Patients with Cancer of the Kidney, Bladder or Lower Abdominal Cavity | 2013 | Prematurely ended; insufficient participants (personal communication with Norgaard) |
| EUCTR2014-001518-25-DE | A study of intravenous iron isomaltoside 1000 (Monofer®) compared to placebo in subjects with iron deficiency anaemia who are Intolerant or unresponsive to oral iron therapy | 2014 | Prematurely ended |
| NCT02189889 | Active Preoperative Anemia Management in Patients Undergoing Cardiac Surgery | 2014 | Terminated; no enrollment since the new hospital opened as there’s no infusion room |
| **PUBLISHED PROTOCOL PAPERS** | | | |
| 1. Borstlap WAA, Buskens CJ, Tytgat KMAJ, Tuynman JB, Consten ECJ, Tolboom RC, Heuff G, van Geloven N, van Wagensveld BA, C A Wientjes CA, Gerhards MF, de Castro SMM, Jansen J, van der Ven AWH, van der Zaag E, Omloo JM, van Westreenen HL, Winter DC, Kennelly RP, Dijkgraaf MGW, Tanis PJ, Bemelman WA. *Multicentre randomized controlled trial comparing ferric (III) carboxymaltose infusion with oral iron supplementation in the treatment of preoperative anaemia in colorectal cancer patients.* BMC Surg 2015, 15:78. doi: 10.1186/s12893-015-0065-6. 2. Lee SH, Shim JK, Soh S, Song JW, Chang BC, Lee S, Kwak YL. *The effect of perioperative intravenously administered iron isomaltoside 1000 (Monofer(R)) on transfusion requirements for patients undergoing complex valvular heart surgery: study protocol for a randomized controlled trial*. Trials 2018, 19(1):350. doi: 10.1186/s13063-018-2545-3. | | | |

### Supplementary Table 4. Overview of adverse events for which data were obtained from the 26 included randomized controlled trials (RCTs).

| **Adverse event category** | **Outcome** | **Included RCTs** | | | | | | | | | | | | | | | | | | | | | | | | | | |
| --- | --- | --- | --- | --- | --- | --- | --- | --- | --- | --- | --- | --- | --- | --- | --- | --- | --- | --- | --- | --- | --- | --- | --- | --- | --- | --- | --- | --- |
|  |  | Biboulet 2018 [1] | Cao 2020 [2] | Christodoulakis 2005 [3] | de Andrade 1996 [4] | Dickson 2020 [5] | Dousias 2003 [6] | Dousias 2005 [7] | Froessler 2016 [8] | Heiss 1996 [9] | Keeler 2017 [10] | Keeler 2019 [11] | Kettelhack 1998 [12] | Khalafallah 2012 [13] | Kim 2009 [14] | Kosmadakis 2003 [15] | Larson 2001 [16] | Lee 2019 [17] | Olijhoek 2001 [18] | Padmanabhan 2019 [19] | Qvist 1999/2000 [20, 21] | Richards 2020 [22] | Scott 2002 [23] | So-Osman 2014 [24] | Stowell 2009 [25] | Urena 2017 [26] | Weltert 2015 [27] | Yoo 2011 [28] |
| Gastro-intestinal | Dyspepsia |  |  |  |  |  |  |  |  |  | 1 |  |  |  | 1 |  |  |  |  |  |  |  |  |  |  |  |  |  |
|  | Nausea |  | 4 |  |  |  |  |  |  |  |  |  |  |  | 1 |  |  |  |  |  |  |  |  |  | 4 |  |  | 4 |
|  | Vomitus |  | 4 |  |  |  |  |  |  |  |  |  |  |  |  |  |  |  |  |  |  |  |  |  | 4 |  |  | 4 |
|  | Obstipation |  |  |  |  |  |  |  |  |  | 1 |  | 4 |  |  |  |  |  |  |  |  |  |  |  | 4 |  |  |  |
|  | Diarrhoea |  |  |  |  |  |  |  |  |  |  |  |  |  |  |  |  |  |  |  |  |  |  |  |  |  |  | 4 |
|  | Flatulence |  |  |  |  |  |  |  |  |  |  |  |  |  |  |  |  |  |  |  |  |  |  |  |  |  |  |  |
|  | Abdominal pain |  |  |  |  |  |  |  |  |  |  |  |  |  |  |  |  |  |  |  |  |  |  |  |  | 4 |  |  |
|  | Gastrointestinal reaction |  |  |  |  |  |  |  |  |  |  |  |  |  |  |  |  |  |  |  |  |  |  |  |  |  |  |  |
|  | (Incomplete) intestinal obstruction |  |  |  |  |  |  |  |  |  |  |  |  |  |  |  |  |  |  |  |  |  |  |  |  |  |  |  |
|  | Gastrointestinal symptoms (incl. diarrhoea and constipation) |  |  |  |  |  |  |  |  |  |  |  |  |  |  |  |  |  |  | 1 |  |  |  |  |  |  |  |  |
|  | Digestive complications | 5 |  |  |  |  |  |  |  |  |  |  |  |  |  |  |  |  |  |  |  |  |  |  |  |  |  |  |
| Muco-cutaneous | Rash |  |  | 4 |  |  |  |  |  |  | 1 |  |  |  |  |  |  |  |  |  |  |  |  |  |  |  |  |  |
|  | Pruritus |  |  |  |  |  |  |  |  |  |  |  |  |  |  |  |  |  |  |  |  |  |  |  | 4 |  |  |  |
|  | Urticaria |  |  | 4 |  |  |  |  |  |  |  |  |  |  |  |  |  |  |  |  |  |  |  |  |  |  |  |  |
|  | Erythema |  |  |  |  |  |  |  |  |  |  |  |  |  |  |  |  |  |  |  |  |  |  |  |  |  |  |  |
|  | Palor |  |  |  |  |  |  |  |  |  |  |  |  |  |  |  |  |  |  |  |  |  |  |  |  |  |  |  |
|  | Flush |  |  |  |  |  |  |  |  |  |  |  |  |  |  |  |  |  |  |  |  |  |  |  |  |  |  |  |
|  | Eczema |  |  |  |  |  |  |  |  |  |  |  |  |  |  |  |  |  |  |  |  |  |  |  |  |  |  |  |
| Autonomic | Fever |  |  |  |  |  |  |  |  |  |  |  |  |  |  |  |  |  |  |  |  |  |  |  |  |  |  |  |
|  | Pyrexia rate (30-day, 90-day, 1-year, overall) |  |  |  |  |  |  |  |  |  |  |  |  |  |  |  |  |  |  |  |  |  |  |  | 4 |  |  |  |
|  | Febrile episodes |  |  |  |  |  | 4 |  |  |  |  |  |  |  |  |  |  |  |  |  |  |  |  |  |  |  |  |  |
|  | Postoperative fever |  |  |  |  |  |  | 4 |  |  |  |  |  |  |  |  |  |  |  |  |  |  |  |  |  |  |  |  |
|  | Chills with fever |  |  |  |  |  |  |  |  |  |  |  | 4 |  |  |  |  |  |  |  |  |  |  |  |  |  |  |  |
| Neuro-psycho-somatic | Post-infusion headache |  |  |  |  |  |  |  |  |  | 1 |  |  |  |  |  |  |  |  |  |  |  |  |  |  |  |  |  |
|  | Headache |  |  |  |  |  |  |  | 2 |  |  |  |  |  |  |  |  |  |  |  |  |  |  |  | 4 |  |  |  |
|  | Myalgia |  |  |  |  |  |  |  |  |  |  |  |  |  | 1 |  |  |  |  |  |  |  |  |  |  |  |  |  |
|  | Injection pain |  |  |  |  |  |  |  |  |  |  |  |  |  | 1 |  |  |  |  |  |  |  |  |  |  |  |  |  |
|  | Light-headedness |  |  |  |  |  |  |  | 2 |  |  |  |  |  |  |  |  |  |  |  |  |  |  |  |  |  |  |  |
|  | Back pain |  |  |  |  |  |  |  | 2 |  |  |  |  |  |  |  |  |  |  |  |  |  |  |  | 4 |  |  |  |
|  | Chest pain |  |  |  |  |  |  |  |  |  |  |  |  |  |  |  |  |  |  |  |  |  |  |  | 4 |  |  |  |
|  | Post-procedural pain |  |  |  |  |  |  |  |  |  |  |  |  |  |  |  |  |  |  |  |  |  |  |  | 4 |  |  |  |
| Neuro-logical | Postoperative ileus |  |  |  |  |  |  |  |  |  |  |  |  |  |  |  |  |  |  |  |  |  |  |  |  |  |  |  |
|  | Cephalgia |  |  |  |  |  |  |  |  |  |  |  |  |  |  |  |  |  |  |  |  |  |  |  |  |  |  |  |
|  | Vertigo |  |  |  |  |  |  |  |  |  |  |  |  |  |  |  |  |  |  |  |  |  |  |  |  |  |  |  |
|  | Paraesthesia |  |  |  |  |  |  |  |  |  |  |  |  |  |  |  |  |  |  |  |  |  |  |  |  |  |  |  |
|  | Dysgeusia |  |  |  |  |  |  |  |  |  |  |  |  |  |  |  |  |  |  |  |  |  |  |  |  |  |  |  |
|  | Neurological complications at 45 days |  |  |  |  |  |  |  |  |  |  |  |  |  |  |  |  |  |  |  |  |  |  |  |  |  | 4 |  |
|  | Focal neurologic damage |  |  |  |  |  |  |  |  |  |  |  |  |  |  |  |  |  |  |  |  |  |  |  |  |  | 4 |  |
|  | Generalized neurologic damage |  |  |  |  |  |  |  |  |  |  |  |  |  |  |  |  |  |  |  |  |  |  |  |  |  | 4 |  |
| Wound healing | Poor wound healing |  |  |  |  |  |  |  |  |  |  |  |  |  |  |  |  |  |  |  |  |  |  |  |  |  |  |  |
|  | (Delayed) suture/wound dehiscence |  |  |  |  | 1 |  |  |  |  |  |  |  |  |  |  |  |  |  |  |  |  |  |  |  |  |  |  |
| Broncho-pulmonary | Dyspnoea |  |  |  |  |  |  |  |  |  |  |  |  |  |  |  |  |  |  |  |  |  |  |  |  |  |  |  |
|  | Hyperventilation |  |  |  |  |  |  |  |  |  |  |  |  |  |  |  |  |  |  |  |  |  |  |  |  |  |  |  |
|  | Pneumonia/respiratory failure |  |  |  |  |  |  |  |  |  |  |  |  |  |  |  |  |  |  |  |  |  |  |  |  |  |  |  |
|  | Respiratory failure |  |  |  |  |  |  |  | 2 |  |  |  |  |  |  |  |  |  |  |  |  |  | 4 |  |  |  |  |  |
|  | Acute respiratory distress syndrome |  |  |  |  |  |  |  |  |  |  |  |  |  |  |  |  |  |  |  |  |  | 4 |  |  |  | 4 |  |
|  | (Acute) pulmonary oedema |  |  |  |  |  |  |  |  |  |  |  |  |  |  |  |  |  |  |  |  |  |  |  |  |  | 4 |  |
|  | Need for re-intubation |  |  |  |  |  |  |  |  |  |  |  |  |  |  |  |  |  |  |  |  |  |  |  |  |  |  |  |
|  | Prolonged ventilation |  |  |  |  |  |  |  |  |  |  |  |  |  |  |  |  |  |  |  |  |  |  |  |  |  |  |  |
| Infectious | Preoperative prostatitis | 5 |  |  |  |  |  |  |  |  |  |  |  |  |  |  |  |  |  |  |  |  |  |  |  |  |  |  |
|  | Infection (30-day, 90-day, 1-year, overall) |  |  |  |  |  |  |  | 2 |  |  |  |  |  |  |  |  |  |  |  |  |  |  |  |  |  |  |  |
|  | Postoperative infection |  |  |  |  |  |  |  |  |  |  | 1 |  |  |  |  |  |  |  | 1 |  |  |  |  |  |  |  |  |
|  | (Surgical/superficial) wound infection |  |  |  |  |  |  |  |  |  |  |  |  |  |  |  | 4 |  |  |  |  |  |  |  | 4 |  | 4 |  |
|  | Intra-abdominal abscess |  |  |  |  |  |  |  |  |  |  |  |  |  |  |  |  |  |  |  |  |  |  |  |  |  |  |  |
|  | Urinary tract infection |  |  |  |  |  |  |  |  |  |  |  |  |  |  |  | 4 |  |  |  |  |  |  |  | 4 |  |  |  |
|  | Infection of the vaginal stump |  |  |  |  |  |  |  |  |  |  |  |  |  |  |  |  |  |  |  |  |  |  |  |  |  |  |  |
|  | Cellulitis after surgery |  |  |  |  |  |  |  |  |  |  |  |  |  |  |  |  |  |  |  |  |  |  |  |  |  |  |  |
|  | Superficial thrombophlebitis |  |  |  |  |  |  |  |  |  |  |  |  |  |  |  |  |  |  |  |  |  |  |  | 4 |  |  |  |
|  | Sepsis rate (30-day, 90-day, 1-year, overall ) or septic shock |  |  |  |  |  |  |  |  | 4 |  |  | 4 |  |  |  | 4 |  |  |  |  |  |  |  |  | 4 |  |  |
|  | Pneumonia (30-day, 90-day, 1-year, overall) |  |  |  |  |  |  |  |  |  |  |  |  |  |  |  |  |  |  |  |  |  |  |  |  |  | 4 |  |
|  | Peritonitis |  |  | 4 |  |  |  |  |  |  |  |  |  |  |  |  |  |  |  |  |  |  |  |  |  |  |  |  |
|  | Endocarditis |  |  |  |  |  |  |  |  |  |  |  |  |  |  |  |  |  |  |  |  |  |  |  |  |  |  |  |
|  | Severe infection (sepsis, pneumonia or mediastinitis) |  |  |  |  |  |  |  |  |  |  |  |  |  |  |  |  |  |  |  |  |  |  |  |  |  |  |  |
|  | Grade of infective complication severity |  |  |  |  |  |  |  |  |  | 1 |  |  |  |  |  |  |  |  |  |  |  |  |  |  |  |  |  |
|  | Infective complication rate |  |  |  |  |  |  |  |  |  |  |  |  | 1 |  |  |  |  |  |  |  |  |  |  |  |  |  |  |
|  | Prevalence of infectious-related codes during hospital stay |  |  |  |  |  |  |  |  |  |  |  |  |  |  |  |  |  |  |  |  |  |  |  |  |  |  |  |
|  | Readmission rate for general infection (discharge to 8 weeks) |  |  |  |  |  |  |  |  |  |  |  |  |  |  |  |  |  |  |  |  | 2 |  |  |  |  |  |  |
|  | Readmission rate for wound infection (discharge to 8 weeks) |  |  |  |  |  |  |  |  |  |  |  |  |  |  |  |  |  |  |  |  | 2 |  |  |  |  |  |  |
| Bleeding | Bleeding |  |  |  |  |  |  |  |  |  |  |  |  |  |  |  |  |  |  |  |  |  |  |  |  |  |  |  |
|  | Upper gastrointestinal bleed |  |  |  |  |  |  |  |  |  |  |  |  |  |  |  |  |  |  |  |  |  |  |  |  |  |  |  |
|  | Surgical wound hematoma |  |  |  |  |  |  |  |  |  |  |  |  |  |  |  |  |  |  |  |  |  |  |  |  |  |  |  |
|  | Postoperative hematoma |  |  |  |  |  |  |  |  |  |  |  |  |  |  |  |  |  |  |  |  |  |  |  |  |  |  |  |
|  | Rectorrhagia/hemoperitoneum |  |  |  |  |  |  |  |  |  |  |  |  |  |  |  |  |  |  |  |  |  |  |  |  |  |  |  |
|  | Anastomosis rupture |  |  |  |  |  |  |  |  |  |  |  |  |  |  |  |  |  |  |  |  |  | 4 |  |  |  |  |  |
|  | Hemarthrosis |  |  |  |  |  |  |  |  |  |  |  |  |  |  |  |  |  |  |  |  |  |  |  |  |  |  |  |
|  | Major or life-threatening bleeding |  |  |  |  |  |  |  |  |  |  |  |  |  |  |  |  |  |  |  |  |  |  |  |  | 4 |  |  |
|  | Haemorrhagic shock |  |  |  |  |  |  |  |  |  |  |  |  |  |  |  |  |  |  |  |  |  |  |  |  |  |  |  |
| Cardio-vascular | Atrial fibrillation (requiring readmission) |  |  | 4 |  | 1 |  |  |  |  |  |  |  |  |  |  |  |  |  | 1 |  |  |  |  |  | 4 | 4 | 4 |
|  | Cardiac failure (30-day, 90-day, 1-year, overall) |  |  | 4 |  |  |  |  |  |  |  |  | 4 |  |  |  |  |  |  |  |  |  |  |  |  | 4 |  |  |
|  | Cardiac or respiratory failure | 5 |  |  |  |  |  |  |  |  |  |  |  |  |  |  |  |  |  |  |  |  |  |  |  |  |  |  |
|  | Cardiac arrest |  |  | 4 |  |  |  |  |  |  |  |  |  |  |  |  |  |  |  |  |  |  |  |  |  |  |  |  |
|  | Perioperative myocardial infarction |  |  |  |  |  |  |  |  |  |  |  |  |  |  |  |  |  |  |  |  |  |  |  |  |  |  |  |
|  | Myocardial injury: peak of troponin and creatine kinase-MB |  |  |  |  |  |  |  |  |  |  |  |  |  |  |  |  |  |  |  |  |  |  |  |  | 4 |  |  |
|  | Acute coronary syndrome |  |  |  |  |  |  |  |  |  |  |  |  |  |  |  |  |  |  |  |  |  |  |  |  |  |  |  |
|  | Arrhythmia |  |  |  |  |  |  |  |  |  |  |  |  |  |  |  |  |  |  |  |  |  |  |  |  |  |  |  |
|  | Tachycardia |  |  |  |  |  |  |  |  |  |  |  |  |  |  |  |  |  |  |  |  |  |  |  |  |  |  | 4 |
|  | Hypertension |  |  |  |  |  | 4 |  |  |  |  |  |  |  |  | 4 |  |  |  |  |  |  | 4 |  |  | 4 | 4 | 4 |
|  | Cardiac tamponade |  |  |  |  |  |  |  |  |  |  |  |  |  |  |  |  |  |  |  |  |  |  |  |  | 4 | 4 |  |
|  | Annulus rupture |  |  |  |  |  |  |  |  |  |  |  |  |  |  |  |  |  |  |  |  |  |  |  |  | 4 |  |  |
|  | Coronary artery occlusion |  |  |  |  |  |  |  |  |  |  |  |  |  |  |  |  |  |  |  |  |  |  |  |  | 4 |  |  |
|  | Moderate aortic regurgitation |  |  |  |  |  |  |  |  |  |  |  |  |  |  |  |  |  |  |  |  |  |  |  |  | 4 |  |  |
|  | Severe hypotension requiring haemodynamic support |  |  |  |  |  |  |  |  |  |  |  |  |  |  |  |  |  |  |  |  |  |  |  |  | 4 |  |  |
|  | Shock with hypotension |  |  |  |  |  |  |  |  |  |  |  |  |  |  |  |  |  |  |  |  |  |  |  |  |  |  |  |
|  | Cardiovascular arrest |  |  |  |  |  |  |  |  |  |  |  |  |  |  |  |  |  |  |  |  |  |  |  |  |  |  |  |
|  | Permanent cerebral vascular accident |  |  |  |  |  |  |  |  |  |  |  |  |  |  |  |  |  |  |  |  |  |  |  |  |  |  |  |
|  | Oedema (30-day) |  |  |  |  |  |  |  |  |  |  |  |  |  |  |  |  |  |  |  |  |  |  |  |  |  |  |  |
|  | Angioedema |  |  |  |  |  |  |  |  |  |  |  |  |  |  |  |  |  |  |  |  |  |  |  |  |  |  |  |
|  | Vascular complications |  |  |  |  |  |  |  |  |  |  |  |  |  |  |  |  |  |  |  |  |  |  |  |  | 4 |  |  |
|  | Cardiovascular events |  | 4 |  |  |  |  |  |  |  |  |  |  |  |  |  |  |  |  |  |  |  |  |  |  |  |  |  |
|  | Major adverse cardiovascular events |  |  |  |  |  |  |  |  |  |  |  |  |  |  |  |  |  |  |  |  |  |  |  |  |  |  |  |
| Renal | Need for renal replacement therapy |  |  |  |  |  |  |  |  |  |  |  |  |  |  |  |  |  |  | 1 |  |  |  |  |  |  | 4 |  |
|  | Need for dialysis |  |  |  |  |  |  |  |  |  |  |  |  |  |  |  |  |  |  |  |  |  |  |  |  | 4 |  |  |
|  | Renal impairment |  |  |  |  |  |  |  | 2 |  |  |  |  |  |  |  |  |  |  |  |  |  |  |  |  |  |  |  |
|  | Renal failure rate (30-day, 90-day, 1-year; new onset, acute) |  |  |  |  |  |  |  |  |  |  |  |  |  |  |  |  |  |  |  |  |  |  |  |  |  | 4 |  |
| Anaemia-associated ischemic events | Myocardial infarction (30-day, 90-day, 1-year, overall) |  |  |  |  |  |  |  |  |  |  |  |  |  |  |  |  |  |  |  |  |  | 4 | 4 | 4 | 4 | 4 |  |
|  | Myocardial ischemia |  |  |  |  |  |  |  |  |  |  |  |  |  |  |  |  |  |  |  |  |  |  |  | 4 |  |  |  |
|  | Stroke (30-day, 90-day, 1-year, overall) |  |  |  |  |  |  |  |  |  |  |  |  |  |  |  |  |  |  |  |  |  | 4 |  | 4 | 4 |  |  |
|  | TIA |  |  |  |  |  |  |  |  |  |  |  |  |  |  |  |  |  |  |  |  |  |  |  | 4 |  |  |  |
|  | Stroke or TIA |  |  |  |  |  |  |  |  |  |  |  |  |  |  |  |  |  |  |  |  |  |  | 4 |  |  |  |  |
|  | Bowel ischemia |  |  |  |  |  |  |  |  |  |  |  |  |  |  |  |  |  |  |  |  |  |  |  |  |  | 4 |  |
|  | Acute kidney injury |  |  |  |  |  |  |  |  |  |  |  |  |  |  |  |  |  |  |  |  | 2 |  |  |  | 4 |  | 4 |
|  | Acute limb ischemia |  |  |  |  |  |  |  |  |  |  |  |  |  |  |  |  |  |  |  |  |  |  |  |  |  |  |  |
|  | Ischemic events |  |  |  |  |  |  |  |  |  |  |  |  |  |  |  |  |  |  |  |  |  |  |  |  | 4 |  |  |
| Thrombo-embolic events | Deep venous thrombosis (requiring readmission) | 5 | 4 |  | 4 | 1 |  |  | 2 | 4 |  |  |  |  |  | 4 |  |  |  |  | 4 |  | 4 | 4 | 4 |  | 4 |  |
|  | Preoperative femoral vein thrombosis | 5 |  |  |  |  |  |  |  |  |  |  |  |  |  |  |  |  |  |  |  |  |  |  |  |  |  |  |
|  | Mesenteric venous thrombosis |  |  |  |  |  |  |  |  | 4 |  |  |  |  |  |  |  |  |  |  |  |  |  |  |  |  |  |  |
|  | Arterial thrombosis |  |  |  |  |  |  |  |  |  |  |  | 4 |  |  |  |  |  |  |  |  |  |  |  |  |  |  |  |
|  | Thrombosis |  |  |  |  |  |  |  |  |  |  |  |  |  |  |  |  |  |  |  |  |  |  |  |  |  |  |  |
|  | Pulmonary embolism |  |  |  |  |  |  |  |  |  |  |  |  |  |  |  |  |  |  |  |  |  |  | 4 | 4 |  |  |  |
|  | Embolism |  |  | 4 |  |  |  |  |  |  |  |  |  |  |  |  |  |  |  |  |  |  |  |  |  |  |  |  |
|  | Valve embolization |  |  |  |  |  |  |  |  |  |  |  |  |  |  |  |  |  |  |  |  |  |  |  |  | 4 |  |  |
|  | Thrombotic and/or vascular events |  |  |  |  |  |  |  |  |  |  |  |  |  |  |  |  |  | 1,4,5 |  |  |  |  |  |  |  |  |  |
| Mortality or survival | Mortality | 5 |  | 4 |  |  |  |  | 2 | 4 | 1 |  | 4 |  | 1 (€) | 4 |  | 3 | 1,4,5 | 1 |  | 2 | 4 |  |  |  | 4 |  |
|  | Survival |  |  |  |  | 1 |  |  |  |  |  |  |  |  |  |  |  |  |  |  |  |  |  |  |  |  |  |  |
| Other | Allergy |  | 4 |  |  |  |  |  |  |  |  |  |  |  |  | 4 |  |  |  |  |  |  |  |  |  |  |  |  |
|  | Severe allergic reactions |  |  |  |  |  |  |  |  |  |  |  |  |  |  |  |  |  |  |  |  |  |  |  |  |  |  |  |
|  | Anaphylactic reactions |  |  |  |  |  |  |  |  |  |  |  |  |  |  |  |  | 3 |  |  |  |  |  |  |  |  |  |  |
|  | Lymph node swelling |  |  |  |  |  |  |  |  |  |  |  |  |  |  |  |  |  |  |  |  |  |  |  |  |  |  |  |
|  | Acute myeloid leukemia |  |  |  |  |  |  |  |  |  |  |  |  |  |  |  |  |  |  |  |  |  |  |  | 4 |  |  |  |
|  | Grade 2 neutropenia |  |  | 4 |  |  |  |  |  |  |  |  |  |  |  |  |  |  |  |  |  |  |  |  |  |  |  |  |
|  | Cachexia |  |  |  |  |  |  |  |  |  |  |  | 4 |  |  |  |  |  |  |  |  |  |  |  |  |  |  |  |
|  | Multiorgan failure |  |  |  |  |  |  |  |  | 4 |  |  |  |  |  |  |  |  |  |  |  |  |  |  |  |  |  |  |
|  | Epileptic seizure |  |  |  |  |  |  |  |  |  |  |  |  |  |  |  |  |  |  |  |  |  |  |  |  |  |  |  |
|  | Convulsions |  |  |  |  |  |  |  |  |  |  |  |  |  |  |  |  |  |  |  |  |  |  |  |  | 4 |  |  |
|  | Muscle spasms |  |  |  |  |  |  |  |  |  |  |  |  |  |  |  |  |  |  |  |  |  |  |  | 4 |  |  |  |
|  | Mild and transient hypertonia |  |  |  |  |  |  |  |  | 4 |  |  |  |  |  |  |  |  |  |  |  |  |  |  |  |  |  |  |
|  | Hypercalcemia |  |  |  |  |  |  |  |  |  |  |  |  |  |  |  |  |  |  |  |  |  |  |  |  |  |  | 4 |
|  | Hypokalemia |  |  |  |  |  |  |  |  |  |  |  |  |  |  |  |  |  |  |  |  |  |  |  | 4 |  |  |  |
|  | Insomnia |  |  |  |  |  |  |  |  |  |  |  |  |  |  |  |  |  |  |  |  |  |  |  | 4 |  |  |  |
|  | Anastomotic leak (requiring readmission) |  |  |  |  | 1 |  |  |  |  |  |  |  |  |  |  |  |  |  |  |  |  |  |  |  |  |  |  |
|  | Chyle leak |  |  |  |  |  |  |  |  |  |  |  |  |  |  |  |  |  |  |  |  |  |  |  |  |  |  |  |
|  | Intra-abdominal collection of fluids |  |  |  |  |  |  |  |  |  |  |  |  |  |  |  |  |  |  |  |  |  |  |  |  |  |  |  |
|  | Conversion to open heart surgery |  |  |  |  |  |  |  |  |  |  |  |  |  |  |  |  |  |  |  |  |  |  |  |  | 4 |  |  |
|  | Need for a second valve |  |  |  |  |  |  |  |  |  |  |  |  |  |  |  |  |  |  |  |  |  |  |  |  | 4 |  |  |
|  | Thrombosis or dysfunction of the prosthesis |  |  |  |  |  |  |  |  |  |  |  |  |  |  |  |  |  |  |  |  |  |  |  |  |  |  |  |
|  | Need for re-exploration |  |  |  |  |  |  |  |  |  |  |  |  |  |  |  |  |  |  | 1 |  |  |  |  |  |  |  |  |
|  | Reoperation |  |  |  |  |  |  |  |  |  |  |  |  |  |  |  |  |  |  |  |  |  |  |  |  |  |  |  |
|  | Hospital readmission |  |  |  |  |  |  |  | 2 |  |  |  |  |  |  |  |  |  |  |  |  |  |  |  |  |  |  |  |
|  | Hypersensitivity reactions |  |  |  |  |  |  |  |  |  |  |  |  |  |  |  |  |  |  |  |  |  |  |  |  | 4 |  |  |
|  | Readmission for general postoperative complications between discharge and 8 weeks |  |  |  |  |  |  |  |  |  |  |  |  |  |  |  |  |  |  |  |  | 2 |  |  |  |  |  |  |
|  | Readmission for complications between discharge and 6 months |  |  |  |  |  |  |  |  |  |  |  |  |  |  |  |  |  |  |  |  | 2 |  |  |  |  |  |  |
|  | Grade of complication severity from recruitment to outpatients |  |  |  |  |  |  |  |  |  | 1 |  |  |  |  |  |  |  |  |  |  |  |  |  |  |  |  |  |
|  | Complication rate from recruitment to outpatients |  |  |  |  |  |  |  |  |  |  | 1 |  |  |  |  |  |  |  |  |  |  |  |  |  |  |  |  |
|  | Postoperative complication rate ≥ Clavien-Dindo grade III |  |  |  |  |  |  |  |  |  |  |  |  |  |  |  |  |  |  |  |  | 2 |  |  |  |  |  |  |
|  | Medical postoperative complications |  |  |  |  |  |  |  |  |  |  |  |  |  |  |  |  |  |  |  |  |  |  |  |  |  |  |  |
|  | Surgical postoperative complications |  |  |  |  |  |  |  |  |  |  |  |  |  |  |  |  |  |  |  |  |  |  |  |  |  |  |  |
|  | Non-thromboembolic complications (composite of prosthesis-related, cardiovascular, allergic, infectious, bleeding etc events) |  |  |  |  |  |  |  |  |  |  |  |  |  |  |  |  |  |  |  |  |  |  | 4 |  |  |  |  |
|  | Postoperative overall complication rate (composite of pulmonic, cardiologic, thrombotic, infectious and neurologic complications) |  |  |  |  |  |  |  |  |  |  |  |  |  |  |  |  |  |  |  |  |  |  |  |  |  |  |  |
|  | Postoperative complicate rate (composite of anastomotic leak, abscess/fistula formation, haemorrhage, wound infection, pulmonary complications, complications from blood transfusions) |  |  |  |  |  |  |  |  |  |  |  |  |  |  | 4 |  |  |  |  |  |  |  |  |  |  |  |  |

Cells shaded in green indicate that the corresponding RCT provided data for the corresponding AE outcome(s). The number mentioned in the green-shaded cell represents the number of the treatment comparison. Comparison 1: Intravenous (IV) iron versus oral iron monotherapy, Comparison 2: IV iron versus usual care/no iron, Comparison 3: IV ferric carboxymaltose versus IV iron sucrose monotherapy, Comparison 4: ESA + iron versus control (placebo and/or iron, no treatment), Comparison 5: ESA + IV iron versus ESA + oral iron, Comparison 6: ESA + IV iron versus ESA + IV iron (different ESA dosing regimens).

If there are no green-shaded cells across an entire row, this means that data on this AE outcome were provided by cohort studies (see Table E).

(€) indicates that additional unpublished data were obtained from the study authors. This enabled the inclusion of data, that otherwise would have been disregarded, on 1 outcome across 1 AE category.

### Supplementary Table 5. Overview of adverse events for which data were obtained from the 16 included cohort studies.

| **Adverse event category** | **Outcome** | **Included cohort studies** | | | | | | | | | | | | | | | |
| --- | --- | --- | --- | --- | --- | --- | --- | --- | --- | --- | --- | --- | --- | --- | --- | --- | --- |
|  |  | Cladellas 2012 [29] | Delasotta 2012 (A) [30] | Delasotta 2012 (B) [31] | Ellermann 2018 [32] | Evans 2021 [33] | Kam 2020 [34] | Klein 2020 [35] | Laso-Morales 2017 [36] | Nandhra 2020 [37] | Pinilla-Gracia 2020 [38] | Quinn 2017 [39] | Rineau 2017 [40] | Triphaus 2019 [41] | Wilson 2018 (A) [42] | Wilson 2018 (B) [43] | Ye 2017 [44] |
| Gastro-intestinal | Dyspepsia |  |  |  | 2 (£) |  |  |  |  |  |  |  |  |  |  |  |  |
|  | Nausea |  |  |  | 2 (£) |  |  | 2 (#) |  |  |  |  |  |  |  |  |  |
|  | Vomitus |  |  |  | 2 (£) |  |  |  |  |  |  |  |  |  |  |  |  |
|  | Obstipation |  |  |  | 2 (£) |  |  |  |  |  |  |  |  |  |  |  |  |
|  | Diarrhoea |  |  |  | 2 (£) |  |  |  |  |  |  |  |  |  |  |  |  |
|  | Flatulence |  |  |  | 2 (£) |  |  |  |  |  |  |  |  |  |  |  |  |
|  | Abdominal pain |  |  |  | 2 (£) |  |  |  |  |  |  |  |  |  |  |  |  |
|  | Gastrointestinal reaction |  |  |  |  |  |  |  |  |  |  |  |  |  |  |  | 2, 4 |
|  | (Incomplete) intestinal obstruction |  |  |  |  |  |  |  |  |  |  |  | 6 ($) |  |  |  | 2, 4 |
|  | Gastrointestinal symptoms (incl. diarrhoea and constipation) |  |  |  |  |  |  |  |  |  |  |  |  | 2 (§) |  |  |  |
|  | Digestive complications |  |  |  |  |  |  |  |  |  |  |  |  |  |  |  |  |
| Muco-cutaneous | Rash |  |  |  |  |  | 2 (ψ) |  |  |  |  |  |  |  |  |  |  |
|  | Pruritus |  |  |  | 2 (£) |  |  |  |  |  |  |  |  |  |  |  |  |
|  | Urticaria |  |  |  | 2 (£) |  |  |  |  |  |  |  |  |  |  |  |  |
|  | Erythema |  |  |  | 2 (£) |  |  |  |  |  |  |  |  |  |  |  |  |
|  | Palor |  |  |  | 2 (£) |  |  |  |  |  |  |  |  |  |  |  |  |
|  | Flush |  |  |  |  |  |  |  |  |  |  |  |  | 2 (§) |  |  |  |
|  | Eczema |  |  |  |  |  |  |  |  |  |  |  |  | 2 (§) |  |  |  |
| Autonomic | Fever |  |  |  |  |  | 2 (ψ) |  |  |  |  |  |  | 2 (§) |  |  | 2,4 |
|  | Pyrexia rate (30-day, 90-day, 1-year, overall) |  |  |  | 2 (£) |  |  |  |  |  |  |  |  |  |  |  |  |
|  | Febrile episodes |  |  |  |  |  |  |  |  |  |  |  |  |  |  |  |  |
|  | Postoperative fever |  |  |  |  |  |  |  |  |  |  |  |  |  |  |  |  |
|  | Chills with fever |  |  |  |  |  |  |  |  |  |  |  |  |  |  |  |  |
| Neuro-psycho-somatic | Post-infusion headache |  |  |  |  |  |  |  |  |  |  |  |  |  |  |  |  |
|  | Headache |  |  |  |  |  |  |  |  |  |  |  |  | 2 (§) |  |  | 2, 4 |
|  | Myalgia |  |  |  |  |  |  |  |  |  |  |  |  |  |  |  | 2, 4 |
|  | Injection pain |  |  |  |  |  |  |  |  |  |  |  |  |  |  |  |  |
|  | Light-headedness |  |  |  |  |  |  |  |  |  |  |  |  |  |  |  |  |
|  | Back pain |  |  |  |  |  |  |  |  |  |  |  |  |  |  |  |  |
|  | Chest pain |  |  |  |  |  |  |  |  |  |  |  |  |  |  |  |  |
|  | Post-procedural pain |  |  |  |  |  |  |  |  |  |  |  |  |  |  |  |  |
| Neuro-logical | Postoperative ileus |  |  |  |  |  | 2 |  | 2 |  |  | 1 (*) |  |  |  |  |  |
|  | Cephalgia |  |  |  | 2 (£) |  |  |  |  |  |  |  |  |  |  |  |  |
|  | Vertigo |  |  |  | 2 (£) |  |  |  |  |  |  |  |  |  |  |  |  |
|  | Paraesthesia |  |  |  | 2 (£) |  |  |  |  |  |  |  |  |  |  |  |  |
|  | Dysgeusia |  |  |  | 2 (£) |  |  |  |  |  |  |  |  |  |  |  |  |
|  | Neurological complications at 45 days |  |  |  |  |  |  |  |  |  |  |  |  |  |  |  |  |
|  | Focal neurologic damage |  |  |  |  |  |  |  |  |  |  |  |  |  |  |  |  |
|  | Generalized neurologic damage |  |  |  |  |  |  |  |  |  |  |  |  |  |  |  |  |
| Wound healing | Poor wound healing |  |  |  |  |  |  |  |  |  |  |  |  |  |  |  | 2, 4 |
|  | (Delayed) suture/wound dehiscence |  |  |  |  |  |  |  | 2 |  |  | 1 |  |  |  |  |  |
| Broncho-pulmonary | Dyspnoea |  |  |  | 2 (£) |  |  |  |  |  |  |  |  | 2 (§) |  |  |  |
|  | Hyperventilation |  |  |  |  |  |  |  |  |  |  |  |  | 2 (§) |  |  |  |
|  | Pneumonia/respiratory failure |  |  |  |  |  | 2 |  |  |  |  |  |  |  |  |  |  |
|  | Respiratory failure |  |  |  |  |  |  |  |  |  |  |  |  |  |  |  |  |
|  | Acute respiratory distress syndrome |  |  |  |  |  |  |  |  |  |  |  |  |  |  |  |  |
|  | (Acute) pulmonary oedema |  |  |  |  |  |  |  |  |  |  |  | 6 ($) |  |  |  |  |
|  | Need for re-intubation |  |  |  |  |  |  |  |  |  |  |  | 6 ($) |  |  |  |  |
|  | Prolonged ventilation | 4 |  |  |  |  |  |  |  |  |  |  | 6 ($) |  |  |  |  |
| Infectious | Preoperative prostatitis |  |  |  |  |  |  |  |  |  |  |  |  |  |  |  |  |
|  | Infection (30-day, 90-day, 1-year, overall) |  |  |  | 2 (£) |  |  |  |  |  |  |  |  |  |  |  |  |
|  | Postoperative infection |  |  |  |  |  |  |  |  |  |  |  |  |  |  |  |  |
|  | (Surgical/superficial) wound infection |  |  |  |  |  | 2 |  | 2 |  |  |  | 6 ($) |  |  |  |  |
|  | Intra-abdominal abscess |  |  |  |  |  |  |  | 2 |  |  |  |  |  |  |  |  |
|  | Urinary tract infection |  |  |  |  |  |  |  | 2 |  |  |  | 6 ($) |  |  |  |  |
|  | Infection of the vaginal stump |  |  |  |  |  |  |  |  |  |  |  |  |  |  |  | 2, 4 |
|  | Cellulitis after surgery |  |  | 4 |  |  |  |  |  |  |  |  |  |  |  |  |  |
|  | Superficial thrombophlebitis |  |  |  |  |  |  |  |  |  |  |  |  |  |  |  |  |
|  | Sepsis rate (30-day, 90-day, 1-year, overall ) or septic shock |  |  |  | 2 (£) |  |  |  |  |  |  |  |  |  |  |  |  |
|  | Pneumonia (30-day, 90-day, 1-year, overall) |  |  |  | 2 (£) |  |  |  | 2 |  |  |  | 6 ($) |  |  |  |  |
|  | Peritonitis |  |  |  |  |  |  |  |  |  |  |  |  |  |  |  |  |
|  | Endocarditis | 4 |  |  |  |  |  |  |  |  |  |  |  |  |  |  |  |
|  | Severe infection (sepsis, pneumonia or mediastinitis) | 4 |  |  |  |  |  |  |  |  |  |  |  |  |  |  |  |
|  | Grade of infective complication severity |  |  |  |  |  |  |  |  |  |  |  |  |  |  |  |  |
|  | Infective complication rate |  |  |  |  |  |  |  |  |  |  |  |  |  |  |  |  |
|  | Prevalence of infectious-related codes during hospital stay |  |  |  | 2 (£) |  |  |  |  |  |  |  |  |  |  |  |  |
|  | Readmission rate for general infection (discharge to 8 weeks) |  |  |  |  |  |  |  |  |  |  |  |  |  |  |  |  |
|  | Readmission rate for wound infection (discharge to 8 weeks) |  |  |  |  |  |  |  |  |  |  |  |  |  |  |  |  |
| Bleeding | Bleeding |  |  |  |  |  | 2 |  |  |  |  |  |  |  |  |  |  |
|  | Upper gastrointestinal bleed |  |  |  |  |  |  |  |  |  |  | 1 (*) |  |  |  |  |  |
|  | Surgical wound hematoma |  |  |  |  |  |  |  | 2 |  |  |  |  |  |  |  |  |
|  | Postoperative hematoma |  |  |  |  |  |  |  |  |  |  |  | 6 ($) |  |  |  |  |
|  | Rectorrhagia/hemoperitoneum |  |  |  |  |  |  |  | 2 |  |  |  |  |  |  |  |  |
|  | Anastomosis rupture |  |  |  |  |  |  |  |  |  |  |  |  |  |  |  |  |
|  | Hemarthrosis |  |  | 4 |  |  |  |  |  |  |  |  |  |  |  |  |  |
|  | Major or life-threatening bleeding |  |  |  |  |  |  |  |  |  |  |  |  |  |  |  |  |
|  | Haemorrhagic shock |  |  |  |  |  |  |  |  |  |  |  | 6 ($) |  |  |  |  |
| Cardio-vascular | Atrial fibrillation (requiring readmission) |  |  |  |  |  |  |  |  |  |  |  | 6 ($) |  |  |  |  |
|  | Cardiac failure (30-day, 90-day, 1-year, overall) | 4 |  |  | 2 (£) |  |  |  |  |  |  | 1 (*) |  |  |  |  |  |
|  | Cardiac or respiratory failure |  |  |  |  |  |  |  |  |  |  |  |  |  |  |  |  |
|  | Cardiac arrest |  |  | 4 |  |  |  |  |  |  |  |  | 6 ($) |  |  |  |  |
|  | Perioperative myocardial infarction | 4 |  |  |  |  |  |  |  |  |  |  |  |  |  |  |  |
|  | Myocardial injury: peak of troponin and creatine kinase-MB |  |  |  |  |  |  |  |  |  |  |  |  |  |  |  |  |
|  | Acute coronary syndrome |  |  |  |  |  |  |  |  |  |  |  | 6 ($) |  |  |  |  |
|  | Arrhythmia |  |  |  |  |  |  |  |  |  |  |  | 6 ($) | 2 (§) |  |  |  |
|  | Tachycardia |  |  |  |  |  | 2 (ψ) |  |  |  |  |  |  | 2 (§) |  |  |  |
|  | Hypertension |  |  |  |  |  |  |  |  |  |  |  |  |  |  |  |  |
|  | Cardiac tamponade | 4 |  |  |  |  |  |  |  |  |  |  |  |  |  |  |  |
|  | Annulus rupture |  |  |  |  |  |  |  |  |  |  |  |  |  |  |  |  |
|  | Coronary artery occlusion |  |  |  |  |  |  |  |  |  |  |  |  |  |  |  |  |
|  | Moderate aortic regurgitation |  |  |  |  |  |  |  |  |  |  |  |  |  |  |  |  |
|  | Severe hypotension requiring haemodynamic support |  |  |  |  |  |  |  |  |  |  |  |  |  |  |  |  |
|  | Shock with hypotension |  |  |  |  |  |  |  |  |  |  |  |  | 2 (§) |  |  |  |
|  | Cardiovascular arrest |  |  |  |  |  |  |  |  |  |  |  |  | 2 (§) |  |  |  |
|  | Permanent cerebral vascular accident | 4 |  |  |  |  |  |  |  |  |  |  |  |  |  |  |  |
|  | Oedema (30-day) |  |  |  | 2 (£) |  |  |  |  |  |  |  |  |  |  |  |  |
|  | Angioedema |  |  |  |  |  |  |  |  |  |  |  |  | 2 (§) |  |  |  |
|  | Vascular complications |  |  |  |  |  |  |  |  |  |  |  |  |  |  |  |  |
|  | Cardiovascular events |  |  |  |  |  |  |  |  |  |  |  |  |  |  |  |  |
|  | Major adverse cardiovascular events | 4 |  |  |  |  |  |  |  |  |  |  |  |  |  |  |  |
| Renal | Need for renal replacement therapy |  |  |  |  |  |  |  |  |  |  |  |  |  |  |  |  |
|  | Need for dialysis |  |  |  |  |  |  |  |  |  |  |  |  |  |  |  |  |
|  | Renal impairment |  |  |  |  |  |  |  |  |  |  |  |  |  |  |  |  |
|  | Renal failure rate (30-day, 90-day, 1-year; new onset, acute) | 4 |  |  | 2 (£) |  | 2 |  |  |  |  |  |  |  |  |  |  |
| Anaemia-associated ischemic events | Myocardial infarction (30-day, 90-day, 1-year, overall) |  |  |  | 2 (£) |  | 2 |  |  |  |  | 1 (*) |  |  |  |  |  |
|  | Myocardial ischemia |  |  |  |  |  |  |  |  |  |  |  |  |  |  |  |  |
|  | Stroke (30-day, 90-day, 1-year, overall) |  |  |  | 2 (£) |  | 2 |  |  |  |  |  | 6 ($) |  |  |  |  |
|  | TIA |  |  |  |  |  |  |  |  |  |  |  |  |  |  |  |  |
|  | Stroke or TIA |  |  |  |  |  |  |  |  |  |  |  |  |  |  |  |  |
|  | Bowel ischemia |  |  |  |  |  |  |  |  |  |  |  |  |  |  |  |  |
|  | Acute kidney injury |  |  |  |  |  |  |  |  |  |  |  | 6 ($) |  |  |  |  |
|  | Acute limb ischemia |  |  |  |  |  |  |  |  |  |  |  | 6 ($) |  |  |  |  |
|  | Ischemic events |  |  |  |  |  |  |  |  |  |  |  |  |  |  |  |  |
| Thrombo-embolic events | Deep venous thrombosis (requiring readmission) |  | 4 | 4 |  |  | 2 |  | 2 |  |  |  | 6 ($) |  |  |  | 2, 4 |
|  | Preoperative femoral vein thrombosis |  |  |  |  |  |  |  |  |  |  |  |  |  |  |  |  |
|  | Mesenteric venous thrombosis |  |  |  |  |  |  |  |  |  |  |  |  |  |  |  |  |
|  | Arterial thrombosis |  |  |  |  |  |  |  |  |  |  |  |  |  |  |  |  |
|  | Thrombosis |  |  |  |  |  |  |  |  |  |  |  |  |  |  |  | 2, 4 |
|  | Pulmonary embolism |  |  | 4 |  |  |  |  |  |  |  |  | 6 ($) |  |  |  |  |
|  | Embolism |  |  |  |  |  |  |  |  |  |  |  |  |  |  |  |  |
|  | Valve embolization |  |  |  |  |  |  |  |  |  |  |  |  |  |  |  |  |
|  | Thrombotic and/or vascular events |  |  |  |  |  |  |  |  |  |  |  |  |  |  |  |  |
| Mortality or survival | Mortality | 4 |  | 4 |  | 2 | 2 | 2 |  | 2 (ξ) | 4 |  |  |  |  |  |  |
|  | Survival |  |  |  |  |  |  |  |  |  |  |  |  |  |  | 2 |  |
| Other | Allergy |  |  |  |  |  |  |  |  |  |  |  | 6 ($) |  |  |  | 2, 4 |
|  | Severe allergic reactions |  |  |  |  |  | 2 (ψ) |  |  |  |  |  |  |  |  |  |  |
|  | Anaphylactic reactions |  |  |  |  |  |  |  |  |  |  |  |  |  |  |  |  |
|  | Lymph node swelling |  |  |  |  |  |  |  |  |  |  |  |  | 2 (§) |  |  |  |
|  | Acute myeloid leukemia |  |  |  |  |  |  |  |  |  |  |  |  |  |  |  |  |
|  | Grade 2 neutropenia |  |  |  |  |  |  |  |  |  |  |  |  |  |  |  |  |
|  | Cachexia |  |  |  |  |  |  |  |  |  |  |  |  |  |  |  |  |
|  | Multiorgan failure |  |  |  |  |  |  |  |  |  |  |  |  |  |  |  |  |
|  | Epileptic seizure |  |  |  |  |  |  |  |  |  |  |  | 6 ($) |  |  |  |  |
|  | Convulsions |  |  |  |  |  |  |  |  |  |  |  |  |  |  |  |  |
|  | Muscle spasms |  |  |  |  |  |  |  |  |  |  |  |  |  |  |  |  |
|  | Mild and transient hypertonia |  |  |  |  |  |  |  |  |  |  |  |  |  |  |  |  |
|  | Hypercalcemia |  |  |  |  |  |  |  |  |  |  |  |  |  |  |  |  |
|  | Hypokalemia |  |  |  |  |  |  |  |  |  |  |  |  |  |  |  |  |
|  | Insomnia |  |  |  |  |  |  |  |  |  |  |  |  |  |  |  |  |
|  | Anastomotic leak (requiring readmission) |  |  |  |  |  | 2 |  |  |  |  |  |  |  |  |  |  |
|  | Chyle leak |  |  |  |  |  | 2 |  |  |  |  |  |  |  |  |  |  |
|  | Intra-abdominal collection of fluids |  |  |  |  |  | 2 |  |  |  |  |  |  |  |  |  |  |
|  | Conversion to open heart surgery |  |  |  |  |  |  |  |  |  |  |  |  |  |  |  |  |
|  | Need for a second valve |  |  |  |  |  |  |  |  |  |  |  |  |  |  |  |  |
|  | Thrombosis or dysfunction of the prosthesis | 4 |  |  |  |  |  |  |  |  |  |  |  |  |  |  |  |
|  | Need for re-exploration |  |  |  |  |  |  |  |  |  |  |  |  |  |  |  |  |
|  | Reoperation | 4 |  |  |  |  |  |  |  |  |  |  |  |  |  |  |  |
|  | Hospital readmission |  |  |  |  |  |  | 2 |  | 2 (ξ) |  |  |  |  |  |  |  |
|  | Hypersensitivity reactions |  |  |  |  |  |  |  |  |  |  |  |  |  |  |  |  |
|  | Readmission for general postoperative complications between discharge and 8 weeks |  |  |  |  |  |  |  |  |  |  |  |  |  |  |  |  |
|  | Readmission for complications between discharge and 6 months |  |  |  |  |  |  |  |  |  |  |  |  |  |  |  |  |
|  | Grade of complication severity from recruitment to outpatients |  |  |  |  |  |  |  |  |  |  |  |  |  |  |  |  |
|  | Complication rate from recruitment to outpatients |  |  |  |  |  |  |  |  |  |  |  |  |  |  |  |  |
|  | Postoperative complication rate ≥ Clavien-Dindo grade III |  |  |  |  |  |  |  |  |  |  |  |  |  |  |  |  |
|  | Medical postoperative complications |  |  |  |  |  |  |  |  |  | 4 |  |  |  |  |  |  |
|  | Surgical postoperative complications |  |  |  |  |  |  |  |  |  | 4 |  |  |  |  |  |  |
|  | Non-thromboembolic complications (composite of prosthesis-related, cardiovascular, allergic, infectious, bleeding etc events) |  |  |  |  |  |  |  |  |  |  |  |  |  |  |  |  |
|  | Postoperative overall complication rate (composite of pulmonic, cardiologic, thrombotic, infectious and neurologic complications) |  |  |  |  |  |  |  |  |  |  |  |  |  | 2 |  |  |
|  | Postoperative complicate rate (composite of anastomotic leak, abscess/fistula formation, hemorrhage, wound infection, pulmonary complications, complications from blood transfusions) |  |  |  |  |  |  |  |  |  |  |  |  |  |  |  |  |

Cells shaded in green indicate that the corresponding cohort study provided data for the corresponding AE outcome(s). The number mentioned in the green-shaded cell represents the number of the treatment comparison. Comparison 1: Intravenous (IV) iron versus oral iron monotherapy, Comparison 2: IV iron versus usual care/no iron, Comparison 3: IV ferric carboxymaltose versus IV iron sucrose monotherapy, Comparison 4: ESA + iron versus control (placebo and/or iron, no treatment), Comparison 5: ESA + IV iron versus ESA + oral iron, Comparison 6: ESA + IV iron versus ESA + IV iron (different ESA dosing regimens).

If there are no green-shaded cells across an entire row, this means that data on this AE outcome were provided by RCTs (see Table D).

(£) indicates that raw data were provided by the study authors, whereas the published paper only contained small bar graphs presenting relative percentages of patients experiencing these events.

(§), (ψ), (ξ) and (#) indicate that additional unpublished data were obtained from the study authors. This enabled the inclusion of data, that otherwise would have been disregarded, on:

- (§) 13 outcomes across 8 AE categories;
- (ψ) 4 outcomes across 4 AE categories;
- (*) 4 outcomes across 4 AE categories.
- (ξ) 2 outcomes across 2 AE categories;
- (#) 1 outcome in 1 AE category.

($) indicates that further clarification was provided by the study authors on the specific types of adverse events studied. This enabled us to report data on 20 separate AEs instead of a single composite measure.

### Supplementary Table 6. Characteristics of included studies.

* In case of missing information, authors were either:

(1) not contacted by the reviewers because no valid email address of any of the authors was available;

(2) contacted by the reviewers via email. However, the authors did not respond to the email;

(3) successfully contacted by the reviewers via email. However, the authors responded that they did no longer have access to the study files and were not able to answer the reviewers’ questions.

| **Study** | | | **Population** | | | **Intervention(s)** | **Comparison** | **Co-interventions** | **RBC transfusion threshold** | **Outcome(s)** |
| --- | --- | --- | --- | --- | --- | --- | --- | --- | --- | --- |
| Author, year, country or registry number | Trial registry or peer-reviewed publication? | Design | Description | Definition anaemia | Definition iron-deficiency |  |  | *Administered to both the intervention and comparison group* |  |  |
| Biboulet, 2018, France | Peer-reviewed publication | Experimental: Randomized controlled trial | 100 adult patients (8% with true iron deficiency) scheduled for elective unilateral primary or revision total hip or knee arthroplasty with Hb levels 10-12.9 g/dl were randomized to one of 2 groups:  1) Epoetin- α + IV iron n=50, median age 67 years (range: 60-75), 42 women, initial median Hb level 12.5 g/dl (range: 11.8-12.7)  2) Epoetin- α + oral iron n=50, median age 71 years (range: 61-78), 38 women, initial median Hb level 12.3 g/dl (range: 11.8-12.8) | Authors: none  Reviewers: included because of initial Hb levels  (10-12.9 g/dl) | Authors:  True iron deficiency: serum ferritin level <30 µg/l in baseline blood sample | Epoetin-α +  IV iron  - 40 000 U epoetin-α (EPREX) subcutaneously on preoperative days -21, -14 and -7  - 1000 mg of ferric carboxymaltose (Ferinject) intravenously over 15 min, immediately after the anesthetic consultation and baseline blood sampling | Epoetin-α + oral iron  - 40 000 U epoetin-α (EPREX) subcutaneously on preoperative days -21, -14 and -7  - oral ferrous sulphate (Tardyferon), two tablets each morning (160 mg iron/day), starting the day after the anesthesiologist’s consultation | *Intraoperative*  Tranexamic acid  1000 mg before skin incision and during wound closure  Cell saver  used in case of revision surgery  Ferric hydroxide sucrose (Venofer)  300 mg administered over the course of 2 hours to all patients  *Postoperative*  Low-molecular-weight heparin  Epoetin-α:  40 000 U subcutaneously if Hb <15 g/dl on day 1  Ferric hydroxide sucrose:  300 mg intravenously on day 2 | Healthy patients in absence of physical activity:  Hb ≤ 7 g/dl  Healthy patients in presence of physical activity:  Hb ≤ 8 g/dl  Patients with cardiac or coronary insufficiency: Hb ≤ 10 g/dl | - Digestive complications: composite measure for nausea, diarrhea, constipation  - Preoperative femoral vein thrombosis  - Preoperative prostatitis  - Postoperative deep venous thrombosis (day 7)  - Cardiac or respiratory failure (within 1 month after surgery) - Mortality during hospitalization  Method of outcome assessment:  - Digestive complications: unclear*(2) (“complications related to ferrous sulfate, ferric carboxymaltose,  or epoetin-α therapy were recorded”) - Preoperative femoral vein thrombosis, preoperative prostatitis, cardiac or respiratory failure: unclear (“Major complications, including  thromboembolic events, cardiac or respiratory failure, and death during hospitalization, were reported on days 3 and 5  for inpatients. The occurrence of complications was further  evaluated by a systematic telephone call 1 month after surgery.”)  - Postoperative deep venous thrombosis:  PD (lower limb ultrasonography was performed in clinically suspected DVT)  Time period:  - Digestive complications: preoperatively  - Preoperative femoral vein thrombosis, preoperative prostatitis, cardiac or respiratory failure, deep venous thrombosis:  until postoperative month 1 |
| Cao, 2020, China | Peer-reviewed publication | Experimental: Randomized controlled trial | 102 adult patients scheduled for unilateral primary total knee arthroplasty with baseline Hb levels between 10 and 13 g/dL (0% iron-deficient) were randomized to one of 3 groups:  1) Preoperative EPO + IV iron n=35, mean age 67.77±8.45 years, 29 women 2) Postoperative EPO + IV iron n=35, mean age 67.43±8.34 years, 30 women 3) IV iron n=32, mean age 69.07±6.46 years, 28 women  [Data on the postoperative EPO + IV iron group were not extracted] | Authors: none  Reviewers: included because of inclusion criteria (Hb 10-13 g/dL) | Authors: none | Preoperative EPO + IV iron 8 daily doses of 10 000 IU rhEPO (150 IU/kg) subcutaneously  and 3 daily doses of 200 mg IV iron sucrose, starting 3 days before surgery | IV iron 3 daily doses of 200 mg IV iron sucrose, starting 3 days before surgery | *Intraoperative*  Tranexamic acid  1g IV prior to tourniquet deflation and  1g topical to bath the surgical site before wound closure  *Postoperative*  Heparin  4000 IU low molecular weight heparin daily, starting on postoperative day 1, for venous thrombosis prophylaxis  Foot pumps  used regularly for thromboembolic prophylaxis  Analgesia Patient-controlled pump and NSAIDs | - Hb <7.5 g/dL  - Cardiovascular and cerebrovascular disease or other symptoms of acute anemia: Hb <9 g/dl | - Deep vein thrombosis - Cardiovascular events - Nausea - Vomiting - Allergic reaction (systemic and/or skin)  Method of outcome assessment:  - Intramuscular vein thrombosis: PD; colour Doppler examination was used to screen for DVT before and between 2 and 4 weeks after surgery - Myocardial infarction, arrhythmia, allergic reaction: unclear,  but probably PD  - Nausea and vomiting:  unclear  Time period: - Intramuscular vein thrombosis: before and between 2 and 4 weeks after surgery - Other outcomes: unclear |
| Christodoulakis, 2005, Greece | Peer-reviewed publication | Experimental: Randomized controlled trial | 223 anaemic patients  (% of iron-deficient patients not reported) undergoing elective colorectal surgery for resectable colorectal cancer, with baseline Hb levels >9 and <12 g/dl, were randomly assigned to one of 3 groups:  1) Epoetin-α 150 IU + oral iron  n=69, 31 men and 38 women, median age 72 years (range 43-91)  2) Epoetin-α 300 IU + oral iron n=67, 30 men and 37 women, median age 71 years (range 36-92)  3) Control n=68, 28 men and 40 women, median age 70 years (range 44-89 | Authors: refer to the patients as “anemic”, but do not provide a definition  Reviewers:  included because of baseline Hb levels  (>9 and <12 g/dl) | Authors:  none | Epoetin-α 150 IU + oral iron  - 150 IU/kg/day Epoetin-α subcutaneously from preoperative day 10 until postoperative day 1  - 200 mg oral elementary iron supplements daily from preoperative day 10 until postoperative day 1  Epoetin-α 300 IU + oral iron - 300 IU/kg/day Epoetin-α subcutaneously from preoperative day 10 until postoperative day 1  - 200 mg oral elementary iron supplements daily from preoperative day 10 until postoperative day 1 | Oral iron  Oral elementary iron supplements 200 mg/day from preoperative day 10 until postoperative day 1 | Folic acid  15 mg/day for the first 10 days after randomization  In patients with iron deficiency only: postoperative IV iron  iron sulphate  40 mg intravenously daily until the day of discharge | *Preoperative*  - Hb <11 g/dl and severe heart disease, chronic obstructive lung disease or arterial disease  - Received β-blockers  - Lost a significant amount of blood  -Younger patients or patients in good health: Hb <9 g/dl  *Intraoperative*  - Blood loss > 300 ml and heart or lung or arterial disease  - Received β-blockers  - Elderly  - Younger patients or patients in good health: blood loss > 400 ml  *Postoperative*  - Hb <10 g/dl and poor prognostic features  - Younger patients or patients in good health: Hb <8 g/dl | Adverse events leading to patient withdrawal: - Local rash - Urticarial allergy to iron  Separate measures for different postoperative complications:  - cardiac arrest  - embolism  - cardio-respiratory failure  - grade 2 atrial fibrillation  - grade 2 neutropenia  - peritonitis  - unexplained grade 2 urticaria  - death  Method of outcome assessment:  unclear*(2)  Time period:  unclear*(2) |
| Cladellas, 2012, Spain | Peer-reviewed publication | Observational: Cohort study | 134 adult anemic patients  (% of iron-deficient patients not reported) undergoing elective heart valve replacement surgery, received one of 2 treatments:  1) Epoetin-β + IV iron  n=75, mean age 73±10 years, 47 women, initial Hb levels 11±1 g/dl  2) No Epoetin-β + no IV iron (historic control group) n=59, mean age 71±8 years, 59 women, initial Hb levels 10.9±0.9 g/dl | Authors:  WHO criteria (Hb<13 g/dl for men and <12g/dl for women) | Authors:  none (although mg of iron deficit determined the dose of iron administered to the patient) | Epoetin-β + IV iron  - 500 IU/kg/day Epoetin-β (Neo-Recormon) intravenously every week for 4 weeks and the 5^th^ dose 48 hours before surgery;  - Iron sucrose intravenously (maximum dose 200 mg/day in a drip infusion for 2 hours; dose depending on body weight and actual Hb levels), together with the Epoetin-β | No Epoetin-β + no IV iron  did not receive any treatment | None reported  Clopidogrel was withheld 10  days before cardiac surgery, and vitamin K antagonist was  replaced by enoxaparin 5 days before surgery | Hb <7  g/dl during or after surgery | In-hospital mortality  Composite and separate measures for major adverse cardiovascular events:  - heart failure - permanent cerebral vascular accident - acute renal failure  - perioperative myocardial infarction  - cardiac tamponade  - reoperation  - severe infection (sepsis, pneumonia, or mediastinitis)  - prolonged ventilation (≥24 hours)  - prosthesis thrombosis or dysfunction  - endocarditis  Method of outcome assessment: unclear*(2),  but probably PD (sometimes in combination with laboratory testing, e.g. measurement of serum creatinine levels to diagnose acute renal failure; see reference Cladellas 2006)  Time period:  unclear*(2)  (probably within 30 days of valve replacement; see reference Cladellas 2006) |
| de Andrade, 1996, USA | Peer-reviewed publication | Experimental: Randomized controlled trial | 316 adult patients (1% iron-deficient) scheduled for major elective orthopaedic surgery of hip or knee, with baseline Hb levels ≤15 g/dl, a serum iron to total iron-binding capacity ratio ≥15%, and a serum ferritin level ≥50 ng/ml, were stratified based on their entry Hb level into:  - Stratum 1 (Hb ≤10 g/dl, n=2)  - Stratum 2 (Hb >10 to ≤13 g/dl, n=96)  - Stratum 3 (Hb >13 g/dl, n=218)  Within each stratum, patients were randomly assigned to one of 3 groups:  1) Epoetin-α 300 IU + oral iron n=112, 74 women, mean age 65.84±12.7 years (range: 29-89 years)  2) Epoetin-α 100 IU + oral iron n=101, 59 women, mean age 65.98±13.44 years (range: 36-86 years)  3) Placebo + oral iron  n=103, 63 women, mean age 67.75±11.12 years (range: 32-92)  As this PICO specifically concerns patients with preoperative anaemia, only outcomes analysed in the stratum 2 patients (too few patients in stratum 1 for analysis) with entry Hb levels >10 to ≤13 g/dl were extracted.  Hence, 100% of the patients were anaemic with 0% being iron-deficient. | Authors:  Hb level <9 g/dl  Reviewers:  included data from the stratum 2 patients because of their entry Hb levels (>10 and ≤13 g/dl). | Authors: none (although the percentage of iron-deficient patients is presented in table I of the peer-reviewed publication) | Epoetin-α 300 IU + oral iron  - 300 IU/kg Epoetin-α subcutaneously for 10 consecutive days prior to the day of surgery, postoperatively on the day of surgery, and for 4 days after the day of surgery;  - ≥150 mg elementary iron orally starting on or before the first day of study medication until hospital discharge  Epoetin-α 100 IU + oral iron - 100 IU/kg Epoetin-α subcutaneously for 10 consecutive days prior to the day of surgery, postoperatively on the day of surgery, and for 4 days after the day of surgery;  - ≥150 mg elementary iron orally starting on or before the first day of study medication until hospital discharge | Placebo + oral iron  - Equivalent volume of placebo subcutaneously, identical to the Epoetin-α diluent for 10 consecutive days prior to the day of surgery, postoperatively on the day of surgery, and for 4 days after the day of surgery;  - ≥150 mg elementary iron orally starting on or before the first day of study medication until hospital discharge | Blood salvaging techniques or hemodilution during surgery and drainage or blood salvaging after surgery Similar across treatment groups and strata  Warfarin (-derivative)  standard anticoagulation therapy  (low-dose [5  mg/d] warfarin or a warfarin derivative)  starting on the day of surgery and continuing  until hospital discharge  (to keep prothrombin  time at 14 to 16 seconds for prophylaxis  against deep vein thrombosis) | Hb ≤9 g/dL, except  where clinical symptoms warranted transfusion | - Deep venous thrombosis rate  Method of outcome assessment:  PD, using ultrasonography (ipsilateral, contralateral, proximal and distal)  Time period:  postoperative day 5, 6, 7 or on the day of discharge (if earlier than day 5) |
| NCT00270088 | Synopsis of study obtained via trial registry |  |  |  |  |  |  |  |  |  |
| Delasotta, 2012, USA (A) | Peer-reviewed publication | Observational: Cohort study | 46 mildly anemic patients  (% of iron-deficient patients not reported) receiving elective revision hip surgery for prosthesis wear out and/or loosening, with pre-operative Hb levels ≥10 and ≤13 g/dl.  Of these, 16 (mean age 70.7 years) received Epoetin-α treatment.  Patients that did not receive Epoetin-α treatment (n=28, mean age 74.1 years)  were patient matched according to age, gender, body mass  index, and ASA score. | Authors: mild anemia = Hb level ≥10 and ≤13 g/dl | Authors:  none | Epoetin-α + oral iron  - 3 weekly doses of Epoetin-α  - Oral iron  No additional information provided | Oral iron  No additional information provided | *Preoperative*  - Oral multivitamins;  - Oral vitamin B12;  -Oral folic acid; - Prophylactic antibiotic  (1 g IV cefazolin or vancomycin) within one hour prior to incision until the first 24 hours after surgery.  *Intraoperative*  No cell-saver or drain.  *Postoperative*  Oral warfarin or subcuteanous enoxaparin | Hb levels ≤8 g/dl, or symptoms consistent with anemia | - Deep venous thrombosis rate  Method of outcome assessment: PD, using ultrasound  Time period: unclear*(2) |
| Delasotta, 2012, USA (B) | Peer-reviewed publication | Observational: Cohort study | 81 mildly anemic patients  (% of iron-deficient patients not reported) receiving elective revision knee surgery for prosthesis wear out and/or loosening, with pre-operative Hb levels ≥10 and ≤13 g/dl.  Of these, 28 (mean age 63.7 years) received Epoetin-α treatment.  Patients that did not receive Epoetin-α treatment (n=53, mean age 64.4 years)  were patient matched (1:2) according to one of 2 attending surgeons, gender, body mass  index, complexity of surgery, ASA score, and age. | Authors: mild anemia = Hb level ≥10 and ≤13 g/dl | Authors: none | Epoetin-α + oral iron  - Epoetin-α at preoperative days 21, 14 and 7  - Oral iron  No additional information provided | Oral iron  No additional information provided | *Preoperative*  - Oral multivitamins;  - Oral vitamin B12;  -Oral folic acid;  - Prophylactic antibiotic  (1 g IV cefazolin or vancomycin) within one hour prior to incision until the first 24 hours after surgery.  *Intraoperative*  No cell-saver or drain.  *Postoperative*  Oral warfarin or subcuteanous enoxaparin for 4 weeks | Based on peri- and postoperative  Hb levels (not specified), the ASA score, and/or clinical symptoms consistent with anemia | - Cellulitis after surgery  - Pulmonary embolism  - Hemarthrosis after surgery  - Deep venous thrombosis - 7-month postoperative mortality  Method of outcome assessment:  unclear*(2)  Time period:  unclear*(2) |
| Dickson, 2020, UK | Peer-reviewed publication | Experimental: Randomized controlled trial | Long-term results of the IVICA trial  (see Keeler, 2017)  110 adult anaemic patients  (% of iron-deficient patients not reported) that underwent elective colorectal cancer surgery for non-metastatic colorectal adenocarcinoma, with Hb levels <11 g/dl (women) or <12 g/dl (men), who were randomized to one of 2 groups during the IVICA trial:  1) IV iron  n=54, 35 men and 19 women,  mean age 74.1±8.8 years  2) Oral iron  n=56, 34 men and 22 women, mean age 75.2±11.1 years | Authors:  WHO criteria (Hb<13 g/dl for men and <12g/dl for women) | Authors: none | IV iron  Ferric carboxymaltose (Ferinject) diluted in 250 ml 0.9% normal saline, infused over 15 minutes.  First dose was administered at initial recruitment visit (≥14 days before surgery).  Dosing regimen:  - Hb ≥10 g/dl:  1000 mg (<70 kg body weight) or 1500 mg (≥70 kg body weight)  - Hb <10 g/dl: 1500 mg (<70 kg body weight) or 2000 mg (≥70 kg body weight). | Oral iron  200 mg ferrous sulphate, twice daily for 2 weeks, starting from initial recruitment visit. | Postoperative oral iron in 30/56 patients in the oral iron group and 4/54 patients in the IV iron group | Transfusion was considered if  Hb levels were < 8.0 g/dl,  was indicated  if Hb was < 7.0 g/dl,  but the decision  to transfuse was made based on the clinical condition of  the patient | - Wound dehiscence  - Anastomotic leak - Atrial fibrillation - Deep vein thrombosis - 2-, 3-, 4- and 5-year overall survival - 2-, 3-, 4- and 5-year disease-free survival - 2-, 3-, 4- and 5-year colorectal cancer-specific survival  Method of outcome assessment: PD "The seven previous participating centres were  contacted to submit anonymized data for their patients  on a standardized case report form. Data were collected from electronic hospital records and patients were not contacted to provide additional information."  Time period:  up to 5-years |
| Dousias, 2003, Greece | Peer-reviewed publication | Experimental: Randomized controlled trial | 50 mildly anemic  (0% iron-deficient, since initial ferritin levels >50 ng/ml) women with benign uterine leiomyomas scheduled for abdominal total hysterectomy, aged between 30 and 60 years old, with initial Hb levels ≥9 and <12 g/dl, were randomly allocated to one of 2 groups:  1) rHuEPO + oral iron  n=23, average age 48±4 years 2) Saline + oral iron n=27, average age 49±5 years | Authors:  refer to the patients as “mildly anemic”, but do not provide a definition  Reviewers:  included because entry Hb levels  (≥9 and <12 g/dl) | Authors: none | rHuEPO + oral iron  - 600 U/ml rHuEPO subcutaneously on preoperative days 14 and 7 and the morning before the operation  - 200 mg oral iron per day throughout the study period (reviewers assume from preoperative day 14 to postoperative day 14) | Saline + oral iron  - Normal saline subcutaneously on preoperative days 14 and 7 and the morning before the operation  - 200 mg oral iron per day throughout the study period (reviewers assume from preoperative day 14 to postoperative day 14) | None reported  None of the women had  received GnRH analogues preoperatively | Not reported | - Febrile episodes - Hypertension  Method of outcome assessment: unclear*(2)  Time period: unclear*(2) |
| Dousias, 2005, Greece | Peer-reviewed publication | Experimental: Randomized controlled trial | 38 women (0% iron-deficient, since mean initial ferritin levels are 56.6 and 61.5 ng/ml) with gynecological cancer scheduled for radical abdominal surgery, were randomly allocated to one of 2 groups:  1) rHuEPO + oral iron n=20, mean age 48.6±7.6 years, mean Hb level on preoperative day 10 10.6±0.8 g/dl  2) Placebo + oral iron n=18, mean age 46.9±7.1 years, mean Hb level on preoperative day 10 10.7±0.7 g/dl | Authors:  none (although one of the paper’s keywords is ‘anemia’)  Reviewers:  included because of mean initial Hb levels (10.6 and 10.7 g/dl) | Authors: none | rHuEPO + oral iron  - 200 U/kg body weight rHuEPO subcutaneously daily from preoperative day 10 until postoperative day 5  - 200 mg oral iron daily from preoperative day 10 until postoperative day 5 | Placebo + oral iron  - placebo (similarly looking subcutaneous injections of only water) daily from preoperative day 10 until postoperative day 5  - 200 mg oral iron daily from preoperative day 10 until postoperative day 5 | Heparin  all women were administered low molecular  weight heparin thromboprophylaxis | Not reported | - Postoperative fever  Method of outcome assessment:  unclear*(2)  Time period:  unclear*(2) |
| Ellermann, 2018, Germany | Peer-reviewed publication | Observational: Cohort study | 666 men (of which 183 (27%) anaemic) and 435 women (of which 136 (31%) anaemic) scheduled for elective surgery (cardiac, thoracic, orthopaedic, gynaecological and obstetric) with a risk of transfusion >10%.  The anaemic patients (n=312, 26% iron-deficiency anaemia and 55.8% anaemia of chronic disease) either received:  1) IV iron  Men: n=76, median age 72 years (Q25%-Q75%: 62-78), median baseline Hb level 10.9 g/dl (10.1-11.8)  Women: n=73, median age 63 years (45-75), median baseline Hb level 10.7 g/dl (9.6-11.3)  2) No IV iron  Men: n=107, median age 73 years (61-77), median baseline Hb level 11.8 g/dl (10.7-12.5)  Women: n=63, median age 66 years (51-77), median baseline Hb level 10.8 g/dl (10.0-11.4)  In the no IV iron patients, intravenous iron was ruled out due to relative and absolute contraindications. | Authors:  WHO criteria (Hb<13 g/dl for men and <12g/dl for women) | Authors:  Iron-deficiency anemia:  Hb >9 g/dl and <13 (men) or <12 g/dl (women), microcytic and hypochromic erythrocytes, transferrin saturation <20% and ferritin levels ≤200 ng/ml | Intravenous iron  500 mg ferric carboxymaltose (Ferinject) diluted in 100 ml 0.9% saline intravenously between preoperative day 28 and 1 | No intravenous iron  no additional information provided | Patients received the same medications and interventions except for the IV iron. Patients did not receive extra vitamins or folic acid prior to surgery to increase Hb. During and after the operation, the concept of PBM was applied to all patients (personal communication with Steinberger). | According to the German guidelines; generally, the trigger was Hb<8 g/dl, or presence of signs of anemic hypoxia (personal communication with Steinberger) | - Prevalence of infectious-related codes  - Adverse events occurring within 30 days after the visit at the anesthesia/PBM clinic (separate measures for pruritus, urticaria, erythema, edema, palor, nausea, vomitus, abdominal pain, obstipation, diarrhoea, flatulence, dyspepsia, dyspnea, cephalgia, vertigo, paresthesia, dysgeusia)  - Health-related events occurring within 30 days after the visit at the anesthesia/PBM clinic (separate measures for myocardial infarction, heart failure, stroke, renal failure, pneumonia, infection, pyrexia, sepsis)  - Health-related events occurring within 90 days after the visit (same separate measures)  - Health-related events occurring within 1 year after the visit (same separate measures)  Method of outcome assessment:  - Infectious-related codes:  LD (codes are entered by a physician or by code experts, but are based on clinical examination, laboratory parameters (leucocytes, CRP and procalcitonin) and the results of specimen testing (sputum, urine, blood culture samples))  - Adverse events at 30 days, health-related events at 30 days, 90 days and at 1 year: PR (standardized telephone interview with the patient or a close relative, following a closed frame with the option to answer yes or no. Corresponding author Steinbicker added by email that all cases were checked for additional records, but not many patients were re-hospitalized or had letters available. In these cases, the authors relied on the patients’ answers)  Time period: - Infectious-related codes: until hospital discharge - Adverse events: 30 days - Health-related events: 30 days, 90 days and 1 year |
| Evans, 2021, UK | Peer-reviewed publication | Observational: Cohort study | 447 patients scheduled for elective cardiac surgery were pre-assessed for iron-deficiency anaemia. Of these, 300 were not anaemic.  Of the 147 anaemic patients, 75 (mean age 71±11 years, 48 men) were treated with intravenous iron, whereas 72 (mean age 72±8 years, 32 men) remained untreated. | Authors: Hb <13 g/dL and ferritin <100 ng/mL | | IV iron single dose of 20 mg/kg body weight iron isomaltoside (Monofer®) at a median (IQR [range]) time of 42 (28-72 [7-262]) days before surgery | No treatment  Reasons:  - no iron deficiency (n=22);  - patient  location too remote (n=1); - insufficient time between pre-assessment and surgery (n=5);  - patient declined the offer of iron (n=1);  - the responsible  surgeon preferred not to offer the patient iron (n=43). | *Preoperatively* Tranexamic  acid  2 g  *Intraoperatively* Tranexamic  acid  2 g Cell salvage  *Postoperatively* Tranexamic  acid  2 g | Hb< 8 g/dL with a single unit transfusion policy | - Mortality within 30 days or during index hospital admission  Method of outcome assessment: N/A  Time period: within 30 days or during index hospital admission |
| Froessler, 2016, Australia | Peer-reviewed publication | Experimental: Randomized controlled trial | 72 adult patients with preoperative iron deficiency anemia scheduled for abdominal surgery (mixture of malignant and non-malignant) were randomly assigned to one of 2 groups:  1) IV iron  n=40, 19 men and 21 women, mean age 64±15 years  2) Usual care  n=32, 17 men and 15 women, mean age 68±15 years | Authors:  Iron-deficiency anemia = ferritin <300 µg/L, transferrin saturation <25%,  Hb<12.0 g/dL (women) or <13.0 g/dL (men) | | IV iron  - Preoperatively: Single dose of ferric carboxymaltose intravenously, given over 15 minutes before surgery (simplified dosing protocol; 15 mg/kg body weight to a maximum  dose of 1000 mg)  - Postoperatively (within 2 days of surgery):  if blood loss >100 ml: 0.5 mg ferric carboxymaltose per recorded ml of blood loss | Usual care  Could consist of:  - No treatment - Continued observations  - Oral iron  - IV iron  - Allogeneic blood transfusion | None reported | Not reported | - Mild adverse events: headache, light-headedness, back pain  - Infection  - Respiratory failure  - Renal impairment  - Deep venous thrombosis  - Hospital readmission  - Mortality  Method of outcome assessment: - Mild adverse events: PR + PD; iron infusion adverse event sheet was completed after the infusion, based on self-reporting and observation (see personal communication with Froessler)  - Other outcomes: PD; Complications, recorded as coded conditions were transferred from the discharge summary and missing information completed at the final interview during the surgical follow up (see personal communication with Froessler)  Time period: During and immediately after iron infusion and at the final interview during appointment 4 weeks post-surgery (see personal communication Froessler) |
| Heiss, 1996, Germany | Peer-reviewed publication | Experimental: Randomized controlled trial | 30 adult moderately anemic patients with primary diagnosis of resectable colorectal cancer were randomly assigned to one of 2 groups:  1) rHuEPO + oral iron  n=17 (3 of the 20 randomized patients dropped out),  7 men and 10 women, median age 66 years (range 42-80), mean baseline Hb levels 12.2±0.39 g/dl  2) Placebo + oral iron  n=10 (0 dropouts), 2 men and 8 women, median age 61 years (range 42-74),  mean baseline Hb levels 12.6±0.74 g/dl  Of the 27 patients included, 66.6% was iron-deficient | Authors:  Moderate anemia = Hb 9-13 g/dl | Authors:  transferrin  saturation ≤ 15% | rHuEPO + oral iron  - 150 IU/kg body weight rHuEPO subcutaneously every 2 days,  from preoperative day 10 until postoperative day 2  - 200 mg ferrous sulphate orally each preoperative day | Placebo + oral iron  - placebo subcutaneously every 2 days, from preoperative day 10 until postoperative day 2  - 200 mg ferrous sulphate orally each preoperative day | Folate  5 mg orally each preoperative day | Indicated by the patient's attending  anesthesiologist or surgeon and recommended at a  Hb≤ 9 g/dl, depending on the  recorded blood loss | - Postoperative septic shock  - Postoperative multiorgan failure  - Postoperative mesenteric venous thrombosis with subtotal small bowel infarction  - Mild and transient hypertonia  - Postoperative deep venous thrombosis  Method of outcome assessment:  unclear*(2) (“medical history, physical examination,  evaluation of vital signs, serum chemistry test, and blood  cell counts were performed every second day until dismissal  from hospital”; unclear if adverse events were diagnosed by a physician or through laboratory testing)  Time period: until hospital discharge |
| Kam, 2020, China (Hong Kong) | Peer-reviewed publication | Observational: Cohort study | 100 anaemic adult patients  undergoing elective colorectal cancer surgery, received one of 2 treatments:  1) IV iron  n=38, median age 70.5 years (range: 45-85), 19 women, (100% iron-deficient)  2) No IV iron (historic control group) n=62, median age 69 years (range: 43-88), 31 women  (% iron-deficient unclear; lab data did not include ferritin and total iron-binding capacity) | Authors:  <10 g/dL before transfusion, or <12 g/dL after recent transfusion | Authors:  serum iron <7 µmol/L, ferritin <34 pmol/L,  and total iron-binding capacity >77 µmol/L (see personal communication Kam) | IV iron  Either:  - 500 mg iron sucrose (Venofer®) in 250 mL normal saline over 210 min intravenously, with a total of 2 doses set at 1 week apart  OR  -1000 mg (or 20 mg/kg if bodyweight <50 kg) iron isomaltoside (Monofer®) in 100 mL normal saline over 15 min as a single dose  IV iron was administered at least 2 weeks prior to elective surgery | No IV iron | None reported  Ongoing medications of patients: - Antiplatelet therapy (IV iron: 13.2% vs control: 16.1%); - Anticoagulant therapy (IV iron: 2.6% vs control: 8.1% );  - Oral iron therapy (IV iron: 31.6% vs control: 29%) | IV iron group Hb ≤8 g/dL or  at surgeon’s or anesthesist’s discretion  Historical control group non-standardised; at the surgeon's or anesthetist's discretion (see personal communication with Kam) | Adverse events due to IV iron infusion (obtained from authors, see personal communication with Kam): - Fever - Tachycardia - Rash - Severe allergic reactions  Complications: -Intra-abdominal collection (i.e. abscess or rim-enhancing collection, infected hematoma, fluid around anastomosis with suspected leakage; simple free fluids or ascites were not considered)  -Anastomotic leak  -Wound infection  -Pneumonia/respiratory failure  -Renal failure  -Prolonged ileus  -Bleeding  -Chyle leak - Myocardial infarction - Stroke - Deep venous thrombosis  -Reoperation rate  -Mortality  Method of outcome assessment:  - Prospective intervention group: all outcomes: PD (Adverse events IV infusion: patients were monitored during and right after IV iron infusion; see personal communication with Kam; Complications: “Patients were  monitored postoperatively for recovery progress and any  adverse outcomes”)  - Retrospective (historic) control group: PD, data obtained from Clinical Data Analysis and Reporting System, patient's electronic records and written clinical notes (see personal communication with Kam)  Timing of outcome assessment (see personal communication with Kam):  - Prospective intervention group: during hospitalisation - Retrospective (historic) control group: during hospitalisation |
| Keeler, 2017, UK | Peer-reviewed publication | Experimental: Randomized controlled trial | 116 adult anaemic patients  (% of iron-deficient patients not reported) scheduled for colorectal cancer surgery for non-metastatic colorectal adenocarcinoma, with Hb levels <11 g/dl (women) or <12 g/dl (men), were randomized to one of 2 groups:  1) IV iron  n=55, 35 men and 20 women,  median age 73.8 years (IQR: 67.4-78.6)  2) Oral iron  n=61, 37 men and 24 women, median age 74.7 years (IQR: 67.9-80.8)  To minimize the risk of including patients with non-iron-deficient anaemia, patients  with metastatic disease, pre-existing haematological disease,  renal failure and those currently undergoing chemotherapy  were excluded. | Authors:  WHO criteria (Hb<13 g/dl for men and <12g/dl for women) | Authors: none | IV iron  Ferric carboxymaltose (Ferinject) diluted in 250 ml 0.9% normal saline, infused over 15 minutes.  First dose was administered at initial recruitment visit (≥14 days before surgery).  Dosing regimen:  - Hb ≥10 g/dl:  1000 mg (<70 kg body weight) or 1500 mg (≥70 kg body weight)  - Hb <10 g/dl: 1500 mg (<70 kg body weight) or 2000 mg (≥70 kg body weight).  A maximum dose of 1000 mg was administered per week and a maximum of 2000 mg during the trial. If patients  required 2 doses, the second dose was administered at least 7 days after the first. | Oral iron  200 mg ferrous sulphate, twice daily for 2 weeks, starting from initial recruitment visit. | Postoperative oral iron  in 30/56 patients in the oral iron group and 4/54 patients in the IV iron group | Transfusion was considered if  Hb levels were < 8.0 g/dl,  was indicated  if Hb was < 7.0 g/dl,  but the decision  to transfuse was made based on the clinical condition of  the patient | - Dyspepsia  - Constipation  - Post-infusion headache  - Rash  - Grade of infective complication severity  - Mortality during study  - 90-day mortality rate  Method of outcome assessment: PR or PD  (“All adverse events occurring during the study observed by the investigator or reported by the  participant, whether or not attributed to study medication, were recorded on the case report form.”; see end of study report)  Time period: until postoperative week 12 (see end of study report) |
| EudraCT 2011-002185-21 | End of study report obtained via trial registry |  |  |  |  |  |  |  |  |  |
| Keeler, 2019, UK | Peer-reviewed publication | Paper on the IVICA trial (see Keeler, 2017) | | | | | | | | More detailed information reported on the infection rates recorded as part of the IVICA trial. Composite measure for: - Wound infection - Lower respiratory infection - Urinary tract infection - Sepsis of unknown source  Method of outcome assessment: No additional information compared to Keeler 2017  Time period: No additional information compared to Keeler 2017 |
| Kettelhack, 1998, Germany | Peer-reviewed publication | Experimental: Randomized controlled trial | 109 anaemic patients with colon cancer scheduled for right hemicolectomy, with Hb levels > 8.5 and ≤13.5 g/dl, were randomly assigned to one of 2 groups:  1) Epoetin-β  n=48, 21 men and 27 women, median age 71 years (range 53-57),  median Hb levels 11.5 g/dl (range: 10.5-12.5), 87% iron-deficient  2) Placebo n=54, 22 men and 32 women, median age 67 years (range 37-91),  median Hb levels 12.0 g/dl (range: 11-12.1), 80% iron-deficient | Authors:  Moderate anaemia = Hb >8.5 and  ≤ 13.5 g/dl  (although the authors use a cut-off level of Hb 11.5 g/dl during multiple logistic regression analysis) | Authors: transferrin saturation <20% | Epoetin-β  - 20 000 IU Epoetin-β subcutaneously for a minimum of 5 (maximum 10) preoperative days until postoperative day 4  - Oral iron in case of iron deficiency (87% of patients) throughout the study | Placebo  - Placebo subcutaneously for a minimum of 5 (maximum 10) preoperative days until postoperative day 4  - Oral iron in case of iron deficiency (80% of patients) throughout the study | IV iron  Postoperative day 1: 40 mg iron sulphate intravenously (regardless of iron status) | Hb ≤7.5 g/dl | - Chills with fever during treatment  - Constipation  - Arterial thrombosis in post-treatment period  - Mycotic sepsis in post-treatment period  - Heart failure due to underlying malignant disease  - Heart failure due to septicaemia  - Cachexia  - Mortality  Method of outcome assessment:  unclear. No information available in the paper. Kettelhack confirmed by email that case report forms were systematically filled out, that outcomes were probably patient reported, that complications noted were documented in the patient records, but that the grade of validation is not certain.  Time period:  until postoperative month 3 |
| Khalafallah, 2012, Australia | Peer-reviewed publication (predatory journal) | Experimental: Randomized controlled trial | 44 patients with iron-deficiency anaemia undergoing elective joint arthroplasty, with Hb levels >9 g/dl and <12 g/dl (women) or <14 g/dl (men), were randomly assigned to one of 2 groups:  1) IV iron  2) Oral iron  Demographics of the 33 patients that completed the trial: 14 men and 19 women, median age 68 years (range 45-91), median initial Hb 11 g/dl (range 9-13) | Authors:  Hb levels <12 g/dl (women) or <14 g/dl (men) | Authors:  ferritin level <30 µg/L | IV iron  single dose of iron polymaltose (Ferrosig) dissolved in normal saline infused over 120 min, 4 weeks before surgery  The total dose was calculated according  to the patient’s body weight at preadmission visit and entry Hb level  according to the product guidelines | Oral iron  325 mg iron sulphate (105 mg elemental iron) daily for 4 weeks preoperatively | No fibrinolytic agents  Epidural anaesthesia used during surgery  (may have contributed to reduced blood loss) | Hb level <8 g/dl in the immediate postoperative period | - Infective complications during and after surgery  Method of outcome assessment: unclear*(2)  Time period: unclear*(2) (probably until hospital discharge) |
| Kim, 2009, South Korea | Peer-reviewed publication | Experimental: Randomized controlled trial | 76 menorrhagic women with established iron-deficiency anemia, with Hb levels <9 g/dl, scheduled to undergo surgical treatment were randomized into one of 2 groups:  1) IV iron  n=39, of which 9 dropped out.  Demographics of 30 remaining patients:  mean age 42.0±7.4 years, mean preoperative Hb levels 7.5±1.2 g/dl  2) Oral iron  n=37, of which 11 dropped out.  Demographics of 26 remaining patients:  mean age 42.3±8.0 years, mean preoperative Hb levels 7.8±1.1 g/dl | Authors: none | | IV iron  2 ampoule infusions of iron sucrose (Venoferrum) 3 times per week, beginning 3 weeks before surgery  Total dose was based on Hb levels and body weight.  In each infusion, the maximum total dose administered was 200 mg of elemental iron in 100 ml of 0.9% normal saline, infused over 20–30 min.  Most patients received iron sucrose at the rate of 200 mg every other day, 3 times per week, beginning 3 weeks before surgery. Treatment was completed after  administration of the calculated dose. | Oral iron  2 ampoules (80 mg in total) iron protein succinylate (Hemo-Q Soln) daily, beginning 3 weeks before surgery until the time of surgery | None reported | Not reported | - Myalgia  - Injection pain  - Nausea  - Dyspepsia  - Mortality  Method of outcome assessment: Myalgia, injection pain, nausea, dyspepsia:  PR  (IV iron patients were actively asked to note any symptoms or adverse effects of treatment before, during and after each infusion)  Time period: - Myalgia, injection pain, nausea, dyspepsia:  during treatment with IV iron (before, during and after each infusion) or oral iron  - Mortality: during study period |
| Klein, 2020, UK | Peer-reviewed publication | Observational: Cohort study | 228 adult patients  undergoing elective cardiac and vascular surgery in 11 UK cardiac centres, receiving:  1) IV iron  n=64, mean age 70.2±10.9 years, 26 women, 100% anaemic (% iron-deficient not reported)  2) No IV iron  n=72, mean age 69.3±11.8 years, 21 women,  100% anaemic  (% iron-deficient not reported)  3) No IV iron  n=92, mean age 67.0±9.7 years, 16 women,  100% non-anaemic (% iron-deficient not reported)  As this PICO specifically concerns patients with preoperative anaemia, outcomes analysed in the third non-anaemic group were excluded | Authors: WHO criteria (Hb<13 g/dL for men and <12g/dL for women) | Authors:  ferritin <100 and transferrin saturation <20% | IV iron  Single dosis of either: - Iron isomaltoside 1000 (Monofer®) at a total dose of calculated at  20 mg/kg  OR  - Ferric carboxymaltose (Ferinject®), to a maximum of 1000 mg  IV iron was administered by infusion over at least 15-30 minutes according to local policy, at least 10 days before surgery ( median of 33 days before surgery (IQR: 15-53, range: 3-303)). | No IV iron lack of treatment with IV iron was mainly due to logistical or geographic barriers | Information obtained from the authors (personal communication with Klein): - All patients received tranexamic acid as standard in all institutions  - Some patients received cell salvage according to institutional protocols - Patients will have received different thromboprophylaxis regimens at different institutions | Information obtained from the authors (personal communication with Klein):  Depending on the local protocols. Nearly always Hb<7 g/dL on bypass and Hb<8 g/dl after surgery | - Mortality - Number of readmissions  - Nausea  [Information obtained from the authors (personal communication with Klein: data on renal function were not collected]  Method of outcome assessment:  - Adverse events related to IV iron therapy: PR or PD (personal communication with Klein: "We monitored any reported side effects e.g. if the patient said they felt nauseous and asked the frequently for they were feeling or of any complaints etc.")  Timing of outcome assessment:  - Mortality: 30 days after the operation  - Adverse events related to IV iron therapy: during and for up to 30 minutes after IV iron infusion |
| Kosmadakis, 2003, Greece | Peer-reviewed publication | Experimental: Randomized controlled trial | 75 moderately anaemic patients  (% of iron-deficient patients not reported) with non-metastatic gastrointestinal tract cancer, with Hb levels between 8.5 and 13 g/dl, were randomly assigned to one of 2 groups: 1) Epoetin-α+ IV iron  2) Control + IV iron  12 randomized patients were excluded because they did not fulfil the inclusion criteria. Therefore, only 63 patients were evaluated:  1) Epoetin-α + iv iron  n=31, 15 men and 16 women, aged on average 67.1±2.1 years, mean Hb 10.6±0.18 g/dl  2) Placebo + iv iron n=32, 19 men and 13 women, aged on average 66.4±2 years, mean Hb 11.1±0.19 g/dl | Authors:  Moderate anemia = Hb 8.5-13 g/dl | Authors: none | Epoetin-α + IV iron  - 300 IU/kg body weight Epoetin-α subcutaneously daily starting from preoperative day 7 until postoperative day 7 - 100 mg intravenous iron (Venofer) daily starting from preoperative day 7 until postoperative day 7 | Placebo + IV iron  - Placebo subcutaneously daily starting from preoperative day 7 until postoperative day 7 - 100 mg intravenous iron (Venofer) daily starting from preoperative day 7 until postoperative day 7 | None reported | Hb ≤8.5 g/dl | - Postoperative complication rate (composite measure of anastomotic leak, abscess/fistula formation, haemorrhage, wound infection, pulmonary complications, complications from blood transfusions)  - Deep venous thrombosis  - Mild and transient hypertension  - Major allergic reactions - 1-year survival  Method of outcome assessment:  - Postoperative complication rate: unclear*(1), but probably PD  - DVT, hypertension and major allergic reactions:  unclear*(1), but probably PD (patients  were admitted for 2 hours in an outpatient manner while  remaining under continuous surveillance for possible adverse  effects)  Time period: probably until hospital discharge, except for 1-year survival |
| Larson, 2001, Sweden | Peer-reviewed publication | Experimental: Randomized controlled trial | 32 anaemic (all iron-deficient) women with uterine myoma scheduled for hysterectomy, with Hb levels <12 g/dl, were randomly assigned to one of 2 groups:  1) Epoetin-β + oral iron  n=15, mean age 46±1 years  2) Oral iron n=16, mean age 44±1 years | Authors:  Hb <12 g/dl | Authors:  mean serum ferritin below the lower reference  value and transferrin saturation <15 % | Epoetin-β + oral iron  - 5000 IU Epoetin-β (NeoRecormon) subcutaneously twice per week during 4 preoperative weeks  - Oral iron succinate 100 mg twice per day during 4 preoperative weeks | Oral iron  Oral iron succinate 100 mg twice per day during 4 preoperative weeks | None reported | Not reported | - Postoperative infection (composite and separate measures for: superficial wound infection, severe streptococcal septicaemia, urinary tract infection)  Method of outcome assessment:  unclear*(2) (“Postoperative infections were recorded”, “The safety and tolerability of EPO was evaluated by reported adverse events, laboratory tests and blood pressure measurements”)  Time period:  until postoperative day 14 |
| Laso-Morales, 2017, Spain | Peer-reviewed publication | Observational: Cohort study | 322 mild-to-moderately anemic (100% iron-deficient) patients undergoing elective colorectal cancer resection surgery received one of 2 treatments:  1) IV iron  n=232, 135 men and 97 women, mean age 71±11 years, mean baseline Hb levels 10.8±1.5 g/dl  2) Standard care  n=90, 45 men and 45 women, mean age 69±15 years, mean baseline Hb levels 12.0±10.9 g/dl  Patients in the standard care group were either:  - not referred to the anemia clinic  - ruled out for IV iron due to contraindications  - refusing therapy  - having logistic problems to attend the clinic and therefore received 100 mg elementary iron per day | Authors:  - Mild anemia  = Hb 11-12.9 g/dl  - Moderate anemia  = Hb 8-10.9 g/dl | Authors: serum ferritin level < 30 ng/mL. In anemic patients with inflammation  (CRP>5 mg/L), transferrin saturation < 20%  also identifies iron deficiency even with normal-to-elevated ferritin levels  (30-300 ng/mL). | Intravenous iron  200 mg iron sucrose intravenously diluted in 100 ml normal saline over 30-60 minutes, up to 3 times per week preoperatively  OR  500-1000 mg ferric carboxymaltose intravenously diluted in 200 ml normal saline over 15-30 minutes, once a week preoperatively | Standard care  Oral iron or no iron | Standardized anesthetic and surgical  protocols, antibiotic and antithrombotic prophylaxes, and  postoperative analgesia were used. | In general:  Hb <8 g/dl  In the presence of cardiac disease or symptoms of acute anemia:  Hb <9 g/dl | - Infectious complication rate (composite and separate measures for: surgical wound, intra-abdominal abscess, suture dehiscence, urinary tract, pneumonia)  - Hemorrhagic complication rate (composite and separate measures for:  surgical wound hematoma, rectorrhagia/hemoperitoneum)  - Paralytic ileus  - Thromboembolic complications (composite measure)  Method of outcome assessment:  - Infectious complications: PD + laboratory, microbiologic and/or radiologic confirmation - Surgical wound hematoma: PD: assessed clinically (see personal communication with Muñoz) - Rectorrhagia/ hemoperitoneum: PD; assessed clinically and via CT scan or abdominal ultrasonography (see personal communication with Muñoz)  - Paralytic ileus: PD; assessed clinically and via abdominal X-ray (see personal communication with Muñoz)  - Deep venous thrombosis:  PD; assessed clinically and via duplex ultrasonography (see personal communication with Muñoz)  Time period:  until postoperative day 30 |
| Lee, 2019, South Korea | Peer-reviewed publication | Experimental: Randomized controlled trial | 101 women with benign uterine diseases, assumed to cause menorrhagia, and iron-deficiency anaemia at the time of preoperative laboratory workup were randomly assigned to one of 2 groups:  1) IV ferric carboxymaltose  n=52, aged on average 44±5.7 years; all anaemic and iron-deficient  2) IV iron sucrose  n=49, aged on average 43.4±5.0 years; all anaemic and iron-deficient | Authors:  Hb levels <10 g/dl | Authors: serum ferritin level <30 ng/ml | IV ferric carboxymaltose  single dose of Ferinject infused over 15 min, based on body weight (<50kg: 500 mg iron; ≥50 kg: 1000mg) | IV iron sucrose  Venoferrum, dosage period based on calculated iron deficit using the Ganzoni formula, up to 3 dosing visits per week, maximum of 600 mg iron per week in 200 mg iron single administration sessions | None reported | Not reported | - Mortality - Anaphylactic reactions  Method of outcome assessment:  unclear*(2)  Time period:  study duration (2 weeks after first treatment administration) |
| Nandhra, 2020, UK | Peer-reviewed publication | Observational: Cohort study | 57 adult anaemic patients undergoing open or endovascular surgery at 10 UK centres that were either treated with IV iron (n=10, 7 men and 3 women, mean age 71.8±7.1 years, 50% iron-deficient) or not treated (n=47, 40 men and 7 women, mean age 74.4±9.0 years, 21% iron-deficient) | Authors:  WHO criteria (Hb<13 g/dl for men and <12g/dl for women) | Authors: ferritin  level <100 μg/L and/or transferrin saturation <20 % | IV iron  at least 10 days before surgery No further information, but probably the same formulation and dose as Klein 2020. | No IV iron | None reported | Not reported | Obtained from study authors (see personal communication Nandhra): - Mortality - Readmission rate  Method of outcome assessment: unclear*(2)  Time period: unclear*(2) |
| Olijhoek, 2001, The Netherlands | Peer-reviewed publication | Experimental: Randomized controlled trial | 110 non-iron-deficient adult patients with baseline Hb levels 10-13 g/dl,  undergoing elective orthopedic surgery estimated to require 2-4 units of blood at one of 18 centers in five countries,  were randomly assigned  to one of 4 groups:  1) Epoetin-α + IV iron n=29, 2 men and 27 women, mean age 64.9±14.7 years, mean baseline Hb 12.0±0.8 g/dl  2) Epoetin-α + oral iron  n=29, 3 men and 26 women, mean age 65.4±13.7 years, mean baseline Hb 12.3±0.7 g/dl  3) Placebo + IV iron n=25, 3 men and 22 women, mean age 65.8±13.3 years, mean baseline Hb 12.6±0.7 g/dl  4) Placebo + oral iron n=27, 3 men and 24 women, mean age 66.9±12.1 years, mean baseline Hb 12.5±0.7 g/dl | Authors:  none  Reviewers:  included because of baseline Hb levels  (10-13 g/dl) | Authors:  Serum total iron-binding capacity (TIBC) ratio  <15 % and serum ferritin level <50 ng/ml | Epoetin-α + IV iron  - 600 IU/kg Epoetin-α subcutaneously on preoperative days 14 and 7  - 200 mg iron saccharate intravenously on preoperative days 14 and 7  Epoetin-α + oral iron  - 600 IU/kg Epoetin-α subcutaneously on preoperative days 14 and 7  - 200 mg oral iron daily during the 14 preoperative days  In addition, both ‘intervention’ groups will be compared to each other as well. | Placebo + IV iron  - Placebo subcutaneously on preoperative days 14 and 7  - 200 mg iron saccharate intravenously on preoperative days 14 and 7  Placebo + oral iron  - Placebo subcutaneously on preoperative days 14 and 7  - 200 mg oral iron daily during the 14 preoperative days  In addition, both ‘comparison’ groups will be compared to each other as well. | None reported | Not reported | - Thrombotic and/or vascular events  - Study mortality  Method of outcome assessment:  unclear*(1): not specified how thrombovascular events are detected  Time period:  until postoperative day 14 |
| Padmanabhan, 2019, UK | Peer-reviewed publication | Experimental: Randomized controlled trial | 50 anemic patients scheduled for elective cardiac surgery (coronary artery bypass graft and/or open valve surgery), with preoperative Hb levels ≥9 and <11.5 g/dl (women) and <12.5 g/dl (men), were randomly assigned to one of 2 groups:  1) IV iron  n=22, 59% men, mean age 73±12 years, mean baseline Hb 11.8±0.89 g/dl,  14% iron-deficient  2) Oral iron  n=22, 64% men, mean age 75±10 years, mean baseline Hb 11.39±1.11 g/dl, 27% iron-deficient | Authors:  WHO criteria (Hb<13 g/dl for men and <12g/dl for women) | Authors:  serum ferritin  <22 µg/l | IV iron  Ferric carboxymaltose (Ferinject) in 250 ml normal saline administered over 30 min during the preoperative clinic visit (at least 3 weeks preoperatively)  Dosing regimen:  - Hb >10 g/dl:  1000 mg (<70 kg body weight) or 1500 mg (>70 kg body weight)  - Hb 7-10 g/dl: 1500 mg (<70 kg body weight) or 2000 mg (>70 kg body weight).  A second dose was offered when required. Patients were free to decline the second dose and were allowed to remain in the study. | Oral iron  200 mg ferrous sulphate twice daily | None reported | Not reported | - Postoperative infection  - Atrial fibrillation  - Renal replacement therapy  - Gastrointestinal symptoms (including diarrhea and constipation) - Mortality  Method of outcome assessment: unclear*(2)  Time period:  until postoperative week 6-8 |
| ISRCTN2215878 | Clinical trial registration |  |  |  |  |  |  |  |  |  |
| Pinilla-Gracia, 2020, Spain | Peer-reviewed publication | Observational: Cohort study | 699 anaemic patients (% iron-deficient patients not reported) scheduled for primary total hip arthroplasty, either before (n=75, 60 women and 15 men, mean age 71±12 years) or after (n=70, 56 women and 14 men, mean age 66±16 years) the implementation of a preoperative Hb optimisation protocol with EPO and IV iron. | Authors: Hb <13 g/dL | Authors: none | Preoperative EPO + IV iron - 40 000 IU rHuEPO (Eprex®) subcutaneously  - 1 g ferric carboxymaltose (Ferinject®) intravenously both administered approximately 4 weeks prior to surgery | No preoperative EPO + IV iron | *Preoperatively, upon hospital admission* IV iron 200 mg iron sucrose per 48 hours, 3 doses  Vitamin B12 1 mg IV  Folic acid 5 mg per day orally during hospitalisation  rHuEPO If Hb <13 g/dL on admission, additional dose of 40 000 IU administered 24 hours before surgery  *Intraoperatively*  Tranexamic acid - Topical administration after skin closure - Single dose of 1 g IV tranexamic acid during the operation (at the anaesthesiologist's discretion)  *Postoperatively* IV iron see above  Folic acid see above  Heparin once-daily, weight-adjusted dosing of low molecular heparin, started 12 hours after surgery and maintained for the first 30 post-operative days | At the discretion of the anaesthesiologist and attending surgeon: - Symptoms of acute anaemia (hypotension, tachycardia, tachypnoea, dizziness, fatigue etc) - In case of no risk factors: Hb <8 g/dL - In case of ischaemic cardiomyopathy or severe peripheral vascular disease:  Hb <10 g/dL | - 6-month mortality  - Medical postoperative complications - Surgical complications  Method of outcome assessment: unclear  Time period:  - Mortality:  until 6 months after surgery  - Complications: until 30 days after surgery |
| Quinn, 2017, UK | Peer-reviewed publication | Observational: cohort study | 15 patients with iron-deficiency anemia (5 men and 10 women, mean age 71 years, mean preoperative Hb levels 9.6 g/dl; personal communication with Quinn)  scheduled for colorectal resection were either treated with:  1) IV iron  n=8  2) Oral iron n=7  Some patients were prescribed oral iron in primary care prior to  undergoing colonoscopy and therefore were treated with oral  rather than IV iron. | Authors:  WHO criteria (Hb<13 g/dl for men and <12g/dl for women)  , but for their study, the authors have lowered the cut-off level to Hb <11 g/dl | Authors:  Ferritin < 30 µg/dl OR  if Ferritin >30 µg/dl but CRP is elevated, then transferrin saturation <20 % | IV iron  Ferric carboxymaltose intravenously administered over 15 minutes, commenced a mean of 17.5 days and maximum of 27 days preoperatively (range 3-50 days).  Dosing regimen (personal communication with Quinn): the first dose was administered a min. of 10 days preoperatively based on the patient’s weight as per manufacturer’s guidelines up to the maximum single dose allowed. If a second dose was required to achieve the total maximum dose based on the patient’s weight, this was administered 7 days later. | Oral iron  Was not part of the study protocol, but was prescribed by treating doctors independently of the study. Hence, consisted of a mixture of preparations and doses at the treating doctor’s discretion (personal communication with Quinn).  Commenced a mean of 101 days preoperatively (range 39-289 days). | None reported | At the  discretion of treating clinicians; majority of clinicians targeted Hb >7 g/dL  (>10 g/dL in high risk cardiac patients) or transfused for active bleeding with  any haemodynamic instability | Postoperative complications:  - delayed wound dehiscence requiring readmission  - postoperative ileus  - postoperative myocardial infarction  - cardiac failure  - upper GI bleed  (personal communication with Quinn)  Method of outcome assessment: unclear*(2)  Time period:  until postoperative day 30.  Any readmission within 3 months was also reviewed. (see personal communication with Quinn) |
| Qvist, 1999, Denmark | Peer-reviewed publication | Experimental: Randomized controlled trial | 100 slightly anemic patients  (% of iron-deficient patients not reported) scheduled for colorectal surgery because of cancer, with Hb levels ≤8.5 mmol/L, were randomly assigned to one of 2 groups:  1) rHuEPO + oral iron n=38, 12 men and 26 women, mean age 69 years (range 48-86),  pre-entry median Hb 7.9 mmol/l (range 5.3-8.5)  2) Placebo + oral iron n=43, 20 men and 23 women, mean age 69 years (range 40-85),  pre-entry median Hb 7.6 mmol/l (range 5.1-8.5) | Authors:  Slight anemia = Hb 5-8.5 mmol/l | Authors: none | rHuEPO + oral iron  - 300 IU/kg rHuEPO (Eprex) subcutaneously on preoperative day 4  - EPO 150 IU/kg subcutaneously daily from preoperative day 3 to postoperative day 3  - 200 mg oral iron daily from preoperative day 4 to preoperative day 1 | Placebo + oral iron  - Placebo subcutaneously daily from preoperative day 4 to postoperative day 3  - 200 mg oral iron daily from preoperative day 4 to preoperative day 1 | Patients that were on drugs with a known effect on erythropoiesis and blood transfusion  within 1 month prior to study entrance were excluded. | Need for transfusion was determined by the attending anesthesiologist and surgeon in cooperation and depended on the clinical condition of each patient. No fixed Hb level was the indication alone. | - Deep venous thrombosis rate  Method of outcome assessment:  PD (see personal communication with Qvist)  Time period:  until 28 days after discharge |
| Qvist, 2000, Denmark | Peer-reviewed publication (duplicate of the 1999 article in Danish) |  |  |  |  |  |  |  |  |  |
| Richards, 2020, Australia | Peer-reviewed publication | Experimental: Randomized controlled trial | 487 adult anaemic patients (screening Hb levels ≥9 and ≤12 g/dL for women or 13 g/dL for men within 4 weeks of randomization, 28.5% iron-deficient) scheduled for elective major (>1 hour with an operative code of major, major plus or complex major operation) open abdominal (34% upper gastrointestinal, 30% gynaecological and 15% colorectal) surgery at 46 UK tertiary care centres were randomized to one of 2 groups:  1) IV iron n=244, mean median age 67 (IQR 57-72) years, 51% women, mean baseline Hb level 11.12±1.18 g/dL (28% iron-deficient)  2) Placebo n=243, mean median age 65 (IQR 50-72) years, 58% women, mean baseline Hb level 11.10±1.19 g/dL (29% iron-deficient) | Authors: WHO criteria (Hb<13 g/dL for men and <12g/dL for women) | Authors: ferritin < 100 ng/mL and  transferrin  saturations < 20% | IV iron single 1000 mg dose of ferric carboxymaltose (Ferinject ®) in 100 mL normal saline, given as an infusion over 15 minutes, a minimum of 10 days and a maximum of 42 days before planned operation | Placebo single 100 mL normal saline, given as an infusion over 15 minutes, a minimum of 10 days and a maximum of 42 days before planned operation | None reported | Information obtained from the authors (personal communication with Richards): Hb <8 g/dL | - 30-day all-cause mortality - 6-month all-cause mortality - Postoperative complication rate (defined as Clavien-Dindo classification grade III or higher)  - Readmission rate for general postoperative complications between discharge and 8 weeks - Readmission rate for general infection between discharge and 8 weeks - Readmission rate for wound infection between discharge and 8 weeks - Any readmission for complications between discharge and 6 months  - Perioperative acute kidney injury  Method of outcome assessment:  - Postoperative complications (defined as Clavien-Dindo classification grade III or higher): PD  -readmission to the hospital for complications: PD (discharge to 8 weeks and discharge to 6 months)  - Perioperative acute kidney injury: unclear  Timing of outcome assessment:  -30-day/6 months mortality: from baseline until 30 days or 6 months after the index operation  - Postoperative complications (defined as Clavien-Dindo classification grade III or higher):  from index operation to date of discharge  - Readmission to the hospital for complications: discharge to 8 weeks and discharge to 6 months)  - Perioperative acute kidney injury: unclear |
| Rineau, 2017, France | Peer-reviewed publication | Observational: Cohort study | 127 patients  (% of iron-deficient patients not reported) scheduled for primary hip or knee prosthetic surgery, with Hb levels at anesthesia visit <13 g/dl (one month before surgery) received one of 2 treatments:  1) IV iron + EPO (3^rd^ dose if Hb <15 g/dl) n=62,  49 women and 13 men,  mean age 74±10 years,  mean Hb levels at anesthesia visit 12±0.9 g/dl  2) IV iron + EPO (3^rd^ dose if Hb <13 g/dl) n=65,  62 women and 3 men, mean age 73±14 years,  mean Hb levels at anesthesia visit 12.1±0.9 g/dl | Authors:  WHO criteria (Hb<13 g/dl for men and <12g/dl for women) | Authors: none | IV iron + EPO  (3^rd^ dose if Hb <15 g/dl) - 1g ferric carboxymaltose intravenously on preoperative day 21  - 40 000 IU EPO subcutaneously on preoperative days 21 and 14 - If Hb <15 g/dl: 40 000 IU EPO subcutaneously on preoperative days 7 and 1 | IV iron + EPO (3^rd^ dose if Hb <13 g/dl) - 1g ferric carboxymaltose intravenously on preoperative day 21  - 40 000 IU EPO subcutaneously on preoperative days 21 and 14 - If Hb <13 g/dl: 40 000 IU EPO subcutaneously on preoperative days 7 and 1 | Thromboprofylaxis  low-molecular-weight heparin from the day  of surgery, 6 to 12 hours after the end of surgery | Hb <8 g/dl in the  presence of comorbidities  Hb <7 g/dl without comorbidity | Perioperative complications (personal communication with Rineau):  -acute coronary syndrome - arrhythmia  - cardiac arrest  -pulmonary oedema  - acute limb ischemia  - venous thrombosis  - pulmonary embolism  - acute kidney injury (creatinine > 25% from baseline)  - need for re-intubation or prolonged ventilation - pneumonia - stroke  - intestinal obstruction  - wound infection  - urinary tract infection  - allergies  - postoperative hematoma  Method of outcome assessment  (see personal communication with Rineau):  - Arrhythmia: postoperative monitoring. In case of heart rate abnormality, an ECG was made by the nurses and shown to the anesthetists - Deep venous thrombosis:  PD; if the surgeon suspected DVT during physical examination (pain, red skin, hot skin), a Doppler echo was performed - Intestinal obstruction:  PD; in case of non-resolutive abdominal pain, an abdominal surgeon’s opinion was asked. An abdominal scan was performed in case of abdominal pain, abdominal distention, ileus and vomiting. - Other: unclear  Time period: during hospitalization |
| Scott, 2002, USA | Peer-reviewed publication | Experimental:  Randomized controlled trial | 60 adult anaemic patients scheduled for major head and neck oncologic surgery, with Hb levels ≥10 and ≤13.5 g/dl, were randomly assigned to one of 2 groups:  1) Epoetin-α + oral iron n=29,  16 men and 13 women, mean age 68±11 years, mean baseline Hb 12.2±1.04 g/dl,  72% iron-deficient  2) Placebo + oral iron n=29, 18 men and 11 women, mean age 62±11 years, mean baseline Hb 12.3±1.15 g/dl (% of iron-deficient patients not reported) | Authors:  do not explicitly define anaemia, but refer to their patients as anaemic (Hb ≥10 and ≤13.5 g/dl) | Authors:  none  (although they state that 72% (21/29) of patients in the Epoetin-α group had baseline serum iron deficiencies) | Epoetin-α + oral iron  - 600 IU/kg Epoetin-α, 3 times: between preoperative days 19 and 10, between preoperative days 12 and 6, on the day of the surgery  - 150 mg oral iron sulphate twice per day, from the time of administration of the first dose of Epoetin-α until the day of surgery | Control + oral iron  - Placebo, 3 times: between preoperative days 19 and 10, between preoperative days 12 and 6, on the day of the surgery  - 150 mg oral iron sulphate twice per day, from the time of administration of the first dose of placebo until the day of surgery | None reported | At the discretion of the attending surgeon; effort was made not to transfuse patients with Hb levels ≥9 g/dl unless clinically indicated | - Prolonged hypertension or exacerbation of preexisting elevated blood pressure during the study  - Cerebrovascular accident  - Intraoperative myocardial infarction - Acute respiratory distress syndrome - Respiratory failure  - Deep venous thrombosis  - Rupture of the anastomosis postoperatively - Perioperative mortality  Method of outcome assessment:  unclear*(1)  Time period: unclear*(1) (probably during study duration) |
| So-Osman, 2014 | Peer-reviewed publication | Experimental: Randomized controlled trial | 730 anaemic patients scheduled for primary or revision total hip or knee replacement surgery, with preoperative Hb levels between 10 and 13 g/dl, were randomly assigned to one of the 4 groups. Of these, 683 were evaluated:  1) Epoetin-α or -β + oral iron  n=125, 113 women and 12 men, 71±12 years, baseline Hb levels 12.5±1.2 g/dL, % of iron-deficient patients not reported.  2) No treatment  n=138, 70 women and 68 men, 71±12 years, baseline Hb levels 12.6±0.8 g/dL, % of iron-deficient patients not reported  3) Epoetin-α or -β + oral iron+ autologous retransfusion  4) Autologous re-transfusion  Data from the two groups that received autologous retransfusion were not extracted (not within the scope of this project). | Authors:  WHO criteria  (Hb levels <13 g/dl (men) or <12 g/dl (women)) | Authors: none | Epoetin-α or -β + oral iron  - Preoperative EPO: 40 000 IU, epoetin-α or -β, subcutaneously once/week for 3 weeks prior and on the day of surgery, 4 injections in total.  If Hb concentrations exceeded 15 g/dL on the day of surgery, the final EPO injection was omitted.  - 200 mg oral ferrofumerate 3 times/day during 3 preoperative weeks. | No treatment | Antiplatelet agents and oral anticoagulants were discontinued - Antiplatelet  agents (nonsteroidal anti-inflammatory drugs, clopidogrel,  acetyl salicylic acid) were discontinued 3 to 10 days before  surgery according to the hospital protocol.  - Oral anticoagulants  (acenocoumarol, phenprocoumon) were discontinued  with monitoring of international normalized ratio values,  which were required to be 1.8 or lower before surgery.  Heparin 6 weeks of postoperative anti-thrombotic prophylaxis with subcutaneous low-molecular weight heparin starting the day before surgery | Transfusion triggers were based Hb levels and age and comorbidities and adopted from existing Dutch guidelines. Patients were transfused if:   1) Hb < 6.4 g/dL when < 60 years and not a risk group* 2) Hb < 8.1 g/dL when ≥ 60 years and not a risk group 3) Hb < 9.7 g/dL when a risk group   * risk group = incapability to increase cardiac output to compensate for anaemia, serious pulmonary disease or symptomatic cerebrovascular disease. | - Total number of serious adverse events  (composite measure for all thromboembolic events + non-thromboembolic events, see below)  -Thromboembolic events: composite measure + individual measures for  myocardial infarction, stroke/TIA, deep venous thrombosis, pulmonary embolism, and other thromboembolic events.  - Non-thromboembolic events: composite measure for prosthesis-related events (hip dislocations, prosthesis infections, wound infections, knee contractures, fractures), cardiovascular events (arrhythmia, blood pressure instability, etc), allergic events, infection/sepsis not prosthesis related, bleeding, and malignancy.  Method of outcome assessment:  - Deep venous thrombosis: PD via ultrasound - Myocardial infarction:  PD + additional ECG and laboratory values - Stroke:  PD + brain CT scan - Infection:  PD + positive culture  Time period:  up to postoperative month 3 |
| Stowell, 2009, USA | Peer-reviewed publication | Experimental: Randomized controlled trial | 681 adult anaemic patients  (% of iron-deficient patients not reported) scheduled for elective spinal surgery for which anticipated blood loss was 2 to 4 units at one of 80 centres in the US, with Hb levels >10 and ≤13 g/dl, were randomly assigned to one of 2 groups:  1) Epoetin-α + oral iron n=341,  36 men and 303 women, mean age 61±14 years,  mean baseline Hb 12.2±0.78 g/dl  2) Standard of care + oral iron n=340,  42 men and 298 women, mean age 59±14 years,  mean baseline Hb 12.2±0.82 g/dl | Authors:  anemia = Hb >10 and ≤13 g/dl | Authors: none | Epoetin alfa + oral iron  - 600 IU/kg Epoetin-α (Procrit) subcutaneously on preoperative days 21, 14 and 7 and on the day of surgery  - Standard of care treatment  - Oral iron therapy from preoperative day 21 until the day of surgery | Standard of care + oral iron - No ESA, treated according to the institution’s policy for blood conservation  - Oral iron therapy from preoperative day 21 until the day of surgery | No perioperative anticoagulation therapy was to be administered, although mechanical deep vein thrombosis prophylaxis was allowed | Not reported | - Death during the study or within 30 days after study completion  - Deep venous thrombosis  - Cerebrovascular accident  - Transient ischaemic attack  - Myocardial ischemia  - Myocardial infarction  - Pulmonary embolism  - Chest pain  - Superficial thrombophlebitis  - Nausea  - Post-procedural pain  - Pyrexia  - Constipation  - Pruritus  - Headache  - Muscle spasms  - Urinary tract infection  - Vomiting  - Hypokalemia  - Back pain  - Insomnia  - Wound infection - Acute myeloid leukemia  Method of outcome assessment:  - Deep vein thrombosis: PD via Color-flow Doppler imaging; - Other outcomes: unclear*(2)  Time period:  - Deep vein thrombosis: postoperative day 4 or within 24 hours of discharge (whichever was first) - Other outcomes: 30 days after the last dose of study medication or 30 days postoperatively (whichever was later) |
| Triphaus, 2019, Germany | Peer-reviewed publication | Observational: Cohort study | 1729 adults scheduled for major surgery were screened for iron-deficiency anaemia. In total, 645 patients were anaemic, of which 234 patients were diagnosed with iron-deficiency anaemia.  Of these 234 patients, 184 (73 women, mean age 63.3±16.1 years) received IV iron supplementation.   [Only data concerning the patients with iron-deficiency anaemia were extracted. Therefore, 100% of the analysed patients were iron-deficient and anaemic.] | Authors:  - Mild: Hb 11-11.9 g/dL (women) and 11-12.9 g/dL (men) - Moderate: Hb 8-10.9 g/dL (women and men)  - Severe:  Hb <8 g/dL (women and men) | Authors: ferritin <100 ng/mL and transferrin saturation <20% | IV iron  500 mg ferric carboxymaltose (Ferinject®) in 100 mL saline over 15 minutes (or 1000 mg in 200 mL saline, depending on the decision of the physician) at the earliest time point before surgery | No IV iron | Standard perioperative care | - Asymptomatic patients: Hb <6 g/dL  - Patients with cardiovascular risk factors or with clinical symptoms of anaemic hypoxia: Hb 6-8 g/dL | Separate measures for different hypersensitivity reactions (obtained from the authors; see personal communication with Meybohm): - Fever - Flush - Eczema - Angioedema - Gastrointestinal symptoms - Shock with hypotension - Tachycardia - Arrhythmia - Dyspnoea - Hyperventilation - Headache - Cardiovascular arrest - Lymph node swelling  Method of outcome assessment: - Hypersensitivity reactions: PD "After drug administration the patient was monitored  for additional 10 to 15 minutes before discharge"  Timing of outcome assessment: 10 to 15 minutes after drug administration |
| Urena, 2017, Canada | Peer-reviewed publication | Experimental: Randomized controlled trial | 104 anaemic patients (≥60 years old) with severe aortic stenosis undergoing transcatheter aortic valve implantation were randomly assigned to one of 2 groups:  1) Darbepoetin-α + IV iron n=51, of which 3 dropped out.  Demographics of 48 remaining patients:  22 men and 26 women, mean age 81±7 years, mean baseline Hb levels 10.7±1.2 g/dl,  21% iron-deficient  2) Placebo n=53, of which 1 dropped out. Demographics of 52 remaining patients: 27 men and 25 women, mean age 81±7 years, mean baseline Hb levels 11.3±1.1 g/dl, 11.5% iron-deficient | Authors:  WHO criteria (Hb<13 g/dl for men and <12g/dl for women) | Authors: ferritin <30 µg/l (personal communication with Rodés-Cabau) | Darbepoetin-α + IV iron  - 0.75 µg/kg Darbepoetin-α (Aranesp) subcutaneously on preoperative day 10(±4) and 1(±1) - 200 mg iron sucrose (Venofer) intravenously on preoperative day 10(±4) and 1(±1) | Placebo  - 0.9% saline subcutaneously on preoperative day 10(±4) and 1(±1) - 0.9% saline intravenously on preoperative day 10(±4) and 1(±1) | Antithrombotic therapy (warfarin, aspirin or clopidogrel) during hospitalization and at hospital discharge, at the discretion of the responsible physician; in patients receiving clopidogrel: - transfemoral procedure: day before surgery - transapical or transaortic procedure: 24 hours after surgery  Intraoperative heparin used during transfemoral procedure, to achieve an activated coagulation time >250 sec  Intraoperative cell saver  used during transapical and transaortic procedure  Intraoperative protamine sulphate at the end of the procedures | - Hb <7 g/dl or Hct <22 %  - Hb 7-8 g/dl and symptoms of anaemia  - Life-threatening bleeding, regardless of Hb levels | - Procedural complications (separate measures for death, conversion to open heart surgery, annulus rupture, valve embolization, need for a second valve, coronary artery occlusion, tamponade, severe hypotension requiring haemodynamic support, vascular complications, major or life-threatening bleeding, ≥ moderate aortic regurgitation)  - Outcomes within 30-days after operation (separate measures for acute kidney injury, need for dialysis, new-onset atrial fibrillation, myocardial injury, myocardial infarction, stroke, death)  - Adverse events (separate measures for ischaemic events, sepsis, heart failure, severe hypertension, hypersensitivity reactions, convulsions, abdominal pain)  Method of outcome assessment:  - Cardiac outcomes were defined and assessed according to the Valve Academic Research Consortium-2 criteria (PD and/or vital signs, imaging, laboratory values) - Adverse events (e.g. hypersensitivity reactions, sepsis, severe hypertension, abdominal pain): unclear*(2)  Time period:  postoperative day 30 |
| Weltert, 2015, Italy | Peer-reviewed publication | Experimental: Randomized controlled trial | 600 patients  (men: 96% anaemic, women: 94% anaemic,  % of iron-deficient patients not reported) undergoing elective cardiac surgery, with preoperative Hb levels ≤14.5 g/dl, were randomly assigned to one of 2 groups:  1) Epoetin-α + oral iron  n=300,  225 men and 75 women, median age 75 years (range: 47-96),  median baseline Hb levels 11.8 g/dl (range: 9.2-16.9)  2) Oral iron n=300,  219 men and 81 women, median age 74 years (range: 40-90), median baseline Hb levels 12.1 g/dl (range: 9.1-15.2) | Authors: Hb <13 g/dl  Reviewers: included because of median baseline Hb levels  (11.8 and 12.1 g/dl) | Authors: none | Epoetin-α + oral iron - 80 000 IU Epoetin-α (Eprex) subcutaneously on preoperative day 2  - 15 ml oral iron daily (Ferrolin; equivalent to 40 mg elemental iron per day) from the day of admission | Oral iron  15 ml oral iron daily (Ferrolin; equivalent to 40 mg elemental iron per day) from the day of admission | Intrapostoperative cell salvage techniques | Strict threshold of Hb <8 g/dl; allowing no discretion to physicians.  All patients requiring RBC transfusion received prestorage leukoreduced allogeneic RBC units. Using a thromboelastometric-guided approach, solvent/detergent  (S/D)-treated plasma and single-donor apheresis PLT units were administered when indicated. | - 45-day mortality (separate measures for all-cause, cardiovascular and not cardiovascular)  - Perioperative myocardial infarction  - Focal neurologic damage - Generalized neurologic damage - Cardiac tamponade - Atrial fibrillation - Acute respiratory distress syndrome - Acute pulmonary edema - Pneumonia  - Bowel ischaemia - Need for renal replacement therapy  - Neurologic complications at 45 days  - Long-term wound infection at 45 days  - Deep vein thrombosis at 45 days  - Acute hypertension onset at 45 days  - New onset renal failure at 45 days  Method of outcome assessment:  - Mortality: direct communication with family or through national or international mortality records  - Other outcomes: unclear*(2)  Time period:  until postoperative day 45 |
| Wilson, 2018, The Netherlands (A) | Peer-reviewed publication | Observational: Cohort study | 318 consecutive anaemic patients  (% of iron-deficient patients not reported) scheduled for colorectal cancer resection received one of 2 treatments:  1) IV iron n=94, 42 men and 52 women, mean age 71.8±11.1 years, mean Hb levels at diagnosis 6.12±0.89 mmol/l  2) Usual care n=224, 131 men and 93 women, mean age 73.7±9.9 years, mean Hb levels at diagnosis 6.61±0.87 mmol/l | Authors:  Hb<8.0 mmol/l, 12.9 g/dl (men) or Hb<7.5 mmol/l,  12.0 g/dl (women) | Authors: none | IV iron  IV iron therapy less than 6 weeks before surgery: 1000 to 2000 mg f iron carboxymaltose (Ferinject) or iron isomaltoside (Monofer) | Usual care  no IV iron therapy less than 6 weeks before surgery; no preoperative oral iron therapy | Some patients received neoadjuvant chemotherapy (IV iron: n=1; usual care: n=14) | According to the 4-5-6 rule, depending on the severity of the anemia and the condition of the patient | - Postoperative overall complications (composite measure for all pulmonic, cardiologic, thrombotic, infectious and neurologic complications)  Method of outcome assessment: unclear*(2)  (dataset collected by the Dutch Surgical Colorectal Audit)  Time period:  until postoperative day 30 day |
| Wilson, 2018, The Netherlands (B) | Peer-reviewed publication | Observational: Cohort study | 320 consecutive anaemic patients  (% of iron-deficient patients not reported) undergoing resection for colorectal cancer received one of 2 treatments:  1) IV iron n=102,  median age 75 years (IQR: 67-80), 52.9% male, median Hb levels at diagnosis 6 mmol/l (IQR: 5.5-7) 2) Usual care n=218,  median age 73.5 years (IQR: 66-80) , 55% male, median Hb levels at diagnosis 6.7 mmol/l (IQR: 6.1-7.3)  To correct for the baseline differences between the treatments  groups, propensity score matching was executed for tumour location,  Hb level at diagnosis, treatment approach and resection type,  and led to 83 patients in each group:  1) IV iron n=83,  median age 75 years (IQR: 67-80), 54.2% male, median Hb levels at diagnosis 6.3 mmol/l (IQR: 5.7-7.1)  2) Usual care  n=83,  median age 72 years (IQR: 66-79), 45.8% male, median Hb levels at diagnosis 6.4 mmol/l (IQR: 5.7-7.1)  Survival analyses were performed on the propensity matched groups. | Authors:  Hb<8.0 mmol/l, 12.9 g/dl (men) or Hb<7.5 mmol/l,  12 g/dl (women) | Authors: none | IV iron  1000 to 2000 mg iron carboxymaltose (Ferinject) or iron isomaltoside (Monofer) intravenously, depending on the clinical assessment and the knowledge of patient blood management of each physician | Usual care  no IV iron therapy, no preoperative oral iron therapy, no ESAs | None reported | Not explicitly mentioned in the paper, probably same as in Wilson 2018 (A) | Overall and disease-free survival (1-, 2-, 3-, 4- and 5-year)  Method of outcome assessment:  Disease-free survival was calculated from the date of surgery to the first date of radiological or pathological evidence of recurrence or metastases or the date of last follow-up, as applicable. Overall survival was calculated as the time from the date of surgery to the date of death or date of last  available follow-up.  Time period:  1, 2, 3, 4 and 5 years postoperatively |
| Ye, 2017, China | Peer-reviewed publication | Observational: Cohort study | 97 anaemic (Hb 7-10 g/dl) patients  (% of iron deficient patients not reported) scheduled for gynecological tumor surgery received one of 3 treatments:  1) rHuEPO + IV iron n=30,  mean age 45.57±3.89 years, mean Hb levels at admission 8.54±0.74 g/dl 2) IV iron n=35,  mean age 44.69±6.58 years, mean Hb levels at admission 8.52±0.90 g/dl 3) Control n=32,  mean age 43.81±7.36 years, mean Hb levels at admission 8.96±0.66 g/dl | Authors:  Hb 7-10 g/dl | Authors: none | rHuEPO + IV iron  - 10 000 IU rHuEPO (Epiao) subcutaneously administered from preoperative day 4 to postoperative day 5 - 100 mg iron sucrose intravenously daily administered from preoperative day 4 to postoperative day 5  IV iron  100 mg iron sucrose intravenously daily administered from preoperative day 4 to postoperative day 5  In addition, both ‘intervention’ groups will be compared to each other as well. | Control  no rHuEPO, no iron sucrose | The patients did not receive folic acid or vitamins. No cell-saver was used during surgery.  However, the use of tranexamic acid and thromboprophylactic medication was not recorded. (personal communication with Qiling Li) | - Hb ≤7 g/dl; - Hb >7 but <9 g/dl and obvious dizziness and fatigue;  - Hb before surgery <9 g/dl and blood loss in surgery ≥400 ml;  - Hb before surgery ≤8 g/dl and blood loss in surgery ≥200 ml. (personal communication with Qiling Li) | - Side effects (headache, mild fever, myalgia, allergy, thrombosis, gastrointestinal reaction)  - Postoperative complications (composite measure and separate measures for infection of the vaginal stump, incomplete intestinal obstruction, poor wound healing, deep venous thrombosis)  Method of outcome assessment: unclear*(2)  Time period: unclear*(2) |
| Yoo, 2011, South Korea | Peer-reviewed publication | Experimental: Randomized controlled trial | 74 anaemic patients (0% iron-deficient) scheduled for valvular heart surgery were randomly assigned to one of 2 groups:  1) rHuEPO + IV iron  n=37,  13 men and 24 women, mean age 56±12 years  2) Control n=37,  14 men and 23 women, mean age 59±12 years | Authors: WHO criteria  (Hb levels <13 g/dl for men or <12 g/dl for women) | Patients with iron deficiency anaemia were excluded from the study | rHuEPO + IV iron  - bolus injection of 500 IU/kg rHuEPO (Epocain) intravenously at 16-24 hours before surgery  - simultaneous administration of 200 mg iron sucrose (Venoferrum) in 100 ml normal saline administered intravenously over 1 hour | Control  - equivalent volume of normal saline administered intravenously over 1 hour 16-24 hours before surgery | *Intraoperative*   Crystalloid and colloid infusion before and after cardiopulmonary bypass  Tranexamic acid 1g as loading dose, followed by infusion of 200 mg per hour during surgery, and another loading dose of 1g with the onset of bypass  Cell saver  blood was reinfused into the patient before the end of surgery | - During cardiopulmonary bypass:  Hb <7 g/dl  - After bypass and postoperatively:  Hb <8 g/dl | - Postoperative death (30-day)  - Postoperative acute kidney injury  - Postoperative atrial fibrillation - Complications associated with rHuEPO therapy (separate measures for hypertension, headache, tachycardia, nausea, vomiting, hypercalcemia, diarrhea)  Method of outcome assessment: unclear*(2)  Time period:  - Postoperative kidney injury: until 48 hours postoperatively  -Atrial fibrillation: until postoperative day 7   - Post-operative death: until postoperative day 30 - Other outcomes/complications: until hospital discharge |

Abbreviations:
DVT: deep venous thrombosis
EPO: erythropoietin
Hb: haemoglobin
IU: international units
IV: intravenous
LD: laboratory-diagnosed
PD: physician-diagnosed
PICO: Population-Intervention-Comparison-Outcome
PR: patient-reported
rHuEPO: recombinant human erythropoietin
WHO: World Health Organization

### Supplementary Table 7. Risk of bias assessment.

Study limitations experimental studies

* Authors were either:

(1) not contacted by the reviewers because no valid email address of any of the authors was available;

(2) contacted by the reviewers via email. However, the authors did not respond to the email;

(3) successfully contacted by the reviewers via email. However, the authors responded that they did no longer have access to the study files and were not able to answer the reviewers’ questions.

| **Author, Year** | **Lack of allocation concealment** | **Lack of blinding** | **Incomplete accounting of outcome events** | **Inadequate measurement of the outcomes** | **Inadequate selection of the reported results** | **Other limitations** |
| --- | --- | --- | --- | --- | --- | --- |
| Biboulet, 2018 | Lack of randomization: **no** block randomization by a computer generated a random number list, and an investigator of the  Department of Medical Statistics  without clinical  involvement in the trial prepared the sequentially numbered, opaque, and sealed envelopes  Lack of allocation concealment: **no** the allocation sequence was concealed  from the anesthesiologist in sequentially numbered, opaque,  and sealed envelopes | Participants: **yes** Patient blinding was impossible. In addition, the oral iron patient group received information on the treatment’s side effects, whereas the IV iron patients were monitored with non-invasive blood pressure and continuous heart rate recording. This may have influenced the results of the participant-reported outcomes, i.e. abdominal complications (nausea, diarrhoea, constipation).  Personnel:  **yes** Although anesthesiologists and surgeons attending to the patient  during surgery were blinded to patients’ allocation, blinding of the personnel administering the oral/IV iron was impossible. This may have influenced the participant-reported outcomes, i.e. abdominal complications. | **no** only 1 of the randomized patients was lost, due to cancellation of the surgery | **yes**  - Digestive complications  (= participant-reported outcomes): The authors do not provide any information on the outcome measurement used for these outcome domains during the preoperative period (“complications related to ferrous sulfate, ferric carboxymaltose,  or epoetin-α therapy were recorded”).  It is unclear if the digestive complications were detected in a systematic (active surveillance) or non-systematic way. It is likely that the measurement may have differed between both groups (monitoring of IV iron patients during infusion). --> ‘high’ in RoB 2.0 tool  - Postoperative complications:  The authors do not provide any information on the outcome measurement used for half of these complications  (no information on cardiac or respiratory failure). It is unclear if the measurement may have differed between both groups. The authors report that they systematically collected data via telephone call 1 month after surgery, but it is unclear if they confirmed the answers of the patient using patient hospital records or not and if these outcomes involved any clinical judgement. Therefore, it is unclear if the assessment of these outcome is potentially influenced by knowledge of the intervention received. --> ‘high’ in RoB 2.0 tool | **yes**  - Digestive complications: This outcome is not mentioned in the Methods section of the paper, so probably was not included in the prespecified analysis plan, but added to the results report post-hoc.  It is unlikely that selective outcome reporting of a particular outcome measurement or of a particular analysis has occurred. --> ‘some concerns’ in RoB 2.0 tool  - Postoperative complications: In the Methods section of the paper, the authors report on the complications of interest (i.e. thromboembolic events, cardiac or respiratory failure, and death during hospitalization), the timing of the recording of these complications (postop day 3 and 5, and after 1 month) and mention that clinically suspected DVT was diagnosed using ultrasonography.  It is unlikely that selective outcome reporting of a particular outcome measurement or of a particular analysis has occurred for any of these postoperative complications. --> ‘low’ in RoB 2.0 tool | The authors mention that “the study population was too weak to properly evaluate the side effects of the preoperative anemia treatment”. This bias may put the result of this review at risk. |
| Cao, 2020 | Lack of randomization: **no**  patients were randomized by a computer randomization system  Lack of allocation concealment: **unclear*(2)**  no information provided | Participants: **yes** Patient blinding was impossible. This may have influenced the results of the participant-reported outcomes, i.e. nausea and vomiting.  Personnel: **unclear*(2)** Blinding of personnel administering the treatment was impossible. It is unclear if outcomes were influenced by this lack of blinding. | **no** all randomized patients completed the final test and were included in the analyses | **yes**  - Participant-reported outcomes (=nausea, vomiting): The authors do not provide any information on the outcome measurement used for these outcome domains during the preoperative period.  It is unclear if the digestive complications were detected in a systematic (active surveillance) or non-systematic way. It is unclear if the measurement may have differed between both groups. As the participants were aware of the intervention received, the results of the participant-reported outcomes may have been biased. --> ‘high’ in RoB 2.0 tool  - Deep venous thrombosis:  "The color  doppler examination was performed before surgery and  between 2 and 4 weeks after surgery to screen deep vein thrombosis.  Research outcomes were assessed by two professional research  assistants who were blinded to the intervention." --> 'low' in RoB 2.0 tool  - Cardiovascular events: The authors do not provide any information on the outcome measurement used.  It is unclear if these events were detected in a systematic (active surveillance) or non-systematic way. It is unclear if the measurement may have differed between both groups. The outcome assessors were blinded to the intervention. --> 'some concerns' in RoB 2.0 tool | **no** The occurrence of adverse events and complications are mentioned as outcomes of interest in the Methods section of the paper ("including perioperative cardiovascular  and cerebrovascular complications, venous thromboembolism  (VTE), allergy and other symptoms related to the adverse effects  of EPO (such as nausea, fever, headache, myalgia etc.)"), so probably were included in the prespecified analysis plan.  It is unlikely that selective outcome reporting of a particular outcome measurement or of a particular analysis has occurred. --> ‘low’ in RoB 2.0 tool | The authors themselves state that the sample size is  relatively small, missing rare treatment-related complications |
| Christodoulakis, 2005 | Lack of randomization:  **unclear*(2)** “patients were randomized (by a third party)”  Lack of allocation concealment:  **unclear*(2)**  no information provided | Participants:  **no**  Patient blinding was impossible as far as the treatment with Epoetin-α and oral iron were concerned. However, none of the adverse effect outcomes are likely to be influenced by this lack of blinding.  In addition, patients in the two EPO groups were blind to the dosage received.  Personnel:  **unclear*(2)** Blinding of personnel administering the treatment was impossible. It is unclear if outcomes were influenced by this lack of blinding. | **yes** all analyses were performed on the per-protocol population.  Of the intention-to-treat population,  19 patients were excluded for protocol violations: refused operation (n=3), refused to complete the study (n=7), underwent operation before the scheduled time (n=4), allocated to the wrong group (n=2), received preoperative transfusion outside study eligibility (n=3). | **yes**  The authors do not provide any information on the outcome measurements used.  It is unclear if measurements may have differed between both groups.  As for the adverse events leading to patient withdrawal (local rash, urticarial allergy):  it is unclear if the outcome assessor remained blinded to the intervention assignment before making the decision to withdraw the patient from the trial. --> ‘high’ in RoB 2.0 tool | **yes**  Adverse events are not mentioned as outcomes of interest in the Methods section of the paper, so were probably not included in the prespecified analysis plan, but added to the results report post-hoc. No information on protocol registration for this trial. It is unlikely that selective outcome reporting of a particular outcome measurement or of a particular analysis has occurred. --> ‘some concerns’ in RoB 2.0 tool |  |
| de Andrade, 1996 | Lack of randomization:  **no** patients were randomly assigned according to a prepared code  Lack of allocation concealment:  **unclear** no information provided | Participants:  **no** The study is called a “double-blind trial” and the authors mention that “the study drug was blinded both for dose (IU /kg) and for identity (Epoetin alfa or placebo)”.  It is unclear if participants are blinded, or if this refers to the study staff and investigators. However, outcomes are not likely to be influenced by this potential lack of blinding.  Personnel:  **no** The study drug was blinded both for dose (IU/kg) and for identity (Epoetin alfa or placebo).  To maintain the blinding among the investigators  and study staff, the reticulocyte count results  (determined by a central laboratory) were blinded  until study termination. | **no** 92% of the enrolled patients received all doses of study medication, underwent surgery and completed the study. The authors clearly report on the number  of patients who discontinued and their reasons  for discontinuing.  In addition, they performed intent-to-treat and modified intent-to-treat analyses. | **yes**  All patients, regardless of group assignment, were systematically assessed for thromboembolic adverse events: all patients underwent ultrasonography (ipsilateral, contralateral, proximal and distal) screening at fixed time points (at baseline and on postsurgery day 5, 6, 7, or on the  day of discharge if before postsurgery day 5) as part of the study protocol, an objective testing that allowed even subclinical DVTs to be detected.  Nevertheless, is it unclear if the assessors that interpreted the ultrasound were blinded to the intervention assignment, which may have substantial influence on the results. --> ‘high’ in RoB 2.0 tool | **yes**  Detection of thromboembolic events through ultrasonography screening is mentioned as a ‘Clinical evaluation’ in the Methods section of the paper. Therefore, this was probably included in the prespecified analysis plan.  However, in the Results section, the authors do not report on potential baseline differences concerning DVT rates between both groups, nor do they explicitly state how many patients were included in the stratum 2 analysis.  --> ‘some concerns’ in RoB 2.0 tool | No information on protocol registration for this trial |
| Dickson, 2020 | Lack of randomization:  **no**  patients were randomized in a 1:1 fashion via a web-based system using variable block allocation, stratified by patient sex and age  Lack of allocation concealment:  **no** Keeler confirmed via email that allocation to the researchers was concealed | Participants:  **no**  Patient blinding was impossible. However, long-term survival rates are highly unlikely to be influenced by this lack of blinding.  Personnel:  **no**  Blinding was not performed. However, long-term survival rates are highly unlikely to be influenced by this lack of blinding. | **no** no indication. Follow-up data to inform survival outcomes  were available for all 110 eligible patients. | **no** The method of measuring the outcome was probably appropriate and probably did not differ between both groups.  Although it is unclear if outcome assessors were aware of the intervention received, it is highly unlikely that the assessment of the outcome might have been influence by this lack of blinding. --> ‘low’ in RoB 2.0 tool | **no** Long-term survival rates are mentioned as primary outcomes in the Methods section of the paper, so probably were included in the prespecified analysis plan.  It is unlikely that selective outcome reporting of a particular outcome measurement or of a particular analysis has occurred. --> 'low' in RoB 2.0 tool | See limitations Keeler 2017 |
| Dousias, 2003 | Lack of randomization:  **no** a random number generator was used  Lack of allocation concealment:  **unclear*(2)** no information provided | Participants:  **no** Patients were unaware of their grouping. Controls were given similarly looking subcutaneous injections with normal saline on the same days.  Personnel:  **no** Operators were unaware of their grouping. Controls were given similarly looking subcutaneous injections with normal saline on the same days. | **unclear*(2)** insufficient information provided | **yes**  The authors do not provide any information on the outcome measurements used.  It is unclear if adverse events were detected in a systematic (active surveillance) or non-systematic way.  It is unclear if this may have differed between both groups.  As febrile episodes and hypertension can be objectively measured and do not require judgement from the observer, the assessment of the outcomes are not likely to be influenced by a potential lack of blinding.  --> ‘some concerns’ in RoB 2.0 tool | **yes**  No information on protocol registration for this trial.  Adverse events are not mentioned as outcomes of interest in the Methods section of the paper, so were probably not included in the prespecified analysis plan, but added to the results report post-hoc.  It is unlikely that selective outcome reporting of a particular outcome measurement or of a particular analysis has occurred.  --> ‘some concerns’ in RoB 2.0 tool | Bias due to selective non-reporting:  the authors use generic statements, such as “local adverse reactions did not occur”, “subcutaneous injections of rHuEPO were generally well tolerated” and “no major postoperative complications occurred” without providing any definitions, which raises at least some concerns concerning incomplete reporting of outcome domains.  This bias may put the result of this review at risk. |
| Dousias, 2005 | Lack of randomization:  **no** a random number generator was used  Lack of allocation concealment:  **no** the rHuEPO or placebo syringes were  contained in sealed, opaque envelopes | Participants:  **no** patients were unaware of their grouping; controls were given similarly looking subcutaneous injections of only water on the same days  Personnel:  **no** administration of the syringes and blood sampling took place at the tertiary institute  or local hospitals by personnel not directly involved in the study | **unclear*(2)** insufficient information provided | **yes**  The authors do not provide any information on the outcome measurement used.  It is unclear if adverse events were detected in a systematic (active surveillance) or non-systematic way.  It is unclear if this may have differed between both groups.  As fever can be objectively measured and does not require judgement from the observer, the assessment of the outcomes are not likely to be influenced by a potential lack of blinding. --> ‘some concerns’ in RoB 2.0 tool | **yes**  Adverse events are not mentioned as outcomes of interest in the Methods section of the paper, so were probably not included in the prespecified analysis plan, but added to the results report post-hoc.  It is unlikely that selective outcome reporting of a particular outcome measurement or of a particular analysis has occurred.  --> ‘some concerns’ in RoB 2.0 tool | No information on protocol registration for this trial  Bias due to selective non-reporting:  the authors use a generic statement: “no adverse reactions attributed to the drug were recorded”, which raises at least some concerns concerning incomplete reporting of outcome domains.  This bias may put the result of this review at risk. |
| Froessler, 2016 | Lack of randomization:  **no**  randomization (1:1) followed a computer-generated number  sequence (see personal communication with Froessler)  Lack of allocation concealment:  **no**  the pharmacy held the computer sequence and randomized for the authors and informed the authors of the group allocation (see personal communication with Froessler) | Participants:  **yes** Patient blinding was impossible. This may have influenced the results of the participant-reported outcomes, i.e. headache, light-headedness and back pain.  Personnel:  **unclear**  The surgeons performing the operations were blinded. Blinding of personnel administering the treatment was impossible. It is unclear if outcomes were influenced by this lack of blinding. | **no** no indication | **yes**  - Participant-reported outcomes:  The authors do not report specific outcome measurements used.  It is unclear if the adverse events were collected in a systematic (active surveillance) or non-systematic way.  It is unclear if the measurement may have differed between both groups.  As the participants were aware of the intervention received, the results of the participant-reported outcomes may have been biased. --> ‘high’ in RoB 2.0 tool  - Other adverse events:  The authors do not report specific outcome measurements used.  It is unclear if the adverse events were collected in a systematic (active surveillance) or non-systematic way.  It is unclear if the measurement may have differed between both groups.  Assessment of the outcomes may have potentially been influenced by the knowledge of the received intervention, as this assessment probably required some clinical judgement. --> ‘high’ in RoB 2.0 tool | **no**  New onset infection, respiratory failure, renal impairment, DVT and 30-day mortality are mentioned in the Methods section as secondary outcomes of interest. Therefore, this was probably included in the prespecified analysis plan.  It is unlikely that selective outcome reporting of a particular outcome measurement or of a particular analysis has occurred. --> ‘low’ in RoB 2.0 tool | Early termination is the main limitation of this study. However, ethical concerns were paramount.  5 participants randomized to the usual care group received IV iron as part of their standard care. Although this may have influenced the results, the final analysis between groups would then represent a more conservative analysis of the effects of IV iron.  Block randomization would have achieved a more equal balance in the allocation of participants. |
| Heiss, 1996 | Lack of randomization:  **unclear*(2)** no information on randomization process, although the authors mention “the sequence of erythropoietin or placebo medication was determined in advance”  Lack of allocation concealment:  **unclear*(2)** no information provided, although the authors mention “the sequence of erythropoietin or placebo medication was determined in advance” | Participants:  **no** patients were blinded for the performed application by use of preparations of EPO or placebo with identical feature and differentiated by successive numbers  Personnel:  **unclear*(2)** although the authors state that the ”investigators were blinded for the performed application by use of preparations of EPO or placebo with identical feature and differentiated by successive numbers”, it is unclear if this refers to the personnel administering the trial medication or to the outcome assessors | **no** although 3 patients in the EPO group dropped out of the study due to deep venous thrombosis at the second day in the study (n=1), refusal to continue (n=1) and suspending of the surgery (n=1) | **yes**  In the Methods section, the authors state that physical examination,  evaluation of vital signs, serum chemistry test, and blood cell counts were performed every second day until dismissal  from hospital. However, they do not provide any information on the outcome measurements for the adverse events they report in the Results section. Therefore, it is also unclear if adverse events were collected in a systematic (active surveillance) or more passive way.  It is unclear if this measurement may have differed between both groups.  Assessment of the outcomes may have potentially been influenced by the knowledge of the received intervention, as this assessment probably required some clinical judgement.  --> ‘high’ in RoB 2.0 tool | **yes**  No information on protocol registration for this trial.  Adverse events are not mentioned as outcomes of interest in the Methods section of the paper, so were probably not included in the prespecified analysis plan, but added to the results report post-hoc.  It is unclear if selective outcome reporting of a particular outcome measurement or of a particular analysis has occurred.  --> ‘some concerns’ in RoB 2.0 tool | Conflict of Interest: Cilag GmbH, the supplier of the EPO, was in some way involved in this trial (see author information). |
| Keeler, 2017 | Lack of randomization:  **no**  patients were randomized in a 1:1 fashion via a web-based system using variable block allocation, stratified by patient sex and age  Lack of allocation concealment:  **no** Keeler confirmed via email that allocation to the researchers was concealed. | Participants:  **yes**  Patient blinding was impossible. This may have influenced the results of the participant-reported outcomes, i.e. dyspepsia, constipation, post-infusion headache.  Personnel:  **unclear*(2)**  There was no blinding of the treatment. It is unclear if the outcomes were influenced by this lack of blinding. | **no**  The authors provide a clear overview of the problems that occurred during follow-up (e.g. did not undergo surgical resection, operation date expedited, dose reduced…). In addition, they performed an intention-to-treat analysis. | **yes**  - Participant-reported outcomes (dyspepsia, constipation, post-infusion headache):  the End of study report states that “All adverse events occurring during the study observed by the investigator or reported by the  participant, whether or not attributed to study medication, were recorded on the CRF”. However, it does not contain any information on the outcome measurements.  Hence, it is unclear if the participants were asked to report adverse events in a systematic (active surveillance) way or not.  It is also unclear if the measurement may have differed between both groups. As the participants were aware of the intervention received, the results of the participant-reported outcomes may have been biased. --> ‘high’ in RoB 2.0 tool  - Other adverse events (rash, grade of infective complication severity):  in the EudraCT End of study report, the authors clearly mention which assessments should be done and when. However, it does not contain any information on the outcome measurements of the different post-operative complications.  Measurements are unlikely to have differed between both groups. Assessment of the outcomes may have potentially been influenced by the knowledge of the received intervention, as this assessment probably required some clinical judgement.  --> ‘high’ in RoB 2.0 tool | **yes**  It is unclear which adverse events were included in the pre-specified analysis plan. In the EudraCT End of study report, the authors elaborate on the reporting procedures for all adverse events, and provide a clear overview of the number and specific types of adverse events that occurred at the different research centers, but it seems as if these adverse events only refer to the ones experienced by the IV iron group.  It is unlikely that selective outcome reporting of a particular outcome measurement or of a particular analysis has occurred.  --> ‘some concerns’ in RoB 2.0 tool | Conflict of interest: *Disclosure:* The research department of M.J.B. has received grant support from Syner-Med (UK) and Vifor Pharma (Switzerland).M.J.B. has received honoraria and travel support for consulting or lecturing from Vifor Pharma and Merck Sharp and Dohme Limited (UK). The research department of A.G.A. has received grant support from Syner-Med (UK), Vifor Pharma (Switzerland) and Pharmacosmos (Denmark). A.G.A. has received honoraria and travel support for consulting or lecturing from Ethicon Endosurgery (UK), Johnson & Johnson (UK), Olympus (UK) and Vifor Pharma (Switzerland). O.N. has received  honoraria and travel support for consulting from Pharmacosmos,  Denmark.  One key limitation of the study was that overall transfusion use was lower than anticipated, which may render the study vulnerable to type II error.  It is possible that the duration of preoperative therapy (21 days) was insufficient to allow this treatment to have maximal effect on haemoglobin levels, which could have influenced the use of  blood transfusion, which may in its turn have affected the occurrence of complications. |
| Keeler, 2019 | Lack of randomization:  **no**  patients were randomized in a 1:1 fashion via a web-based system using variable block allocation, stratified by patient sex and age  Lack of allocation concealment:  **no** Keeler confirmed via email that allocation to the researchers was concealed. | Participants:  **no**  Patient blinding was impossible. However, it is highly unlikely that this lack of blinding would have influenced infection outcomes.  Personnel:  **unclear*(2)**  There was no blinding of the treatment. It is unclear if the outcomes were influenced by this lack of blinding. | **no**  see Keeler 2017: The authors provide a clear overview of the problems that occurred during follow-up (e.g. did not undergo surgical resection, operation date expedited, dose reduced…). In addition, they performed an intention-to-treat analysis. | **yes** The authors do not provide any information on the outcome measurements of the different post-operative complications.  Measurements are unlikely to have differed between both groups. Assessment of the outcomes may have potentially been influenced by the knowledge of the received intervention, as this assessment probably required some clinical judgement.  --> ‘high’ in RoB 2.0 tool | **yes**  see Keeler 2017: it is unclear which adverse events were included in the pre-specified analysis plan. In the EudraCT End of study report, the authors elaborate on the reporting procedures for all adverse events, and provide a clear overview of the number and specific types of adverse events that occurred at the different research centers, but it seems as if these adverse events only refer to the ones experienced by the IV iron group.  It is unlikely that selective outcome reporting of a particular outcome measurement or of a particular analysis has occurred.  --> ‘some concerns’ in RoB 2.0 tool | See Keeler 2017 |
| Kettelhack, 1998 | Lack of randomization:  **unclear*(3)** no information on randomization process  Lack of allocation concealment:  **unclear*(3)** no information provided | Participants:  **unclear*(3)** the study is called a double-blind study, but no information is provided on the blinding process. This potential lack of blinding may have influenced the results of the participant-reported outcomes, e.g. constipation.  Personnel:  **unclear*(3)** the study is called a double-blind study, but no information is provided on the blinding process. It is unclear if the outcomes were influenced by this lack of blinding. | **no** although 7 patients (4 in Epoetin beta group and 3 in placebo group) were excluded from analysis, due to adverse events before surgery (Epoetin beta n=2), preoperative transfusion (Placebo n=2), no hemicolectomy because colonic carcinoma was not confirmed at operation (Epoetin beta n=2), performance of palliative ileotransverse colostomy (Placebo n=1) | **yes**  No information available in the paper. Kettelhack confirmed by email that case report forms were systematically filled out, that outcomes were probably patient-reported, that complications noted were documented in the patient records, but that the grade of validation is not certain.  - Participant-reported complications (constipation, chills):  The authors do not report specific outcome measurements used.  It is unclear if the adverse events were collected in a systematic (active surveillance) or non-systematic way.  It is unclear if the measurement may have differed between both groups.  As the participants were aware of the intervention received, the results of the participant-reported outcomes may have been biased. --> ‘high’ in RoB 2.0 tool  - Other complications:  The authors do not report specific outcome measurements used.  It is unclear if the adverse events were collected in a systematic (active surveillance) or non-systematic way.  It is unclear if the measurement may have differed between both groups.  Assessment of the outcomes may have potentially been influenced by the knowledge of the received intervention, as this assessment probably required some clinical judgement. --> ‘high’ in RoB 2.0 tool | **yes** No information on protocol registration for this trial.  Adverse events are not mentioned as outcomes of interest in the Methods section of the paper, so were probably not included in the prespecified analysis plan, but added to the results report post-hoc.  It is unclear if selective outcome reporting of a particular outcome measurement or of a particular analysis has occurred.  --> ‘some concerns’ in RoB 2.0 tool |  |
| Khalafallah, 2012 | Lack of randomization:  **no**  randomization in blocks of 4  Lack of allocation concealment: **unclear*(2)**  no information provided | Participants:  **no**  patient blinding was impossible, but adverse event outcome data are not likely to be influenced by this lack of blinding  Personnel: **unclear*(2)**  no information provided on blinding. It is unclear if adverse event outcome data were influenced by this potential lack of blinding. | **no**  the authors provide a clear overview of the reasons of patient drop-out (see Figure 1: reaction to oral iron, changed their mind against IV iron, operation delayed or cancelled). Moreover, drop-out was similar between both treatment groups (oral iron: n=5, IV iron: n=6) and reasons for drop-out are unlikely to be connected with the studied outcomes. | **yes**  The authors do not report specific outcome measurements used.  It is unlikely that the measurement may have differed between both groups.  Assessment of the outcomes may have potentially been influenced by the knowledge of the received intervention, as this assessment probably required some clinical judgement. --> ‘high’ in RoB 2.0 tool | **yes**  Trial registration ACTRN12609000596 only contains a general note on the assessment of “complications” as a secondary outcome.  The assessment of postoperative complications is mentioned as a secondary outcome in the Patients and Methods section of the paper.  Therefore, this was probably included in the prespecified analysis plan.  It is unclear if selective outcome reporting of a particular outcome measurement or of a particular analysis has occurred. --> ‘some concerns’ in RoB 2.0 tool | Short preoperative duration of treatment with oral iron in some patients  The epidural anaesthesia used during the surgery may have contributed to reduced blood loss, and therefore reduced rates of blood transfusions, which may have influenced postoperative complication rates.  Bias due to selective non-reporting:  the authors use a generic statement: “Assessment of infection complications during and after the operative procedure showed no statistical significance between patients in both  treatment groups.”  and do not provide any further information on the type of infectious complications studied, which raises at least some concerns concerning selective non-reporting.  This bias may put the result of this review at risk. |
| Kim, 2009 | Lack of randomization:  **no**  a computer-generated randomization table was used, and group allocation was determined by one of the authors who was not involved in patient care  Lack of allocation concealment: **unclear**  no information provided | Participants:  **yes**  Patient blinding was impossible. In addition, IV iron patients were actively asked to note any symptoms or adverse effects of treatment before, during and after each infusion.  This may have influenced the results of the participant-reported outcomes, i.e. nausea, dyspepsia, myalgia, injection pain.  Personnel:  **unclear**  no information provided. It is unclear if outcomes were influenced by this potential lack of blinding. | **no**  Fifty-six participants  with >80% compliance completed the trial in the intravenous iron (n = 30, 76.9%) and oral iron (n = 26, 70.3%) groups. | **yes**  Physical findings, including vital signs, were measured before, during and after each infusion of IV iron, and “by regular contact with study nurses” in the oral iron group. In the IV iron group, patients were asked to note any symptoms or adverse effects of treatment. It is unclear if this was also the case in the oral iron group. It is therefore likely that the measurement may have differed between both groups.  --> ‘high’ in RoB 2.0 tool | **yes** No information on protocol registration for this trial. Recording of any symptoms or adverse effects of treatment is mentioned in the Methods section. Therefore, this was probably included in the prespecified analysis plan.  It is unclear if selective reporting of a particular outcome measurement and/or particular analysis has occurred.  --> ‘some concerns’ in RoB 2.0 tool | 3 weeks of follow-up may be insufficient for oral iron treatment in general medical conditions  Premature cessation of the oral iron arm.  Bias due to selective non-reporting:  the authors use generic statements, such as “no severe adverse events were observed in the 2 groups, and only some tolerable adverse events were observed in each group” without providing any definitions, which raises at least some concerns concerning selective non-reporting.  This bias may put the result of this review at risk. |
| Kosmadakis, 2003 | Lack of randomization:  **unclear*(1)** no information on randomization process  Lack of allocation concealment:  **unclear*(1)** no information provided | Participants:  **no**  patients are said to be blinded to the performed application; this is unlikely to have influenced adverse event data  Personnel:  **unclear*(1)** “investigators” are said to be blinded to the performed application, but it is not clear whether this refers to the personnel or the outcome assessors. | **no**  although 12 randomized patients were excluded from the study because of blood transfusions within 1 month before the study (n=2), personal reasons (n=4), protocol violation (n=2) and distant metastases proven at operation (n=4) | **yes**  The authors report that patients were admitted for 2 hours in an outpatient manner while  remaining under continuous surveillance for possible adverse  effects. However, they do not report on the methods used to detect possible post-operative complications, nor do they report specific outcome measurements used.  It is unclear if the outcome measurements used differed between both groups.  Assessment of the outcomes may have potentially been influenced by the knowledge of the received intervention, as this assessment probably required some clinical judgement. --> ‘high’ in RoB 2.0 tool | **yes**  No information on protocol registration for this trial.  Continuous surveillance for adverse effects during treatment was mentioned in the Methods section. Postoperative complications were not mentioned as outcomes of interest, so were probably not included in the prespecified analysis plan, but added to the results report post-hoc.  It is unclear if selective outcome reporting of a particular outcome measurement or of a particular analysis has occurred.  --> ‘some concerns’ in RoB 2.0 tool | Bias due to selective non-reporting:  the authors use a generic statement: “no major allergic reactions were reported”, without providing any definitions, which raises at least some concerns concerning selective non-reporting.  This bias may put the result of this review at risk. |
| Larson, 2001 | Lack of randomization:  **unclear*(2)** no information on randomization process  Lack of allocation concealment:  **unclear*(2)** no information provided | Participants:  **no** patient blinding was impossible, but outcomes are not likely to be influenced by this lack of blinding  Personnel:  **unclear*(2)** Blinding of personnel administering the treatment was impossible. It is unclear if outcomes were influenced by this lack of blinding. | **no** but one patients from the oral iron group was lost to follow-up due to severe streptococcal septicemia | **yes** The authors mention that the safety and tolerability of EPO was evaluated  by reported adverse events, laboratory tests and blood pressure measurements. They do not provide any additional specific information. In addition, it is unclear if this was also evaluated in the oral iron group.  It is therefore likely that the measurement may have differed between both groups.  --> ‘high’ in RoB 2.0 tool | **yes**  In the Methods section, the authors mention that the safety and tolerability of EPO was evaluated. However, they do not specify which outcomes were considered relevant. This suggests that the outcomes were probably not included in the prespecified analysis plan, but added to the results report post-hoc. It is unclear if selective reporting of a particular outcome measurement and/or a particular analysis has occurred.  --> ‘some concerns’ in RoB 2.0 tool | No information on protocol registration for this trial  Bias due to selective non-reporting:  the authors use the generic statement “no adverse events related to the study drug were observed”, which raises at least some concerns concerning selective non-reporting.  This bias may put the result of this review at risk. |
| Lee, 2019 | Lack of randomization:  **no**  patients were randomized 1:1 by a computer randomization system  Lack of allocation concealment: **unclear*(2)**  no information provided | Participants:  **no**  no blinding (open-label trial), but the outcomes of anaphylaxis and mortality cannot be influenced by this lack of blinding  Personnel:  **no**  No blinding (open-label trial). However, it is unlikely that the outcome of anaphylaxis is influenced by this lack of blinding. | **no**  no drop-outs in both groups reported | **yes**  The authors mention that “safety was assessed throughout the study by monitoring the severity of adverse events. The  National Cancer Institute Common Terminology Criteria  for Adverse Events (CTCAE version 4.0) were used to report adverse event and safety data.”.  It is unclear if the outcome measurements used differed between both groups.  Although adverse events are adjudicated by physicians via the CTCAE rating system, assessment of anaphylaxis is unlikely to have been influenced by the knowledge of the received intervention.  --> ‘some concerns’ in RoB 2.0 tool | **yes**  No information on protocol registration for this trial.  Adverse events are not mentioned as outcomes of interest in the Methods section of the paper. This suggests that they were probably not included in the prespecified analysis plan, but added to the results report post-hoc.  It is unclear if selective outcome reporting of a particular outcome measurement or of a particular analysis has occurred. --> ‘some concerns’ in RoB 2.0 tool | Lack of measurement of long-term outcomes  This study was supported by an unrestricted educational grant from JW Pharma and Vifor Pharma. The analysis of this study was not affected by any employees of the pharma companies.  Bias due to selective non-reporting:  the authors use generic statements, such as “no patients withdrew from this study due to a major adverse event” and “All adverse events were mild in  severity, not requiring any intervention (CTCAE  grade 1), and the most common adverse event was  headache (n = 4)”. Moreover, they do not provide any definition on “major adverse event” (neither does the CTCAE), and do not provide information on the other adverse events besides headache. |
| Olijhoek, 2001 | Lack of randomization:  **no** patients were randomly assigned in a 1:1:1:1 ratio according to a randomization schedule  Lack of allocation concealment:  **unclear*(1)** no information provided | Participants:  **no** The study is called “a double-blind study” and the authors mention that “the study was blinded for identity of study drug  (epoetin alfa or placebo), but not for iron administration  (oral or IV)”. It is unclear if participants are blinded, or if this refers to the study staff and investigators. However, adverse event data are not likely to be influenced by this potential lack of blinding.  Personnel:  **no** The study is called “a double-blind study” and the authors mention that “the study was blinded for identity of study drug  (epoetin alfa or placebo), but not for iron administration  (oral or IV)”.  However, adverse event data are not likely to be influenced by this potential lack of blinding. | **no** 107 out of the 110 patients (97%) completed the study. The authors describe the reasons for withdrawal of the 3 patients in detail. | **yes**  The authors mention that “Safety  evaluations included assessment of clinical laboratory tests, vital signs, physical examinations, and the incidence and severity of adverse events, including thrombotic/vascular  events. On Day 14 or at the time of early withdrawal, the physical exam (including vital signs) and the clinical laboratory  tests (including hematology, serum chemistry, urinalysis,  serum iron, TIBC, and serum ferritin) were repeated.”  However, it is unclear which outcome measurement was used. In addition, it is unclear if the outcome measurements used differed between both groups. It is unclear if the measurement is likely to be influenced by knowledge of the received intervention. --> ‘high’ in RoB 2.0 tool | **yes**  No information on protocol registration for this trial.  Recording of adverse events was part of the study schedule. Therefore, this was probably included in the prespecified analysis plan.  It is unclear if selective reporting of a particular outcome measurement and/or particular analysis has occurred.  --> ‘some concerns’ in RoB 2.0 tool | Bias due to selective non-reporting:  The authors do not provide clear data on the adverse events in the 4 different groups (e.g. only compares adverse events in the IV iron patients to those in the oral iron patients, even though both groups consist of 2 subgroups (Epoetin-alfa and placebo).  In addition, they provide rather generic statements: “Only nausea, constipation,  and diarrhea were reported by more than one patient.  There were no clinically meaningful changes in laboratory values, vital signs, or findings from physical examination.”  This raises at least some concerns concerning selective non-reporting.  This type of bias may put the result of this review at risk. |
| Padmanabhan, 2019 | Lack of randomization: **unclear*(2)**  treatment allocation  was decided by random numbers  Lack of allocation concealment:  **no**  opaque concealed envelopes were used | Participants:  **yes**  Patient blinding was impossible. All patients received information on the most common side effects of both treatments (see clinical trial registration ISRCTN2215878). This may have influenced the results of the participant-reported outcomes, i.e. gastrointestinal symptoms (diarrhoea, constipation).  Personnel:  **yes**  Blinding of the personnel administering the oral/IV iron was impossible. This may have influenced the participant-reported outcomes, i.e. gastrointestinal symptoms (diarrhoea, constipation). | **yes**  in the Discussion, the authors report that there were  high rates of patients randomized to intravenous iron declining their second dosage of intravenous iron.  In addition, the intention-to-treat analysis includes patients who withdrew from the study but had undergone iron therapy and/or surgery. Including patients who did not undergo surgery may have influenced the postoperative outcome data. | **yes**  - Gastrointestinal symptoms  (= participant-reported outcomes): The authors do not provide any information on the outcome measurement used for these outcome domains.  It is unclear if they were detected in a systematic (active surveillance) or non-systematic way. It is likely that the measurement may have differed between both groups (monitoring of IV iron patients before, during and after infusion).  --> ‘high’ in RoB 2.0 tool  - Postoperative complications:  The authors do not provide any information on the outcome measurement used for these outcome domains.  It is unclear if they were detected in a systematic (active surveillance) or non-systematic way.  It is unclear if the outcome measurements used differed between both groups.  Assessment of the outcomes may have potentially been influenced by the knowledge of the received intervention, as this assessment probably required some clinical judgement.  --> ‘high’ in RoB 2.0 tool | **yes**  Postoperative complications  (acute kidney injury, atrial fibrillation and any infection,  such as chest infection, surgical site infection and  septicaemia) are mentioned as secondary outcomes of interest in the Methods section of the paper. However, other types of adverse events are not, so may not have included in the prespecified analysis plan, but added to the results report post-hoc.  It is unclear if selective outcome reporting of a particular outcome measurement or of a particular analysis has occurred. --> ‘some concerns’ in RoB 2.0 tool | There were high rates of patients randomized to IV iron that declined their second dosage. |
| Qvist 1999/2000 | Lack of randomization:  **no** no information on randomization process in the manuscript.  The author confirmed via email that consecutively numbered envelopes with corresponding inclusion were used (see personal communication Qvist).  Lack of allocation concealment:  **no** no information provided in the manuscript. The author confirmed via email that opaque sealed envelopes were used (see personal communication Qvist). | Participants:  **no** The study is called a double-blind study, but no information is provided on the blinding process. However, DVT rates are not likely to be influenced by this potential lack of blinding.  Personnel:  **unclear** the study is called a double-blind study, but no information is provided on the blinding process | **no** but 19 patients (11 in the EPO, 8 in the placebo group) were excluded from analysis, because of death within 2 weeks after surgery due to widespread  neoplastic disease (n=2),  personal reasons (n=6) and  protocol violation (n=11). | **yes**  DVTs were assessed via physical examination (see personal communication with Qvist).  It is however unclear if they were detected in a systematic (active surveillance) or non-systematic way. It’s also unclear if the authors looked at pre-existing DVTs.  It is unclear if this measurement used differed between both groups.  As outcome assessors were blinded to the received intervention, the assessment will not have been influenced.  --> ‘some concerns’ in RoB 2.0 tool | **yes**  No information on protocol registration for this trial.  Adverse events are not mentioned as outcomes of interest in the Methods section of the paper. This suggests that the outcomes were probably not included in the prespecified analysis plan, but added to the results report post-hoc.  It is unclear if selective outcome reporting of a particular outcome measurement or of a particular analysis has occurred. --> ‘some concerns’ in RoB 2.0 tool | Conflict of interest: the study was funded by Janssen-Cilag, the company supplying the EPO  Bias due to selective non-reporting: the authors mention that “No adverse events were reported to be drug-related by the investigators.”. This phrase raises at least some concerns concerning selective non-reporting.  This type of bias may put the result of this review at risk. |
| Richards, 2020 | Lack of randomization:  **no** 1:1 randomization via a secure web-based service using minimization, considering baseline Hb (<100 vs ≥100 g/L), age  (<70 vs ≥70 years), centre, and operation type (major, major  plus, complex major)  Lack of allocation concealment:  **no** The randomisation allocation was set up by Sealed Envelope (an internet randomisation service provider). The unblinded staff completed the concealed treatment allocation. | Participants:  **no** their skin  was swabbed with iodine, and the study treatment was shielded from vision (light protection bags) and infused  through black tubing  Personnel:  **no** dedicated unblinded study personnel  were responsible for the preparation and administration of the study drug but had no other involvement in the trial. Other clinical and research staff were blinded to treatment allocation. | **no** Even at postoperative month 6, only 7% of all participants was lost to follow-up. The authors provide a clear overview of the reasons for drop-out (see Figure 1). | **yes**  The authors do not provide any information on the outcome measurement used.  It is unclear if they were detected in a systematic (active surveillance) or non-systematic way.  It is unclear if the outcome measurements used differed between both groups.  Outcome assessors were blinded to treatment allocation. --> 'some concerns' in RoB 2.0 tool | **yes** The outcomes reported do not completely correspond with the outcomes mentioned in the [published protocol](https://preventt.lshtm.ac.uk/protocol-3/). As for the postoperative complications, the protocol mentioned the use of the Post-Operative Morbidity Survey (POMS), consisting of 18 items. Nevertheless, these data are not reported in the published paper or appendix. The authors only report on the number of complications with a CD grade III or above. Moreover, in the appendix, they mention that they only identified the most severe postoperative complication for each patient. This raises some concerns concerning selective outcome reporting. --> 'some concerns' in RoB 2.0 tool | Preoperative iron deficiency was not specifically defined as an inclusion criterion.  Conflict of interest: Multiple authors report grants and support from multiple pharma companies |
| Scott, 2002 | Lack of randomization:  **unclear*(1)** no information on randomization process  Lack of allocation concealment:  **unclear*(1)**  no information provided | Participants:  **no** patients were blinded to the drug administered by use of identical Epoetin alfa and placebo preparations that were successively numbered  Personnel:  **no**  investigators were blinded to the drug administered by use of identical Epoetin alfa and placebo preparations that were successively numbered | **no** but one patient from each group was disqualified from the study as a result of surgery cancellation after enrolment and receiving the study medication. | **yes**  The authors do not provide any information on the outcome measurement used for the perioperative adverse events.  It is unclear if they were detected in a systematic (active surveillance) or non-systematic way.  It is unclear if the outcome measurements used differed between both groups.  Assessment of the outcomes may have potentially been influenced by the knowledge of the received intervention, as this assessment probably required some clinical judgement.  --> ‘high’ in RoB 2.0 tool | **yes**  No information on protocol registration for this trial. Adverse events are not mentioned as outcomes of interest in the Methods section of the paper. This suggests that the outcomes were probably not included in the prespecified analysis plan, but added to the results report post-hoc.  It is unclear if selective outcome reporting of a particular outcome measurement or of a particular analysis has occurred. --> ‘some concerns’ in RoB 2.0 tool | The study was funded by Ortho Biotech Products, L.P.. Although it is not explicitly mentioned in the text, this company is probably the provider of the Epoetin alfa.  Bias due to selective non-reporting:  The authors mention that: “Overall, the use of epoetin alfa in head and neck  cancer patients was well tolerated in this study. No patients discontinued the study medication as a result of side effects during the preoperative dosing period.”. This may indicate that patients experienced some minor side effects that were not reported by the authors, which raises at least some concerns concerning selective non-reporting.  This type of bias may put the result of this review at risk. |
| So-Osman, 2014 | Lack of randomization:  **no** computer-generated randomization was used  Lack of allocation concealment:  **no** for each subject to be randomized, a sheet of paper  with all relevant stratification and group-allocation information  was produced and placed in a sealed opaque envelope.  The exact moment of opening  the envelope and its associated sequence number was verified  against a centrally stored randomization list to check for  selection bias. | Participants:  **no** Due to the nature of the interventions, to avoid protocol  violations, clinical-site staff members, clinicians, research  nurses, and patients were aware of study group assignments. However, outcomes are not likely to be influenced by this lack of patient blinding.  Personnel:  **no** Due to the nature of the interventions, to avoid protocol  violations, clinical-site staff members, clinicians, research nurses, and patients were aware of study group assignments.  The chart data were written on the Case Report Form by the research nurses. All written information was transferred  from the paper Case Report Form to the secure online Webbased  data management system (ProMISe) of the department  of Medical Statistics and BioInformatics in Leiden. A  built-in quality management system checked for irregularities,  inconsistencies, and coding errors, and clarification was asked for whenever necessary.  Hence, adverse event data are not likely to be influenced by this potential lack of blinding. | **no** of the 47  not-evaluated patients, for the majority (83%) surgery had  been cancelled or performed elsewhere, six of these patients  had received at least one erythropoietin dose. | **yes**  The authors do not provide any information on the outcome measurement used for the perioperative adverse events, except for DVT (diagnosed by ultrasound and not based on active surveillance). It is unclear if the outcome measurements used differed between both groups.  Assessment of the outcomes may have been influenced by the knowledge of the received intervention, as this assessment probably required some clinical judgement. --> ‘high’ in RoB 2.0 tool | **yes**  Trial registration (ISRCTN96327523) mentions the assessment of postoperative complications. Adverse events are mentioned in the Method’s “outcome measures” section of the paper, but are not explicitly designated primary or secondary outcomes.  However, it is likely that they were included in the prespecified analysis plan, given that the authors explicitly state that “serious adverse events were reported up to 3 months and were defined as death, life-threatening events, (prolongation of) hospitalization, and/or events resulting in persistent disability, and categorized into prosthesis related (dislocation, wound  infection or deep prosthetic infection, fractures, or limitation in movement), thromboembolic (deep venous thrombosis diagnosed by ultrasound and not based on active surveillance, pulmonary  emboli, stroke or transient ischemic attack, myocardial  infarction), other cardiovascular events, allergy, infection/sepsis  (not prosthesis related), malignancy, and other events.”  It is unclear if selective outcome reporting of a particular outcome measurement or of a particular analysis has occurred.  --> ‘some concerns’ in RoB 2.0 tool | The study was not powered  for safety evaluation. Hence, the authors were unable to draw valid conclusions  on the incidence of adverse complications.  In addition, all patients  received thrombosis-prophylaxis, which may have influenced the proportion of thromboembolic complications in the EPO group.  Bias due to selective non-reporting: the authors use a composite measure for non-thromboembolic events and do not specify the different types of events. This raises at least some concerns concerning selective non-reporting.  This type of bias may put the result of this review at risk. |
| Stowell, 2009 | Lack of randomization: **no** patients were randomized using a computer-generated randomization schedule and an interactive voice-response system in a 1:1 ratio  Lack of allocation concealment:  **yes** no allocation concealment | Participants:  **yes**  Participants were not blinded. This may have influenced the participant-reported outcomes, i.e. nausea, constipation, headache, back pain etc.  Personnel: **yes**  The personnel was not blinded. This may have influenced the participant-reported outcomes (i.e. nausea, constipation, headache, back pain etc.) | **no**  In total, 99 of the 680 patients (14.5%) discontinued the study (17.6% in the EPO group, 9.7% in the standard of care group).  However, in the paper, the authors have reported data on adverse events in the intent-to-treat population.  In the Clinical Study Report (PR97-19-002), the authors mention that they have also performed analysis on the evaluable population (all patients who had surgery and underwent postoperative Doppler imaging for evaluation of symptomatic and asymptomatic DVT) and the per-protocol population (the evaluable population with no major protocol deviations and with interpretable Doppler imaging). Here, they briefly mention that “Similar results were seen in the evaluable and per-protocol populations”. | **yes**  All patients, regardless of group assignment, were systematically assessed for DVT: all patients underwent Doppler imaging on postoperative day 4 or within 24 hours of discharge (whichever came first), as part of the study protocol, an objective testing that allowed even subclinical DVTs to be detected.  Doppler  images were reanalyzed by an independent reviewer at a core  laboratory who was blinded to the local findings and the study arm assignment. If the local and core laboratory interpretations  differed, a third party adjudicator blinded to the prior interpretations  and the study arm assignment reviewed the image and  rendered the definitive interpretation.  Unfortunately, the study investigators did not perform baseline ultrasound scanning to exclude or balance preexisting deep vein thrombosis.  As for the other types of adverse events, outcome measurements are not reported.  It is unclear if this may have differed between both groups.  As participants were not blinded, the participant-reported outcomes (e.g. headache, pruritus, nausea etc.) may have been influenced. For the other outcomes (e.g. urinary tract infection, wound infection) it is unclear if the assessment of these outcome is potentially influenced by knowledge of the intervention received.  --> ‘high’ in RoB 2.0 tool | **no** In the Methods section of the paper, the authors clearly indicate that Color-flow Doppler imaging is systematically used to screen for DVT.  From the Clinical Study Report (PR97-19-002), it is clear that the detection of DVT, other thromboembolic events and other types of adverse events, were part of the study protocol, and hence were included in the prespecified analysis plan.  It is highly unlikely that selective outcome reporting of a particular outcome measurement or of a particular analysis has occurred. --> ‘low’ in RoB 2.0 tool |  |
| Urena, 2017 | Lack of randomization:  **no** patients were randomized by a technician who was not otherwise involved in the study using a computer-generated randomisation  schema in a 1:1 ratio, stratified by planned surgical route  Lack of allocation concealment:  **no** treatment assignments  were concealed from the investigators gathering data and assessing outcome and safety events until the study was complete | Participants:  **unclear*(2)** The study is called a double-blind trial, but there is no specific mentioning of the blinding process and whether this applies to the participants. The participant-reported outcome (i.e. abdominal pain) may have been influenced by this potential lack of blinding.  Personnel:  **no** Both the EPO (+iron sucrose) and placebo  were administered by a nurse not involved in the screening, randomisation  or assessing of patient outcomes. | **no** no missing data; there were no patient withdrawals between the time of randomisation and treatment administration, and no patients were lost at follow-up | **yes**  In the Methods section, the authors mention that their secondary outcomes include stroke, myocardial infarction, new-onset atrial fibrillation etc. They provide definitions and methods for diagnosis for each of these outcomes by referring to the Valve Academic Research Consortium-2 criteria.  However, they do not provide any definition or information on the detection of other adverse events (e.g. hypersensitivity reactions, sepsis, severe hypertension). It is unlikely that the outcome measurements used differed between both groups.  Assessment of the outcomes may have potentially been influenced by the knowledge of the received intervention, as this assessment probably required clinical judgement. --> ‘high’ in RoB 2.0 tool | **yes**  No information on protocol registration for this trial.  In the Methods section of the paper, the authors mention that their secondary outcomes include rates of 30-day mortality,  myocardial infarction, stroke, and the combined endpoint of red cell transfusion, myocardial infarction or stroke, the incidence of acute kidney injury, need for haemodialysis and new-onset atrial  fibrillation.  It is likely that these adverse event outcomes were included in the prespecified analysis plan.  The other outcomes (hypersensitivity, sepsis, severe hypertension) were probably not included in the prespecified analysis plan, but added to the results report post-hoc.  It is unclear if selective outcome reporting of a particular outcome measurement or of a particular analysis has occurred. --> ‘some concerns’ in RoB 2.0 tool | Conflict of interest: this trial was partially sponsored by unrestricted grants from Edwards Lifesciences (medical company specialized in artificial heart valves)  The response to EPO might be different in younger and lower-risk patients  Patients underwent TAVI using older-generation  devices; these results might be different when using newer-generation  transcatheter heart valves |
| Weltert, 2015 | Lack of randomization:  **no** a permuted block randomization scheme with 1:1 allocation was used to keep the group sizes equal  Lack of allocation concealment:  **no** group assignments were not  disclosed to the investigators until the study was complete | Participants:  **no** the patients knew whether EPO was being  administered or not, but outcomes are not likely to be influenced by this lack of blinding  Personnel:  **no** all investigators, surgeons, anesthesiologists, perfusionists,  and intensive care unit physicians responsible for  the administration of blood products  were blinded with respect to group  assignment thereby creating a single-blinded study | **no**  no missing data (no loss to follow up).  In addition, an intention-to-treat analysis was performed. | **no** The authors do not provide any information on the outcome measurement used for most of these complications  (e.g. focal neurologic damage, acute respiratory distress syndrome…).  It is unlikely that the measurement may have differed between both groups. In addition, the authors explicitly state that everyone involved in the evaluation of outcomes were blinded with respect to group assignment. --> ‘low’ in RoB 2.0 tool | **yes**  In the Methods section, the authors clearly specify that recording of the occurrence of common acute complications after heart surgery, post-discharge complications and morality rates was part of the study schedule. Therefore, this was probably included in the prespecified analysis plan.  It is unclear if selective reporting of a particular outcome measurement and/or particular analysis has occurred.  --> ‘some concerns’ in RoB 2.0 tool | This study reports a single-centre experience so, by definition, it gives no guarantee for results to be generalizable to other centres |
| Yoo, 2011 | Lack of randomization:  **no** computer-generated randomization was used and was performed by a ward physician not involved in the current trial  Lack of allocation concealment:  **no** a ward physician not involved in the current trial performed the assignment | Participants:  **unclear*(2)** no information provided  Personnel:  **no**  medications were prepared and administered  by a ward physician recognizing the patient’s group but not involved in the current study, whereas the surgeon and anesthesiologist involved in the study and patient management  were blinded to the patients’ groups until the end of the  study | **no** no missing data | **yes**  - Participant-reported outcomes (headache, nausea, diarrhea, vomiting):  It is unclear if these adverse events were collected in a systematic (active surveillance) or non-systematic way.  It is unclear if the measurement may have differed between both groups.  As it is unclear if the participants were aware of the intervention received, it is unclear if the results of the participant-reported outcomes may have been biased. --> ‘high’ in RoB 2.0 tool  - Other outcomes (tachycardia, thromboembolism, acute kidney injury, atrial fibrillation, duration of ventilator care etc.): The authors do not provide any information on the outcome measurement used for most of these outcomes (only for acute kidney injury: through serum creatinine levels).  It is unclear if the measurement may have differed between both groups. It is unclear if the assessment of these outcomes is potentially influenced by knowledge of the intervention received.  --> ‘high’ in RoB 2.0 tool | **yes**  No information on protocol registration for this trial.  In the Methods section of the paper, the occurrence of postoperative complications is mentioned as a secondary outcome. However, further on in this section, they also mention that this study was not designed to validate the difference in outcome variables such as acute kidney injury, atrial fibrillation, duration of ventilator care, intensive care unit, and hospital stay, and surgical mortality, but that it could be informative to provide postoperative outcome variables to demonstrate that EPO therapy did not appear to be associated with adverse outcomes.  Nevertheless, these outcomes were probably included in the prespecified analysis plan.  It is unclear if selective reporting of a particular outcome measurement and/or particular analysis has occurred.  --> ‘some concerns’ in RoB 2.0 tool |  |

Study limitations observational studies

| **Author, Year** | **Inappropriate eligibility criteria** | **Inappropriate methods for exposure variables** | **Inadequate measurement of the outcomes** | **Inadequate selection of the reported result** | **Not controlled for confounding** | **Incomplete or inadequate follow-up** | **Other limitations** |
| --- | --- | --- | --- | --- | --- | --- | --- |
| Cladellas, 2012 | **yes** The intervention cohort consisted of statistically significantly more diabetic individuals. This may have confounded the effects of the treatment on the occurrence of adverse events. | **no** no indication: surgical techniques, anesthetic care and transfusion triggers were the same in both periods | **yes**  The authors refer to a previous paper for definitions on the major postoperative adverse cardiovascular events. However, this paper does not provide any information on the outcome measurements used.  It is unclear if measurements may have differed between both groups.  It is highly unlikely that the outcome assessors were blinded to the intervention status. Assessment of the outcomes will have probably been influenced by the knowledge of the received intervention, as this assessment probably required some clinical judgement. --> ‘high’ in RoB 2.0 tool | **yes**  In the Methods section, the authors report that they looked at the composite and separate measures for major postoperative adverse cardiovascular events. Therefore, this was probably included in the prespecified analysis plan.  It is unclear if selective reporting of a particular outcome measurement and/or particular analysis and/or a particular subgroup has occurred.  --> ‘some concerns’ in RoB 2.0 tool | **yes** Although the authors did not find any significant  changes in surgical technique or anesthetic care between both  periods, this long period analyzed could lead to a bias owing to some unrecognized confounders.  Although the authors performed multivariable regression, this was only the case for in-hospital mortality and the composite measure of major adverse cardiac events, so not for the individual measures of the adverse cardiac events. | **no** no indication | **no** |
| Delasotta, 2012 (A) | **unclear*(2)** insufficient demographic information provided | **no** no indication | **yes**  The authors report that ultrasound was used to diagnose an uncomplicated DVT. However, it is unclear if the study investigators ruled out any preexisting DVTs.  It is unclear if patients were systematically assessed for the occurrence of adverse events.  It is unclear if the measurement outcome may have differed between both groups. It is unclear if the assessor that interpreted the ultrasound was blinded to the intervention assignment, which may have substantially influenced his judgement.  --> ‘high’ in RoB 2.0 tool | **yes**  Adverse events are not mentioned as outcomes of interest in the Methods section of the paper. This suggests that the outcomes were probably not included in the prespecified analysis plan, but added to the results report post-hoc.  It is unclear if selective outcome reporting of a particular outcome measurement and/or of a particular analysis and/or of a particular subgroup has occurred.  --> ‘some concerns’ in RoB 2.0 tool | **yes** although the authors patient-matched on age, gender, BMI, ASA score and procedure, other factors may have confounded the results (e.g. estimated blood loss, preoperative administration of vitamins).  Simple student’s t-test and chi-square test were used for comparing the continuous and categorical variables. | **no** no indication | **no**  Retrospective data collection |
| Delasotta, 2012 (B) | **unclear*(2)** insufficient demographic information provided | **no** no indication | **yes**  The authors do not provide any information on the outcome measurements used. In addition, it is unclear if the study investigators ruled out any preexisting DVTs or emboli.  It is unclear if patients were systematically assessed for the occurrence of adverse events.  It is unclear if the measurement outcomes may have differed between both groups. It is unclear if the outcome assessors were blinded to the intervention assignment, which may have substantially influenced the results.  --> ‘high’ in RoB 2.0 tool | **yes**  Adverse events are not mentioned as outcomes of interest in the Methods section of the paper. This suggests that the outcomes were probably not included in the prespecified analysis plan, but added to the results report post-hoc.  It is unclear if selective outcome reporting of a particular outcome measurement and/or of a particular analysis and/or of a particular subgroup has occurred.  --> ‘some concerns’ in RoB 2.0 tool | **yes** although the authors patient-matched on age, gender, BMI, ASA score and procedure, other factors may have confounded the results (e.g. preoperative administration of folic acid, iron, vitamin B12 and multivitamins). Simple student’s t-test and chi-square test were used for comparing the continuous and categorical variables. | **no** no indication | The authors included a consecutive series of case patients |
| Ellermann, 2018 | **unclear** insufficient demographic information provided | **no** no indication | **yes**  The authors have systematically telephoned patients after 30 days to interview them about possible adverse events. A second telephone interview was performed 90 days after the visit, and a third 1 year after the visit. Telephone interviews were standardized. Measurements did not differ between both groups.  For some outcomes (e.g. dyspepsia, nausea, abdominal pain), the results may have been biased since the participants were the outcome assessors, and they were aware of the intervention received. For other outcomes, it is unclear if outcome assessors were blinded. --> ‘high’ in RoB 2.0 tool | **yes**  Adverse events were clearly an outcome of interest for this study, as systematic telephone interviewing was part of the study protocol. Therefore, this was probably included in the prespecified analysis plan.  It is unclear if selective reporting of a particular outcome measurement and/or particular analysis and/or a particular subgroup has occurred.  --> ‘some concerns’ in RoB 2.0 tool | **yes** although multivariable analyses were performed adjusting for the possible confounders age, gender, and time,  there may still be residual confounding (e.g. medication taken, previous surgeries, bleeding disorder) | **no** no indication | **no** |
| Evans, 2021 | **yes** The intervention cohort consisted of statistically significantly more male individuals. This may have confounded the effects of the treatment on 30-day mortality. | **no** no indication | **no** It is highly unlikely that the method of measuring procedural (in-hospital or within 30 days after surgery) was inappropriate.  It is unlikely that the measurement outcomes may have differed between both groups. It is unclear if outcome assessors were blinded to intervention status. However, it is highly unlikely that results are influenced by knowledge of the intervention received.  --> ‘low’ in RoB 2.0 tool | **no**  Mortality is clearly mentioned as an outcome of interest for this study in the Methods section. Therefore, this was probably included in the prespecified analysis plan.  It is unlikely that selective reporting of a particular outcome measurement and/or particular analysis and/or a particular subgroup has occurred. --> ‘low’ in RoB 2.0 tool | **yes** Although insufficient information is available on between-group differences concerning potentially relevant confounding variables, it is likely that analyses should have been corrected for multiple confounding variables (e.g. age, gender, comorbidities) | **no** no indication | **no**  Retrospective data collection  Analyses were  exploratory in nature and not corrected for multiple testing  Potential conflict of interest: The supplier of the IV iron (Pharmacosmos) has granted honoraria and subsidies, as well as fees for undertaking statistical analysis to several study authors. |
| Kam, 2020 | **yes** The intervention cohort consisted of statistically significantly more smokers. This may have confounded the effects of the treatment on the occurrence of adverse events. In addition, it remains unclear if similar inclusion/exclusion criteria were applied to the historic cohort (compared to the intervention cohort). Via personal email communication, it was indicated that “the historical cohort also consisted of patients with iron-deficiency anemia but the data collection was not complete - meaning, some patients only had low serum iron, but at that point the lab data did not include ferritin / TIBC, because they were not requested specifically.” | **yes** While the  technical aspects of the operations were not changed, learning  curves of individual surgeons differed or improved and  may have affected the control of hemostasis and use of transfusions in the intervention cohort compared to the historic control cohort. This may have also resulted in changes in adverse events. Moreover, the period in which IV iron therapy was introduced coincided with a period of shortage of blood products in Hong Kong. This may have distorted the data on the number of transfusions and subsequently the number of adverse events. In addition, via personal email communication, it was indicated that the transfusion policy in the historic cohort was not standardized (compared to the intervention cohort). | **yes**  From the personal communication with the authors, it is clear that patients in the IV iron group were systematically assessed by a study nurse for the occurrence of adverse events during and after infusion.  It is unlikely that the measurement of complications was done inappropriately, but it is unclear if this measurement may have differed between both groups. It is unclear if the outcome assessors were blinded and to which extent this may have influenced the outcome assessment. --> ‘high’ in RoB 2.0 tool | **yes**  Adverse events are not mentioned as outcomes of interest in the Methods section of the paper. This suggests that the outcomes were probably not included in the prespecified analysis plan, but added to the results report post-hoc.  It is unclear if selective outcome reporting of a particular outcome measurement and/or of a particular analysis and/or of a particular subgroup has occurred.  --> ‘some concerns’ in RoB 2.0 tool | **yes** although the authors have used propensity score matching to create groups that are comparable, the data on the occurrence of adverse events may have been confounded by other variables that have not been taken into account | **no** no indication | **no**  Retrospective data collection (historic control group) |
| Klein, 2020 | **yes** Patients treated with IV iron were more likely to have chronic kidney disease | **yes** Information obtained from the authors (personal communication with Klein): Across the 11 NHS hospitals, all patients received tranexamic acid as standard in all institutions and some patients received cell salvage according to institutional protocols. However,  patients have received different thromboprophylaxis regimens at different institutions. This may have impacted adverse events data. | **yes** From the personal communication with the authors, it is clear that patients in the IV iron group were systematically assessed for the occurrence of adverse events during and after infusion.  It is highly unlikely that the measurement of readmission rates and mortality was done inappropriately, but it is unclear if this measurement may have differed between both groups. For the outcome of nausea, the results may have been biased since the participants were the outcome assessors, and they were aware of the intervention received. For readmission and mortality rates, it is unclear if outcome assessors were blinded, although it is highly unlikely that assessment was influenced by this potential lack of blinding. --> 'some concerns' (readmission and mortality) and ‘high’ (nausea) in RoB 2.0 tool | **yes** Complications during hospital stay were mentioned as secondary outcomes of interest in published protocol (Chau 2017). However, this also mentioned renal function as an outcome of interest. During personal communication with the authors, it became clear that this outcome was not measured. This raises concerns about potential selective outcome reporting of a particular outcome measurement and/or of a particular analysis. --> 'some concerns' in RoB 2.0 tool | **yes** analyses were not controlled for confounding. The authors themselves state that: "There were significant differences  in baseline characteristics between the groups that could plausibly lead to changes in the treatment effect and measured outcomes". | **no** no indication | **no**  The study was designed to detect an increase  in Hb concentration and therefore underpowered to  detect outcome changes.  Conflict of interest: although the authors declared no conflict of interest, Pharmacosmos provided funding for the study |
| Laso-Morales, 2017 | **unclear**  Although the authors have investigated if there were any baseline differences in comorbidities between both groups, they only reported a p-value for the comparison of all comorbidities combined, not for the separate comorbidities (e.g. diabetes, hypertension). | **no** no indication | **yes**  The authors do not provide any information on the outcome measurements used, except for infection (diagnosed by a senior member + confirmed by laboratory/radiologic/microbiological evidence). In addition, it is unclear if the study investigators ruled out any preexisting DVTs or emboli.  It is unclear if patients were systematically assessed for the occurrence of adverse events.  It is unclear if the measurement outcomes may have differed between both groups. It is unclear if the outcome assessors were blinded to the intervention assignment (although the attending surgeon who assessed the DVT was not necessarily aware of the treatment given by the anaesthesiologist), which may have substantially influenced the results.  --> ‘high’ in RoB 2.0 tool | **yes**  It is unclear if the reported results are consistent with an a priori analysis plan.  It is unclear if selective reporting of a particular outcome measurement and/or particular analysis and/or a particular subgroup has occurred.  --> ‘some concerns’ in RoB 2.0 tool | **yes** although multivariable analyses were performed adjusting for the possible confounders gender, BMI, surgical wound hematoma, paralytic ileus, baseline anemia, IVI administration and ASA scores,  not all possible confounders were  analyzed (e.g. estimated blood loss) | **no** no indication | **no**  This study may not be powered  to detect significant differences in postoperative complications |
| Nandhra, 2020 | **yes**  Although the authors have investigated if there were any baseline differences in comorbidities between both groups, they only reported a p-value for the comparison of anaemic vs non-anaemic and anaemic non-treated vs non-anaemic. Nevertheless, they report that the patients receiving IV iron therapy were at higher surgical risk, indicating an apparent selection bias for inclusion in the trial. | **yes** No direct information provided in the manuscript or via the authors. However, since this study population is part of the CAVIAR study population, the information from the Klein 2020 study is probably applicable here. | **yes**  The authors do not provide information on the outcome measurements used.  It is unclear if patients were systematically assessed for the occurrence of adverse events.  It is unclear if the measurement outcomes may have differed between both groups. It is unclear if outcome assessors were blinded to the intervention assignment, although it is unlikely that the outcomes (readmission, mortality) would be influenced by a lack of blinding.  --> ‘some concerns’ in RoB 2.0 tool | **yes**  Although adverse events are not mentioned as outcomes of interest in the Methods section of the paper, complications during hospital stay were mentioned as secondary outcomes of interest in published protocol (Chau 2017). This suggests that the outcomes were probably included in the prespecified analysis plan.  Raw data on mortality and readmission were obtained through personal communication with the study author.  It is unclear if selective reporting of a particular outcome measurement and/or particular analysis has occurred.  --> ‘some concerns’ in RoB 2.0 tool | **yes** Although insufficient information is available on between-group differences concerning potentially relevant confounding variables, it is likely that analyses should have been corrected for a number of variables | **no** no indication | **no**  Conflict of interest: although the authors declared no conflict of interest, Pharmacosmos provided funding for the study |
| Pinilla-Gracia, 2020 | **yes** the subjects in the ESA + IV iron group were significantly younger than the subjects in the control group | **no** No indication. Perioperative management (regional anesthesia, antibiotic prophylaxis, choice of prosthetic implants, transfusion protocol, etc) was standardized between the intervention and control cohort. | **yes**  The authors do not provide information on the outcome measurements used.  It is unclear if patients were systematically assessed for the occurrence of adverse events.  It is unclear if the measurement outcomes may have differed between both groups. It is unclear if outcome assessors were blinded to the intervention assignment, which could have influenced the assessment of the outcomes.  --> ‘high’ in RoB 2.0 tool | **yes** In the Methods section of the paper, the authors mention that postoperative thromboembolic and infectious complications and medical or prosthetic complications, as well as 6-month mortality were outcomes of interest.  Therefore, this was probably included in the prespecified analysis plan. It is unclear if selective reporting of a particular outcome measurement and/or particular analysis and/or a particular subgroup has occurred.  --> ‘some concerns’ in RoB 2.0 tool | **yes** analyses were not controlled for confounding | **no** no indication | **no**  Conflict of interest:  Manuel Muñoz has received honoraria for lectures and/or consultancies  from Vifor Pharma, the supplier of IV iron |
| Quinn, 2017 | **unclear** insufficient demographic information provided | **no** no indication | **yes**  The authors do not provide information on the outcome measurements used (“Data was  collected on patients' pre- and postoperative Hb levels, iron-deficiency  rates, methods and success of iron-deficiency correction,  perioperative transfusion rates, postoperative complications  and length of postoperative stay”).  It is unclear if patients were systematically assessed for the occurrence of adverse events.  It is unclear if the measurement outcomes may have differed between both groups. It is unlikely that outcome assessors were blinded to the intervention assignment, which may have substantially influenced the results.  --> ‘high’ in RoB 2.0 tool | **yes**  Although postoperative complications were reported to be of interest in the Methods section, no information is provided in the Results section. Data on complications were obtained through personal communication with the study author.  It is unclear if selective reporting of a particular outcome measurement and/or particular analysis and/or a particular subgroup has occurred.  --> ‘some concerns’ in RoB 2.0 tool | **yes** Although insufficient information is available on between-group differences concerning potentially relevant confounding variables, it is likely that analyses should have been corrected for multiple confounding variables (e.g. age, gender, comorbidities) | **no** no indication | **no** |
| Rineau, 2017 | **yes** there were significantly more women in the EPO<13 group compared to the EPO<15 group | **no** no indication | **yes**  The authors do not provide information on the outcome measurements used (“Secondary outcomes were the  number of EPO injections per patient, perioperative Hb levels, length of hospital stay, and complications”).  It is unclear if patients were systematically assessed for the occurrence of adverse events.  It is unclear if the measurement outcomes may have differed between both groups. It is unlikely that outcome assessors were blinded to the intervention assignment, which may have substantially influenced the results.  --> ‘high’ in RoB 2.0 tool | **yes**  In the paper, only a composite measure of the different perioperative complications is reported. Data on individual complications were obtained via personal communication with the study author.  It is unclear if selective reporting of a particular outcome measurement and/or particular analysis and/or a particular subgroup has occurred.  --> ‘some concerns’ in RoB 2.0 tool | **yes** the data on the occurrence of perioperative complications may have been confounded by several variables that have not been taken into account | **no** no indication | **no** |
| Triphaus, 2019 | **unclear** insufficient demographic information provided | **no** No indication. All patients received standard perioperative care. | **yes**  It is obvious from the Methods section and the personal communication with the authors that patients in the IV iron group were systematically assessed by a study nurse for the occurrence of adverse events during and after infusion. However, it is unclear if an appropriate assessment was also present in the control group.  Hence, it is unclear if the measurement outcomes may have differed between both groups. For some outcomes (e.g. headache, gastrointestinal symptoms), the results may have been biased since the participants were the outcome assessors, and they were aware of the intervention received. For other outcomes, it is unclear if outcome assessors were blinded. --> ‘high’ in RoB 2.0 tool | **yes** In the Methods section, the authors report that IV iron-related adverse events and in-hospital mortality were secondary outcomes of interest.  Therefore, this was probably included in the prespecified analysis plan.  Data on the IV iron-related adverse events studies were obtained via personal communication with the study authors.  In the clinical trial registration (NCT02147795), a composite measure for in-hospital myocardial infarction, stroke, acute renal failure, death, pneumonia and sepsis until discharge from hospital was mentioned. However, this is not reported in the paper. This raises some concern about selective outcome reporting.  --> ‘some concerns’ in RoB 2.0 tool | **yes** the data on the occurrence of adverse events may have been confounded by several variables that have not been taken into account | **no** no indication | **no**  The authors state that the dosing regimen of 500 or 1000 mg IV iron might have been a bit  too low to substitute severe iron-deficiency in some patients and therefore may have weakened the study results. This may also have implications on the occurrence/detection rate of adverse events.  Conflict of interest:  Disclosure: P.M. and K.Z. received grants from B. Braun Melsungen, CSL  Behring, Fresenius Kabi, and Vifor Pharma for the implementation of Frankfurt‘  s Patient Blood Management program and honoraria for scientific lectures  from B. Braun Melsungen, Vifor Pharma, Ferring, CSL Behring, and Pharmacosmos. |
| Wilson, 2018 (A) | **yes** The usual care cohort consisted of statistically significantly more men, more people with comorbidities as well as people with higher Hb levels at diagnosis. This may have confounded the data on the occurrence of adverse events. | **no** no indication | **yes** The authors report that data on postoperative complications were collected by the Dutch Surgical Colorectal Audit. However, they do not provide information on the outcome measurements used originally.  It is unclear if patients were systematically assessed for the occurrence of adverse events.  It is unclear if the measurement outcomes may have differed between both groups. It is highly unlikely that outcome assessors were blinded to the intervention assignment, which may have substantially influenced the results.  --> ‘high’ in RoB 2.0 tool | **yes**  In the Methods section, the authors report that complications were secondary outcomes of interest. Therefore, this was probably included in the prespecified analysis plan.  It is unclear if selective reporting of a particular outcome measurement and/or particular analysis and/or a particular subgroup has occurred. However, the fact that they only report on a composite measure raises some concerns.  --> ‘some concerns’ in RoB 2.0 tool | **yes** The significant differences  between the IV iron and UC group (e.g. baseline Hb levels) could, despite correction  in the multivariable regressions analyses, potentially  indicate selection bias and have significant impact on the outcome. Moreover, iron status was not consistently monitored  in each patient, and blood transfusion strategies have changed over time. | **no** no indication | **no**  This study included a series of consecutive patients.  Only short-term effects of IV iron therapy were studied. |
| Wilson, 2018 (B) | **yes** Both groups showed statistically significant differences in tumour location,  Hb level at diagnosis, treatment approach and resection type. This may have confounded the data on survival. | **no** no indication | **no**  It is highly unlikely that the method of measuring overall and disease-free survival was inappropriate.  It is unlikely that the measurement outcomes may have differed between both groups. It is unclear if outcome assessors were blinded to intervention status. However, it is highly unlikely that results are influenced by knowledge of the intervention received.  --> ‘low’ in RoB 2.0 tool | **no**  Survival and disease-free survival are clearly mentioned as the primary outcomes for this study. In addition, in the Methods section, the authors provide an extensive explanation on how this was calculated. Therefore, the results were probably analysed in accordance with a pre-specified analysis plan.  It is unlikely that selective reporting of a particular outcome measurement and/or particular analysis and/or a particular subgroup has occurred. --> ‘low’ in RoB 2.0 tool | **yes** although survival analyses were performed on propensity matched groups, possible confounders may be hidden and therefore selection bias cannot be completely ruled out | **no** no indication | **no**  Surgery and tumour eradication may neutralise the possible detrimental effects of intravenous  iron on tumour growth and long-term prognosis |
| Ye, 2017 | **yes**  The control group (no ESA, no iron) had significantly higher baseline Hb levels. This may have confounded the data on the occurrence of adverse events. | **no** no indication | **yes**  The authors do not provide information on the outcome measurements used (“In accordance with our study, the following data were collected: […]; (g) side effects observed (headache, mild fever, myalgia, allergy, thrombosis, and gastrointestinal reaction); (h) postoperative complications; […]  ”).  It is unclear if patients were systematically assessed for the occurrence of adverse events.  It is unclear if the measurement outcomes may have differed between both groups. For some outcomes (e.g.headache, myalgia), the results may have been biased since the participants were the outcome assessors, and they were aware of the intervention received. For other outcomes, it is unclear if outcome assessors were blinded. --> ‘high’ in RoB 2.0 tool | **yes**  In the Methods section, the authors report that complications were outcomes of interest. Therefore, this was probably included in the prespecified analysis plan.  It is unclear if selective reporting of a particular outcome measurement and/or particular analysis and/or a particular subgroup has occurred  --> ‘some concerns’ in RoB 2.0 tool | **yes** not controlled for confounding | **no** no indication | **no**  No conflicts of interest |

### Supplementary Table 8. Synthesis of findings.

| **Outcome** | **Comparison** | **Effect Size** | **#studies, # participants** | **Reference** |
| --- | --- | --- | --- | --- |
| **IRON ADMINISTRATION ONLY** | | | | |
| **COMPARISON 1: IV IRON VS ORAL IRON** | | | | |
| **1.1 Gastrointestinal (Figure 1.1)** | | | | |
| Gastrointestinal symptoms including diarrhoea and constipation | IV iron vs oral iron | Not statistically significant:  0/20 vs 3/20 §  RR: 0.14, 95%CI [0.01;2.60] ¥ (p=0.19)* | 1, 20 vs 20 | Padmanabhan, 2019 |
| Dyspepsia |  | *Meta-analysis:* Not statistically significant:  0/85 vs 2/87 §  RR: 0.33, 95%CI [0.03;3.08] ¥  (p=0.33)* | 2, 85 vs 87 | Keeler, 2017;  Kim, 2009 |
| Nausea |  | Not statistically significant:  0/30 vs 1/26 §  RR: 0.29, 95%CI [0.01;6.83] ¥ (p=0.44)* | 1, 30 vs 26 | Kim, 2009 |
| Obstipation |  | Not statistically significant:  0/55 vs 1/61 §  RR: 0.37, 95%CI [0.02;8.88] ¥ (p=0.54)* | 1, 55 vs 61 | Keeler, 2017 |
| *Figure 1.1: IV iron vs oral iron – Gastrointestinal adverse events*  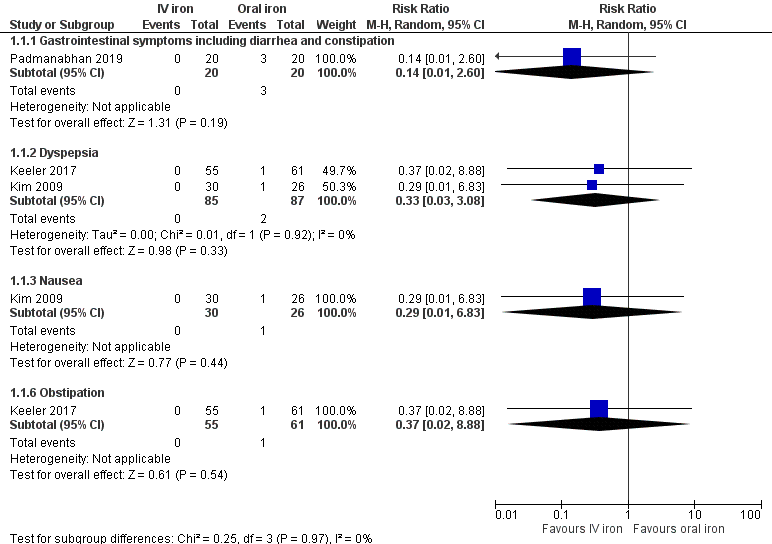 | | | | |

| **1.2 Mucocutaneous (Figure 1.2)** | | | | |
| --- | --- | --- | --- | --- |
| Rash | IV iron vs oral iron | Not statistically significant:  1/55 vs 0/61 §  RR: 3.32, 95%CI [0.14;79.88] ¥ (p=0.46)* | 1, 55 vs 61 | Keeler, 2017 |
| *Figure 1.2: IV iron vs oral iron – Mucocutaneous adverse events*  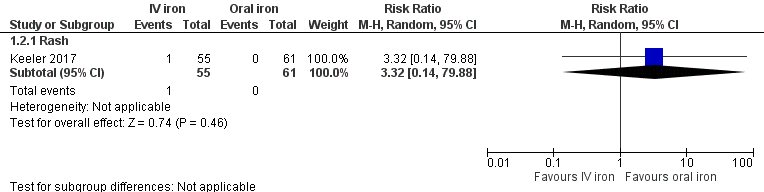 | | | | |
| **1.3 Neuro-psychosomatic (Figure 1.3)** | | | | |
| Post-infusion headache | IV iron vs oral iron | Not statistically significant:  3/55 vs 0/61 §  RR: 7.75, 95%CI [0.41;146.76] ¥ (p=0.17)* | 1, 55 vs 61 | Keeler, 2017 |
| Myalgia |  | Not statistically significant:  2/30 vs 0/26 §  RR: 4.35, 95%CI [0.22;86.79] ¥ (p=0.34)* | 1, 30 vs 26 | Kim, 2009 |
| Injection pain |  | Not statistically significant:  1/30 vs 0/26 §  RR: 2.61, 95%CI [0.11;61.51] ¥ (p=0.55)* |  |  |
| *Figure 1.3: IV iron vs oral iron – Neuro-psychosomatic adverse events*  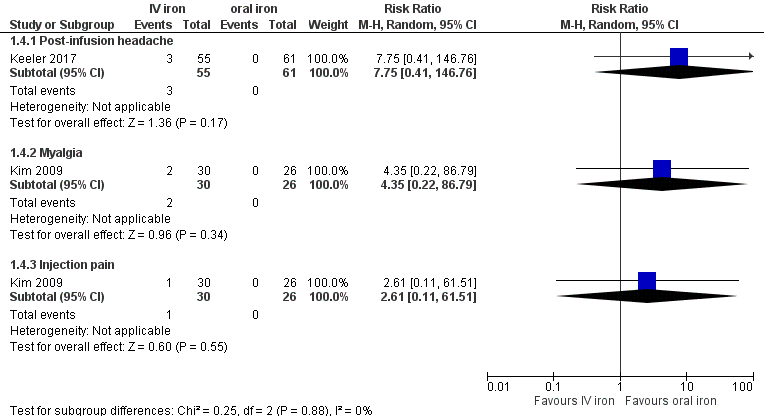 | | | | |

| **1.4 Neurological (Figure 1.4)** | | | | |
| --- | --- | --- | --- | --- |
| Postoperative ileus | IV iron vs oral iron | Not statistically significant: 1/8 vs 0/7 §  RR: 2.67, 95%CI [0.13;56.63] ¥ (p=0.53)* | 1, 8 vs 7 | Quinn, 2017 (cohort) |
| *Figure 1.4: IV iron vs oral iron – Neurological adverse events*  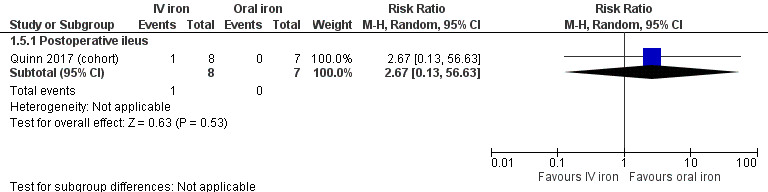 | | | | |
| **1.5 Wound healing (Figure 1.5)** | | | | |
| Wound dehiscence requiring readmission | IV iron vs oral iron | Not statistically significant: 1/50 vs 1/56 § RR: 1.12, 95%CI [0.07;17.44] ¥ (p=0.94)* | 1, 50 vs 56 | Dickson, 2020 |
| Delayed wound dehiscence requiring readmission |  | Not statistically significant:  0/8 vs 1/7 §  RR: 0.30, 95%CI [0.01;6.29] ¥ (p=0.44)* | 1, 8 vs 7 | Quinn, 2017 (cohort) |
| *Figure 1.5: IV iron vs oral iron – Adverse events related to wound healing*  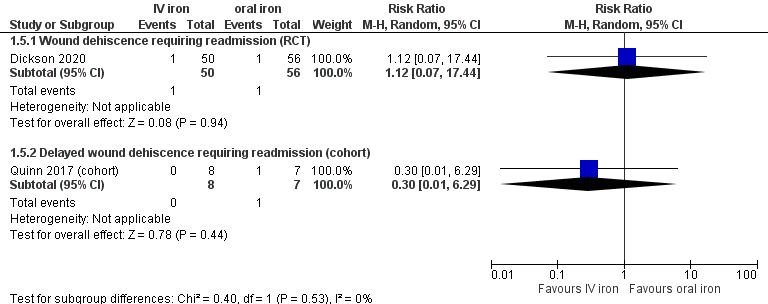 | | | | |

| **1.6 Infection** | | | | |
| --- | --- | --- | --- | --- |
| ***Findings presented in Figure 1.6*** | | | | |
| Postoperative infection | IV iron vs oral iron | *Meta-analysis:*  Not statistically significant:  25/75 vs 19/81 §  RR: 1.38, 95%CI [0.74;2.58] ¥ (p=0.31)* | 2, 75 vs 81 | Keeler, 2019; Padmanabhan, 2019 |
| ***Additional findings (not presented in Figure 1.6)*** | | | | |
| Grade of infective complication severity | IV iron vs oral iron | Not statistically significant:  £†  (p=0.083) | 1, 55 vs 61 § | Keeler, 2017 |
| Infective complication rate |  | Not statistically significant: £†  (p>0.05) | 1, 16 vs 17 § | Khalafallah, 2012 |
| *Figure 1.6: IV iron vs oral iron – Infectious adverse events*  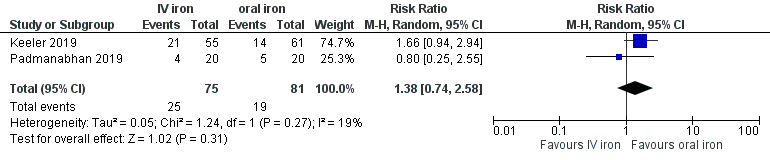 | | | | |
| **1.7 Bleeding (Figure 1.7)** | | | | |
| Upper gastrointestinal bleed | IV iron vs oral iron | Not statistically significant:  1/8 vs 0/7 §  RR: 2.67, 95%CI [0.13;56.63] ¥ (p=0.53)* | 1, 8 vs 7 | Quinn, 2017 (cohort) |
| *Figure 1.7: IV iron vs oral iron – Bleeding adverse events*  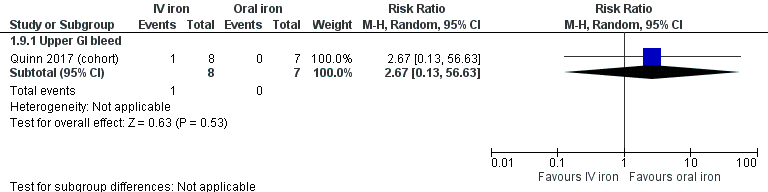 | | | | |

| **1.8 Cardiovascular (Figure 1.8)** | | | | |
| --- | --- | --- | --- | --- |
| Atrial fibrillation | IV iron vs oral iron | Not statistically significant: 10/20 vs 11/20 §  RR: 0.91, 95%CI [0.50;1.64] ¥ (p=0.75)* | 1, 20 vs 20 | Padmanabhan, 2019 |
| Atrial fibrillation requiring readmission |  | Not statistically significant: 0/50 vs 1/56 § RR: 0.37, 95%CI [0.02;8.94] ¥ (p=0.54)* | 1, 50 vs 56 | Dickson, 2020 |
| Cardiac failure |  | Not statistically significant: 1/8 vs 0/7 § RR: 2.67, 95%CI [0.13;56.63] ¥ (p=0.53)* | 1, 8 vs 7 | Quinn, 2017 (cohort) |
| *Figure 1.8: IV iron vs oral iron – Cardiovascular adverse events*  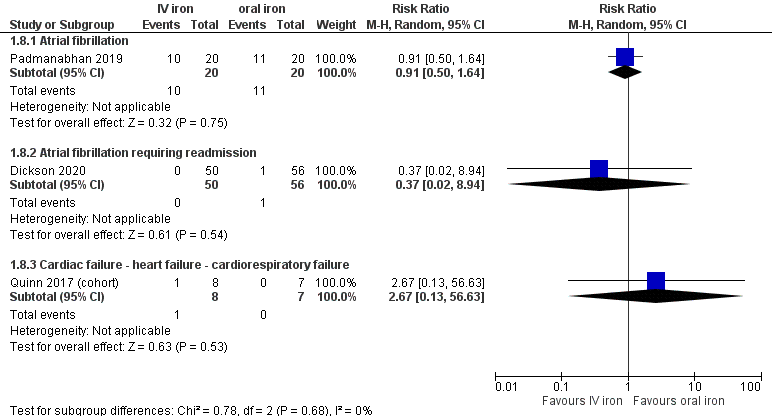 | | | | |
| **1.9 Renal (Figure 1.9)** | | | | |
| Need for renal replacement therapy | IV iron vs oral iron | Not statistically significant:  1/20 vs 1/20 §  RR: 1.00, 95%CI [0.07;14.90] ¥ (p=1.00)* | 1, 20 vs 20 | Padmanabhan, 2019 |
| *Figure 1.9: IV iron vs oral iron – Renal adverse events*  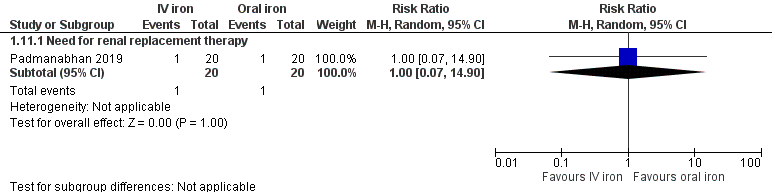 | | | | |

| **1.10 Anaemia-associated ischemic events (Figure 1.10)** | | | | |
| --- | --- | --- | --- | --- |
| Myocardial infarction | IV iron vs oral iron | Not statistically significant:  1/8 vs 0/7 §  RR: 2.67, 95%CI [0.13;56.63] ¥ (p=0.53)* | 1, 8 vs 7 | Quinn, 2017 (cohort) |
| *Figure 1.10: IV iron vs oral iron – Anaemia-associated ischemic events*  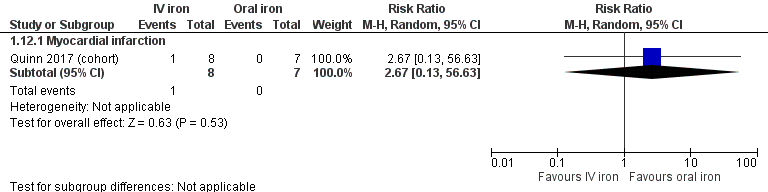 | | | | |
| **1.11 Thromboembolic events (Figure 1.11)** | | | | |
| Deep vein thrombosis requiring readmission | IV iron vs oral iron | Not statistically significant: 0/50 vs 1/56 § RR: 0.37, 95%CI [0.02;8.94] ¥ (p=0.54)* | 1, 50 vs 56 | Dickson, 2020 |
| Thrombotic and/or vascular events |  | 0/25 vs 0/27 § Effect size not estimable | 1, 25 vs 27 | Olijhoek, 2001 |
| *Figure 1.11: IV iron vs oral iron – Thromboembolic events*  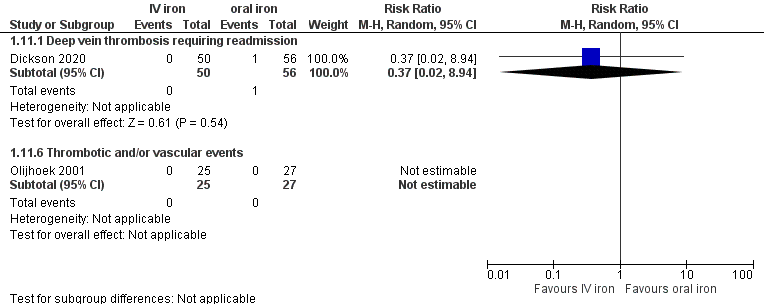 | | | | |

| **1.12 Mortality (Figure 1.12)** | | | | |
| --- | --- | --- | --- | --- |
| Mortality | IV iron vs oral iron | *Meta-analysis:* Not statistically significant:  6/130 vs 4/134 §  RR: 1.54, 95%CI [0.48;4.98] ¥ (p=0.47)* | 4, 130 vs 134 | Keeler, 2017; Kim, 2009; Olijhoek, 2001; Padmanabhan, 2019 |
| *Figure 1.12: IV iron vs oral iron – Mortality*  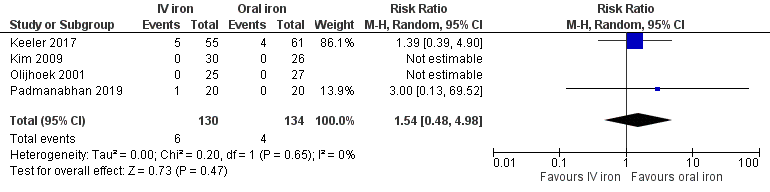 | | | | |
| **1.13 Survival (Figure 1.13)** | | | | |
| 2-year overall survival | IV iron vs oral iron | Not statistically significant: 45/54 vs 48/56 § RR: 0.97, 95%CI [0.83;1.14]  (p=0.73)* | 1, 54 vs 56 | Dickson, 2020 |
| 3-year overall survival |  | Not statistically significant: 42/54 vs 45/56 § RR: 0.97, 95%CI [0.80;1.17]  (p=0.74)* |  |  |
| 4-year overall survival |  | Not statistically significant: 37/54 vs 43/56 § RR: 0.89, 95%CI [0.71;1.12] ¥ (p=0.33)* |  |  |
| 5-year overall survival |  | Not statistically significant: 29/54 vs 33/56 § RR: 0.91, 95%CI [0.65;1.27] ¥ (p=0.58)* |  |  |
| 2-year disease-free survival |  | Not statistically significant: 45/54 vs 48/56 § RR: 0.97, 95%CI [0.83;1.14]  (p=0.73)* |  |  |
| 3-year disease-free survival |  | Not statistically significant: 42/54 vs 45/56 § RR: 0.97, 95%CI [0.80;1.17]  (p=0.74)* |  |  |
| 4-year disease-free survival |  | Not statistically significant: 38/54 vs 43/56 § RR: 0.92, 95%CI [0.73;1.15] ¥ (p=0.45)* |  |  |
| 5-year disease-free survival |  | Not statistically significant: 31/54 vs 35/56 § RR: 0.92, 95%CI [0.68;1.25] ¥ (p=0.59)* |  |  |
| 2-year colorectal cancer-specific survival |  | Not statistically significant: 40/54 vs 41/56 § RR: 1.01, 95%CI [0.81;1.27] ¥ (p=0.92)* |  |  |
| 3-year colorectal cancer-specific survival |  | Not statistically significant: 37/54 vs 36/56 § RR: 1.07, 95%CI [0.82;1.39] ¥ (p=0.64)* |  |  |
| 4-year colorectal cancer-specific survival |  | Not statistically significant: 34/54 vs 35/56 § RR: 1.01, 95%CI [0.76;1.34] ¥ (p=0.96)* |  |  |
| 5-year colorectal cancer-specific survival |  | Not statistically significant: 28/54 vs 29/56 § RR: 1.00, 95%CI [0.70;1.44] ¥ (p=0.99)* |  |  |
| *Figure 1.13: IV iron vs oral iron – Survival*  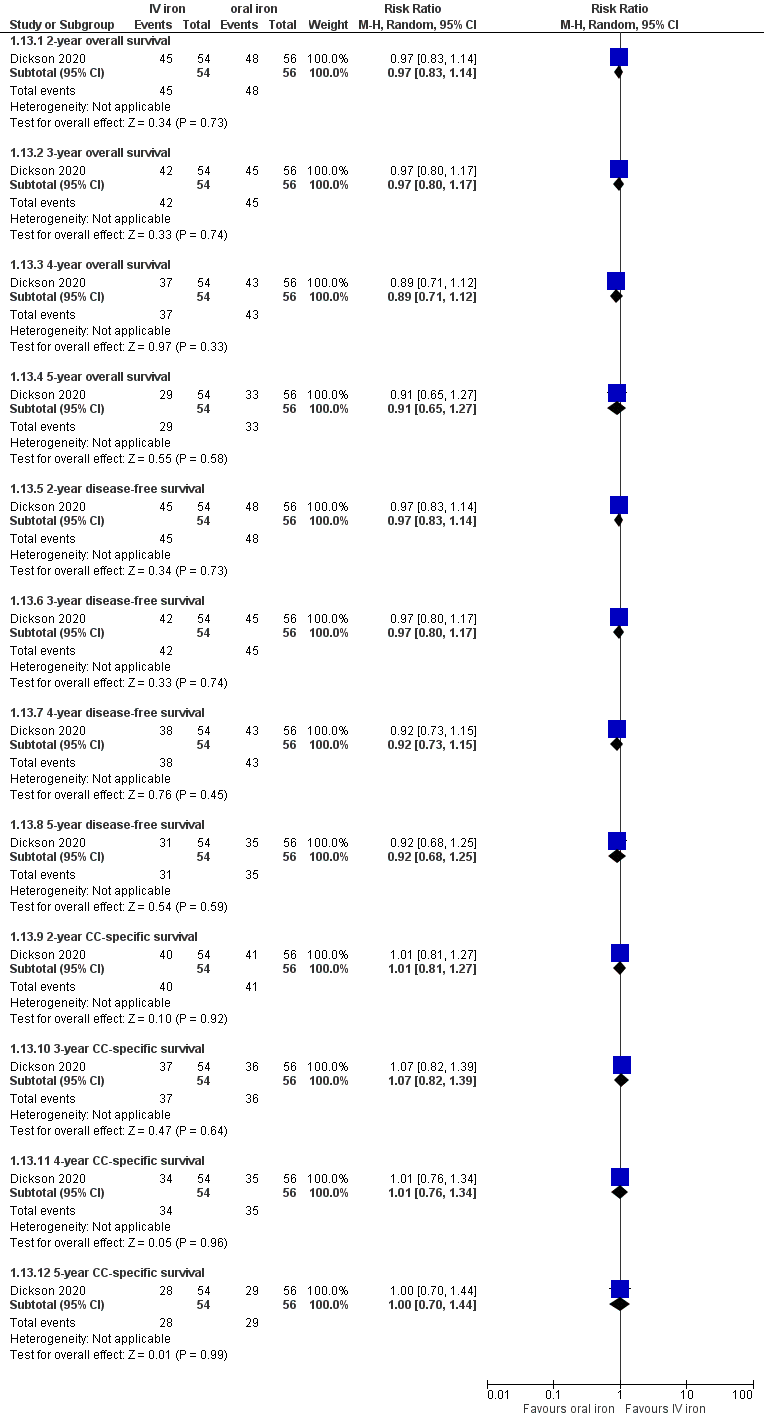 | | | | |
| **1.14 Other adverse events (Figure 1.14)** | | | | |
| Anastomotic leak requiring readmission | IV iron vs oral iron | Not statistically significant: 0/50 vs 1/56 § RR: 0.37, 95%CI [0.02;8.94] ¥ (p=0.54)* | 1, 50 vs 56 | Dickson, 2020 |
| Need for re-exploration |  | Not statistically significant: £†  (p>0.05) | 1, 20 vs 20 § | Padmanabhan, 2019 |
| Grade of complication severity from recruitment to outpatients |  | Not statistically significant:  £†  (p=0.995) | 1, 55 vs 61 § | Keeler, 2017 |
| Complication rate from recruitment to outpatients |  | Not statistically significant:  £†  (p=0.305) |  |  |
| *Figure 1.14: IV iron vs oral iron – Other adverse events*  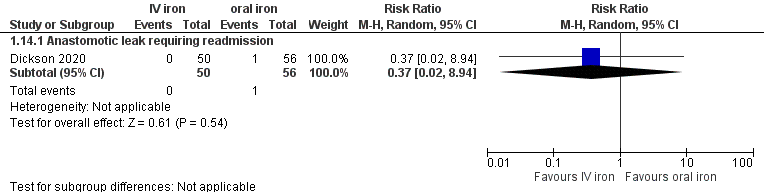 | | | | |

| **COMPARISON 2: IV IRON VS USUAL CARE/NO IRON** | | | | |
| --- | --- | --- | --- | --- |
| **2.1 Gastrointestinal (Figure 2.1)** | | | | |
| Nausea | IV iron  vs usual care/no iron | *Meta-analysis:* Not statistically significant: 7/166 vs 10/175 §  RR: 0.71, 95%CI [0.28;1.78] ¥ (p=0.46)* | 2, 166 vs 175 | Ellermann, 2018 (cohort);  Klein, 2020 (cohort) |
| Dyspepsia |  | Statistically significant: 2/102 vs 13/103 §  RR: 0.16, 95%CI [0.04;0.67] (p=0.01)* *in favour of IV iron* | 1, 102 vs 103 | Ellermann, 2018 (cohort) |
| Vomitus |  | Not statistically significant: 6/102 vs 7/103 §  RR: 0.87, 95%CI [0.30;2.49] ¥ (p=0.79)* |  |  |
| Abdominal pain |  | Not statistically significant: 4/102 vs 8/103 §  RR: 0.50, 95%CI [0.16;1.62] ¥ (p=0.25)* |  |  |
| Obstipation |  | Not statistically significant: 18/102 vs 20/103 §  RR: 0.91, 95%CI [0.51;1.61] ¥ (p=0.74)* |  |  |
| Diarrhoea |  | Not statistically significant:  7/102 vs 10/103 §  RR: 0.71, 95%CI [0.28;1.78] ¥ (p=0.46)* |  |  |
| Flatulence |  | Not statistically significant: 5/102 vs 3/103 §  RR: 1.68, 95%CI [0.41;6.86] ¥ (p=0.47)* |  |  |
| Gastrointestinal symptoms |  | 0/184 vs 0/50 § Effect size not estimable | 1, 184 vs 50 | Triphaus, 2019 (cohort) |
| Gastrointestinal reaction |  | 0/35 vs 0/32 §  Effect size not estimable | 1, 35 vs 32 | Ye, 2017 (cohort) |
| Incomplete intestinal obstruction |  | Not statistically significant: 2/35 vs 0/32 §  RR: 4.58, 95%CI [0.23;92.00] ¥ (p=0.32)* |  |  |

*Figure 2.1: IV iron vs usual care/no iron – Gastrointestinal adverse events*

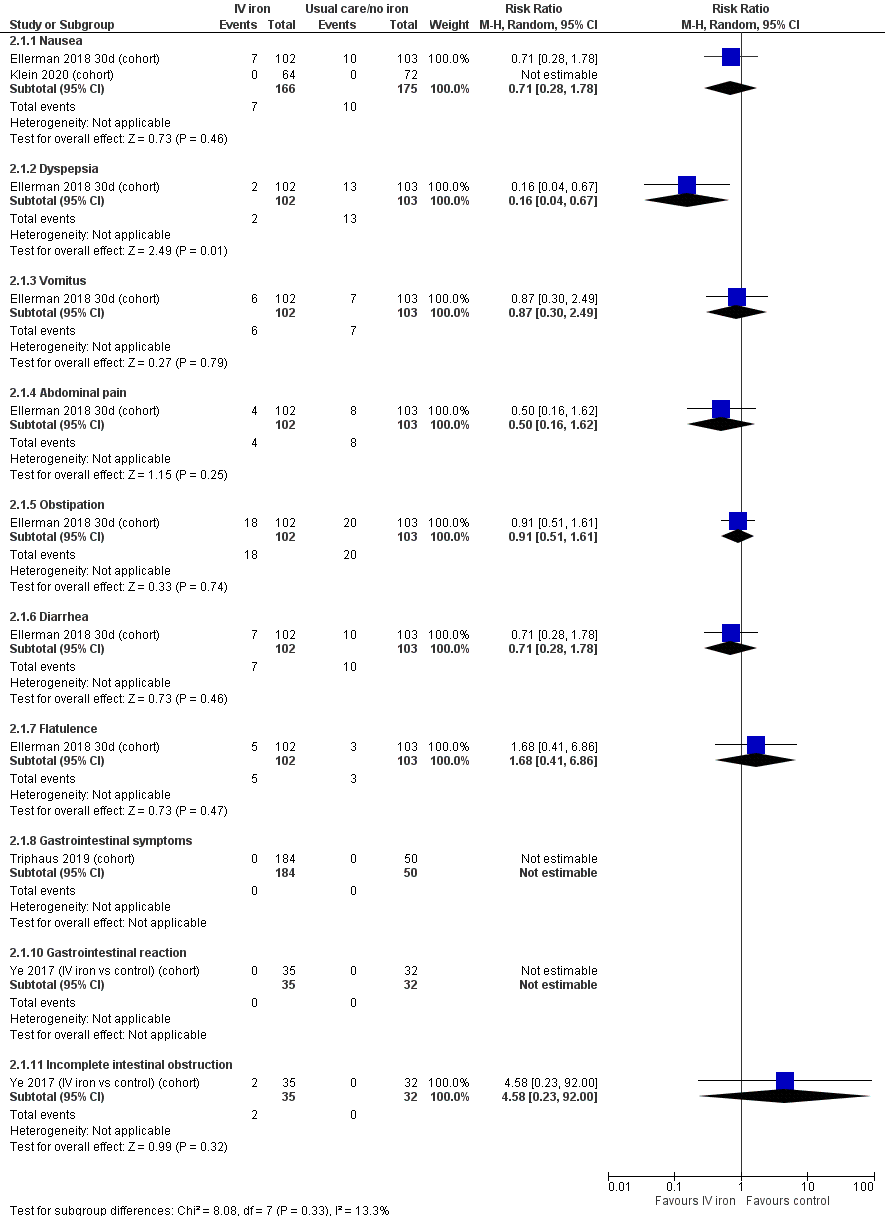

| **2.2 Mucocutaneous (Figure 2.2)** | | | | |
| --- | --- | --- | --- | --- |
| Pruritus | IV iron  vs usual care/no iron | Not statistically significant: 4/102 vs 3/103 §  RR: 1.35, 95%CI [0.31;5.87] ¥ (p=0.69)* | 1, 102 vs 103 | Ellermann, 2018 (cohort) |
| Urticaria |  | Not statistically significant: 5/102 vs 2/103 §  RR: 2.52, 95%CI [0.50;12.72] ¥ (p=0.26)* |  |  |
| Erythema |  | Not statistically significant: 3/102 vs 2/103 §  RR: 1.51, 95%CI [0.26;8.88] ¥ (p=0.65)* |  |  |
| Palor |  | 0/102 vs 0/103 § Effect size not estimable |  |  |
| Flush |  | Not statistically significant: 1/184 vs 0/50 § RR: 0.83, 95%CI [0.03;20.00] ¥ (p=0.91)* | 1, 184 vs 50 | Triphaus, 2019 (cohort) |
| Eczema |  | 0/184 vs 0/50 § Effect size not estimable |  |  |
| Rash |  | 0/38 vs 0/62 § Effect size not estimable | 1, 38 vs 62 | Kam, 2020 (cohort) |

*Figure 2.2: IV iron vs usual care/no iron – Mucocutaneous adverse events*

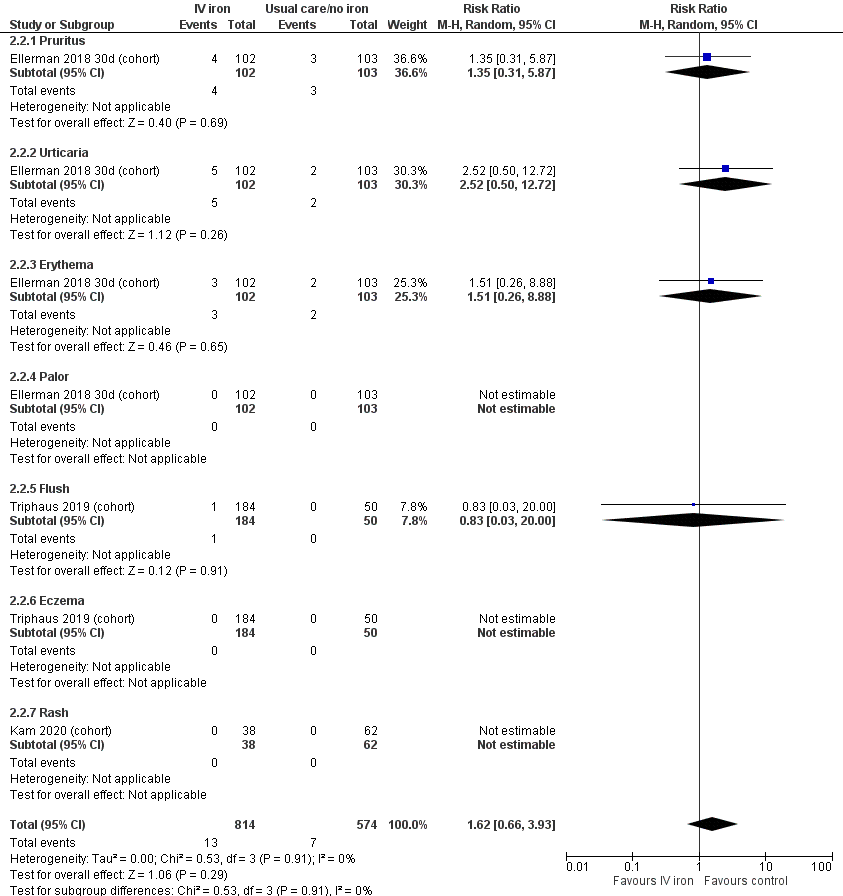

| **2.3 Autonomic (Figure 2.3)** | | | | |
| --- | --- | --- | --- | --- |
| Fever | IV iron  vs usual care/no iron | 0/257 vs 0/144 § Effect size not estimable | 3, 257 vs 144 | Kam, 2020 (cohort); Triphaus, 2019 (cohort); Ye, 2017 (cohort) |
| 30-day pyrexia rate |  | Not statistically significant: 10/102 vs 7/103 §  RR: 1.44, 95%CI [0.57;3.64] ¥ (p=0.44)* | 1, 102 vs 103 | Ellermann, 2018 (cohort) |
| 90-day pyrexia rate |  | Not statistically significant: 7/99 vs 4/101 § RR: 1.79, 95%CI [0.54;5.91] ¥ (p=0.34)* | 1, 99 vs 101 |  |
| 1-year pyrexia rate |  | Not statistically significant:  3/59 vs 3/59 §  RR: 1.00, 95%CI [0.21;4.75] ¥ (p=1.00)* | 1, 59 vs 59 |  |
| *Figure 2.3: IV iron vs usual care/no iron – Autonomic adverse events*  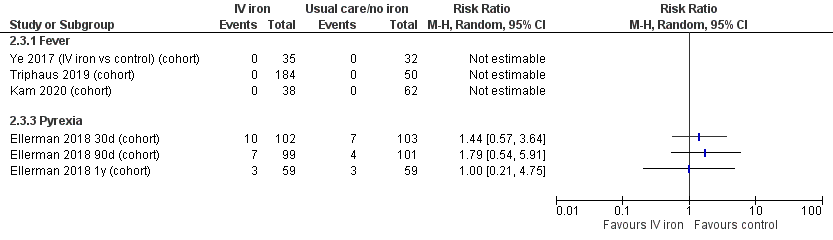 | | | | |

| **2.4 Neuro-psychosomatic (Figure 2.4)** | | | | |
| --- | --- | --- | --- | --- |
| Myalgia | IV iron  vs usual care/no iron | 0/35 vs 0/32 § Effect size not estimable | 1, 35 vs 32 | Ye, 2017 (cohort) |
| Headache |  | Not statistically significant: 1/40 vs 0/32 §  RR: 2.41, 95%CI [0.10;57.35] ¥ (p=0.59)* | 1, 40 vs 32 | Froessler, 2016 |
|  |  | *Meta-analysis:* Not statistically significant: 1/219 vs 0/82 §  RR: 0.83, 95%CI [0.03;20.00] ¥ (p=0.91)* | 2, 219 vs 82 | Ye, 2017 (cohort); Triphaus, 2019 (cohort) |
| Light-headedness |  | Not statistically significant: 1/40 vs 0/32 §  RR: 2.41, 95%CI [0.10;57.35] ¥ (p=0.59)* | 1, 40 vs 32 | Froessler, 2016 |
| Back pain |  | Not statistically significant: 1/40 vs 0/32 §  RR: 2.41, 95%CI [0.10;57.35] ¥ (p=0.59)* |  |  |
| *Figure 2.4: IV iron vs usual care/no iron – Neuro-psychosomatic adverse events*  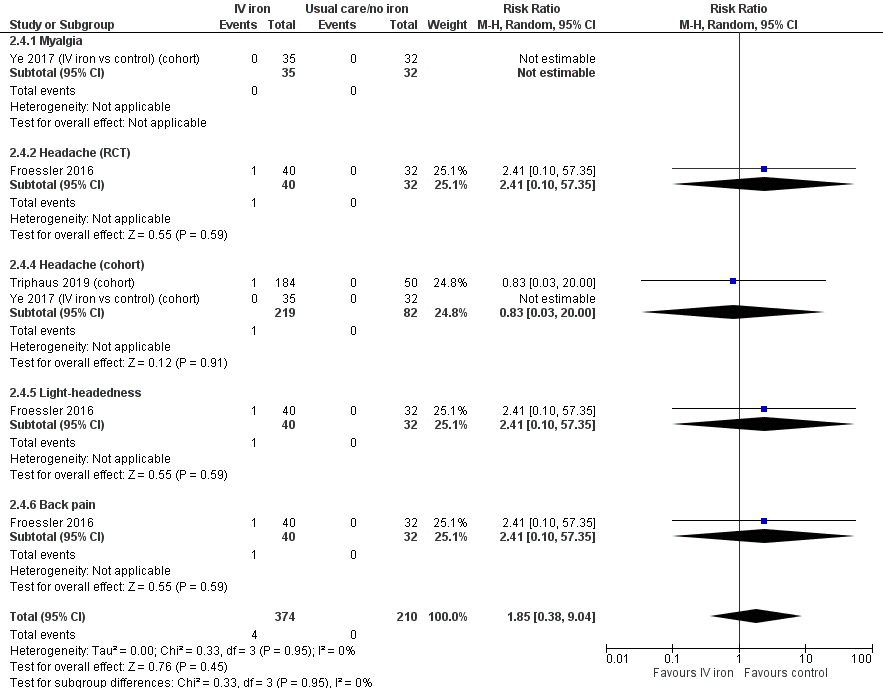 | | | | |

| **2.5 Neurological (Figure 2.5)** | | | | |
| --- | --- | --- | --- | --- |
| Ileus | IV iron  vs usual care/no iron | *Meta-analysis:* Not statistically significant: 23/270 vs 8/152 §  RR: 1.30, 95%CI [0.60;2.82] ¥ (p=0.51)* | 2, 270 vs 152 | Kam, 2020 (cohort); Laso-Morales, 2017 (cohort) |
| Cephalgia |  | Not statistically significant: 7/102 vs 8/103 §  RR: 0.88, 95%CI [0.33;2.35] ¥ (p=0.80)* | 1, 102 vs 103 | Ellermann, 2018 (cohort) |
| Vertigo |  | Not statistically significant: 8/102 vs 8/103 §  RR: 1.01, 95%CI [0.39;2.59] ¥ (p=0.98)* |  |  |
| Paresthesia |  | Not statistically significant: 3/102 vs 3/103 §  RR: 1.01, 95%CI [0.21;4.89] ¥ (p=0.99)* |  |  |
| Dysgeusia |  | Not statistically significant: 11/102 vs 12/103 §  RR: 0.93, 95%CI [0.43;2.00] ¥ (p=0.84)* |  |  |
| *Figure 2.5: IV iron vs usual care/no iron – Neurological adverse events*  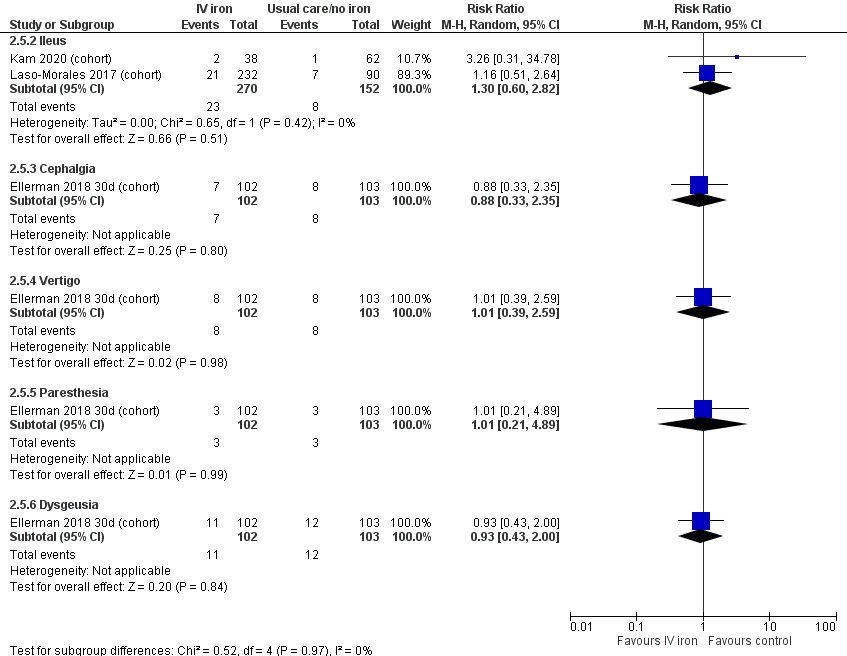 | | | | |

| **2.6 Wound healing (Figure 2.6)** | | | | |
| --- | --- | --- | --- | --- |
| Poor wound healing | IV iron  vs usual care/no iron | Not statistically significant: 1/35 vs 1/32 §  RR: 0.91, 95%CI [0.06;14.02] ¥ (p=0.95)* | 1, 35 vs 32 | Ye, 2017 (cohort) |
| Suture dehiscence |  | Not statistically significant:  19/232 vs 11/90 § RR: 0.67, 95%CI [0.33;1.35] ¥ (p=0.26)* | 1, 232 vs 90 | Laso-Morales, 2017 (cohort) |
| *Figure 2.6: IV iron vs usual care/no iron – Adverse events related to wound healing*  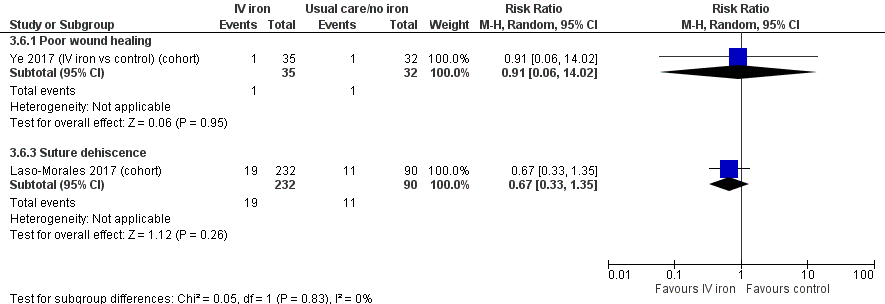 | | | | |

| **2.7 Bronchopulmonary (Figure 2.7)** | | | | |
| --- | --- | --- | --- | --- |
| Dyspnea | IV iron  vs usual care/no iron | *Meta-analysis:*  Statistically significant:  4/286 vs 15/153 §  RR: 0.24, 95%CI [0.08;0.75] (p=0.01)* *in favour of IV iron* | 2, 286 vs 153 | Ellermann, 2018 (cohort); Triphaus, 2019 (cohort) |
| Hyperventilation |  | Not statistically significant:  1/184 vs 0/50 § RR: 0.83, 95%CI [0.03;20.00] ¥ (p=0.91)* | 1, 184 vs 50 | Triphaus, 2019 (cohort) |
| Respiratory failure |  | Not statistically significant: 3/40 vs 3/32 §  RR: 0.80, 95%CI [0.17;3.70] ¥ (p=0.78)* | 1, 40 vs 32 | Froessler, 2016 |
| Pneumonia/respiratory failure |  | Not statistically significant: 0/38 vs 1/62 § RR: 0.54, 95%CI [0.02;12.89] ¥ (p=0.70)* | 1, 38 vs 62 | Kam, 2020 (cohort) |
| *Figure 2.7: IV iron vs usual care/no iron – Bronchopulmonary adverse events*  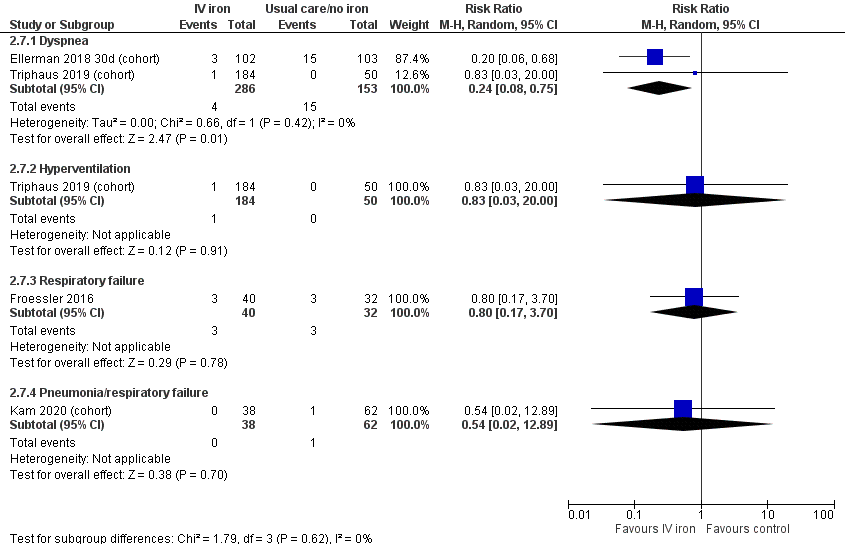 | | | | |

| **2.8 Infection (Figure 2.8)** | | | | |
| --- | --- | --- | --- | --- |
| Surgical/superficial wound infection | IV iron  vs usual care/no iron | *Meta-analysis:* Not statistically significant: 13/270 vs 10/152 §  RR: 0.60, 95%CI [0.27;1.36] ¥ (p=0.22)* | 2, 270 vs 152 | Kam, 2020 (cohort); Laso-Morales, 2017 (cohort) |
| Intra-abdominal abscess |  | Not statistically significant: 2/232 vs 1/90 §  RR: 0.78, 95%CI [0.07;8.45] ¥ (p=0.84)* | 1, 232 vs 90 | Laso-Morales, 2017 (cohort) |
| Urinary tract infection |  | Not statistically significant: 5/232 vs 3/90 § RR: 0.65, 95%CI [0.16;2.65] ¥ (p=0.54)* |  |  |
| Infection of the vaginal stump |  | Not statistically significant:  1/35 vs 0/32 §  RR: 2.75, 95%CI [0.12;65.18] ¥ (p=0.53)* | 1, 35 vs 32 | Ye, 2017 (cohort) |
| 30-day sepsis rate |  | Not statistically significant:  0/102 vs 1/103 §  RR: 0.34, 95%CI [0.01;8.17] ¥ (p=0.50)* | 1, 102 vs 103 | Ellermann, 2018 (cohort) |
| 90-day sepsis rate |  | 0/99 vs 0/101 §  Effect size not estimable | 1, 99 vs 101 |  |
| 1-year sepsis rate |  | 0/59 vs 0/59 §  Effect size not estimable | 1, 59 vs 59 |  |
| Pneumonia |  | Not statistically significant: 3/232 vs 3/90 §  RR: 0.39, 95%CI [0.08;1.89] ¥ (p=0.24)* | 1, 232 vs 90 | Laso-Morales, 2017 (cohort) |
| 30-day pneumonia rate |  | Not statistically significant:  2/102 vs 1/103 §  RR: 2.02, 95%CI [0.19;21.93] ¥ (p=0.56)* | 1, 102 vs 103 | Ellermann, 2018 (cohort) |
| 90-day pneumonia rate |  | Not statistically significant: 2/99 vs 2/101 §  RR: 1.02, 95%CI [0.15;7.10] ¥ (p=0.98)* | 1, 99 vs 101 |  |
| 1-year pneumonia rate |  | Not statistically significant:  3/59 vs 1/59 §  RR: 3.00, 95%CI [0.32;28.02] ¥ (p=0.33)* | 1, 59 vs 59 |  |
| Infection |  | Not statistically significant: 4/40 vs 5/32 §  RR: 0.64, 95%CI [0.19;2.19] ¥ (p=0.48)* | 1, 40 vs 32 | Froessler, 2016 |
| 30-day infection rate |  | Not statistically significant: 21/102 vs 25/103 § RR: 0.85, 95%CI [0.51;1.41] ¥ (p=0.53)* | 1, 102 vs 103 | Ellermann, 2018 (cohort) |
| 90-day infection rate |  | Not statistically significant:  16/99 vs 18/101 §  RR: 0.91, 95%CI [0.49;1.68] ¥ (p=0.75)* | 1, 99 vs 101 |  |
| 1-year infection rate |  | Statistically significant: 6/59 vs 17/59 §  RR: 0.35, 95%CI [0.15;0.83] (p=0.0173)* *in favour of IV iron* | 1, 59 vs 59 |  |
| Prevalence of infectious-related codes during hospital stay |  | Statistically significant:  47/149 vs 72/170 §  RR: 0.74, 95%CI [0.55;0.99] (p=0.0498)* *in favour of IV iron* | 1, 149 vs 170 |  |
| Readmission rate for general infection between discharge and 8 weeks |  | Not statistically significant:  6/234 vs 7/234 § RR: 0.86, 95%CI [0.29;2.51] ¥ (p=0.78)* | 1, 234 vs 234 | Richards, 2020 |
| Readmission rate for wound infection between discharge and 8 weeks |  | Statistically significant: 1/234 vs 8/234 § RR: 0.13, 95%CI [0.02;0.99] (p=0.0491)* *in favour of IV iron* |  |  |
| *Figure 2.8: IV iron vs usual care/no iron – Infectious adverse events*  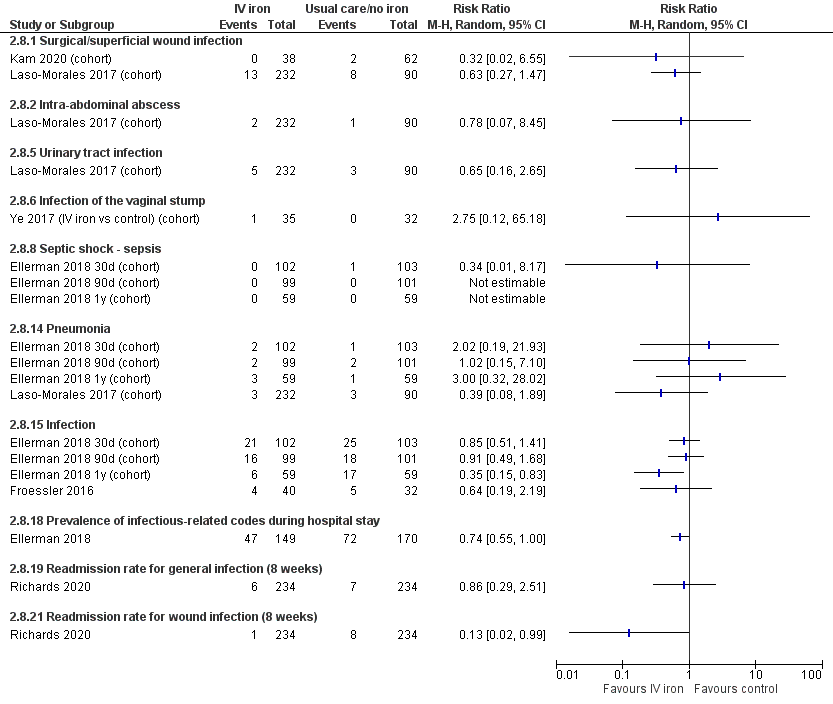 | | | | |

| **2.9 Bleeding (Figure 2.9)** | | | | |
| --- | --- | --- | --- | --- |
| Surgical wound hematoma | IV iron  vs usual care/no iron | Not statistically significant:  15/232 vs 8/90 §  RR: 0.73, 95%CI [0.32;1.66] ¥ (p=0.45)* | 1, 232 vs 90 | Laso-Morales, 2017 (cohort) |
| Rectorrhagia/ hemoperitoneum |  | Not statistically significant:  11/232 vs 5/90 §  RR: 0.85, 95%CI [0.31;2.39] ¥ (p=0.76)* |  |  |
| Bleeding |  | Not statistically significant:  0/38 vs 2/62 § RR: 0.32, 95%CI [0.02;6.55] ¥ (p=0.46)* | 1, 38 vs 62 | Kam, 2020 (cohort) |
| *Figure 2.9: IV iron vs usual care/no iron – Bleeding adverse events*  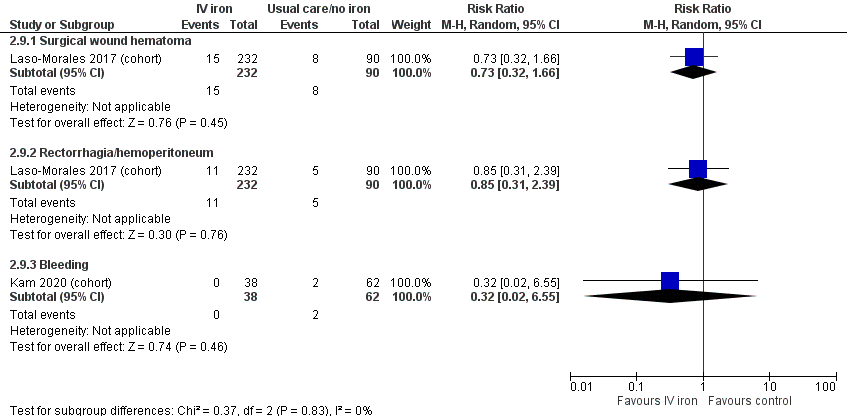 | | | | |

| **2.10 Cardiovascular (Figure 2.10)** | | | | |
| --- | --- | --- | --- | --- |
| 30-day heart failure rate | IV iron  vs usual care/no iron | Not statistically significant: 1/102 vs 0/103 §  RR: 3.03, 95%CI [0.12;73.50] ¥ (p=0.50)* | 1, 102 vs 103 | Ellermann, 2018 (cohort) |
| 90-day heart failure rate |  | 0/99 vs 0/101 §  Effect size not estimable | 1, 99 vs 101 |  |
| 1-year heart failure rate |  | 0/59 vs 0/59 §  Effect size not estimable | 1, 59 vs 59 |  |
| 30-day edema rate |  | Not statistically significant: 1/102 vs 1/103 §  RR: 1.01, 95%CI [0.06;15.93] ¥ (p=0.99)* | 1, 102 vs 103 |  |
| Angioedema |  | Not statistically significant: 1/184 vs 0/50 § RR: 0.83, 95%CI [0.03;20.00] ¥ (p=0.91)* | 1, 184 vs 50 | Triphaus, 2019 (cohort) |
| Shock with hypotension |  | 0/184 vs 0/50 §  Effect size not estimable |  |  |
| Tachycardia |  | 0/222 vs 0/112 §  Effect size not estimable | 2, 222 vs 112 | Kam, 2020 (cohort); Triphaus, 2019 (cohort) |
| Arrhythmia |  | 0/184 vs 0/50 §  Effect size not estimable | 1, 184 vs 50 | Triphaus, 2019 (cohort) |
| Cardiovascular arrest |  | 0/184 vs 0/50 §  Effect size not estimable |  |  |
| *Figure 2.10: IV iron vs usual care/no iron – Cardiovascular adverse events*  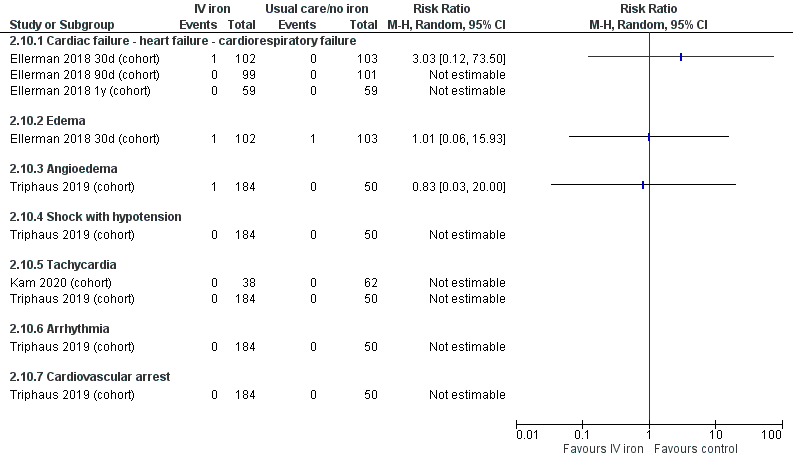 | | | | |

| **2.11 Renal (Figure 2.11)** | | | | |
| --- | --- | --- | --- | --- |
| Renal impairment | IV iron  vs usual care/no iron | Not statistically significant:  1/40 vs 1/32 §  RR: 0.80, 95%CI [0.05;12.30] ¥ (p=0.87)* | 1, 40 vs 32 | Froessler, 2016 |
| Renal failure |  | Not statistically significant: 0/38 vs 1/62 § RR: 0.54, 95%CI [0.02;12.89] ¥ (p=0.70)* | 1, 38 vs 62 | Kam, 2020 (cohort) |
| 30-day renal failure rate |  | Not statistically significant:  0/102 vs 2/103 §  RR: 0.20, 95%CI [0.01;4.16] ¥ (p=0.30)* | 1, 102 vs 103 | Ellermann, 2018 (cohort) |
| 90-day renal failure rate |  | Not statistically significant:  1/99 vs 1/101 §  RR: 1.02, 95%CI [0.06;16.09] ¥ (p=0.99)* | 1, 99 vs 101 |  |
| 1-year renal failure rate |  | Not statistically significant:  1/59 vs 0/59 §  RR: 3.00, 95%CI [0.12;72.18] ¥ (p=0.50)* | 1, 59 vs 59 |  |
| *Figure 2.11: IV iron vs usual care/no iron – Renal adverse events*  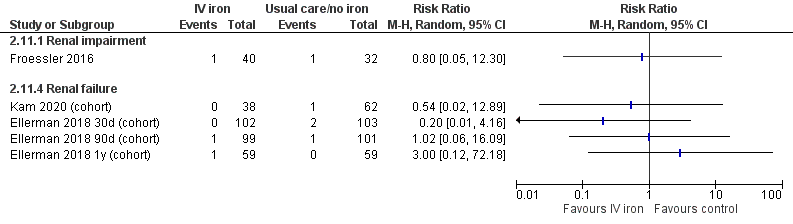 | | | | |

| **2.12 Anaemia-associated ischemic events (Figure 2.12)** | | | | |
| --- | --- | --- | --- | --- |
| Myocardial infarction | IV iron  vs usual care/no iron | 0/38 vs 0/62 § Effect size not estimable | 1, 38 vs 62 | Kam, 2020 (cohort) |
| 30-day myocardial infarction rate |  | Not statistically significant:  0/102 vs 1/103 §  RR: 0.34, 95%CI [0.01;8.17] ¥ (p=0.50)* | 1, 102 vs 103 | Ellermann, 2018 (cohort) |
| 90-day myocardial infarction rate |  | Not statistically significant:  0/99 vs 1/101 §  RR: 0.34, 95%CI [0.01;8.25] ¥ (p=0.51)* | 1, 99 vs 101 |  |
| 1-year myocardial infarction rate |  | Not statistically significant:  0/59 vs 1/59 §  RR: 0.33, 95%CI [0.01;8.02] ¥ (p=0.50)* | 1, 59 vs 59 |  |
| Stroke |  | 0/38 vs 0/62 § Effect size not estimable | 1, 38 vs 62 | Kam, 2020 (cohort) |
| 30-day stroke rate |  | Not statistically significant:  1/102 vs 1/103 §  RR: 1.01, 95%CI [0.06;15.93] ¥ (p=0.99)* | 1, 102 vs 103 | Ellermann, 2018 (cohort) |
| 90-day stroke rate |  | Not statistically significant:  1/99 vs 0/101 §  RR: 3.06, 95%CI [0.13;74.23] ¥ (p=0.49)* | 1, 99 vs 101 |  |
| 1-year stroke rate |  | 0/59 vs 0/59 §  Effect size not estimable | 1, 59 vs 59 |  |
| Perioperative acute kidney injury |  | Not statistically significant:  11/137 vs 13/122 §  RR: 0.75, 95%CI [0.35;1.62] ¥ (p=0.47)* | 1, 137 vs 122 | Richards, 2020 |
| *Figure 2.12: IV iron vs usual care/no iron – Anaemia-associated ischemic events*  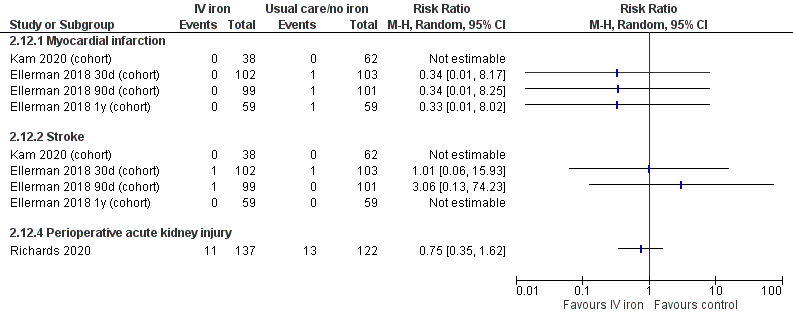 | | | | |

| **2.13 Thromboembolic events (Figure 2.13)** | | | | |
| --- | --- | --- | --- | --- |
| Deep venous thrombosis | IV iron  vs usual care/no iron | Not statistically significant:  0/40 vs 1/32 §  RR: 0.27, 95%CI [0.01;6.37] ¥ (p=0.42)* | 1, 40 vs 32 | Froessler, 2016 |
|  |  | *Meta-analysis:*  Not statistically significant:  0/305 vs 2/184 §  RR: 0.20, 95%CI [0.02;1.89] ¥ (p=0.16)* | 3, 305 vs 184 | Kam, 2020 (cohort); Laso-Morales, 2017 (cohort); Ye, 2017 (cohort) |
| Thrombosis |  | 0/35 vs 0/32 §  Effect size not estimable | 1, 35 vs 32 | Ye, 2017 (cohort) |
| *Figure 2.13: IV iron vs usual care/no iron – Thromboembolic events*  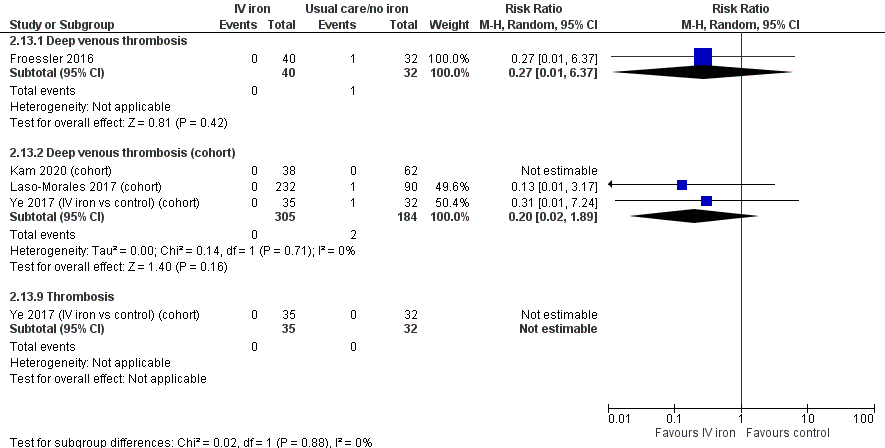 | | | | |

| **2.14 Mortality** | | | | |
| --- | --- | --- | --- | --- |
| ***Findings presented in Figure 2.14*** | | | | |
| Mortality | IV iron  vs usual care/no iron | *Meta-analysis:* Not statistically significant:  3/279 vs 2/273 §  RR: 1.28, 95%CI [0.24;6.76] ¥ (p=0.77)* | 2, 279 vs 273 | Froessler, 2016; Richards, 2020 |
|  |  | *Meta-analysis:*  Not statistically significant:  3/187 vs 6/253 §  RR: 0.54, 95%CI [0.14;2.16] ¥ (p=0.39)* | 4, 187 vs 253 | Evans, 2021 (cohort); Kam, 2020 (cohort); Klein, 2020 (cohort); Nandhra, 2020 (cohort) |
| *Figure 2.14: IV iron vs usual care/no iron – Mortality*  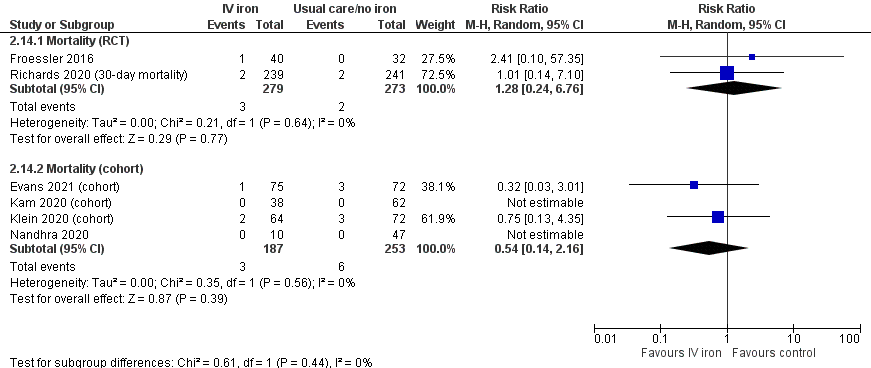 | | | | |
| ***Additional findings (not presented in Figure 2.14)*** | | | | |
| 6-month mortality | IV iron vs usual care/no iron | Not statistically significant: 12/238 vs 10/236 § RR: 1.19, 95%CI [0.52;2.70] ¥ (p=0.68)* | 1, 238 vs 236 | Richards, 2020 |

| **2.15 Survival (Figure 2.15)** | | | | |
| --- | --- | --- | --- | --- |
| 1-year overall survival | IV iron  vs usual care/no iron | Not statistically significant:  76/83 vs 78/83 §  RR: 0.97, 95%CI [0.89;1.06]  (p=0.55)* | 1, 83 vs 83 | Wilson, 2018 B (cohort) |
| 2-year overall survival |  | Not statistically significant:  56/83 vs 64/83 §  RR: 0.88, 95%CI [0.72;1.06]  (p=0.17)* |  |  |
| 3-year overall survival |  | Not statistically significant:  40/83 vs 48/83 §  RR: 0.83,95%CI [0.62;1.11]  (p=0.22)* |  |  |
| 4-year overall survival |  | Statistically significant:  22/83 vs 40/83 §  RR: 0.55, 95%CI [0.36;0.84]  (p=0.005)* *in favour of usual care/no iron* |  |  |
| 5-year overall survival |  | Statistically significant:  10/83 vs 25/83 §  RR: 0.40, 95%CI [0.21;0.78]  (p=0.007)* *in favour of usual care/no iron* |  |  |
| 1-year disease-free survival |  | Not statistically significant:  68/79 vs 64/78 §  RR: 1.05, 95%CI [0.92;1.20]  (p=0.49)* | 1, 79 vs 78 |  |
| 2-year disease-free survival |  | Not statistically significant:  47/79 vs 46/78 §  RR: 1.01, 95%CI [0.78;1.31] ¥  (p=0.95)* |  |  |
| 3-year disease-free survival |  | Not statistically significant:  27/79 vs 34/78 §  RR: 0.78, 95%CI [0.53;1.17]  (p=0.23)* |  |  |
| 4-year disease-free survival |  | Statistically significant:  13/79 vs 29/78 §  RR: 0.44, 95%CI [0.25;0.79]  (p=0.005)*  *in favour of usual care/no iron* |  |  |
| 5-year disease-free survival |  | Statistically significant:  4/79 vs 18/78 §  RR: 0.22, 95%CI [0.08;0.62]  (p=0.004)*  *in favour of usual care/no iron* |  |  |

*Figure 2.15: IV iron vs usual care/no iron – Survival*

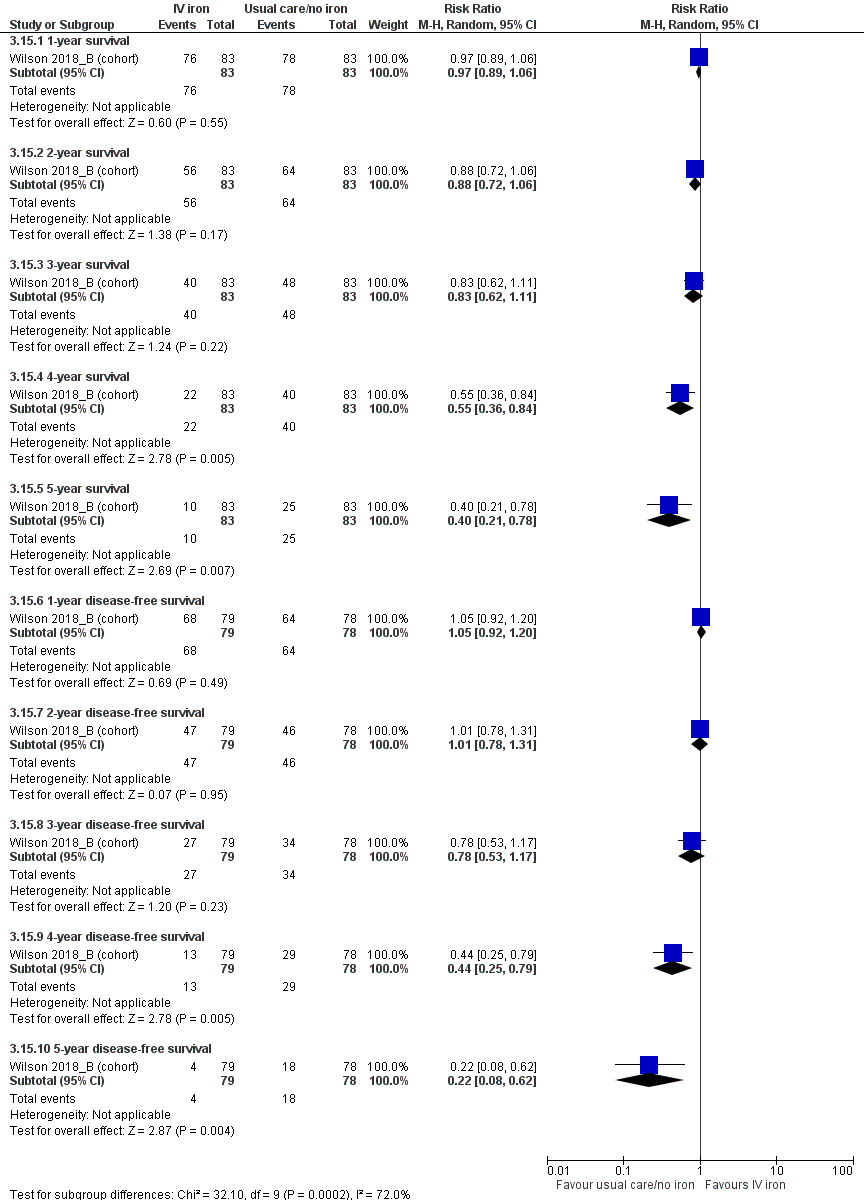

| **2.16 Other adverse events (Figure 2.16)** | | | | |
| --- | --- | --- | --- | --- |
| Allergy | IV iron  vs usual care/no iron | 0/35 vs 0/32 §  Effect size not estimable | 1, 35 vs 32 | Ye, 2017 (cohort) |
| Severe allergic reactions |  | 0/38 vs 0/62 §  Effect size not estimable | 1, 38 vs 62 | Kam, 2020 (cohort) |
| Lymph node swelling |  | 0/184 vs 0/50 §  Effect size not estimable | 1, 184 vs 50 | Triphaus, 2019 (cohort) |
| Postoperative overall complication rate (composite of pulmonic, cardiologic, thrombotic, infectious and neurologic complications) |  | Not statistically significant:  24/94 vs 77/224 §  RR: 0.74, 95%CI [0.50;1.10] (p=0.13)* | 1, 94 vs 224 | Wilson, 2018 A (cohort) |
| Postoperative complication rate ≥CD grade III |  | Not statistically significant: 22/233 vs 24/227 § RR: 0.89, 95%CI [0.52; 1.55] ¥ (p=0.69)* | 1, 233 vs 277 | Richards, 2020 |
| Hospital readmission |  | Not statistically significant:  6/40 vs 3/32 §  RR: 1.60, 95%CI [0.43;5.90] ¥ (p=0.48)* | 1, 40 vs 32 | Froessler, 2016 |
|  |  | *Meta-analysis:* Not statistically significant:  13/74 vs 11/119 §  RR: 2.13, 95%CI [0.94;4.86] ¥ (p=0.07)* | 2, 74 vs 119 | Klein, 2020 (cohort); Nandhra, 2020 (cohort) |
| Readmission for general postoperative complications between discharge and 8 weeks |  | Not statistically significant:  25/234 vs 36/234 § RR: 0.69, 95%CI [0.43;1.12] ¥ (p=0.13)* | 1, 234 vs 234 | Richards, 2020 |
| Readmission for complications between discharge and 6 months |  | Not statistically significant:  58/227 vs 73/223 § RR: 0.78, 95%CI [0.58;1.04] ¥ (p=0.10)* | 1, 227 vs 223 |  |
| Reoperation rate |  | Not statistically significant:  1/38 vs 3/62 § RR: 0.54, 95%CI [0.06;5.04] ¥ (p=0.59)* | 1, 38 vs 62 | Kam, 2020 (cohort) |
| Intra-abdominal collection of fluids (due to abscess, rim enhancement, fluid around anastomosis with suspected leakage) |  | Not statistically significant:  2/38 vs 3/62 §  RR: 1.09, 95%CI [0.19;6.22] ¥ (p=0.92)* |  |  |
| Anastomotic leak |  | Not statistically significant:  1/38 vs 2/62 §  RR: 0.82, 95%CI [0.08;8.69] ¥ (p=0.87)* |  |  |
| Chyle leak |  | Not statistically significant:  1/38 vs 0/62 §  RR: 4.85, 95%CI [0.20;116.02] ¥ (p=0.33)* |  |  |

*Figure 2.16: IV iron vs usual care/no iron – Other adverse events*

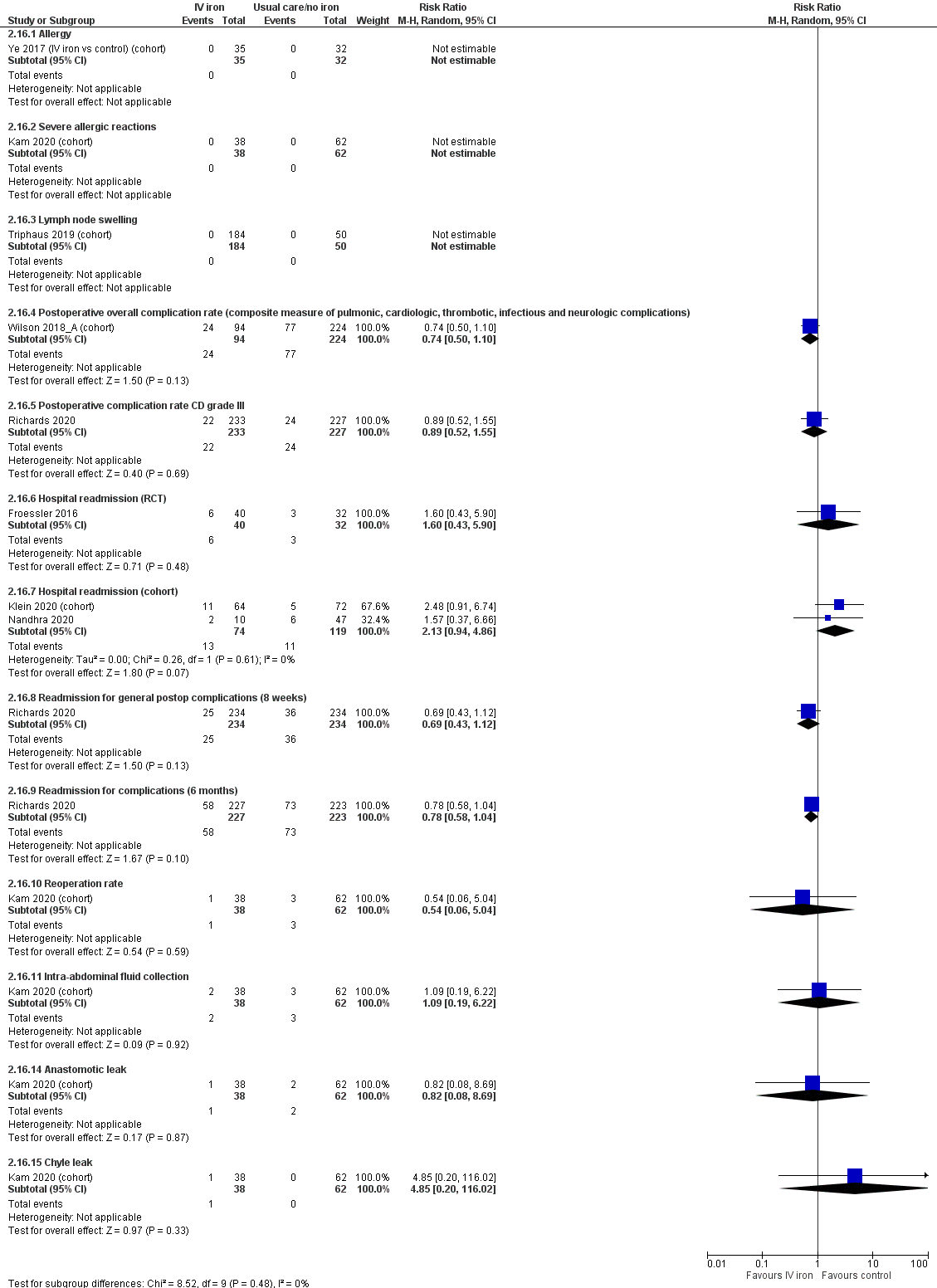

| **COMPARISON 3: IV FERRIC CARBOXYMALTOSE VS IRON SUCROSE** | | | | |
| --- | --- | --- | --- | --- |
| **3.1 Mortality** | | | | |
| Mortality | IV ferric carboxymaltose  vs IV iron sucrose | 0/52 vs 0/49  Effect size not estimable | 1, 52 vs 49 | Lee, 2019 |
| **3.2 Other adverse events** | | | | |
| Anaphylactic reactions | IV ferric carboxymaltose  vs IV iron sucrose | 0/52 vs 0/49  Effect size not estimable | 1, 52 vs 49 | Lee, 2019 |

| **ESA + IRON ADMINISTRATION** | | | | |
| --- | --- | --- | --- | --- |
| **COMPARISON 4: ESA + IRON VS CONTROL (PLACEBO AND/OR IRON OR NO TREATMENT)** | | | | |
| **4.1 Gastrointestinal (Figure 4.1)** | | | | |
| Nausea | ESA + oral iron  vs control  (placebo and/or oral iron or no treatment) | Not statistically significant:  91/340 vs 99/340 §  RR: 0.92, 95%CI [0.72;1.17] (p=0.49)* | 1, 340 vs 340 | Stowell, 2009 |
|  | ESA + IV iron  vs control  (placebo and/or IV iron or no treatment) | *Meta-analysis:* Not statistically significant:  1/72 vs 2/69 §  RR: 0.46, 95%CI [0.04;4.80] ¥ (p=0.51)* | 2, 72 vs 69 | Cao, 2020; Yoo, 2011 |
| Vomitus | ESA + oral iron  vs control  (placebo and/or oral iron or no treatment) | Not statistically significant:  21/340 vs 23/340 §  RR: 0.91, 95%CI [0.52;1.62] ¥ (p=0.76)* | 1, 340 vs 340 | Stowell, 2009 |
|  | ESA + IV iron  vs control  (placebo and/or IV iron or no treatment) | *Meta-analysis:* Not statistically significant:  1/72 vs 2/69 §  RR: 0.46, 95%CI [0.04;4.80] ¥ (p=0.51)* | 2, 72 vs 69 | Cao, 2020; Yoo, 2011 |
| Diarrhoea | ESA + IV iron  vs control  (placebo and/or IV iron or no treatment) | 0/37 vs 0/37 §  Effect size not estimable | 1, 37 vs 37 | Yoo, 2011 |
| Obstipation | ESA + oral iron  vs control  (placebo and/or oral iron or no treatment) | *Meta-analysis:* Not statistically significant:  58/392 vs 55/397 §  RR: 1.05, 95%CI [0.75;1.47] ¥ (p=0.78)* | 2, 392 vs 397 | Kettelhack, 1998; Stowell, 2009 |
| Abdominal pain | ESA + IV iron  vs control  (placebo and/or IV iron or no treatment) | Not statistically significant:  1/48 vs 0/52 §  RR: 3.24, 95%CI [0.14;77.79] ¥ (p=0.47)* | 1, 48 vs 52 | Urena, 2017 |
| Gastrointestinal reaction |  | 0/30 vs 0/67 §  Effect size not estimable | 1, 30 vs 67 | Ye, 2017 (cohort) |
| Incomplete intestinal obstruction |  | Not statistically significant:  0/30 vs 2/67 §  RR: 0.44, 95%CI [0.02;8.87] ¥ (p=0.59)* |  |  |

*Figure 4.1: ESA + iron vs control (placebo and/or iron or no treatment) – Gastrointestinal adverse events*

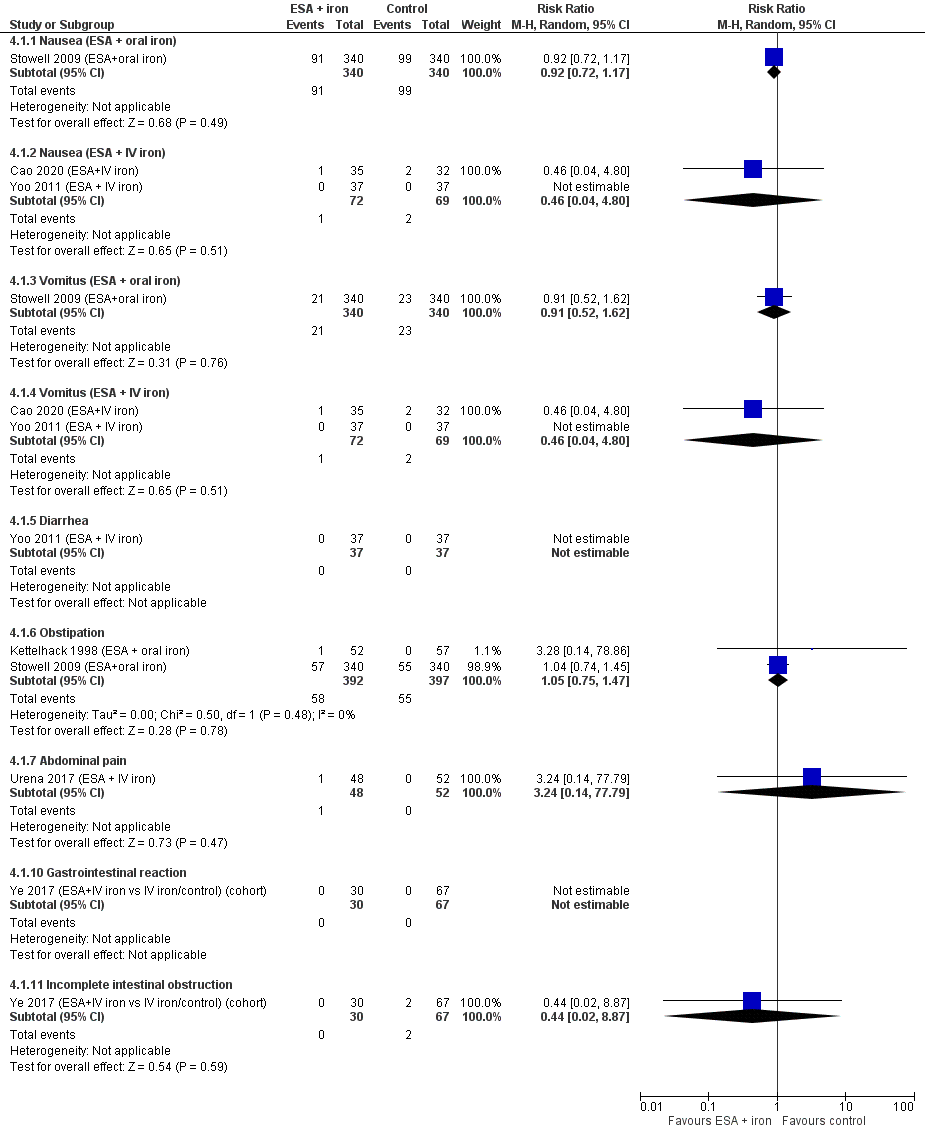

| **4.2 Mucocutaneous (Figure 4.2)** | | | | |
| --- | --- | --- | --- | --- |
| Rash | ESA + oral iron  vs control  (placebo and/or oral iron or no treatment) | Not statistically significant:  1/136 vs 0/68 §  RR: 1.51, 95%CI [0.06;36.61] ¥ (p=0.80)* | 1, 136 vs 68 | Christodoulakis, 2005 |
| Urticarial allergy to iron |  | Not statistically significant:  0/136 vs 1/68 §  RR: 0.17, 95%CI [0.01;4.07] ¥ (p=0.27)* |  |  |
| Unexplained grade 2 urticaria |  | Not statistically significant:  1/136 vs 0/68 §  RR: 1.51, 95%CI [0.06;36.61] ¥ (p=0.80)* |  |  |
| Pruritus |  | Not statistically significant:  39/340 vs 43/340 §  RR: 0.91, 95%CI [0.60;1.36] ¥ (p=0.64)* | 1, 340 vs 340 | Stowell, 2009 |
| *Figure 4.2: ESA + iron vs control (placebo and/or iron or no treatment) – Mucocutaneous adverse events*  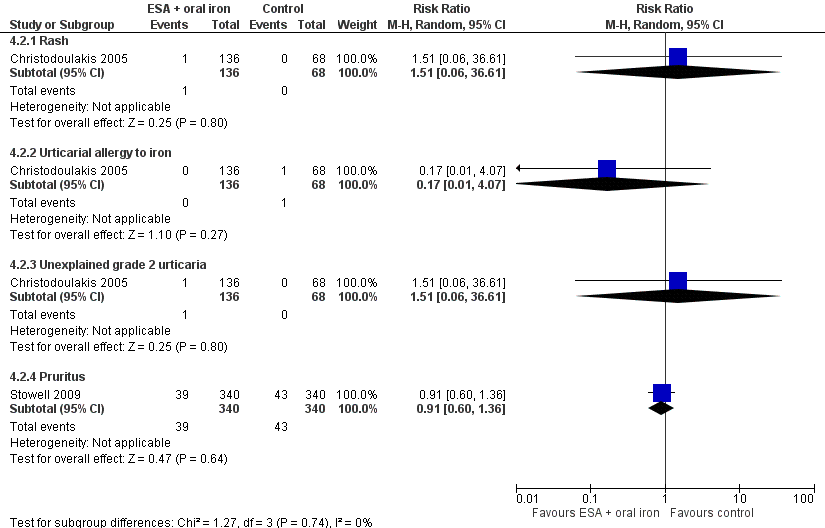 | | | | |

| **4.3 Autonomic (Figure 4.3)** | | | | |
| --- | --- | --- | --- | --- |
| Febrile episodes | ESA + oral iron  vs control  (placebo and/or oral iron or no treatment) | Not statistically significant:  2/23 vs 2/27 §  RR: 1.17, 95%CI [0.18;7.69] ¥ (p=0.87)* | 1, 23 vs 27 | Dousias, 2003 |
| Postoperative fever |  | Not statistically significant:  3/20 vs 3/18 §  RR: 0.90, 95%CI [0.21;3.91] ¥ (p=0.89)* | 1, 20 vs 18 | Dousias, 2005 |
| Chills with fever |  | Not statistically significant:  1/52 vs 0/57 §  RR: 3.28, 95%CI [0.14;78.86] ¥ (p=0.46)* | 1, 52 vs 57 | Kettelhack, 1998 |
| Pyrexia |  | Not statistically significant:  57/340 vs 58/340 §  RR: 0.98, 95%CI [0.70;1.37] ¥ (p=0.92)* | 1, 340 vs 340 | Stowell, 2009 |
| Mild fever | ESA + IV iron  vs control  (placebo and/or IV iron or no treatment) | 0/30 vs 0/67 §  Effect size not estimable | 1, 30 vs 67 | Ye, 2017 (cohort) |
| *Figure 4.3: ESA + iron vs control (placebo and/or iron or no treatment) – Autonomic adverse events*  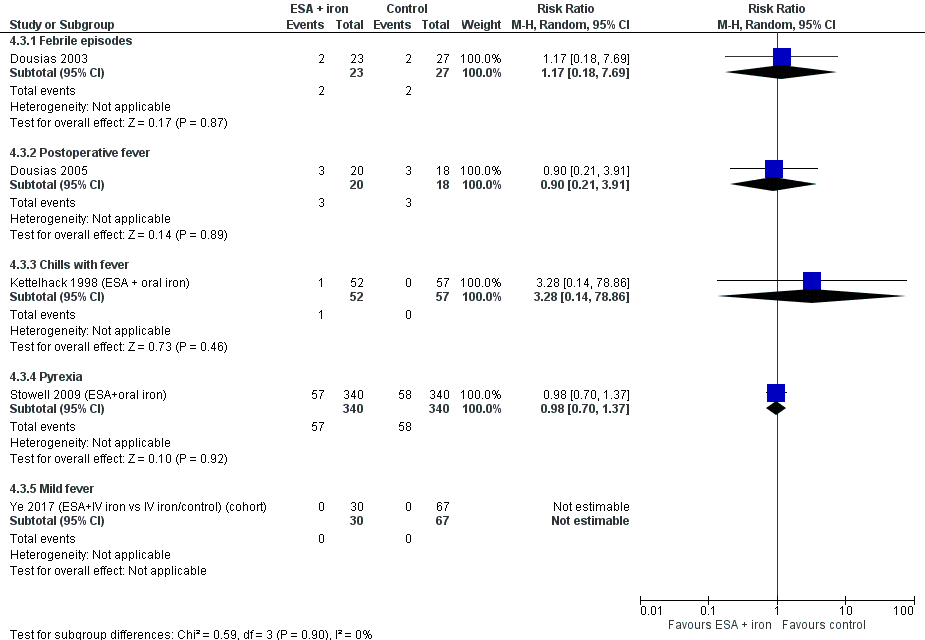 | | | | |

| **4.4 Neuro-psychosomatic (Figure 4.4)** | | | | |
| --- | --- | --- | --- | --- |
| Headache | ESA + oral iron  vs control  (placebo and/or oral iron or no treatment) | Not statistically significant:  24/340 vs 21/340 §  RR: 1.14, 95%CI [0.65;2.01] ¥ (p=0.64)* | 1, 340 vs 340 | Stowell, 2009 |
|  | ESA + IV iron  vs control  (placebo and/or IV iron or no treatment) | 0/30 vs 0/67 §  Effect size not estimable | 1, 30 vs 67 | Ye, 2017 (cohort) |
|  |  | 0/37 vs 0/37 §  Effect size not estimable | 1, 37 vs 37 | Yoo, 2011 |
| Back pain | ESA + oral iron  vs control  (placebo and/or oral iron or no treatment) | Statistically significant:  18/340 vs 7/340 §  RR: 2.57, 95%CI [1.09;6.08]  (p=0.03)*  *in favour of control* | 1, 340 vs 340 | Stowell, 2009 |
| Chest pain |  | Not statistically significant:  5/340 vs 4/340 §  RR: 1.25, 95%CI [0.34;4.61] ¥ (p=0.74)* |  |  |
| Post-procedural pain |  | Not statistically significant:  71/340 vs 72/340 § RR: 0.99, 95%CI [0.74;1.32] ¥ (p=0.93)* |  |  |
| Myalgia | ESA + IV iron  vs control  (placebo and/or IV iron or no treatment) | 0/30 vs 0/67 §  Effect size not estimable | 1, 30 vs 67 | Ye, 2017 (cohort) |

*Figure 4.4: ESA + iron vs control (placebo and/or iron or no treatment) – Neuro-psychosomatic adverse events*

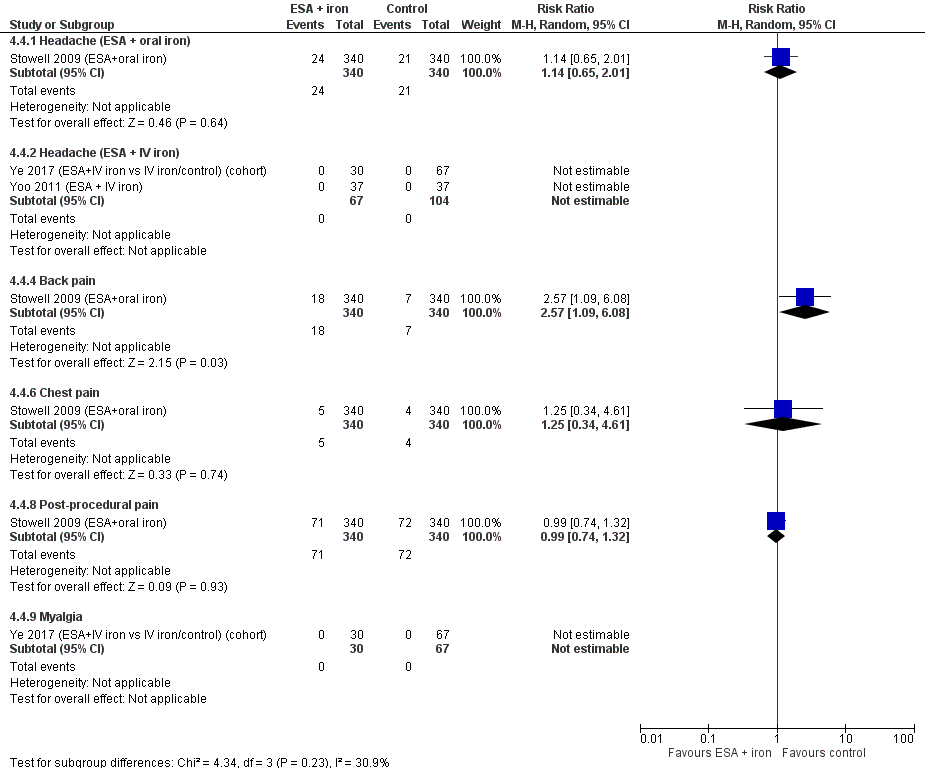

| **4.5 Neurological (Figure 4.5)** | | | | |
| --- | --- | --- | --- | --- |
| Neurological complications at 45 days | ESA + oral iron  vs control  (placebo and/or oral iron or no treatment) | Not statistically significant:  4/300 vs 5/300 §  RR: 0.80, 95%CI [0.22;2.95] ¥ (p=0.74)* | 1, 300 vs 300 | Weltert, 2015 |
| Focal neurologic damage |  | Not statistically significant:  5/300 vs 6/300 §  RR: 0.83, 95%CI [0.26;2.70] ¥ (p=0.76)* |  |  |
| Generalized neurologic damage |  | Not statistically significant:  0/300 vs 1/300 §  RR: 0.33, 95%CI [0.01;8.15] ¥ (p=0.50)* |  |  |
| *Figure 4.5: ESA + oral iron vs control (placebo and/or oral iron or no treatment) – Neurological adverse events*  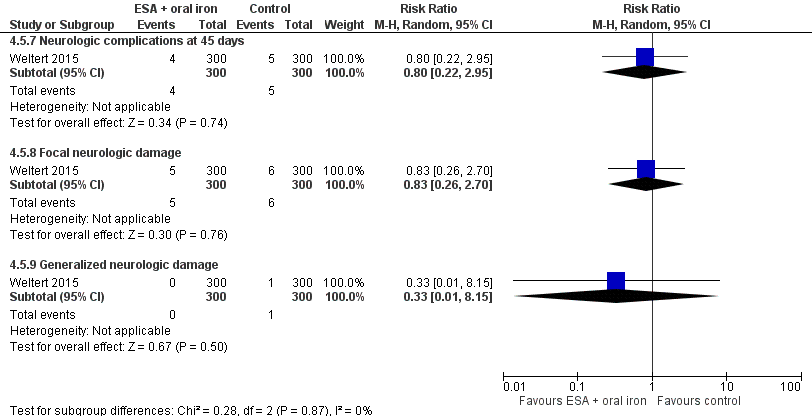 | | | | |
| **4.6 Wound healing (Figure 4.6)** | | | | |
| Poor wound healing | ESA + IV iron  vs control  (placebo and/or IV iron or no treatment) | Not statistically significant:  0/30 vs 2/67 §  RR: 0.44, 95%CI [0.02;8.87] ¥ (p=0.59)* | 1, 30 vs 67 | Ye, 2017 (cohort) |
| *Figure 4.6: ESA + iron vs control (placebo and/or iron or no treatment) – Adverse events related to wound healing*  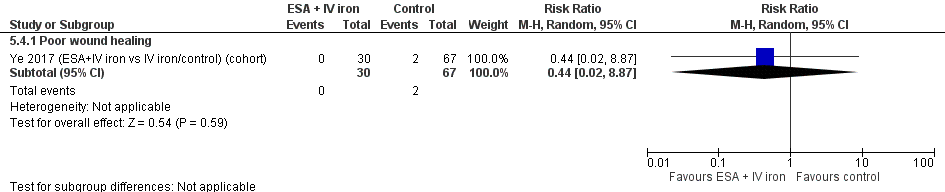 | | | | |

| **4.7 Bronchopulmonary (Figure 4.7)** | | | | |
| --- | --- | --- | --- | --- |
| Respiratory failure | ESA + oral iron  vs control  (placebo and/or oral iron or no treatment) | Not statistically significant:  1/29 vs 0/29 §  RR: 3.00, 95%CI [0.13;70.74] ¥ (p=0.50)* | 1, 29 vs 29 | Scott, 2002 |
| Acute respiratory distress syndrome |  | Not statistically significant:  14/329 vs 13/329 §  RR: 1.06, 95%CI [0.51;2.20] ¥ (p=0.87)* | 1, 329 vs 329 | Scott, 2002; Weltert, 2015 |
| Acute pulmonary edema |  | Not statistically significant:  2/300 vs 2/300 §  RR: 1.00, 95%CI [0.14;7.05] ¥ (p=1.00)* | 1, 300 vs 300 | Weltert, 2015 |
| Prolonged ventilation | ESA + IV iron  vs control  (placebo and/or IV iron or no treatment) | Not statistically significant:  11/75 vs 17/59 §  RR: 0.51, 95%CI [0.26;1.00] ¥ (p=0.0507)* | 1, 75 vs 59 | Cladellas, 2012 (cohort) |
| *Figure 4.7: ESA + iron vs control (placebo and/or iron or no treatment) – Bronchopulmonary adverse events*  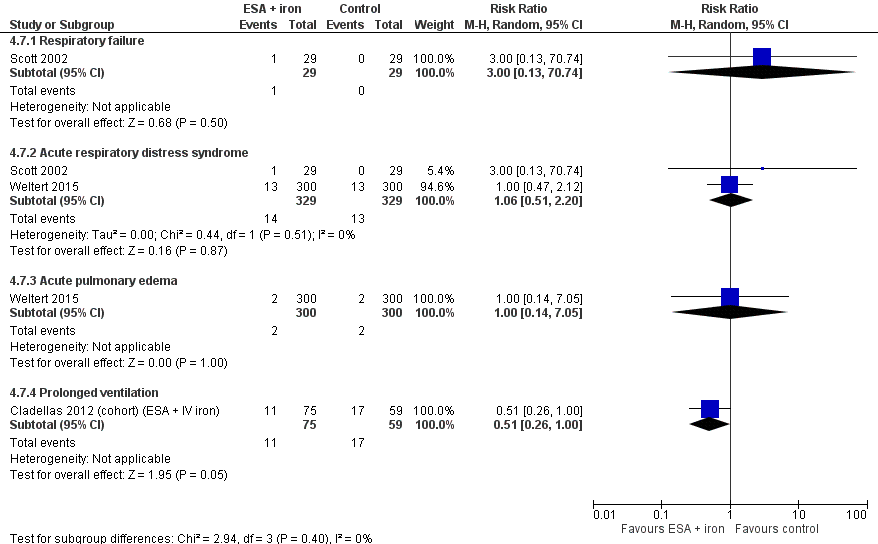 | | | | |

| **4.8 Infection** | | | | |
| --- | --- | --- | --- | --- |
| ***Findings presented in Figure 4.8*** | | | | |
| Surgical/superficial wound infection | ESA + oral iron  vs control  (placebo and/or oral iron or no treatment) | *Meta-analysis:* Not statistically significant:  9/655 vs 5/656 §  RR: 1.63, 95%CI [0.55;4.85] ¥ (p=0.38)* | 3, 655 vs 656 | Larson, 2001; Stowell, 2009; Weltert, 2015 |
| Urinary tract infection |  | *Meta-analysis:* Not statistically significant:  22/355 vs 17/356 §  RR: 1.30, 95%CI [0.71;2.41] ¥ (p=0.40)* | 2, 355 vs 356 | Larson, 2001; Stowell, 2009 |
| Cellulitis after surgery |  | Not statistically significant:  0/28 vs 1/53 §  RR: 0.62, 95%CI [0.03;14.76] ¥ (p=0.77)* | 1, 28 vs 53 | Delasotta, 2012 B (cohort) |
| Superficial thrombophlebitis |  | Not statistically significant:  2/340 vs 0/340 §  RR: 5.00, 95%CI [0.24;103.76] ¥ (p=0.30)* | 1, 340 vs 340 | Stowell, 2009 |
| Septic shock |  | Not statistically significant:  1/17 vs 0/10 §  RR: 1.83, 95%CI [0.08;41.17] ¥ (p=0.70)* | 1, 17 vs 10 | Heiss, 1996 |
| Sepsis | ESA + IV iron  vs control  (placebo and/or IV iron or no treatment) | Not statistically significant:  2/48 vs 0/52 §  RR: 5.41, 95%CI [0.27;109.87] ¥ (p=0.27)* | 1, 48 vs 52 | Urena, 2017 |
| Mycotic sepsis | ESA + oral iron  vs control  (placebo and/or oral iron or no treatment) | Not statistically significant:  0/52 vs 1/57 §  RR: 0.36, 95%CI [0.02;8.76] ¥ (p=0.53)* | 1, 52 vs 57 | Kettelhack, 1998 |
| Severe streptococcal septicaemia |  | Not statistically significant:  0/15 vs 1/16 §  RR: 0.35, 95%CI [0.02;8.08] ¥ (p=0.52)* | 1, 15 vs 16 | Larson, 2001 |
| Pneumonia |  | Not statistically significant:  10/300 vs 10/300 §  RR: 1.00, 95%CI [0.42;2.37] ¥ (p=1.00)* | 1, 300 vs 300 | Weltert, 2015 |
| Peritonitis |  | Not statistically significant:  1/136 vs 0/68 §  RR: 1.51, 95%CI [0.06;36.61] ¥ (p=0.80)* | 1, 136 vs 68 | Christodoulakis, 2005 |
| Severe infection (sepsis, pneumonia or mediastinitis) | ESA + IV iron  vs control  (placebo and/or IV iron or no treatment) | Statistically significant:  6/75 vs 14/59 §  RR: 0.34, 95%CI [0.14;0.82] (p=0.02)*  *in favour of ESA + IV iron* | 1, 75 vs 59 | Cladellas, 2012 (cohort) |
| Infection of the vaginal stump |  | Not statistically significant:  0/30 vs 1/67 § RR: 0.73, 95%CI [0.03;17.45] ¥ (p=0.85)* | 1, 30 vs 67 | Ye, 2017 (cohort) |
| ***Additional findings (not presented in Figure 4.8)*** | | | | |
| Endocarditis | ESA + IV iron vs control  (placebo and/or IV iron or no treatment) | Not statistically significant: £†  (p>0.05) | 1, 75 vs 59 § | Cladellas, 2012 (cohort) |
| *Figure 4.8: ESA + iron vs control (placebo and/or iron or no treatment) – Infectious adverse events*  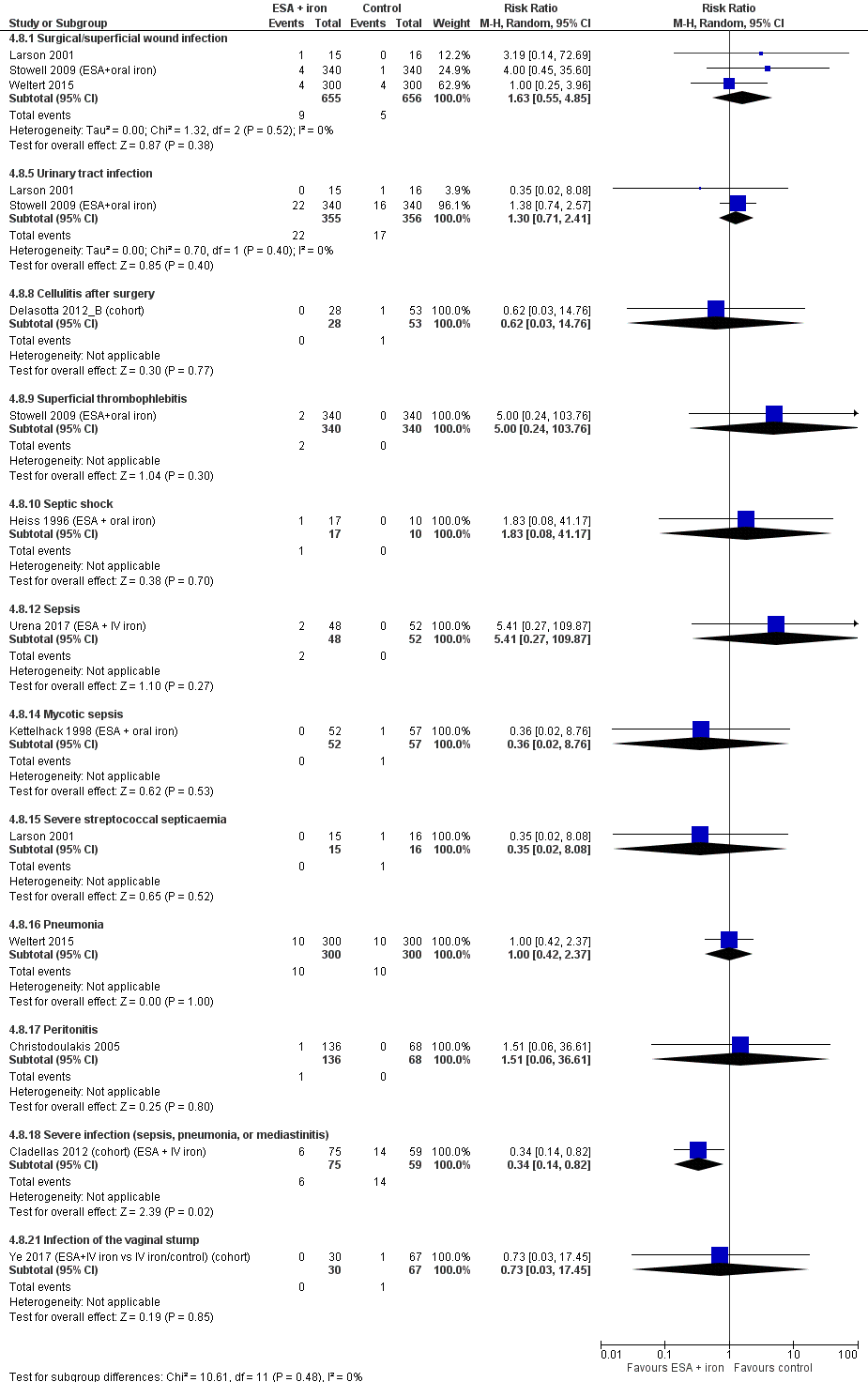 | | | | |

| **4.9 Bleeding (Figure 4.9)** | | | | |
| --- | --- | --- | --- | --- |
| Anastomosis rupture | ESA + oral iron  vs control  (placebo and/or oral iron or no treatment) | Not statistically significant: 1/29 vs 0/29 § RR: 3.00, 95%CI [0.13;70.74] ¥ (p=0.50)* | 1, 29 vs 29 | Scott, 2002 |
| Hemarthrosis |  | Not statistically significant: 0/28 vs 1/53 § RR: 0.62, 95%CI [0.03;14.76] ¥ (p=0.77)* | 1, 28 vs 53 | Delasotta 2012 B (cohort) |
| Major or life-threatening bleeding | ESA + IV iron  vs control  (placebo and/or IV iron or no treatment) | Not statistically significant:  7/48 vs 6/52 §  RR: 1.26, 95%CI [0.46;3.50] ¥ (p=0.65)* | 1, 48 vs 52 | Urena, 2017 |
| *Figure 4.9: ESA + iron vs control (placebo and/or iron or no treatment) – Bleeding adverse events*  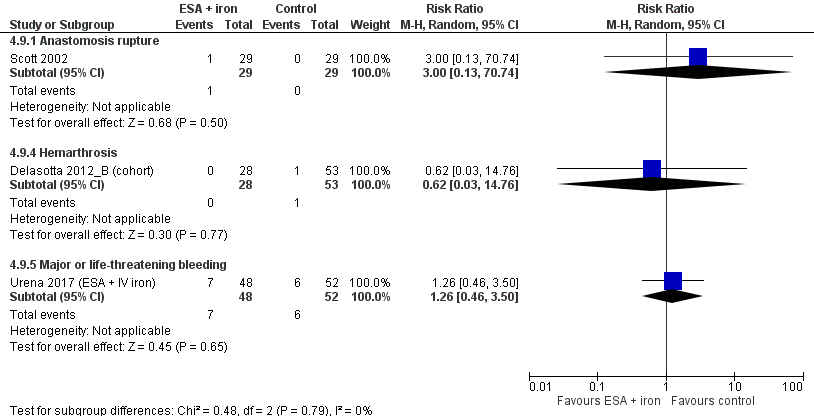 | | | | |

| **4.10 Cardiovascular** | | | | |
| --- | --- | --- | --- | --- |
| **Atrial fibrillation (Figure 4.10 A)** | | | | |
| Atrial fibrillation | ESA + iron  vs control  (placebo and/or iron or no treatment) | *Meta-analysis:*  Not statistically significant: 93/512 vs 113/450 §  RR: 0.84, 95%CI [0.67;1.06] ¥ (p=0.14)* | 4, 512 vs 450 | Christodoulakis, 2005;  Weltert, 2015;  Urena, 2017;  Yoo, 2011 |
|  |  | *Sensitivity analysis excluding studies that are not in line with the  WHO definition of anaemia  (excluding Weltert)* Not statistically significant: 8/212 vs 16/150 §  RR: 0.56, 95%CI [0.26;1.19] ¥ (p=0.13)* | 3, 212 vs 150 | Christodoulakis, 2005;  Urena, 2017;  Yoo, 2011 |
|  |  | *Sensitivity analysis excluding studies with high risk of bias on the items ‘inadequate measurement of the outcomes’ and ‘inadequate selection of the reported results’*  Not performed, as Weltert 2015 is the only study with low risk of bias on the item ‘inadequate measurement of the outcomes’ | N/A | N/A |
|  | ESA + oral iron  vs control  (placebo and/or oral iron or no treatment) | *Subgroup analysis ESA + oral iron:*  Not statistically significant: 85/436 vs 98/368 §  RR: 0.85, 95%CI [0.54;1.33] ¥ (p=0.48)* | 2, 85 vs 98 | Christodoulakis, 2005;  Weltert, 2015 |
|  | ESA + IV iron  vs control  (placebo and/or IV iron or no treatment) | *Subgroup analysis ESA + IV iron:*  Not statistically significant:  8/76 vs 15/82 §  RR: 0.60, 95%CI [0.28;1.31] ¥ (p=0.20)* | 2, 76 vs 82 | Urena, 2017;  Yoo, 2011 |
| *Figure 4.10 A: ESA + iron vs control (placebo and/or iron or no treatment) – Atrial fibrillation*  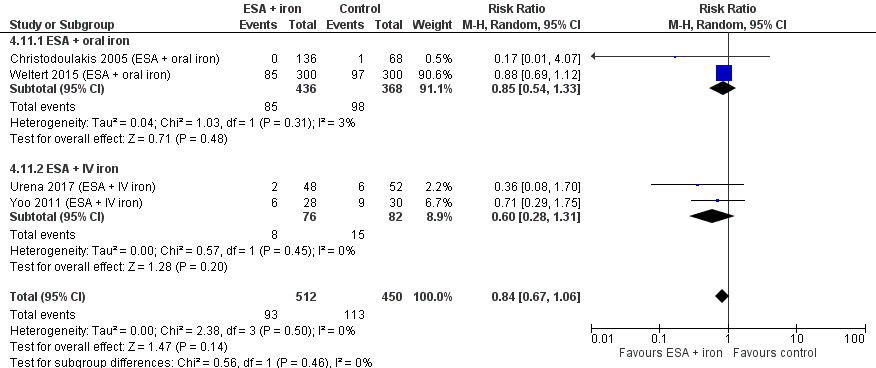 | | | | |

| **Heart failure (Figure 4.10 B)** | | | | |
| --- | --- | --- | --- | --- |
| Heart failure | ESA + iron  vs control  (placebo and/or iron or no treatment) | *Meta-analysis:* Not statistically significant: 8/236 vs 4/177 § RR: 1.45, 95%CI [0.41;5.15] ¥ (p=0.56)* | 3, 236 vs 177 | Christodoulakis, 2005;  Kettelhack, 1998; Urena, 2017 |
|  |  | *Sensitivity analysis excluding studies that are not in line with the  WHO definition of anaemia*  Not performed, as Kettelhack 1998 is the only study that used an alternative definition of anaemia (Hb>8.5 and ≤13.5 g/dl), and exclusion of this study would not have any influence on the results | N/A | N/A |
|  |  | *Sensitivity analysis excluding studies with high risk of bias on the items ‘inadequate measurement of the outcomes’ and ‘inadequate selection of the reported results’*  Not performed, as all 3 studies are at high risk of bias for both items | N/A | N/A |
|  | ESA + oral iron  vs control  (placebo and/or oral iron or no treatment) | *Subgroup analysis ESA + oral iron:*  Not statistically significant: 7/188 vs 2/125 § RR: 2.15, 95%CI [0.48;9.64] ¥ (p=0.32)* | 2, 188 vs 125 | Christodoulakis, 2005;  Kettelhack, 1998 |
|  | ESA + IV iron  vs control  (placebo and/or IV iron or no treatment) | *Subgroup analysis ESA + IV iron:*  Not statistically significant: 1/48 vs 2/52 § RR: 0.54, 95%CI [0.05;5.78] ¥ (p=0.61)* | 1, 48 vs 52 | Urena, 2017 |
| *Figure 4.10 B: ESA + iron vs control (placebo and/or iron or no treatment) – Heart failure*  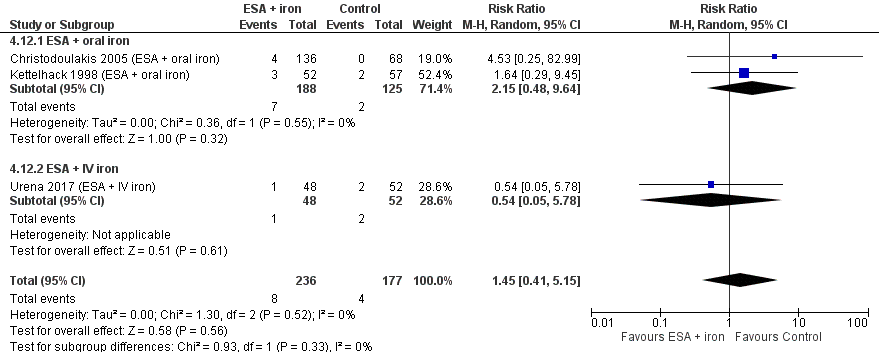 | | | | |
| Heart failure | ESA + IV iron  vs control  (placebo and/or IV iron or no treatment) | Statistically significant:  19/75 vs 26/59 §  RR: 0.57, 95%CI [0.35;0.93] (p=0.02)*  *in favour of ESA + IV iron* | 1, 75 vs 59 | Cladellas, 2012 (cohort) |

| **Hypertension** | | | | |
| --- | --- | --- | --- | --- |
| ***Findings presented in Figure 4.10 C*** | | | | |
| Hypertension | ESA + iron  vs control  (placebo and/or iron or no treatment) | *Meta-analysis:* Not statistically significant: 4/439 vs 4/448 § RR: 1.05, 95%CI [0.26;4.26] ¥ (p=0.94)* | 5, 439 vs 448 | Dousias, 2003; Weltert, 2015; Kosmadakis, 2003;  Urena, 2017; Yoo, 2011 |
|  |  | *Sensitivity analysis excluding studies that are not in line with the  WHO definition of anaemia*  Not performed, as 3 of these 5 studies have non-estimable effect sizes | N/A | N/A |
|  |  | *Sensitivity analysis excluding studies with high risk of bias on the items ‘inadequate measurement of the outcomes’ and ‘inadequate selection of the reported results’*  Not performed, as 3 of these 5 studies have non-estimable effect sizes | N/A | N/A |
|  | ESA + oral iron  vs control  (placebo and/or oral iron or no treatment) | *Subgroup analysis ESA + oral iron:*  Not statistically significant: 1/323 vs 2/327 § RR: 0.50, 95%CI [0.05;5.48] ¥ (p=0.57)* | 2, 323 vs 327 | Dousias, 2003; Weltert, 2015 |
|  | ESA + IV iron  vs control  (placebo and/or IV iron or no treatment) | *Subgroup analysis ESA + IV iron:*  Not statistically significant: 3/116 vs 2/121 § RR: 1.55, 95%CI [0.28;8.64] ¥ (p=0.62)* | 3, 116 vs 121 | Kosmadakis, 2003; Urena, 2017; Yoo, 2011 |
| *Figure 4.10 C: ESA + iron vs control (placebo and/or iron or no treatment) – Hypertension*  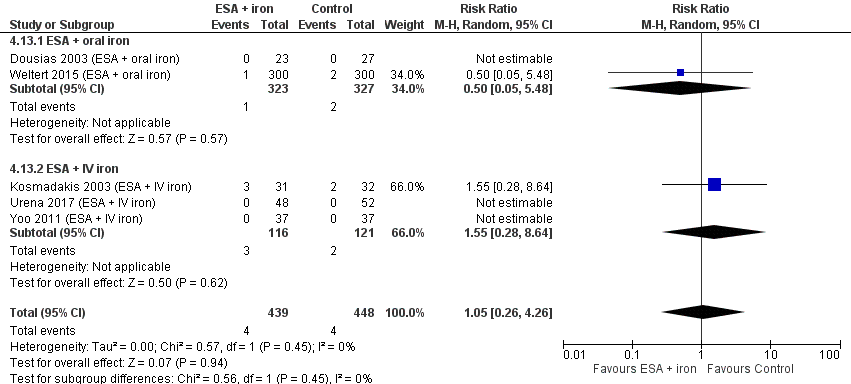 | | | | |
| ***Additional findings (not presented in Figure 4.10 C)*** | | | | |
| Prolonged hypertension or exacerbation of preexisting elevated blood pressure | ESA + oral iron  vs control  (placebo and/or oral iron or no treatment) | 0/29 vs 0/29 §  Effect size not estimable | 1, 29 vs 29 | Scott, 2002 |

| **Other cardiovascular events** | | | | |
| --- | --- | --- | --- | --- |
| ***Findings presented in Figure 4.10 D*** | | | | |
| Cardiac tamponade | ESA + iron vs control  (placebo and/or IV iron or no treatment) | *Meta-analysis:*  Not statistically significant: 4/348 vs 6/352 §  RR: 0.68, 95%CI [0.19;2.40] ¥ (p=0.55)* | 2, 348 vs 352 | Weltert, 2015;  Urena, 2017 |
| Cardiac arrest | ESA + oral iron  vs control  (placebo and/or oral iron or no treatment) | Not statistically significant: 4/136 vs 0/68 §  RR: 4.53, 95%CI [0.25;82.99] ¥ (p=0.31)* | 1, 136 vs 68 | Christodoulakis, 2005 |
|  |  | Not statistically significant:  1/28 vs 0/53 §  RR: 5.59, 95%CI [0.24;132.81] ¥ (p=0.29)* | 1, 28 vs 53 | Delasotta 2012 B (cohort) |
| Tachycardia | ESA + IV iron  vs control  (placebo and/or IV iron or no treatment) | 0/37 vs 0/37 §  Effect size not estimable | 1, 37 vs 37 | Yoo, 2011 |
| Annulus rupture |  | Not statistically significant:  0/48 vs 1/52 §  RR: 0.36, 95%CI [0.02;8.64] ¥ (p=0.53)* | 1, 48 vs 52 | Urena, 2017 |
| Coronary artery occlusion |  | 0/48 vs 0/52 §  Effect size not estimable |  |  |
| Vascular complications |  | Not statistically significant:  5/48 vs 3/52 §  RR: 1.81, 95%CI [0.46;7.15] ¥ (p=0.40)* |  |  |
| Moderate aortic regurgitation |  | Not statistically significant:  5/48 vs 8/52 §  RR: 0.68, 95%CI [0.24;1.93] ¥ (p=0.47)* |  |  |
| Severe hypotension requiring haemodynamic support |  | Not statistically significant:  3/48 vs 4/52 §  RR: 0.81, 95%CI [0.19;3.45] ¥ (p=0.78)* |  |  |
| Major adverse cardiovascular events |  | Statistically significant:  30/75 vs 47/59 §  RR: 0.50, 95%CI [0.37;0.68] (p<0.0001)*  *in favour of ESA + IV iron* | 1, 75 vs 59 | Cladellas, 2012 (cohort) |
| Cardiovascular events |  | Not statistically significant:  1/35 vs 0/32 § RR: 2.75, 95%CI [0.12;65.18] ¥ (p=0.53)* | 1, 35 vs 32 | Cao, 2020 |
| ***Additional findings (not presented in Figure 4.10 D)*** | | | | |
| Myocardial injury: peak of troponin (µg/l, median (IQR)) | ESA + IV iron  vs control  (placebo and/or IV iron or no treatment) | Not statistically significant:  0.22 (0.15-0.55) vs 0.18 (0.11-0.38) β: 0.12, 95%CI [-0.33;0.279] (adjusted p=0.12) | 1, 48 vs 52 § | Urena, 2017 |
| Myocardial injury: peak of creatine kinase-MB (µg/l, median (IQR)) |  | Not statistically significant:  11.25 (6.52-15.93) vs 8.80 (5.60-15.15) β: 1.87, 95%CI [-1.44;5.14] (adjusted p=0.27) |  |  |
| Perioperative myocardial infarction |  | Not statistically significant: £†  (p>0.05) | 1, 75 vs 59 § | Cladellas, 2012 (cohort) |
| Cardiac tamponade |  | Not statistically significant: £†  (p>0.05) |  |  |
| Permanent cerebral vascular accident |  | Not statistically significant: £†  (p>0.05) |  |  |

*Figure 4.10 D: ESA + iron vs control (placebo and/or iron or no treatment) – Cardiovascular adverse events*

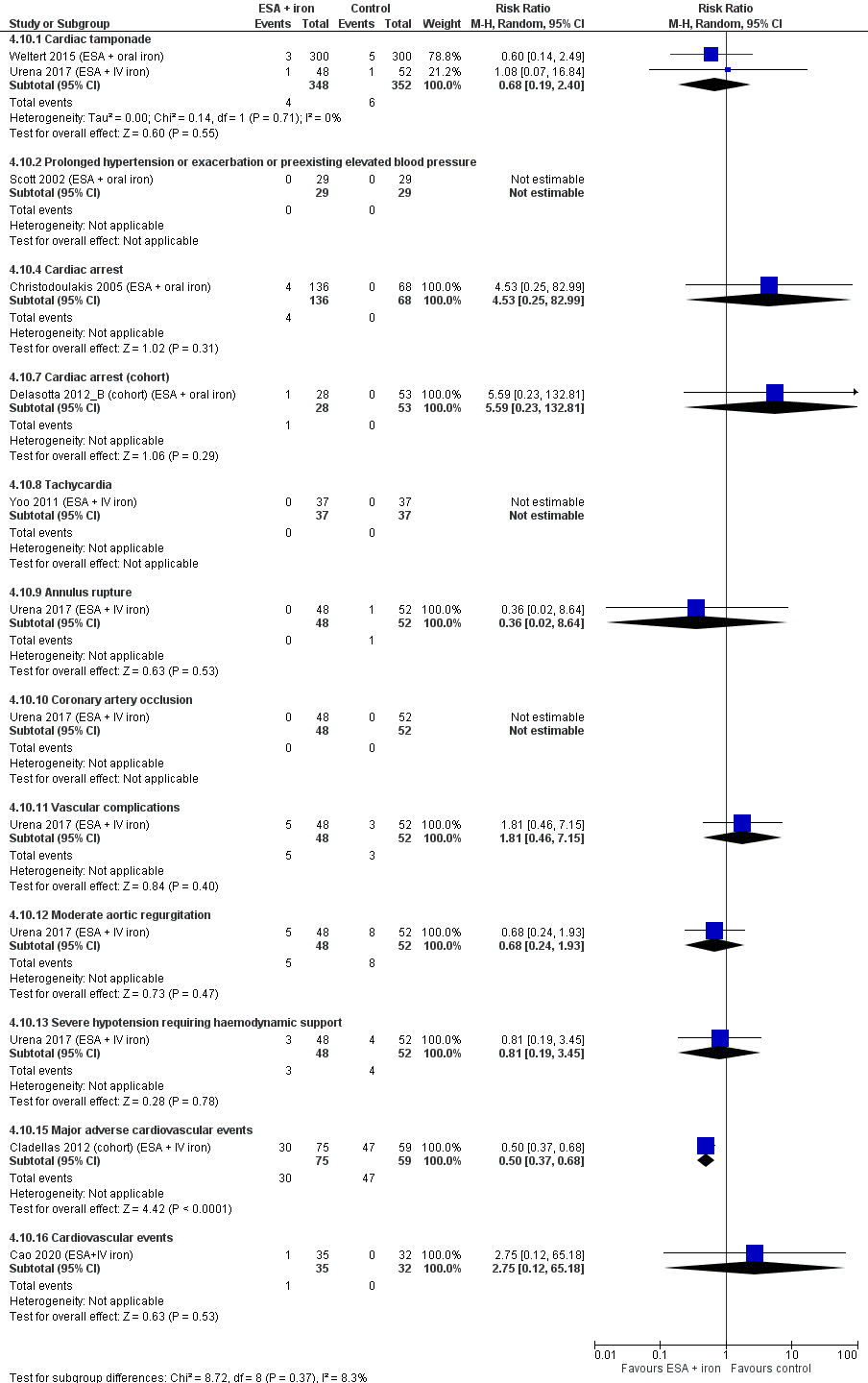

| **4.11 Renal (Figure 4.11)** | | | | |
| --- | --- | --- | --- | --- |
| Need for renal replacement therapy | ESA + oral iron  vs control  (placebo and/or oral iron or no treatment) | Not statistically significant:  14/300 vs 15/300 §  RR: 0.93, 95%CI [0.46;1.90] ¥ (p=0.85)* | 1, 300 vs 300 | Weltert, 2015 |
| New onset renal failure |  | Not statistically significant:  2/300 vs 1/300 §  RR: 2.00, 95%CI [0.18;21.94] ¥ (p=0.57)* |  |  |
| Acute renal failure | ESA + IV iron  vs control  (placebo and/or IV iron or no treatment) | Statistically significant:  22/75 vs 32/59 §  RR: 0.54, 95%CI [0.35;0.83] (p=0.004)* *in favour of ESA + IV iron* | 1, 75 vs 59 | Cladellas, 2012 (cohort) |
| Need for dialysis |  | 0/48 vs 0/52 §  Effect size not estimable | 1, 48 vs 52 | Urena, 2017 |
| *Figure 4.11: ESA + iron vs control (placebo and/or iron or no treatment) – Renal adverse events*  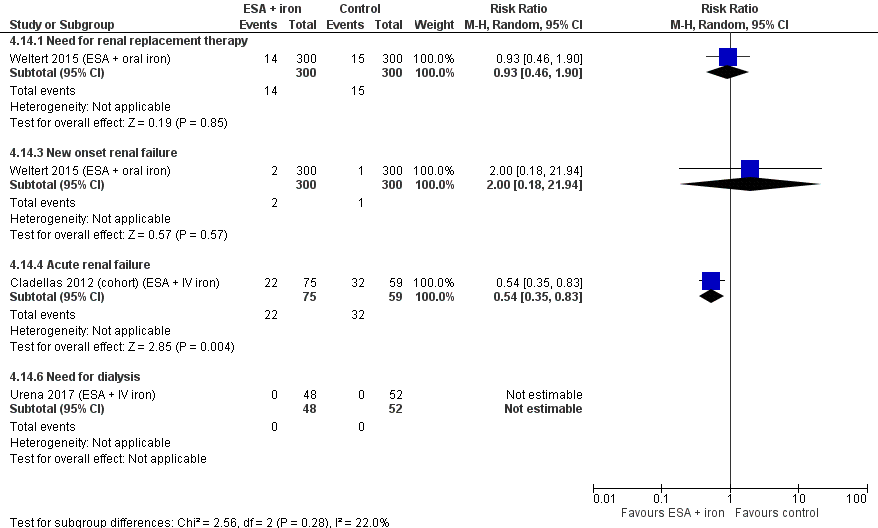 | | | | |

| **4.12 Anaemia-associated ischemic events** | | | | |
| --- | --- | --- | --- | --- |
| **Myocardial infarction (Figure 4.12 A)** | | | | |
| Myocardial infarction | ESA + iron  vs control  (placebo and/or iron or no treatment) | *Meta-analysis:*  Not statistically significant: 13/842 vs 9/859 §  RR: 1.40, 95%CI [0.61;3.17] ¥ (p=0.43)* | 5, 842 vs 859 | Scott, 2002; So-Osman, 2014; Stowell, 2009; Weltert, 2015; Urena, 2017 |
|  |  | *Sensitivity analysis excluding studies that are not in line with the  WHO definition of anaemia (only including So-Osman and Urena)* Not statistically significant: 4/173 vs 2/190 §  RR: 2.19, 95%CI [0.41;11.75] ¥ (p=0.36)* | 2, 173 vs 190 | So-Osman, 2014; Urena, 2017 |
|  |  | *Sensitivity analysis excluding studies with high risk of bias on the items ‘inadequate measurement of the outcomes’ and ‘inadequate selection of the reported results’*  Not performed, as Stowell 2009 and Weltert 2015 are the only studies that are at low risk of bias for one of the items, and only including these studies would not influence the results | N/A | N/A |
|  | ESA + oral iron  vs control  (placebo and/or oral iron or no treatment) | *Subgroup analysis ESA + oral iron:*  Not statistically significant: 11/794 vs 8/807 §  RR: 1.31, 95%CI [0.55;3.15] ¥ (p=0.54)* | 4, 794 vs 807 | Scott, 2002; So-Osman, 2014; Stowell, 2009; Weltert, 2015 |
|  | ESA + IV iron  vs control  (placebo and/or IV iron or no treatment) | *Subgroup analysis ESA + IV iron:*  Not statistically significant: 2/48 vs 1/52 §  RR: 2.17, 95%CI [0.20;23.14] ¥ (p=0.52)* | 1, 48 vs 52 | Urena, 2017 |
| *Figure 4.12 A: ESA + iron vs control (placebo and/or iron or no treatment) – Myocardial infarction*  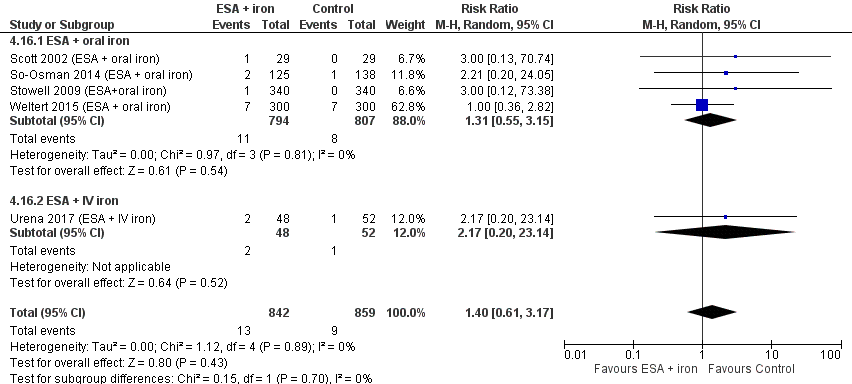 | | | | |
| **Stroke (Figure 4.12 B)** | | | | |
| Stroke | ESA + iron  vs control  (placebo and/or iron or no treatment) | *Meta-analysis:*  Not statistically significant: 7/417 vs 1/421 §  RR: 4.07, 95%CI [0.87;18.99] ¥ (p=0.07)* | 3, 417 vs 421 | Scott, 2002; Stowell, 2009; Urena, 2017 |
|  |  | *Sensitivity analysis excluding studies that are not in line with the  WHO definition of anaemia*  Not performed, as Urena 2017 is the only study in line with the WHO definition of anaemia, and only including these studies would not influence the results | N/A | N/A |
|  |  | *Sensitivity analysis excluding studies with high risk of bias on the items ‘inadequate measurement of the outcomes’ and ‘inadequate selection of the reported results’*  Not performed, as Stowell 2009 is the only study that is at low risk of bias for one of the items | N/A | N/A |
|  | ESA + oral iron  vs control  (placebo and/or oral iron or no treatment) | *Subgroup analysis ESA + oral iron:*  Not statistically significant: 4/369 vs 0/369 §  RR: 5.00, 95%CI [0.59;42.10] ¥ (p=0.14)* | 2, 369 vs 369 | Scott, 2002; Stowell, 2009 |
|  | ESA + IV iron  vs control  (placebo and/or IV iron or no treatment) | *Subgroup analysis ESA + IV iron:*  Not statistically significant: 3/48 vs 1/52 § RR: 3.25, 95%CI [0.35;30.19] ¥ (p=0.30)* | 1, 48 vs 52 | Urena, 2017 |
| *Figure 4.12 B: ESA + iron vs control (placebo and/or iron or no treatment) – Stroke*  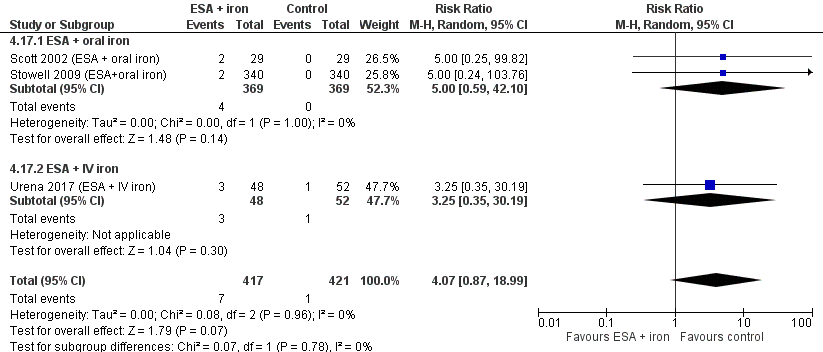 | | | | |
| **Other anaemia-associated ischemic events (Figure 4.12 C)** | | | | |
| Myocardial ischemia | ESA + oral iron  vs control  (placebo and/or oral iron or no treatment) | Not statistically significant:  1/340 vs 0/340 §  RR: 3.00, 95%CI [0.12;73.38] ¥ (p=0.50)* | 1, 340 vs 340 | Stowell, 2009 |
| TIA |  | Not statistically significant:  1/340 vs 0/340 §  RR: 3.00, 95%CI [0.12;73.38] ¥ (p=0.50)* |  |  |
| Stroke/TIA |  | Not statistically significant:  2/125 vs 0/138 §  RR: 5.52, 95%CI [0.27;113.80] ¥ (p=0.27)* | 1, 125 vs 138 | So-Osman, 2014 |
| Bowel ischemia |  | Not statistically significant:  2/300 vs 4/300 §  RR: 0.50, 95%CI [0.09;2.71] ¥ (p=0.42)* | 1, 300 vs 300 | Weltert, 2015 |
| Acute kidney injury | ESA + IV iron  vs control  (placebo and/or IV iron or no treatment) | *Meta-analysis:* Not statistically significant:  13/85 vs 22/87 §  RR: 0.67, 95%CI [0.22;2.01] ¥ (p=0.48)* | 2, 85 vs 87 | Urena, 2017; Yoo, 2011 |
| Ischemic events |  | Not statistically significant:  0/48 vs 1/52 §  RR: 0.36, 95%CI [0.02;8.64] ¥ (p=0.53)* | 1, 48 vs 52 | Urena, 2017 |

*Figure 4.12 C: ESA + iron vs control (placebo and/or iron or no treatment) – Other anaemia-associated ischemic events*

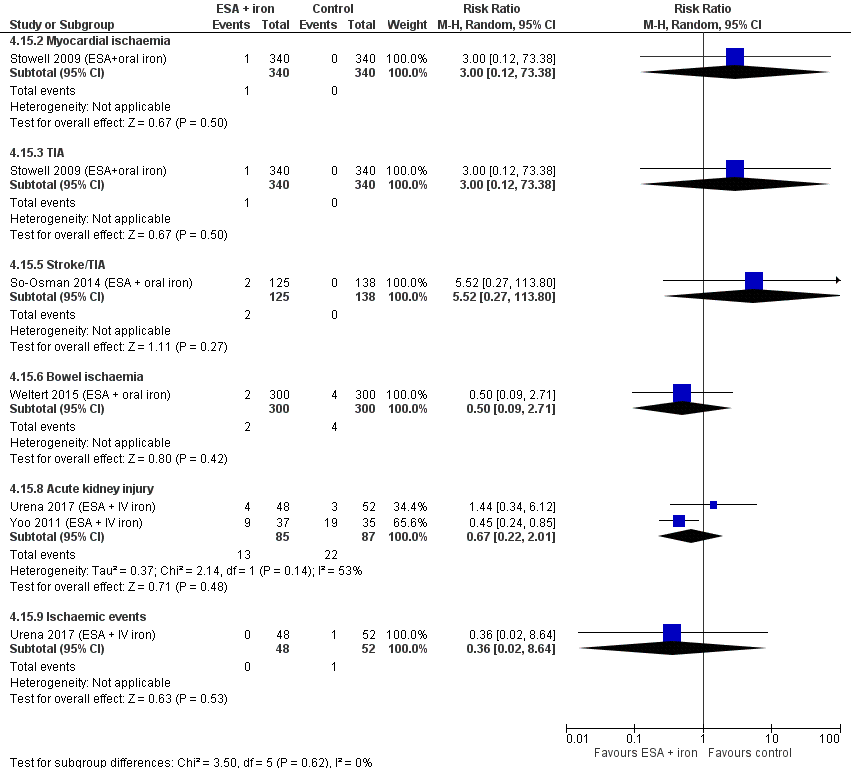

| **4.13 Thromboembolic events** | | | | |
| --- | --- | --- | --- | --- |
| **Deep venous thrombosis - RCTs** | | | | |
| ***Findings presented in Figure 4.13 A*** | | | | |
| Deep venous thrombosis | ESA + iron  vs control  (placebo and/or iron or no treatment) | *Meta-analysis:*  Not statistically significant: 25/915 vs 15/924 §  RR: 1.62, 95%CI [0.85;3.09] ¥ (p=0.14)* | 8, 915 vs 924 | Cao, 2020; Heiss, 1996; Qvist 1999/2000; Scott, 2002; So-Osman, 2014; Stowell, 2009; Weltert, 2015; Kosmadakis, 2003 |
|  |  | *Sensitivity analysis excluding studies that are not in line with the  WHO definition of anaemia*  Not performed, as Cao 2020 and So-Osman 2014 are the only studies in line with the WHO definition of anaemia, and So-Osman's effect size is not estimable | N/A | N/A |
|  |  | *Sensitivity analysis excluding studies with high risk of bias on the items ‘inadequate measurement of the outcomes’ and ‘inadequate selection of the reported results’*  Not performed, as Cao 2020, Stowell 2009 and Weltert 2015 are the only studies that are at low risk of bias for one of the items, and only including these studies would not influence the results | N/A | N/A |
|  | ESA + oral iron  vs control  (placebo and/or oral iron or no treatment) | *Subgroup analysis ESA + oral iron:*  Not statistically significant: 20/849 vs 12/860 §  RR: 1.49, 95%CI [0.59;3.75] ¥ (p=0.40)* | 6, 849 vs 860 | Heiss, 1996; Qvist 1999/2000; Scott, 2002; So-Osman, 2014; Stowell, 2009; Weltert, 2015 |
|  | ESA + IV iron  vs control  (placebo and/or IV iron or no treatment) | *Subgroup analysis ESA + IV iron:*  Not statistically significant: 5/66 vs 3/64 § RR: 1.58, 95%CI [0.39;6.35] ¥ (p=0.52)* | 2, 66 vs 64 | Cao, 2020; Kosmadakis, 2003 |

| *Figure 4.13 A: ESA + iron vs control (placebo and/or iron or no treatment) – Deep venous thrombosis*  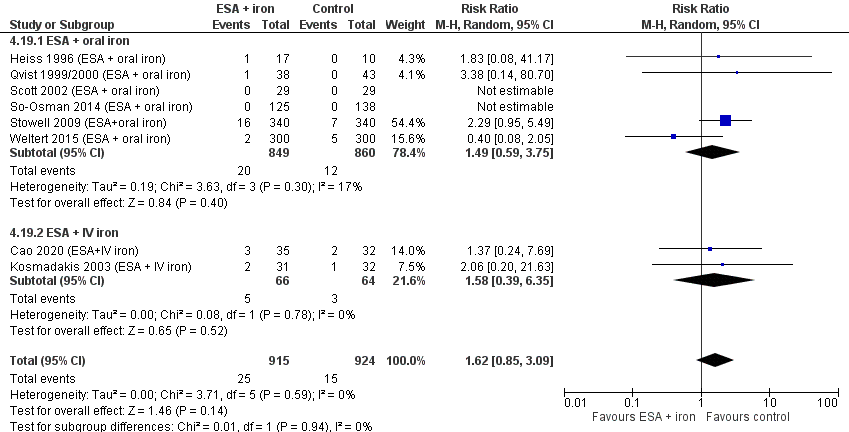 | | | | |
| --- | --- | --- | --- | --- |
| ***Additional findings (not presented in Figure 4.13 A)*** | | | | |
| Deep venous thrombosis rate (%) | ESA 300 IU/kg + oral iron vs placebo + oral iron | Not statistically significant: 11 vs 16 £† (p>0.05) | 1, number of participants not reported ££ | de Andrade, 1996 |
|  | ESA 100 IU/kg + oral iron vs placebo + oral iron | Not statistically significant: 3 vs 16 £† (p>0.05) | 1, number of participants not reported ££ |  |

| **Deep venous thrombosis – Cohort studies (Figure 4.13 B)** | | | | |
| --- | --- | --- | --- | --- |
| Deep venous thrombosis | ESA + iron  vs control  (placebo and/or iron or no treatment) | *Meta-analysis:*  Not statistically significant: 2/74 vs 1/148 §  RR: 2.77, 95%CI [0.45;17.17] ¥ (p=0.27)* | 3, 74 vs 148 | Delasotta, 2012 A (cohort);  Delasotta, 2012 B (cohort); Ye, 2017 (cohort) |
|  | ESA + oral iron  vs control  (placebo and/or oral iron or no treatment) | *Subgroup analysis ESA + oral iron:*  Not statistically significant: 2/44 vs 0/81 §  RR: 5.34, 95%CI [0.57;49.80] ¥ (p=0.14)* | 2, 44 vs 81 | Delasotta, 2012 A (cohort);  Delasotta, 2012 B (cohort) |
|  | ESA + IV iron  vs control  (placebo and/or IV iron or no treatment) | *Subgroup analysis ESA + IV iron:*  Not statistically significant: 0/30 vs 1/67 §  RR: 0.73, 95%CI [0.03;17.45] ¥ (p=0.85)* | 1, 30 vs 67 | Ye, 2017 (cohort) |
| *Figure 4.13 B: ESA + iron vs control (placebo and/or iron or no treatment) – Deep venous thrombosis – cohort studies*  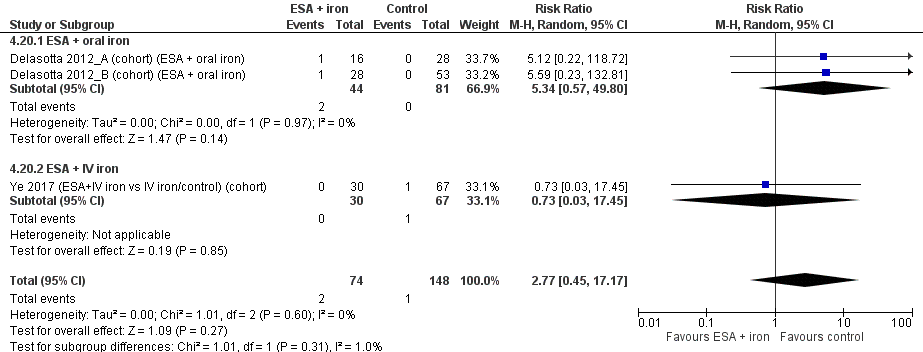 | | | | |

| **Other thromboembolic events (Figure 4.13 C)** | | | | |
| --- | --- | --- | --- | --- |
| Mesenteric venous thrombosis | ESA + oral iron  vs control  (placebo and/or oral iron or no treatment) | Not statistically significant:  0/17 vs 1/10 §  RR: 0.20, 95%CI [0.01;4.57] ¥ (p=0.32)* | 1, 17 vs 10 | Heiss, 1996 |
| Arterial thrombosis |  | Not statistically significant:  1/52 vs 0/57 §  RR: 3.28, 95%CI [0.14;78.86] ¥ (p=0.46)* | 1, 52 vs 57 | Kettelhack, 1998 |
| Thrombosis | ESA + IV iron  vs control  (placebo and/or IV iron or no treatment) | 0/30 vs 0/67 §  Effect size not estimable | 1, 30 vs 67 | Ye, 2017 (cohort) |
| Pulmonary embolism | ESA + oral iron  vs control  (placebo and/or oral iron or no treatment) | *Meta-analysis:* Not statistically significant:  0/465 vs 3/478 §  RR: 0.62, 95%CI [0.03;14.76] ¥ (p=0.20)* | 2, 465 vs 478 | So-Osman, 2014; Stowell, 2009 |
|  |  | Not statistically significant:  0/28 vs 1/53 §  RR: 0.62, 95%CI [0.03;14.76] ¥ (p=0.77)* | 1, 28 vs 53 | Delasotta, 2012 B |
| Embolism |  | Not statistically significant:  1/136 vs 0/68 §  RR: 1.51, 95%CI [0.06;36.61] ¥ (p=0.80)* | 1, 136 vs 68 | Christodoulakis, 2005 |
| Valve embolization | ESA + IV iron  vs control  (placebo and/or IV iron or no treatment) | Not statistically significant: 0/48 vs 2/52 § RR: 0.22, 95%CI [0.01;4.39] ¥ (p=0.32)* | 1, 48 vs 52 | Urena, 2017 |
| Thrombotic and/or vascular events | ESA + oral iron  vs control  (placebo and/or oral iron or no treatment) | 0/29 vs 0/27 §  Effect size not estimable | 1, 29 vs 27 | Olijhoek, 2001 |
|  | ESA + IV iron  vs control  (placebo and/or IV iron or no treatment) | 0/29 vs 0/25 §  Effect size not estimable | 1, 29 vs 25 |  |

*Figure 4.13 C: ESA + iron vs control (placebo and/or iron or no treatment) – Thromboembolic events*

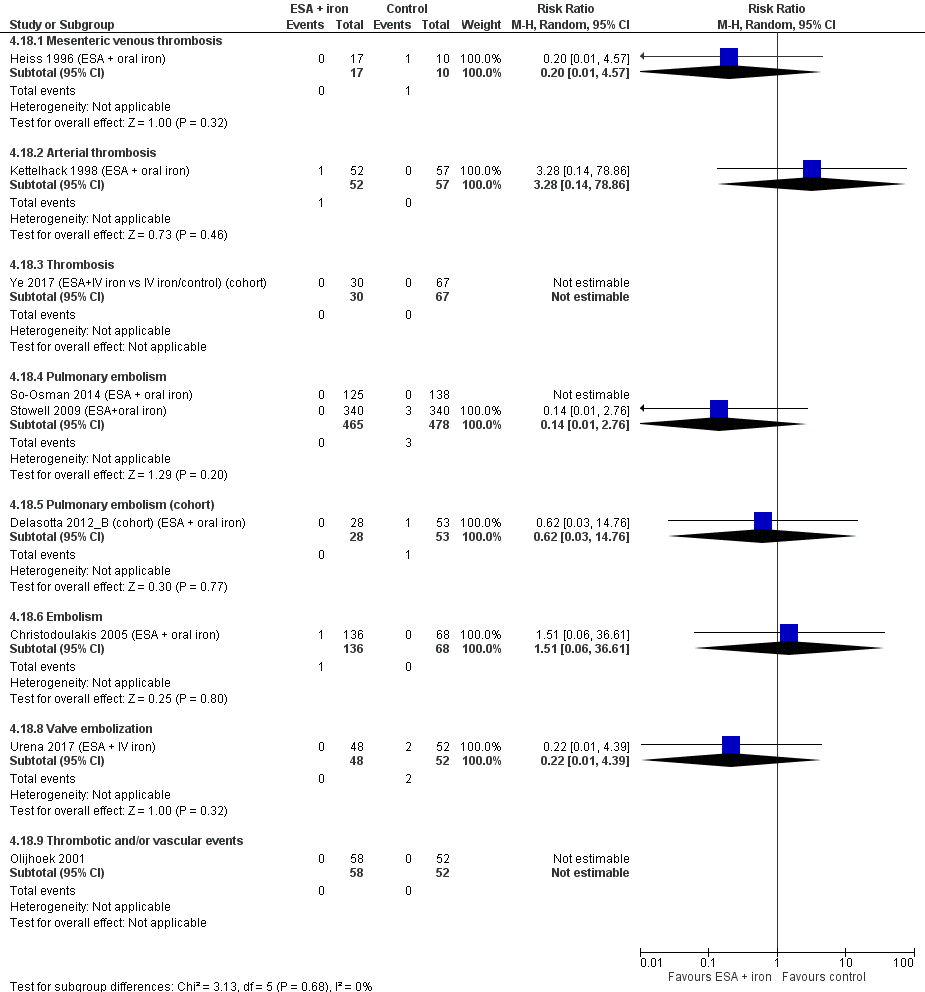

| **4.14 Mortality** | | | | |
| --- | --- | --- | --- | --- |
| **Mortality - RCTs (Figure 4.14 A)** | | | | |
| Mortality | ESA + iron  vs control  (placebo and/or iron or no treatment) | *Meta-analysis:*  Not statistically significant: 32/623 vs 26/548 §  RR: 1.32, 95%CI [0.56;3.12] ¥ (p=0.53)* | 7, 623 vs 548 | Christodoulakis, 2005;  Heiss, 1996; Kettelhack, 1998; Kosmadakis, 2003; Olijhoek, 2001; Scott, 2002; Weltert, 2015 |
|  |  | *Sensitivity analysis excluding studies that are not in line with the  WHO definition of anaemia*  Not performed, as none of the studies are in line with the WHO definition of anaemia | N/A | N/A |
|  |  | *Sensitivity analysis excluding studies with high risk of bias on the items ‘inadequate measurement of the outcomes’ and ‘inadequate selection of the reported results’*  Not performed, as Weltert 2015 is the only study that is at low risk of bias for one of the items, and only including this study would not influence the results | N/A | N/A |
|  | ESA + oral iron  vs control  (placebo and/or oral iron or no treatment) | *Subgroup analysis ESA + oral iron:*  Not statistically significant: 26/563 vs 13/491 §  RR: 1.75, 95%CI [0.76;4.02] ¥ (p=0.18)* | 6, 563 vs 491 | Christodoulakis, 2005;  Heiss, 1996; Kettelhack, 1998; Olijhoek, 2001; Scott, 2002; Weltert, 2015 |
|  | ESA + IV iron  vs control  (placebo and/or IV iron or no treatment) | *Subgroup analysis ESA + IV iron:*  Not statistically significant: 6/60 vs 13/57 §  RR: 0.48, 95%CI [0.21;1.09] ¥ (p=0.08)* | 2, 60 vs 57 | Kosmadakis, 2003; Olijhoek, 2001 |
| *Figure 4.14 A: ESA + iron vs control (placebo and/or iron or no treatment) – Mortality RCTs*  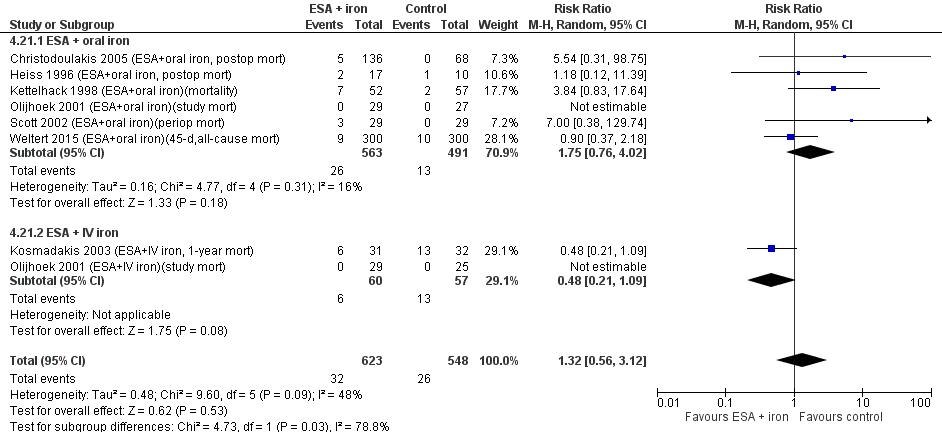 | | | | |
| **Mortality – Cohort studies (Figure 4.14 B)** | | | | |
| 7-month postoperative mortality | ESA + oral iron  vs control  (placebo and/or oral iron or no treatment) | Not statistically significant: 1/28 vs 0/53 §  RR: 5.59, 95%CI [0.23;132.81] ¥ (p=0.29)* | 1, 28 vs 53 | Delasotta, 2012 B (cohort) |
| Mortality | ESA + IV iron  vs control  (placebo and/or IV iron or no treatment) | *Meta-analysis:* Statistically significant:  7/145 vs 14/134 §  RR: 0.39, 95%CI [0.17;0.91] (p=0.03)* *in favour of ESA + IV iron* | 2, 145 vs 134 | Cladellas, 2012 (cohort); Pinilla-Gracia, 2020 (cohort) |
| *Figure 4.14 B: ESA + iron vs control (placebo and/or oral iron or no treatment) – Mortality cohort studies*  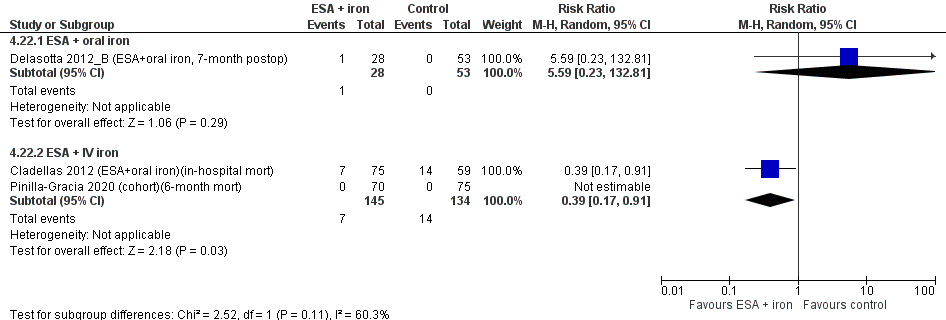 | | | | |

| **4.15 Other adverse events** | | | | |
| --- | --- | --- | --- | --- |
| ***Findings presented in Figure 4.15*** | | | | |
| Grade 2 neutropenia | ESA + oral iron  vs control  (placebo and/or oral iron or no treatment) | Not statistically significant:  1/136 vs 0/68 §  RR: 1.51, 95%CI [0.06;36.61] ¥ (p=0.80)* | 1, 136 vs 68 | Christodoulakis, 2005 |
| Multiorgan failure |  | Not statistically significant:  1/17 vs 0/10 §  RR: 1.83, 95%CI [0.08;41.17] ¥ (p=0.70)* | 1, 17 vs 10 | Heiss, 1996 |
| Muscle spasms |  | Not statistically significant:  24/340 vs 21/340 §  RR: 1.14, 95%CI [0.65;2.01] ¥ (p=0.64)* | 1, 340 vs 340 | Stowell, 2009 |
| Hypokalemia |  | Not statistically significant:  21/340 vs 19/340 §  RR: 1.11, 95%CI [0.61;2.02] ¥ (p=0.74)* |  |  |
| Insomnia |  | Not statistically significant:  17/340 vs 23/340 §  RR: 0.74, 95%CI [0.40;1.36] ¥ (p=0.33)* |  |  |
| Acute myeloid leukemia |  | Not statistically significant:  0/340 vs 1/340 §  RR: 0.33, 95%CI [0.1;8.15] ¥ (p=0.50)* |  |  |
| Mild and transient hypertonia |  | Not statistically significant:  2/17 vs 0/10 §  RR: 3.06, 95%CI [0.16;57.93] ¥ (p=0.46)* | 1, 17 vs 10 | Heiss, 1996 |
| Cachexia |  | Not statistically significant:  1/52 vs 0/57 §  RR: 3.28, 95%CI [0.14;78.86] ¥ (p=0.46)* | 1, 52 vs 57 | Kettelhack, 1998 |
| Non-thromboembolic complications (composite measure of prosthesis-related, cardiovascular, allergic, infectious, bleeding etc events) |  | Not statistically significant:  1/125 vs 4/138 §  RR: 0.28, 95%CI [0.03;2.44] ¥ (p=0.25)* | 1, 125 vs 138 | So-Osman, 2014 |
| Hypersensitivity reactions | ESA + IV iron  vs control  (placebo and/or IV iron or no treatment) | 0/48 vs 0/52 §  Effect size not estimable | 1, 48 vs 52 | Urena, 2017 |
| Allergy |  | 0/31 vs 0/32 §  Effect size not estimable | 1, 31 vs 32 | Kosmadakis, 2003 |
|  |  | 0/30 vs 0/67 §  Effect size not estimable | 1, 30 vs 67 | Ye, 2017 (cohort) |
|  |  | 0/35 vs 0/32 §  Effect size not estimable | 1, 35 vs 32 | Cao, 2020 |
| Hypercalcemia |  | 0/37 vs 0/37 §  Effect size not estimable | 1, 37 vs 37 | Yoo, 2011 |
| Convulsions |  | 0/48 vs 0/52 §  Effect size not estimable | 1, 48 vs 52 | Urena, 2017 |
| Conversion to open heart surgery |  | Not statistically significant:  1/48 vs 2/52 §  RR: 0.54, 95%CI [0.05;5.78] ¥ (p=0.61)* | 1, 48 vs 52 | Urena, 2017 |
| Need for a second valve |  | Not statistically significant:  1/48 vs 3/52 §  RR: 0.36, 95%CI [0.04;3.35] ¥ (p=0.37)* | 1, 48 vs 52 | Urena, 2017 |
| Postoperative complication rate (composite measure of anastomotic leak, abscess/fistula formation, hemorrhage, wound infection, pulmonary complications, complications from blood transfusions) |  | Statistically significant:  4/31 vs 13/32 §  RR: 0.32, 95%CI [0.12;0.87] (p=0.03)* *in favour of ESA + IV iron* | 1, 31 vs 32 | Kosmadakis, 2003 |
| Medical postoperative complications |  | Not statistically significant:  3/70 vs 10/75 § RR: 0.32, 95%CI [0.09;1.12] ¥ (p=0.07)* | 1, 70 vs 75 | Pinilla-Gracia, 2020 (cohort) |
| Surgical postoperative complications |  | Not statistically significant:  8/70 vs 6/75 §  RR: 1.43, 95%CI [0.52;3.91] ¥ (p=0.49)* |  |  |
| ***Additional findings (not presented in Figure 4.15)*** | | | | |
| Reoperation | ESA + IV iron  vs control  (placebo and/or IV iron or no treatment) | Not statistically significant: £†  (p>0.05) | 1, 75 vs 59 § | Cladellas, 2012 (cohort) |
| Thrombosis or dysfunction of the prosthesis |  | Not statistically significant: £†  (p>0.05) |  |  |

*Figure 4.15: ESA + iron vs control (placebo and/or iron or no treatment) – Other adverse events*

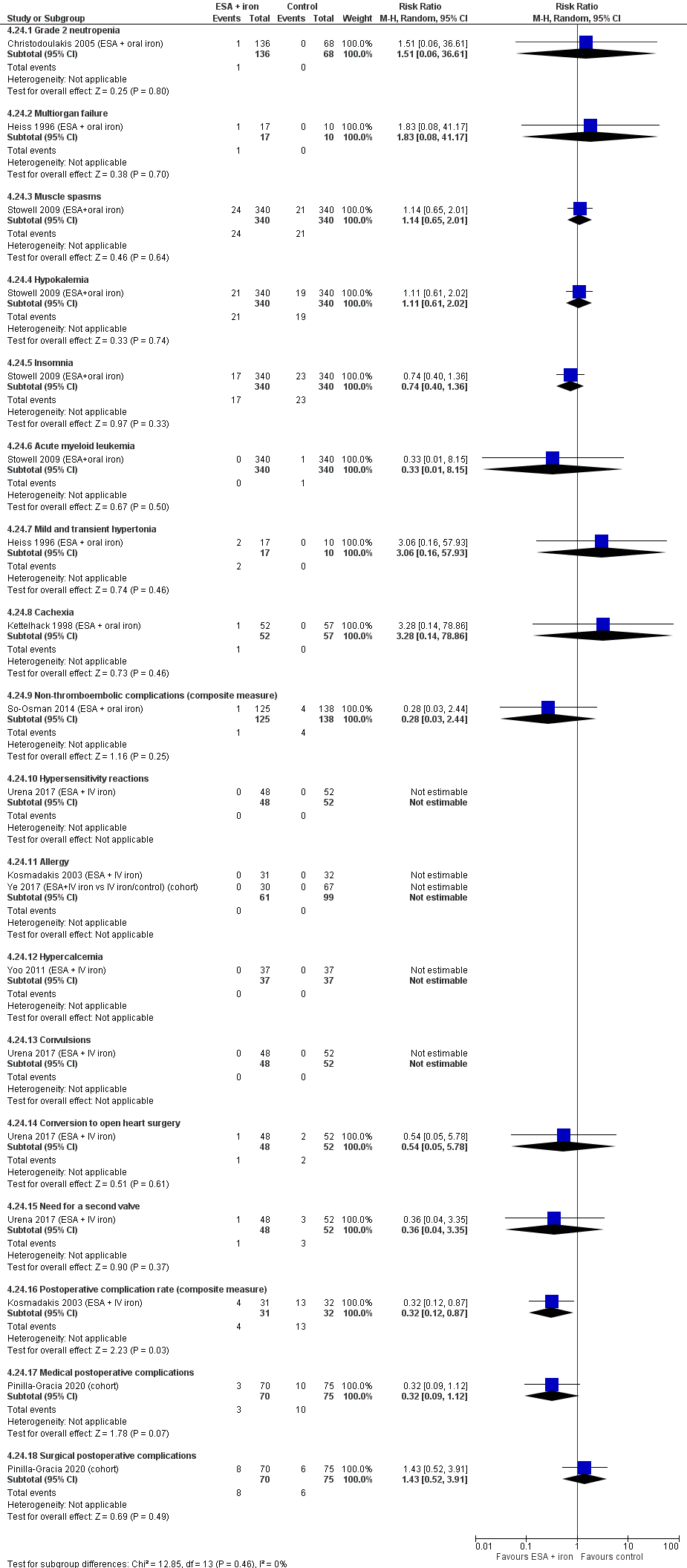

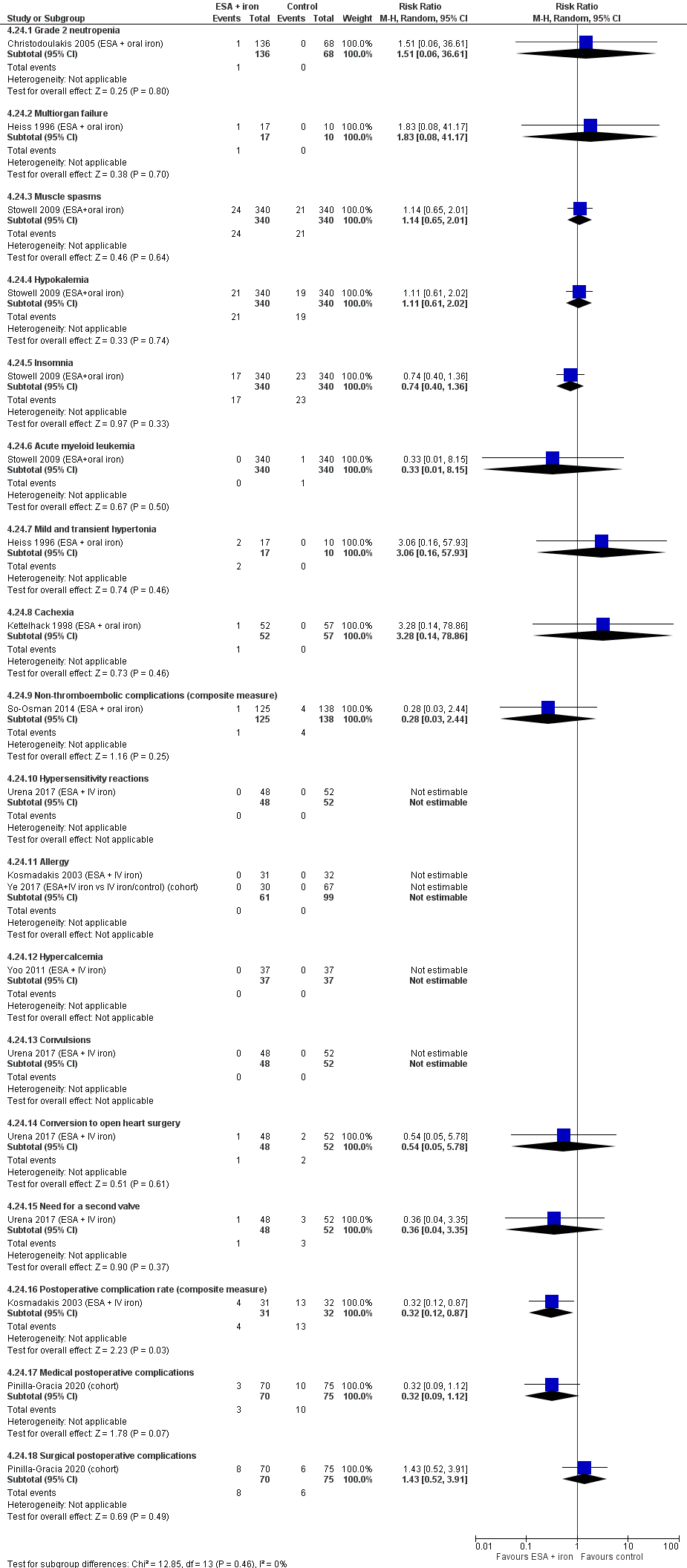

| **COMPARISON 5: ESA + IV IRON VS ESA + ORAL IRON** | | | | |
| --- | --- | --- | --- | --- |
| **5.1 Gastrointestinal (Figure 5.1)** | | | | |
| Digestive complications | ESA + IV iron vs ESA + oral iron | Statistically significant: 1/50 vs 26/50 § RR: 0.04, 95%CI [0.01;0.27] (p=0.001)* *in favour of ESA + IV iron* | 1, 50 vs 50 | Biboulet, 2018 |
| *Figure 5.1: ESA + IV iron vs ESA + oral iron – Gastrointestinal adverse events*  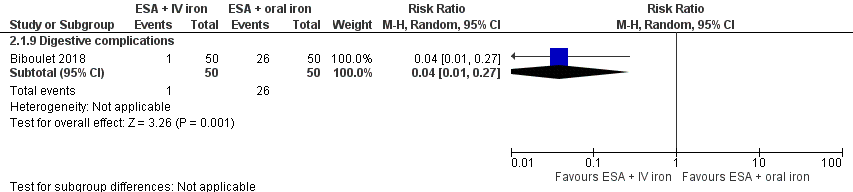 | | | | |
| **5.2 Infection (Figure 5.2)** | | | | |
| Preoperative prostatitis | ESA + IV iron vs ESA + oral iron | Not statistically significant: 1/50 vs 0/50 § RR: 3.00, 95%CI [0.13;71.92] ¥ (p=0.50)* | 1, 50 vs 50 | Biboulet, 2018 |
| *Figure 5.2: ESA + IV iron vs ESA + oral iron – Infectious adverse events*  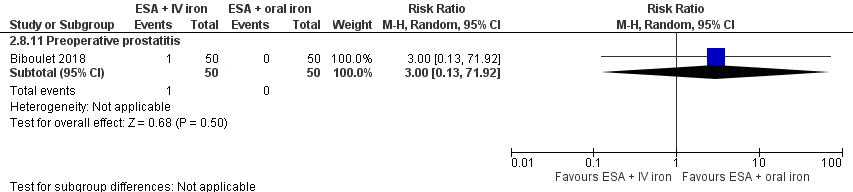 | | | | |
| **5.3 Cardiovascular** | | | | |
| Cardiac or respiratory failure | ESA + IV iron vs ESA + oral iron | 0/50 vs 0/50 § Effect size not estimable | 1, 50 vs 50 | Biboulet, 2018 |
| **5.4 Thromboembolic events (Figure 5.4)** | | | | |
| Deep venous thrombosis | ESA + IV iron vs ESA + oral iron | Not statistically significant:  1/50 vs 0/50 § RR: 3.00, 95%CI [0.13;71.92] ¥ (p=0.50)* | 1, 50 vs 50 | Biboulet, 2018 |
| Thrombotic and/or vascular events |  | 0/29 vs 0/29 § Effect size not estimable | 1, 29 vs 29 | Olijhoek, 2001 |
| Preoperative femoral vein thrombosis |  | Not statistically significant:  1/50 vs 0/50 § RR: 3.00, 95%CI [0.13;71.92] ¥ (p=0.50)* | 1, 50 vs 50 | Biboulet, 2018 |

| *Figure 5.4: ESA + IV iron vs ESA + oral iron – Thromboembolic adverse events*  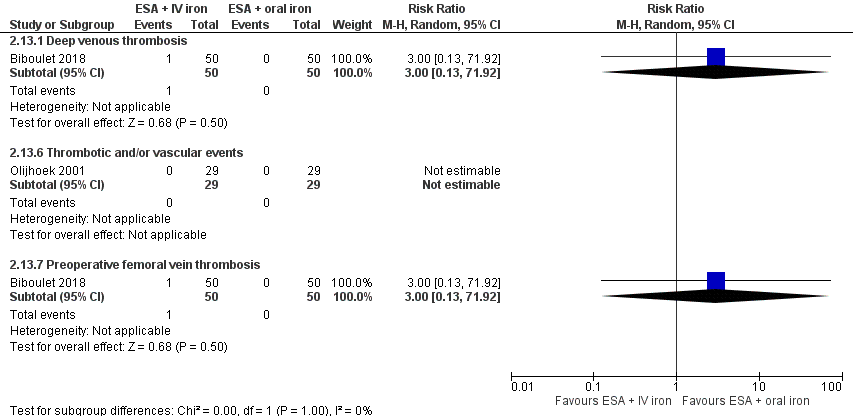 | | | | |
| --- | --- | --- | --- | --- |
| **5.5 Mortality** | | | | |
| Study mortality | ESA + IV iron vs ESA + oral iron | 0/29 vs 0/29 §  Effect size not estimable | 1, 29 vs 29 | Olijhoek, 2001 |
| Mortality during hospitalization |  | 0/50 vs 0/50 § Effect size not estimable | 1, 50 vs 50 | Biboulet, 2018 |

| **COMPARISON 6: ESA + IV IRON VS ESA + IV IRON (DIFFERENT ESA DOSES)** | | | | |
| --- | --- | --- | --- | --- |
| **6.1 Gastrointestinal (Figure 6.1)** | | | | |
| Intestinal obstruction | EPO (3^rd^ dose if Hb<15) + IV iron vs EPO (3^rd^ dose if Hb<13) + IV iron | Not statistically significant: 1/62 vs 0/65 §  RR: 3.14, 95%CI [0.13;75.72] ¥ (p=0.48)* | 1, 62 vs 65 | Rineau, 2017 (cohort) |
| *Figure 6.1: EPO (3^rd^ dose if Hb<15) + IV iron vs EPO (3^rd^ dose if Hb<13) + IV iron – Gastrointestinal adverse events*  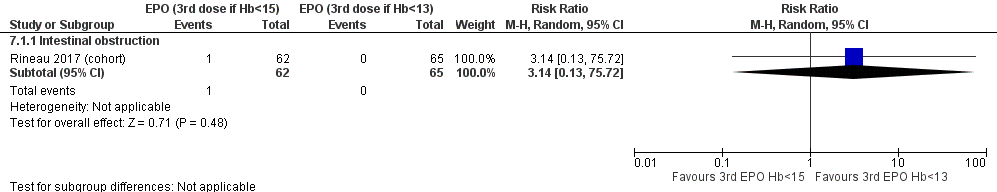 | | | | |
| **6.2 Bronchopulmonary** | | | | |
| Pulmonary edema | EPO (3^rd^ dose if Hb<15) + IV iron vs EPO (3^rd^ dose if Hb<13) + IV iron | 0/62 vs 0/65 §  Effect size not estimable | 1, 62 vs 65 | Rineau, 2017 (cohort) |
| Need for re-intubation |  | 0/62 vs 0/65 §  Effect size not estimable |  |  |
| Need for prolonged ventilation |  | 0/62 vs 0/65 §  Effect size not estimable |  |  |
| **6.3 Infection** | | | | |
| Pneumonia | EPO (3^rd^ dose if Hb<15) + IV iron vs EPO (3^rd^ dose if Hb<13) + IV iron | 0/62 vs 0/65 §  Effect size not estimable | 1, 62 vs 65 | Rineau, 2017 (cohort) |
| Wound infection |  | 0/62 vs 0/65 §  Effect size not estimable |  |  |
| Urinary tract infection |  | 0/62 vs 0/65 §  Effect size not estimable |  |  |
| **6.4 Bleeding (Figure 6.4)** | | | | |
| Postoperative hematoma | EPO (3^rd^ dose if Hb<15) + IV iron vs EPO (3^rd^ dose if Hb<13) + IV iron | 0/62 vs 0/65 §  Effect size not estimable | 1, 62 vs 65 | Rineau, 2017 (cohort) |
| Hemorrhagic shock |  | Not statistically significant:  0/62 vs 1/65 §  RR: 0.35, 95%CI [0.01;8.41] ¥ (p=0.52)* |  |  |

| *Figure 6.4: EPO (3^rd^ dose if Hb<15) + IV iron vs EPO (3^rd^ dose if Hb<13) + IV iron – Bleeding adverse events*  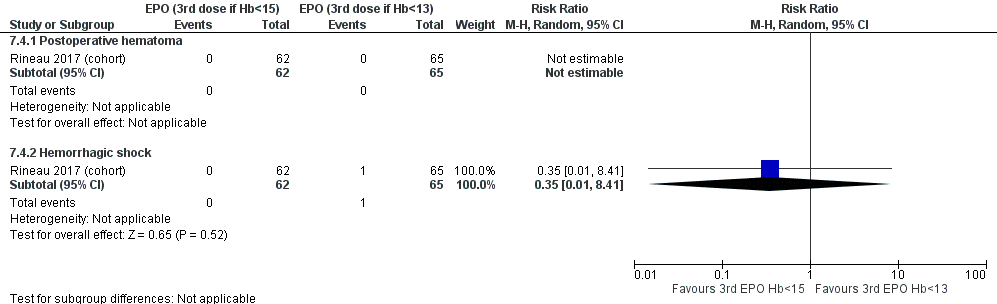 | | | | |
| --- | --- | --- | --- | --- |
| **6.5 Cardiovascular (Figure 6.5)** | | | | |
| Acute coronary syndrome | EPO (3^rd^ dose if Hb<15) + IV iron vs EPO (3^rd^ dose if Hb<13) + IV iron | 0/62 vs 0/65 §  Effect size not estimable | 1, 62 vs 65 | Rineau, 2017 (cohort) |
| Arrhythmia |  | 0/62 vs 0/65 §  Effect size not estimable |  |  |
| Cardiac arrest |  | 0/62 vs 0/65 §  Effect size not estimable |  |  |
| Atrial fibrillation |  | Not statistically significant:  1/62 vs 1/65 §  RR: 1.05, 95%CI [0.07;16.40] ¥ (p=0.97)* |  |  |
| *Figure 6.5: EPO (3^rd^ dose if Hb<15) + IV iron vs EPO (3^rd^ dose if Hb<13) + IV iron – Cardiovascular adverse events*  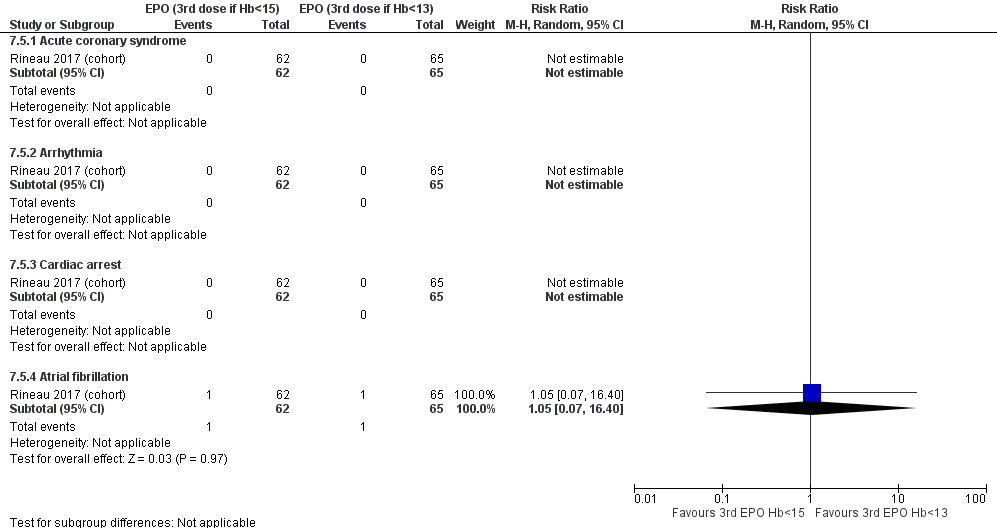 | | | | |

| **6.6 Anaemia-associated ischemic events** | | | | |
| --- | --- | --- | --- | --- |
| Acute limb ischemia | EPO (3^rd^ dose if Hb<15) + IV iron vs EPO (3^rd^ dose if Hb<13) + IV iron | 0/62 vs 0/65 §  Effect size not estimable | 1, 62 vs 65 | Rineau, 2017 (cohort) |
| Stroke |  | 0/62 vs 0/65 §  Effect size not estimable |  |  |
| Acute kidney injury |  | 0/62 vs 0/65 §  Effect size not estimable |  |  |
| **6.7 Thromboembolic events (Figure 6.7)** | | | | |
| Deep venous thrombosis | EPO (3^rd^ dose if Hb<15) + IV iron vs EPO (3^rd^ dose if Hb<13) + IV iron | Not statistically significant:  1/62 vs 0/65 §  RR: 3.14, 95%CI [0.13;75.72] ¥ (p=0.48)* | 1, 62 vs 65 | Rineau, 2017 (cohort) |
| Pulmonary embolism |  | 0/62 vs 0/65 §  Effect size not estimable |  |  |

| *Figure 6.7: EPO (3^rd^ dose if Hb<15) + IV iron vs EPO (3^rd^ dose if Hb<13) + IV iron – Thromboembolic adverse events*  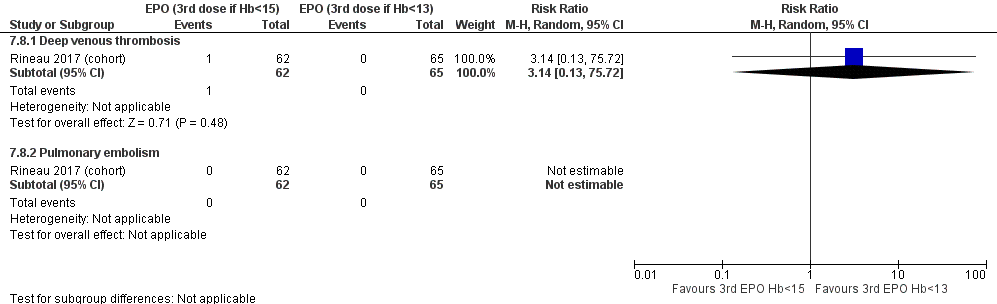 | | | | |
| --- | --- | --- | --- | --- |
| **6.8 Other adverse events (Figure 6.8)** | | | | |
| Allergy | EPO (3^rd^ dose if Hb<15) + IV iron vs EPO (3^rd^ dose if Hb<13) + IV iron | 0/62 vs 0/65 §  Effect size not estimable | 1, 62 vs 65 | Rineau, 2017 (cohort) |
| Epileptic seizure |  | Not statistically significant:  0/62 vs 1/65 §  RR: 0.35, 95%CI [0.01;8.41] ¥ (p=0.52)* |  |  |
| *Figure 6.8: EPO (3^rd^ dose if Hb<15) + IV iron vs EPO (3^rd^ dose if Hb<13) + IV iron – Other adverse events*  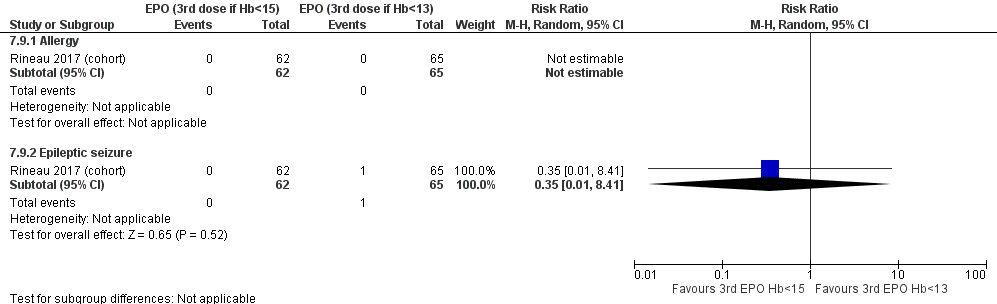 | | | | |

Table legend:

IQR: interquartile range
N/A: not applicable
£ No raw data available
££ Number of participants not reported

† Imprecision (lack of data)
§ Imprecision (limited sample size or low number of events)

¥ Imprecision (large variability of results)
